# Interplay of Human Metabolome and Gut Microbiome in Major Depression

**DOI:** 10.1101/2022.06.21.22276700

**Authors:** Najaf Amin, Jun Liu, Bruno Bonnechere, Siamak MehmoudianDehkordi, Matthias Arnold, Richa Batra, Yu-Jie Chiou, Marco Fernandes, M. Arfan Ikram, Robert Kraaij, Jan Krumsiek, Danielle Newby, Kwangsik Nho, Djawad Radjabzadeh, Andrew J Saykin, Liu Shi, William Sproviero, Laura Winchester, Yang Yang, Alejo J Nevado-Holgado, Gabi Kastenmüller, Rima F Kaddurah-Daouk, Cornelia M van Duijn

## Abstract

The pathogenesis of depression is complex involving the interplay of genetic and environmental risk factors including diet, lifestyle and the gut microbiome. Metabolomics studies may shed light on the interplay of these factors. We study over 63,000 individuals including 8462 cases with a lifetime major depression and 5403 cases with recurrent major depression from the UK Biobank profiled for nuclear magnetic resonance (NMR) spectroscopy based metabolites with the Nightingale platform. We identify 124 metabolites that are associated with major depressive disorder (MDD), including 49 novel associations. No differences were seen between the metabolic profiles of lifetime and recurrent MDD. We find that metabolites involved in the tricarboxylic acid (TCA) cycle are significantly altered in patients with MDD. Integrating the metabolic signatures of major depression and the gut microbiome, we find that the gut microbiome might play an important role in the relationship between these metabolites, lipoproteins in particular, and MDD. The order *Clostridiales*, and the phyla *Proteobacteria* and *Bacteroidetes* were the most important taxa, which link the lipoprotein particles to MDD. Our study shows that at the molecular level energy metabolism is disturbed in patients with MDD and that the interplay between the gut microbiome and blood metabolome may play a key role in the pathogenesis of MDD.

## Introduction

Major depression is an important determinant of population health, affecting people across the life span from adolescence to old age. The disease is associated with a plethora of debilitating symptoms beyond emotional dysregulation^1^, spanning from cognition, motoric function, neurovegetative symptoms to inflammation and disturbances of the immune system. MDD is further linked to increased risks of cardiometabolic disorders and mortality^2^. Currently, most antidepressant therapies modulate the monoamine pathway, but evidence is increasing for a more complex interplay of multiple pathways involving a wide range of metabolic alterations spanning energy metabolism^3^ lipid metabolism^4^ and inflammation^5^.

There is increased interest in energy metabolism and mitochondrial dysregulation as a contributor to the pathogenesis of major depression^6,7^. The brain has high aerobic activity requiring 20 times more energy than the rest of the body^8^ and is vulnerable to impaired energy production^6^. A decreased brain energy production has been found in depressed patients ^9^. An excess of structural mutations in the mitochondrial DNA have been reported in individuals diagnosed with depression compared to controls^6^. There is a high prevalence (54%) of depression in patients with mitochondrial diseases^10^. Symptoms of depression such as loss of energy and appetite, tiredness, weakness, cognitive impairments, and sleep disturbance are also frequent in mitochondrial diseases^9,11^ Mitochondria are cell organelles that provide energy in the form of adenosine triphosphate via oxidative phosphorylation. Simultaneously, mitochondria are also responsible for generating reactive oxygen species (ROS) and anti-oxidants such as creatine, coenzyme Q10, niconitamide and glutathione, which protect cells from various deleterious effects of ROS^12^. The imbalance in the generation of ROS and antioxidants leads to oxidative stress, damages to lipids and proteins, inflammation and apoptosis ^13,14^.

Disturbed plasma lipid concentrations have been implicated in the development of mitochondrial dysfunction^13^, with high density lipoprotein (HDL) inversely correlating with mitochondrial DNA damage^15^. A recent study using Nightingale’s proton nuclear magnetic resonance (NMR) metabolomics platform in nine Dutch cohorts (5,283 patients with depression and 10,145 controls) showed a shift towards decreased levels of high density lipoprotein (HDL)-cholesterol and increased levels of very low density lipoprotein (VLDL)-cholesterol and tri- and diglycerides particles in patients with depression ^16^. The relevance and molecular mechanisms underlying this shift are not understood. Gut microbiome has been shown to be a major determinant of the circulating lipids, specifically triglycerides and HDL^17^ and is also known to regulate mitochondrial function through the production of microbial metabolites including short chain fatty acids (SCFA), lipids, vitamins and amino acids.^18^ Disruptions to the gut microbiome have been found in patients with major depressive disorder^19,20^. Metabolic signatures of the gut microbiome can be found in feces and blood^17,21,22^. In our recent study connecting the gut microbiome with the Nightingale’s metabolites^21^ we observed association of metabolites in plasma with 32 gut microbial groups. A higher abundance of family *Christensenellaceae*, genera *ChristensenellaceaeR7 group, Ruminococcaceae* (*UCG002, UCG003, UCG005, UCG010, UCG014*), *Coprococcus, RuminococcaceaeNK4A214group* and *Ruminiclostridium6* associated with a favourable lipid profile, i.e., decreased VLDLs and increased HDLs. Interestingly, decreased abundance of the same groups of gut microbiota we also find associated with higher scores on depressive symptoms in our study of gut microbiome and depression^20^. This raises the questions 1) whether the gut microbiome explains part of the shift in VLDL and HDL levels seen in patients with depression ^16^ and 2) can we use the metabolic signatures of the disease based on Nightingale’s metabolites as a tool to infer the association between gut microbiome and the disease.

In this study, we harness the power of the UK biobank (UKB) to study over 63000 individuals including 8462 patients with MDD and 5403 patients with recurrent MDD, who are profiled for NMR spectroscopy based metabolites with the Nightingale platform. The largest study conducted to date discovers dysregulation of metabolites involved in the mitochondrial functioning in patients with MDD. When integrating the data with those of the Rotterdam Study to understand the interplay between the blood metabolome, gut microbiome and MDD, we find evidence that the altered gut microbiome in patients is a key player in the shift of the VLDL/HDL axis in MDD patients.

## Methods

### Study design and participants

The study was performed in the UK Biobank dataset, which comprises of more than 500,000 participants aged from 37 to 73 years during recruitment (2006 to 2010) for whom blood sampling was performed^23^. A random subset of 118,466 individuals was profiled for metabolites using a high-throughput ^1^H-NMR metabolomics (Nightingale Health, Helsinki, Finland) platform. The participants were registered with the UK National Health service and from 22 assessment centres across England, Wales, and Scotland using standardised procedures for data collection which included a wide range of questionnaires, anthropological measurement, clinical biomarkers, genotype data, etc. All participants provided electronically signed informed consent. UK Biobank has approval from the Northwest Multi-centre Research Ethics Committee, the Patient Information Advisory Group, and the Community Health Index Advisory Group. Further detail on the rationale, study design, survey methods, data collection are available elsewhere^23^. The current study is a part of UK Biobank projects 30418 and 54520.

### Definition of traits

For the initial analyses we considered two phenotypes including 1) lifetime major depressive disorder (MDD) and 2) recurrent major depressive disorder. Both lifetime and recurrent major depressive disorder were defined using the UK Biobank field code 20126, ICD10 codes F32 (single episode) and F33 (recurrent) or if participants were on antidepressant therapy at the baseline. Individuals who reported any other mental illnesses (e.g., bipolar disorder, schizophrenia, psychosis, etc.) were excluded from the study. Controls included individuals who had not reported depression at the baseline.

### Definition of covariates

The covariates considered in the analysis included baseline age, sex, ethnicity, fasting time, assessment center, lifestyle factors including body mass index (BMI), smoking status, alcohol intake frequency, education and status regarding multiple medication from touchscreen or verbal interview and technical variables during the NMR measurement, i.e., batch and spectrometer. Fasting time was defined as the time interval between the consumption of food or drink and blood sampling and natural log-transformed. Ethnicity was categorized to White, Asian (excluding Chinese), Black, Chinese, mixed and others. Smoking status was categorized to never, previous and current. Alcohol intake frequency was categorized to 1) daily or almost daily, 2) three to four times a week, 3) once or twice a week, 4) less than once a week. Education was categorized to 1) College or University degree, 2) A levels, advanced subsidiary (AS) levels or equivalent, 3) Certificated of secondary education (CSEs) or equivalent, 4) National vocational qualification (NVQ) or higher national diploma (HND) or higher national certificate (HNC) or equivalent, 5) O levels, general certificate of secondary education (GCSEs) or equivalent, 6) Other professional qualifications, and 7) none of the above based on the highest qualification. Information for those who chose “prefer not to answer”, was put as missing. Medication status was based on the medication codes collected from the verbal interview which were further coded to Anatomical Therapeutic Chemical (ATC) codes^9^. The medications considered in the covariates were selected based on our previous publication^24^, including five anti-hypertensives (C08, C09, C07, C03 and C02), anti-diabetes (metformin and other anti-diabetes under A10), lipid-lowering drugs (C10), digoxin (C01AA), anti-thrombotic (B01AC06), proton pump inhibitors (PPI, A02BC), hypnotics and sedatives (N05) and antidepressants (N06).

### Imputation of missing values in the covariates

Fast imputation of missing values by chained random forests was performed through the R package *missRanger* to impute the missing values for the shared covariates, including smoking status, BMI, alcohol intake frequency, education, and ethnicity. The information used in the imputation included baseline age, sex, smoking status, pack-years of smoking, alcohol intake frequency, physical activity from International Physical Activity Questionnaire (IPAQ) groups, ethnicity, BMI, education, blood pressure and waist-hip ratio. In brief, the large matrix was imputed with maximum of ten chaining interactions and 200 trees and weighted by the number of non-missing values; three candidate non-missing values were selected from in the predictive mean matching steps.

### Metabolite profiling

The metabolites were measured in plasma using the targeted high-throughput ^1^H-NMR metabolomics platform of Nightingale (Nightingale Health Ltd; biomarker quantification version 2020)^25,26^ which includes 249 metabolites. They include clinical lipids, lipoprotein subclass profiling with lipid concentrations within 14 subclasses, fatty acid composition, and various low-molecular weight metabolites such as amino acids, ketone bodies and glycolysis metabolites quantified in molar concentration units. The technology is based a standardized protocol of sample quality control and sample preparation, data storage and automated spectral analyses.

The data obtained from the baseline sampling was used^25^. For the samples with repeated measurements of the metabolites, one of the values was extracted at random. The metabolite values which were suggested to be technical errors in the quality control provided by Nightingale Health during the measurement procedure were treated as missing. A natural logarithm transformation of each metabolite was performed for the analysis. The zero values were replaced by the lowest value except for zero. Finally the transformed values were scaled to standard deviation units.

### Replication

For replication we considered the results from the previously published study performed in Dutch cohorts by the BBMRI-NL consortium^16^. The study included 5,283 patients with depression and 10,145 controls, who were characterized using the Nightingale platform. We further looked up the association of metabolites in the Predictors of Remission in Depression to Individual and Combined Treatments (PReDICT) study. The design and clinical outcomes of PReDICT have been detailed previously ^27^. Briefly, the PReDICT study aimed to identify predictors and moderators of response to 12 weeks of randomly-assigned treatment with duloxetine (30-60 mg/day), escitalopram (10-20 mg/day) or cognitive behaviour therapy (CBT, 16 one-hour individual sessions). Eligible participants were adults aged 18-65 years with an active untreated major depressive episode without psychotic features. Severity of depression at the randomization visit was assessed with the 17-item Hamilton Depression Rating Scale (HRSD_17_) ^28^. Eligibility required an HRSD_17_ score ≥18 at the screening visit and ≥15 at the randomization visit, indicative of moderate-to-severe depression. Active significant suicide risk, current illicit drug use (assessed with urine drug screen) or a history of substance abuse in the three months prior to randomization, pregnancy, lactation, and uncontrolled general medical conditions were all excluded. Details on metabolomics profiling and statistical analysis are provided in the **Supplemental text**.

### Statistical analysis

All analyses were performed in R statistical software. Descriptive analysis was performed using the ‘CBCgrps’ ^29^ library of R.

#### Metabolome-wide association analysis

We used logistic regression to test the association of the metabolite levels with lifetime and recurrent depression. We considered four models with increasing number of covariates in the subsequent models to identify the effects of most known confounders in the regression analysis. Model 1 was adjusted for age, sex, fasting time, ethnicity, assessment centre, and technical variables during the NMR measurement, i.e., batch and spectrometer; model 2 was additionally adjusted for BMI; model 3 further for antidepressant use and model 4 additionally adjusted for most known lifestyle factors including smoking status, alcohol intake frequency, physical activity, level of education and medication use for most chronic diseases. The current analysis included all 249 metabolites measured by Nightingale. False discovery rate (FDR) of 0.05 was used to identify significantly associated metabolites. We further performed a sensitivity analysis by removing those on antidepressant therapy. Finally, we performed forward regression analysis in R on model 4 adjusted residuals of depression-associated metabolites to identify independent metabolites. A multivariate regression analysis was performed including all metabolites selected from forward regression analysis. Multiple testing correction was performed using false discovery rate (FDR).

#### Integration of metabolic signatures of human gut microbiome and MDD

To identify patterns of correlation in the metabolic signatures of MDD and the human gut microbiome we estimated correlation coefficients from the effect estimates from linear mixed regression of 1) Z-score_MDD_ on Z-score_Microbe_ and 2) Z-score_Microbe_ on Z-score_MDD_. Z-score_MDD_ are the results of association analysis of MDD with Nightingale metabolites from the current study while Z-score_Microbe_ are the results of association analysis of the gut microbiome ascertained with 16S RNA sequencing and with the Nightingale metabolites published earlier by Vojinovic et al.^21^. Metabolites were clustered to 20 groups based on the method of Li & Ji ^30^. These groups were used as random effects in the linear mixed regression. This step was performed to control for inflation in the statistic because of high correlation between the metabolites. Correlation between the metabolic profiles of the 361 gut microbial taxa and MDD was estimated as the square root of the product of the effect estimates from the two regressions described above. Significance of the correlation was tested using the Student’s T test. FDR was applied to correct for multiple testing. Next, we compared these T-Statistics (proxy association for MDD-Microbiome based on the MDD-metabolome) as obtained in the present study with the Z-scores (direct association of MDD-Microbiome) from Djawad et al.^20^.

### Mendelian Randomisation (MR)

To elucidate the causal relationships between depression and the associated metabolites, we performed bi-directional two-sample MR using the R package of *TwoSampleMR* for the inverse variance weighted MR, heterogeneity test and pleiotropy test from MR-Egger regression^31^. The default pipelines in the packages were used. In brief, for the analysis using *TwoSampleMR*, the genetic score was based on the top SNPs (P-value < 10^−6^) with linkage disequilibrium R^2^ < 0.001 within 10,000kbps clumping distance. The overlapping SNPs were used without seeking proxy SNPs as we assumed that each meaningful locus should have multiple SNPs significant and overlapped. The metabolite GWAS were obtained from the MRC IEU OpenGWAS in MRbase^31^ using all the UK Biobank participants who were profiled for Nightingale metabolites (n=118,000). For MDD we used the publicly available results of the largest GWAS by Howard et al. ^32^.

## Results

Baseline characteristics of the studied samples are provided in **Table 1**. Cases consisted of 8462 individuals with a lifetime major depression and 5403 patients with recurrent major depression and the controls included > 55420 participants. Patients are significantly younger, are more often female, smokers, have higher education and have a higher body mass index, are less physically active, consume less alcohol and have a black/mixed ethnic background compared to the controls (**Table 1**). Of the patients with history of depression, 1958 (23%) were using anti-depressants at the time blood was drawn for the metabolomic characterization. The patients were found to use more often medication related to gastric diseases, pain and addiction (**Table 1**).

**Table 1:**
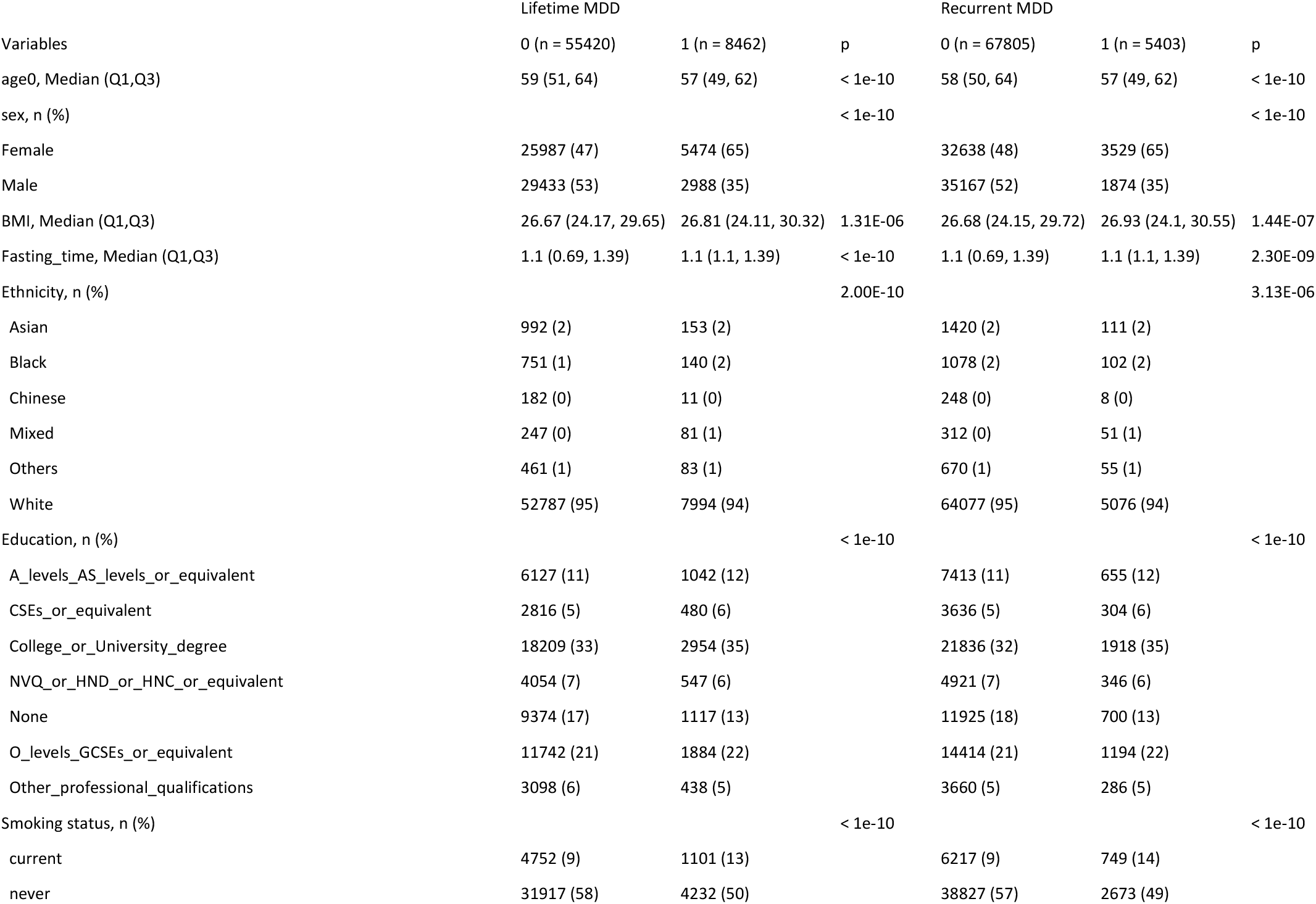
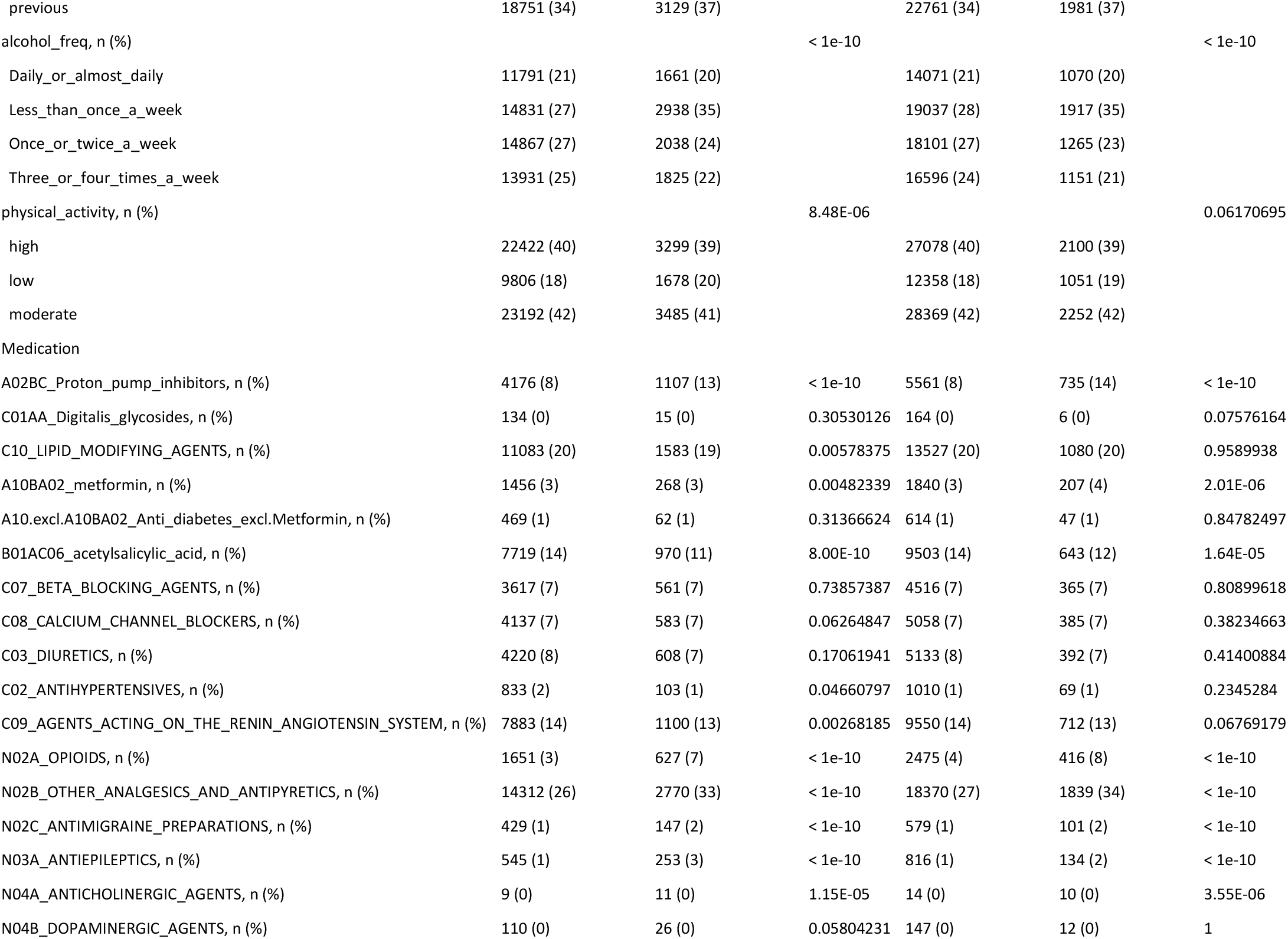
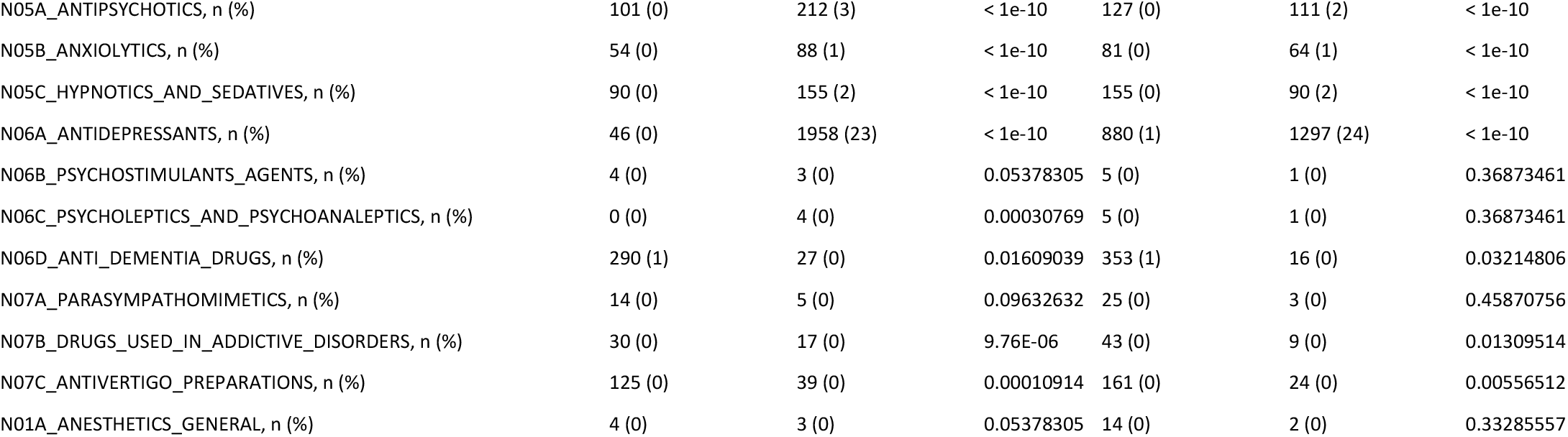
Descriptive Statistics for the patients with MDD and controls and recurrent depression

### Metabolome-wide association analysis for major depression using the Nightingale platform

Results of the analysis are shown in Figure 1 and Supplementary Figures 1-4. Adjusting for age, sex and technical covariates, 178 (71.0%) metabolites of the 249 metabolites tested were significantly (false discovery rate (FDR) < 0.05) associated with MDD in the basic model 1. When adjusting for BMI (model 2), 163 (65.5%) metabolites remained significantly associated with MDD and further adjusting for antidepressant use (model 3) yielded 132 (53.0%) metabolites significantly associated (FDR<0.05). In the full model further adjusted for lifestyle factors including physical activity, alcohol consumption, smoking, education and medication use for cardio-vascular morbidity (model 4), a total of 124 (49.8%) metabolites remained significantly associated with MDD (**Figure 1, Supplementary Table 1**). These include 27 small to extremely large VLDL particles (chylomicrons) that were increased in individuals with MDD patients, 18 medium to very large sized HDL particles all of which were decreased in MDD, except the **triglycerides** in the small and medium HDL particles, and 5 intermediate density lipoprotein (IDL) particles all of which, except the **triglyceride content in IDL**, were decreased in MDD patients. Among fatty acids, total monounsaturated fatty acids (MUFA) and its ratio to total fatty acids was significantly increased in MDD while the ratios of linoleic acid (LA), omega 6 and polyunsaturated fatty acid (PUFA) to total fatty acids were significantly decreased in MDD patients. Further, apolipoprotein A1 (ApoA1), cholesteryl esters, citrate and sphingomyelins were significantly decreased in MDD while alanine and pyruvate were significantly increased in MDD patients (**Figure 1, Supplementary Table 1**). Findings were very similar when excluding those with antidepressant use instead of adjusting for antidepressants (**Supplementary Figure 5, Supplementary Table 2**). Comparing the results of lifetime MDD with that of the recurrent depression, the metabolic profiles were found to be highly correlated (**Supplementary Figures 1-4**).

**Figure 1:**
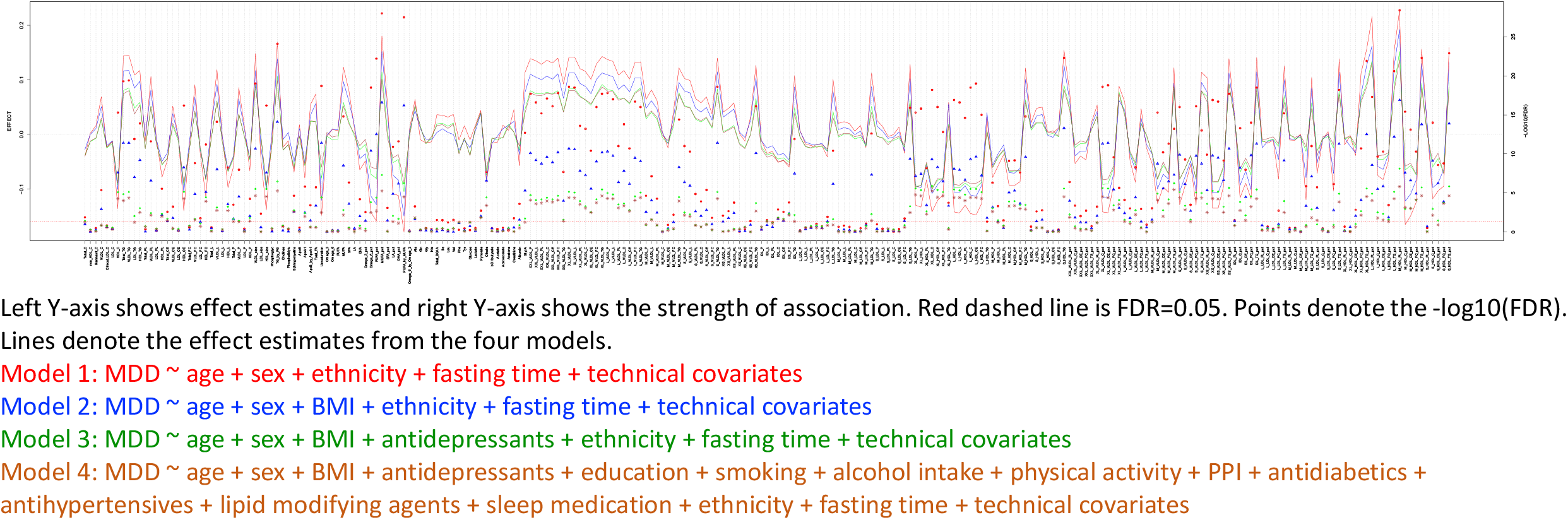
Results of metabolome-wide association analysis

Comparing our results with those of the BBMRI-NL ^16^, 113 of the metabolites that we find associated to major depression were also studied by BBMRI-NL. Supplementary Figure 6 shows that the effect direction of the identified metabolites from model 4 in our study is consistent with that seen in the previous study by the BBMRI-NL consortium (**Supplementary Figure 6, Supplementary Table 3**. In the present study, we identified 49 metabolites that were FDR significant that were not reported in the BBMRI-NL study (**Supplementary Table 4)**. These include metabolites involved in mitochondrial functioning, including alanine, citrate, pyruvate, fatty acids including PUFA%, LA% and omega6%, sphingomyelins, IDL subfractions in addition to some VLDL and HDL subfractions that were not associated earlier. The association of depression to omega 6, PUFA, citrate and pyruvate was replicated in the PReDICT study (**Supplementary Table 5**). We could not replicate the finding on alanine and no data were available on sphingomyelins, IDL or other lipid fractions, although most of these show consistency in direction of effects in the BBMRI-NL study (**Supplementary Figure 6**).

### Mendelian Randomization (MR) analysis

Results of bidirectional MR are provided in the **Figure 2** and **Supplementary Table 6**. No significant pleiotropy was observed (**Supplementary Table 7)** in the MR analysis. However, significant heterogeneity was observed for most metabolites (**Supplementary Table 8**) suggesting violation of assumptions of MR. We therefore considered the results of the weighted median method as these have shown to produce consistent results in the presence of heterogeneity^33^. Significant MR results were obtained after multiple testing correction using FDR < 0.05 (**Figure 2**) when MDD was used as the exposure and metabolites as the outcome. Changes in all metabolites except ApoA1, citrate, pyruvate, alanine, total cholesteryl ester, sphingomyelins, medium and large HDL particles, HDL cholesterol, total concentration of HDL particles, total lipids in HDL and average diameter of HDL particles, and a few VLDL subfractions **Figure 2**) appeared to be associated to the genes that are associated to depression.

**Figure 2:**
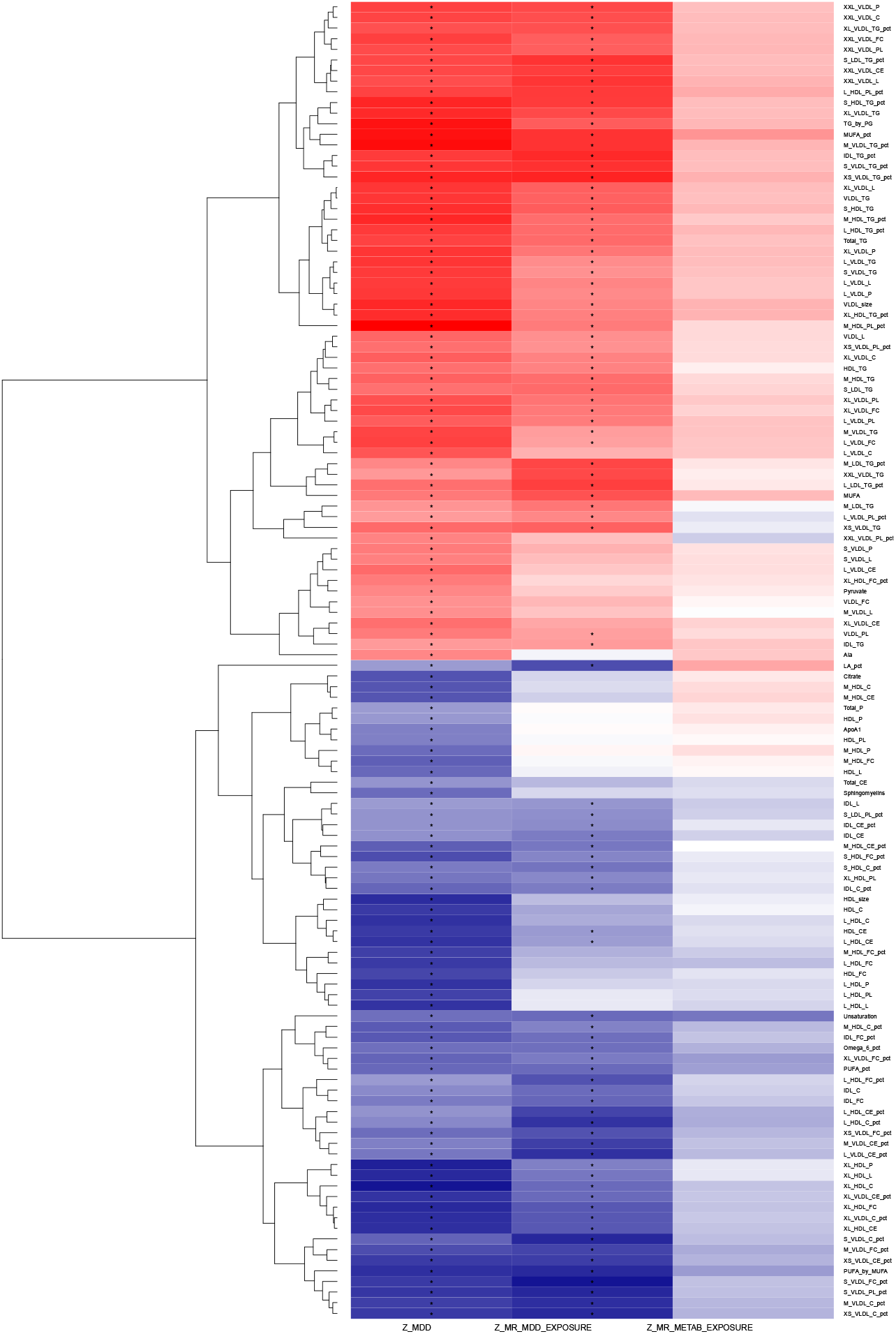
Results of bi-directional Mendelian Randomization The first column from the left shows the direction of association of 124 significantly associated metabolites with MDD (Z-scores). Blue is negative association and red is positive association. The second column shows the results of the Mendelian randomization (MR) analysis when MDD is the exposure and microbiome is the outcome. The third column depicts the results of MR when metabolites were used as exposure and MDD outcome. Black stars represent significant ones after correcting for multiple testing using FDR.

### Integration of human gut microbiome and metabolic signatures

We compared the pattern of association of MDD with the metabolites (metabolic signature of MDD) to the pattern of association of human gut microbial taxa with the metabolites (metabolic signatures of the gut microbial taxa) (**Supplementary Table 9, Figures 3 & 4)**. The association of the gut microbiota and the Nightingale metabolites is based on an independent study published earlier^21^. **Figure 3** shows all taxa whose metabolic signatures show significant positive correlation (r > 0.50 & FDR < 0.05) with that of MDD and **Figure 4** illustrates all taxa whose metabolic signatures show significant negative correlation (r < - 0.50 & FDR < 0.05) with the metabolic signature of MDD. We henceforth refer to this correlation of the metabolic signatures of MDD and gut microbial taxa as ‘proxy association’ between MDD and gut microbiome. When comparing this proxy association with the direct association of depression with the gut microbiome from a previous independent study^20^, we find a highly significant correlation (r=0.58, p-value < 10^−16^)(**Figure 5)**. This finding suggests that the Nightingale platform can be used as a proxy measure for the microbiome and that the correlation of the microbiome and MDD are mainly driven by lipoproteins, especially VLDL and HDL particles. Of note is that the bacteria which were associated with a healthy lipid profile, i.e., increased levels of HDL subfractions and decreased VLDL lipid levels, were found to be decreased in MDD (**Figure 3**). Conversely, bacteria which were associated with an unhealthy lipid profile, i.e., decreased levels of lipids in HDL particles and increased levels of lipids in VLDL, were found to be increased in patients with MDD (**Figure 4**).

**Figure 3:**
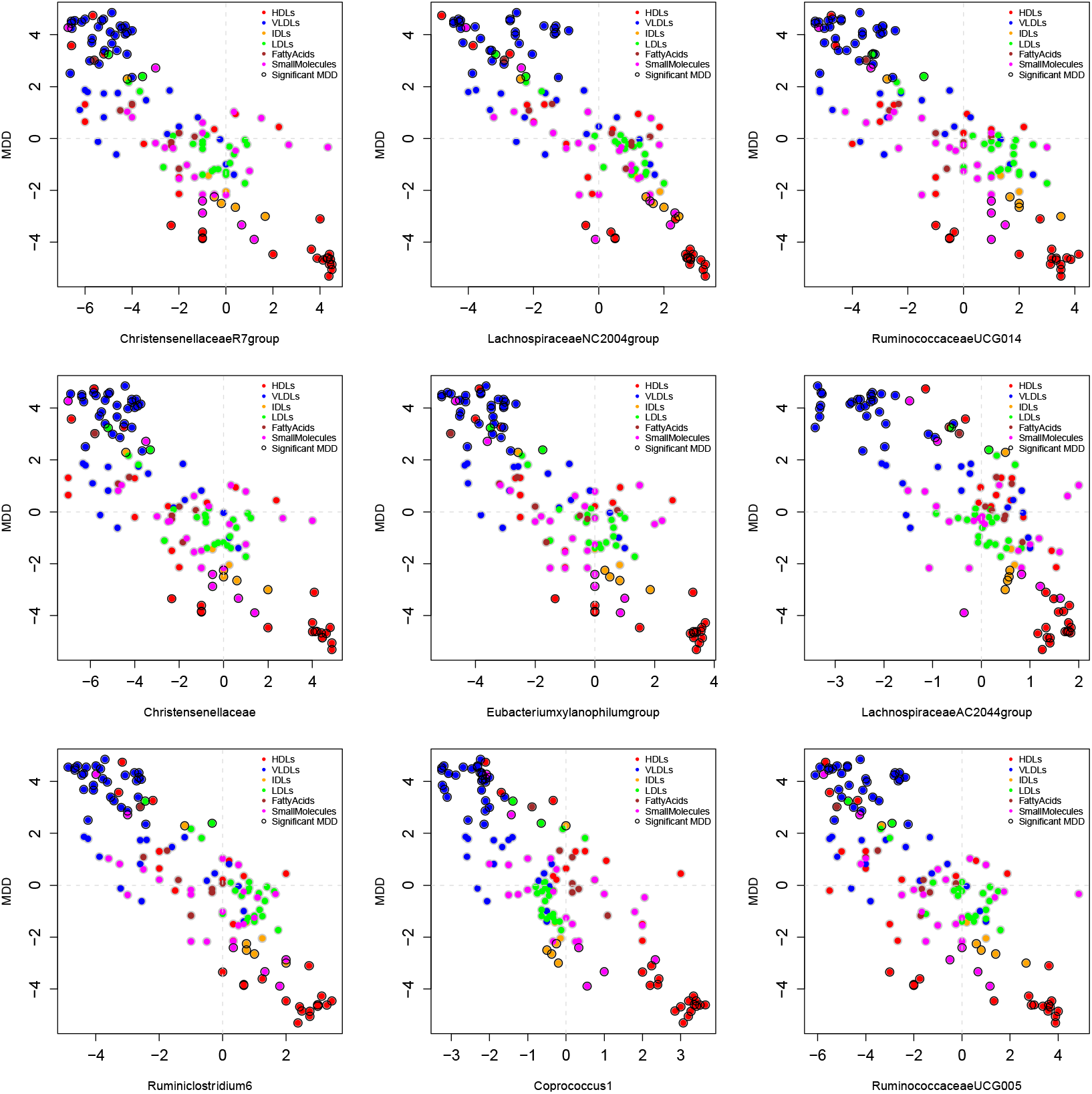
Association of metabolic profiles (z-scores) of ‘healthy’ gut microbiota and MDD A scatter plot showing the correlation between the metabolic profiles of microbial taxa that are negatively associated the metabolic profile of MDD. Each dot represents a metabolite. X-axis shows the association of the metabolite to microbial taxa and Y-axis shows the association of the metabolite to MDD. Different colours of the dots represent the class of the metabolite and the dots highlighted with black circles are the significantly associated metabolites with MDD in model 4.

**Figure 4:**
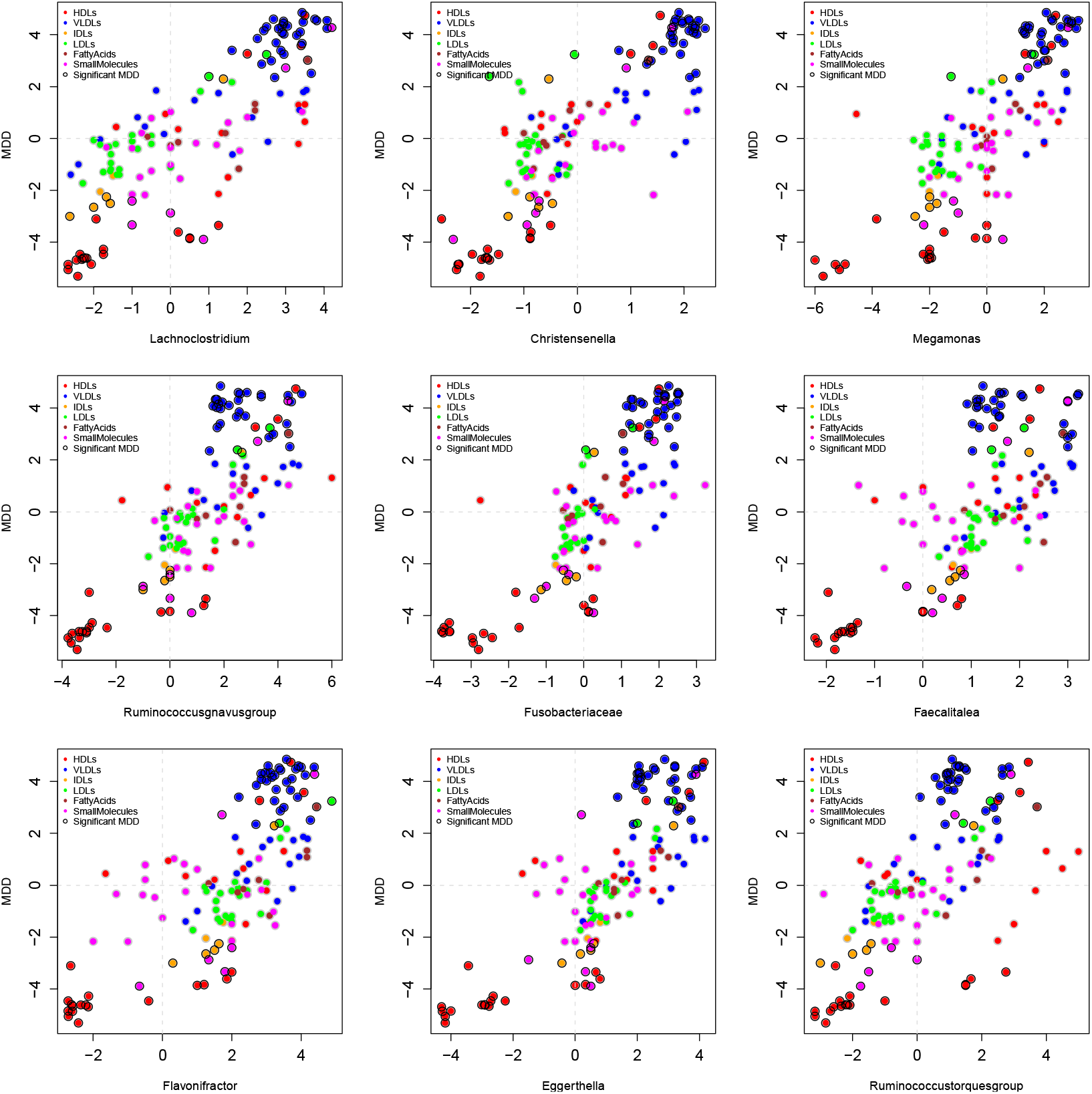
Association of metabolic profiles (z-scores) of unhealthy microbiota and MDD A scatter plot showing the correlation between the metabolic profiles of microbial taxa that are positively associated the metabolic profile of MDD. Each dot represents a metabolite. X-axis shows the association of the metabolite to microbial taxa and Y-axis shows the association of the metabolite to MDD. Different colours of the dots represent the class of the metabolite and the dots highlighted with black circles are the significantly associated metabolites with MDD in model 4.

**Figure 5:**
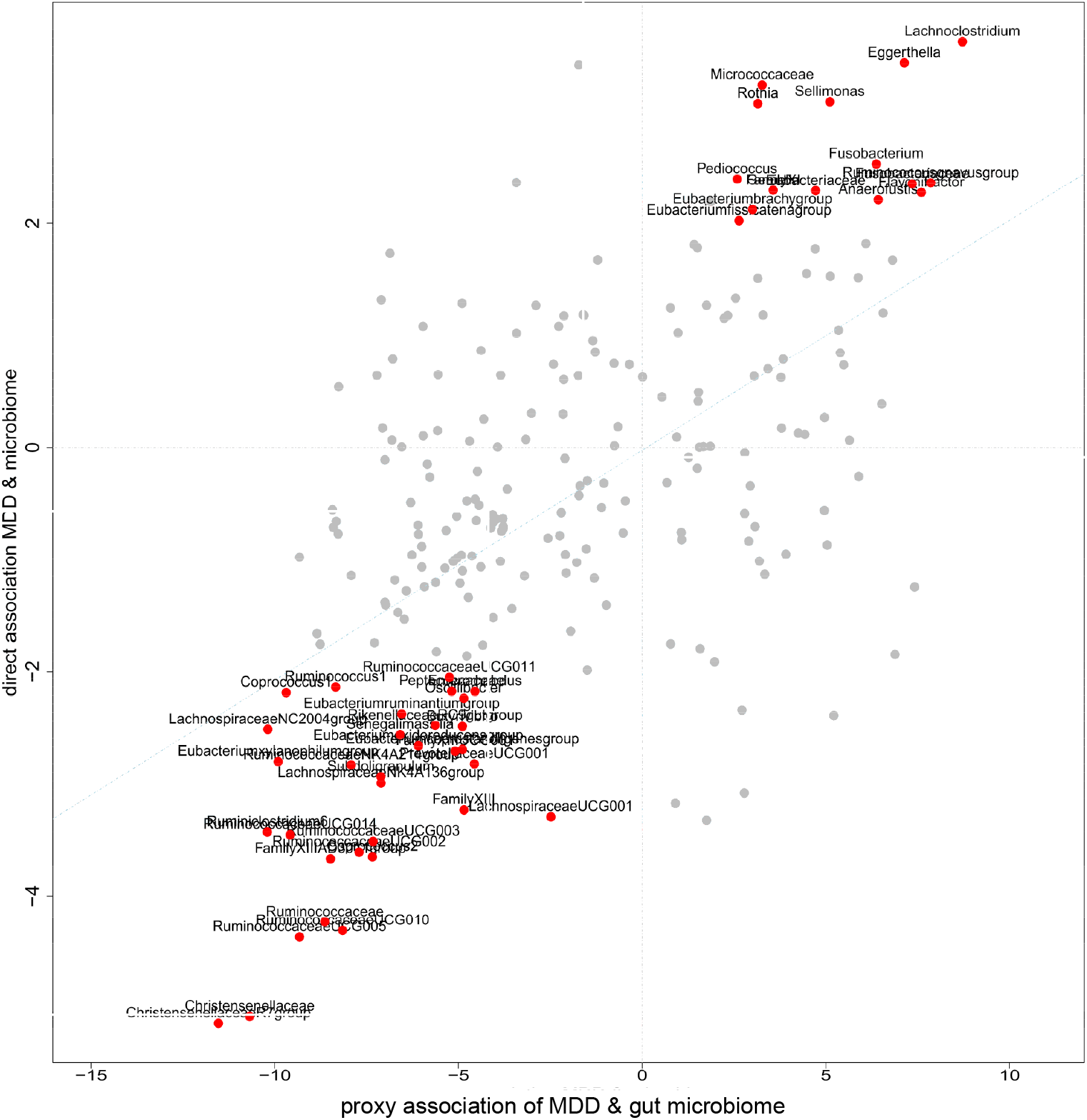
Scatter plot of direct and proxy association of microbiome with major depression. Taxa that have a p-value < 0.05 in both proxy and direct association are annotated. Each dot represents a microbial taxon. X-axis depicts the proxy association (T-scores) between microbial taxa and MDD inferred through their metabolic signatures and Y-axis depicts the direct association of microbial taxa with MDD (Z-scores) from the Rotterdam study performed in Radjabzadeh et al^20^. Taxa that are nominally significant in both proxy and direct associations are annotated.

Overall, we observed 223 bacterial taxa significantly associated with MDD using proxy association (FDR_corr_ < 0.05) (**Figure 6, Supplementary Table 9**). Figure 6 shows the complete gut microbiome profile of MDD at hierarchical levels. Family *Ruminococcaceae* (r=-0.60, FDR=2.9*10^−13^) and most of its genera were significantly negatively correlated with MDD patients. Several other families belonging to the order *Clostridiales* (*Clostridiaceae* (r=-0.59, FDR=8.8*10^−13^), *Christensenellaceae* (−0.68, FDR=1.8*10^−17^), *Peptostreptococcaceae* (r=-0.50, FDR=2.1*10^−09^), *Defluviitaleaceae* (r=-0.45, FDR=9.4*10^−08^) and *Peptococcaceae* (r=-0.41, FDR=2.3*10^−06^)) also showed significant negative correlation with MDD. Families *Lachnospiraceae* (r=0.43, FDR= 6.3*10^−07^) and *Eubacteriaceae* (r=0.38, FDR=1.3*10^−05^) were significantly positively correlated with MDD, however, some genera belonging to these families were significantly negatively correlated with MDD (**Figure 6**). Further, several families belonging to the phylum *Proteobacteria* including *Methanobacteriaceae, Rhodospirillaceae, Desulfovibrionaceae, Pasteurellaceae, Neisseriaceae* and *Oxalobacteraceae* and phylum *Bacteroidetes* including *Porphyromonadaceae, Rikenellaceae* and *Prevotellaceae* were significantly negatively correlated with MDD (**Figure 6, Supplementary Table 9**).

**Figure 6:**
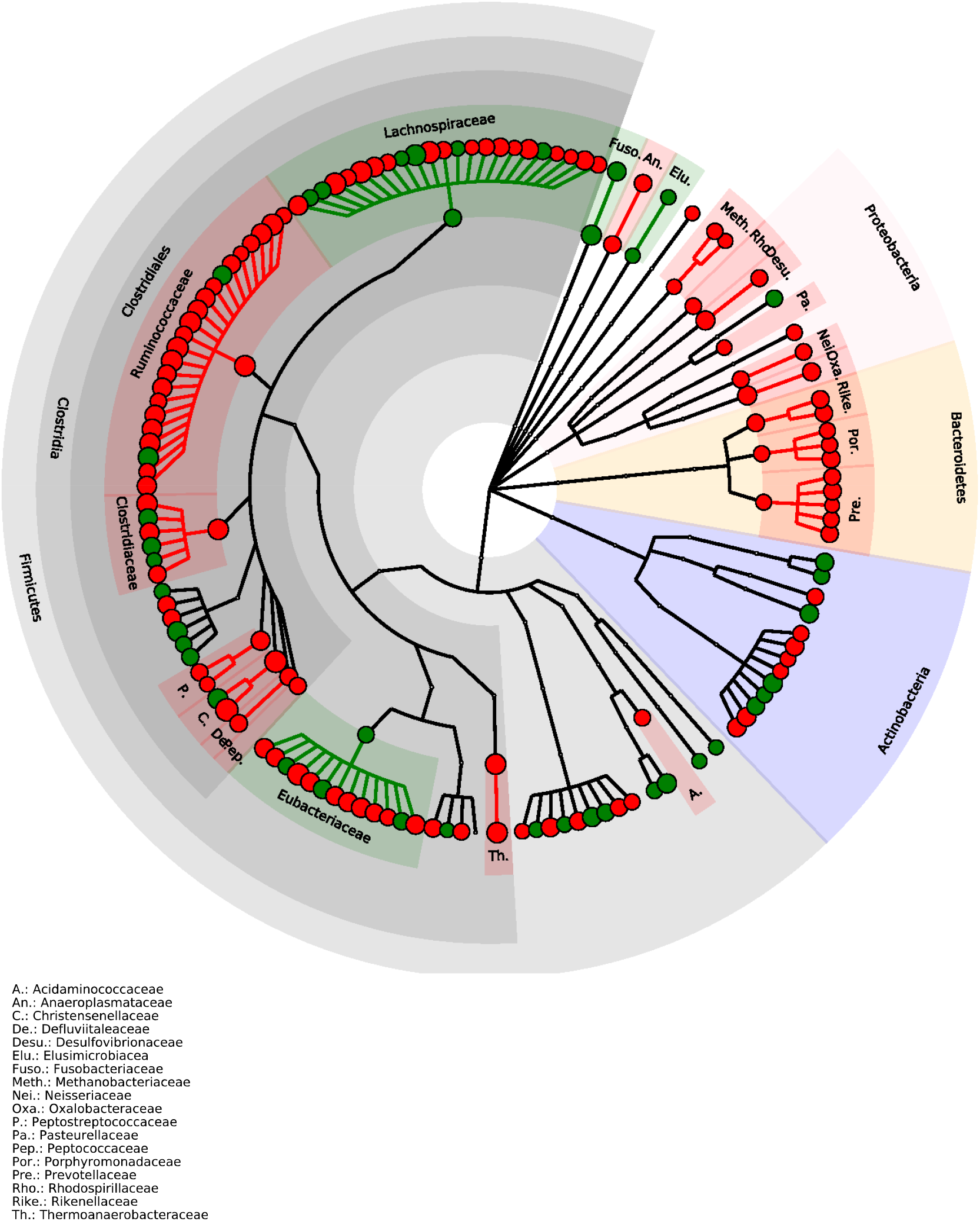
Hierarchical illustration of all healthy and pathogenic bacteria that showed significant correlation (r > 0.3 & FDR < 0.05) with MDD metabolic profile Red dots represent significant negative proxy association between microbial taxa and MDD and green dots represent significant positive proxy association between microbial taxa and MDD. The outermost layer depicts the phylum followed by, class, order, family and genus.

## Discussion

We have identified 124 metabolites associated with major depression. No differences were observed between the metabolic profiles of lifetime MDD and recurrent MDD. The findings of our study corroborate well with those of the previously published large study of the BBMRI-NL consortium^16^ in that most metabolites (90%) show consistency in the direction of association. Compared to the study of the BBMRI consortium, we find 49 metabolites that were not significantly associated to MDD earlier, including the amino acid alanine as well as citrate and pyruvate, which are two key metabolites of the energy metabolism pathway. The shift of the metabolites observed in depression was associated with bacterial taxa belonging to order *Clostridiales*, and phyla *Proteobacteria* and *Bacteroidetes*.

Our findings extend previous reports on the major changes in lipid metabolism seen in patients with MDD. These include novel associations with polyunsaturated fatty acids (PUFA%, LA%, omega6%), sphingomyelins and various lipoprotein subfractions. In this study we replicate the shift in the VLDL / HDL axis in patients with MDD^16^. VLDLs are produced by the liver and are rich in triglyceride. The removal of triglycerides from VLDL by muscle and adipose tissue results in the formation of IDL particles, which we also find significantly associated with MDD in the present study (not reported earlier in BBMRI-NL). VLDL, IDL and LDL particles are atherogenic and are found to be increased in cardio-vascular morbidities^34^. HDL, on the other hand is synthesized in the liver and the intestine^35^. Apolipoprotein A1 (ApoA1) is the major structural protein of HDL accounting for 70% of HDL protein^35^. HDL particles are responsible for reverse cholesterol transport and have anti-oxidant, anti-inflammatory, anti-thrombotic, and anti-apoptotic properties^34^. HDLs have been shown to be protective against mitochondrial dysfunction and positively associated with mitochondrial oxidative function^15^.

The finding that various VLDL particles and MUFA are increased and HDL particles and APOA1 are decreased in depression are consistent with those of previous studies^16,36^. Our study provides evidence suggesting that the microbiome is a key player in the shift in VLDL/HDL axis in MDD. Integrating metabolic profiles of MDD and the gut microbiome, we find that the shift in most metabolites, particularly the HDL and VLDL fractions are associated to several families of gut microbiota. Those belonging to the order *Clostridiales*, and phylum *Proteobacteria* and *Bacteroidetes* are decreased in MDD patients and are associated with high HDL lipid levels and low VLDL lipid levels in blood while families *Lachnospiracea* and *Eubacteriaceae* are predicted to be increased in MDD patients and are associated with low levels of HDL subfractions and high levels of VLDL subfractions. Earlier studies have shown that gut microbiome is a major determinant of the circulating lipids and has a bidirectional relationship with mitochondrial function^17,18^. Members of microbiota from order *Clostridiales* are known to provide transforming growth factor *β* enriched environment for promotion and accumulation of regulatory T cells in the gut^37^. These regulatory T cells have been found to be reduced in mood disorders^38^. It is interesting to note that we find several families belonging to the order *Clostridiales* associated with MDD in our proxy association. Our findings are in line with the previous study where *Clostridiales* were found to be the predominant microbes mediating psychiatric disorders including depression^39^.

This significant link between the microbiome and MDD may also shed light on the interpretation of the Mendelian Randomization experiments that did not yield significant evidence that any of the human genes associated to the metabolites investigated here. However, we find evidence that changes in lipoproteins and their subfractions and fatty acids associate with genes involved in MDD. We hypothesise that the metabolic change we observe is part of the disease process and that the genes that have been implicated in MDD explain the shift towards increased VLDL and decreased HDL subfractions (except medium and large HDL particles) in MDD. This may be explained by the pleiotropic effects of the genes determining lipid metabolism or by the changes in diet and physical activity that are consequences of the changes in a patient’s mood. Alternatively, we hypothesise that the change in these metabolites may be due to altered composition of the gut microbiome in depressed individuals, which may be driven by the genes that determine MDD. Such a genetically driven shift in microbiome is also seen in transgenic models for Alzheimer’s disease (AD) in which the introduction of the major genes involved in AD in mice, resulted in a shift of the microbiome^40^. Although we cannot exclude that the VLDL levels in the blood drive the gut microbiome and MDD, this mechanism is not supported by the Mendelian Randomization. Based on the Mendelian Randomization analysis, we hypothesize that MDD genes drive the gut microbiome, which determines the metabolic spectrum (VLDLs and small HDLs) in the blood^17^. The exact mechanism is to be determined in future experiments.

The second major finding is the disruption in the mitochondrial metabolism, more specifically the tricarboxylic acid (TCA)/Krebs cycle. We find significantly increased levels of pyruvate and decreased levels of citrate, which are major components of the mitochondrial Krebs cycle^41^. To our knowledge, there is no evidence in human data for an association of these metabolites to MDD. Decreased citrate levels were found in the urine of rats in chronic unpredictable mild stress depression model^42,43^. Further, increased mitochondrial activity generating citrate and reduced oxidative stress parameters has been observed in patients with bipolar depression treated with lithium compared to untreated patients^44^. In the brain, citrate contributes to the regulation of neuronal excitability chelating and controlling the availability of divalent ions such as Ca^+2^ and Mg^+2 41,45^. Extracellular citrate is also used for neurotransmitter synthesis^41,45^. In animal models of induced oxidative stress, oral administration of citrate decreased brain lipid peroxidation and inflammation, liver damage, and DNA fragmentation^46^. 90% of the total citrate in the body is localized in bones^47,48^. Osteoblast metabolic production of citrate provides the source of citrate in bone in the presence of zinc, which inhibits the oxidation of citrate in mitochondria^48,49^. Citrate is released in the plasma during bone resorption^47^. Decreased levels of blood citrate (hypocitricemia) causes loss of bone citrate and osteoporosis. The relationship of depression and osteoporosis is well established^50^, however, Mendelian randomisation study showed no causal effect of depression on osteoporosis^51^. Interestingly, administration of vitamin D increases the plasma and bone citrate concentrations by inhibiting mitochondrial citrate oxidation^47^. This implies that, if plasma citrate deficiency has a causal influence on major depression, the subgroup of individuals with MDD exhibiting low citrate levels may very well be treated with zinc and/or vitamin D supplements, both of which have shown to ameliorate symptoms of depression^52,53^.

Low citrate levels may also be a result of intestinal dysbiosis or impairment of the pyruvate dehydrogenase complex that catalyses the conversion of pyruvate into acetyl-CoA^54^. One of the questions to answer in future studies is whether acetyl-CoA links the findings on lipids and amino acids. Cholesterol synthesis initiates from acetyl coenzyme A (acetyl-CoA)^55^. Acetyl-CoA is synthesized in mitochondria from the oxidative decarboxylation of pyruvate, oxidation of fatty acids or oxidative degradation of some amino acids (e.g., phenylalanine, tyrosine, leucine, lysine, and tryptophan)^56^. Acetyl-CoA is transported out of the mitochondria after being converted into citrate^56^. Cytosolic acetyl-CoA is used in fatty acid metabolism and lipid biosynthesis ^57^. We hypothesize that citrate, which we found to be decreased in depression may be the key metabolite connecting both the lipid and energy metabolism pathways via Acetyl-CoA.

In conclusion, we have performed the largest and most comprehensive study investigating the association of NMR metabolites with major depression. We find that metabolites involved in the tricarboxylic acid (TCA) cycle are significantly altered in patients with MDD, suggesting perturbations in metabolites involved in energy metabolism in patients with MDD. Our finding that the interplay between the gut microbiome and the blood metabolome may play a key role in MDD and suggests that the gut microbiome may be a target for novel preventive and therapeutic interventions for MDD.

## Data Availability

All data belonging to the UK Biobank is publicly available.

## Acknowledgements

“Metabolomics data is provided by the Alzheimer’s disease Metabolomics Consortium (ADMC) and funded wholly or in part by the following grants and supplements thereto: NIA R01AG046171, RF1AG051550, RF1AG057452, R01AG059093, RF1AG058942, U01AG061359, U19AG063744 and FNIH: #DAOU16AMPA awarded to Dr. Kaddurah-Daouk at Duke University in partnership with a large number of academic institutions. As such, the investigators within the ADMC, not listed specifically in this publication’s author’s list, provided data along with its pre-processing and prepared it for analysis, but did not participate in analysis or writing of this manuscript. A complete listing of ADMC investigators can be found at: https://sites.duke.edu/adnimetab/team/.”

In order to accurately acknowledge data gathering and pre-processing by the ADMC, data users must include a laboratory specific acknowledgement statement in the methods section of manuscripts. These study/dataset-specific acknowledgements can be found in the Study Pages. Depending upon the length and focus of the article, it may be appropriate to include more or less of these statements. Draft: “The Nightingale datasets have been generated at Nightingale Health.”

## Conflicts of interest

“Dr. Kaddurah-Daouk in an inventor on a series of patents on use of metabolomics for the diagnosis and treatment of CNS diseases and holds equity in Metabolon Inc., Chymia LLC and PsyProtix.”Alternatively, “R.K-D. formed Chymia LLC and PsyProtix, a Duke University biotechnology spinout aiming to transform the treatment of mental health disorders.”All other authors declare no conflicts of interest.

Dr. Saykin receives support from multiple NIH grants (P30 AG010133, P30 AG072976, R01 AG019771, R01 AG057739, U01 AG024904, R01 LM013463, R01 AG068193, T32 AG071444, and U01 AG068057 and U01 AG072177). He has also received support from Avid Radiopharmaceuticals, a subsidiary of Eli Lilly (in kind contribution of PET tracer precursor); Bayer Oncology (Scientific Advisory Board); Eisai (Scientific Advisory Board); Siemens Medical Solutions USA, Inc. (Dementia Advisory Board); Springer-Nature Publishing (Editorial Office Support as Editor-in-Chief, Brain Imaging and Behavior).

M.A. and G.K. received funding (through their institutions) from the National Institutes of Health/National Institute on Aging through grants RF1AG058942, RF1AG059093, U01AG061359, U19AG063744, and R01AG069901.

M.A. and G.K. are co-inventors (through Duke University/Helmholtz Zentrum München) on patents on applications of metabolomics in diseases of the central nervous system; M.A. and G.K. hold equity in Chymia LLC and IP in PsyProtix and Atai that are exploring the potential for therapeutic applications targeting mitochondrial metabolism in treatment-resistant depression.

## Notes

### Funding Statement

Metabolomics data is provided by the Alzheimers disease Metabolomics Consortium (ADMC) and funded wholly or in part by the following grants and supplements thereto: NIA R01AG046171, RF1AG051550, RF1AG057452, R01AG059093, RF1AG058942, U01AG061359, U19AG063744 and FNIH: DAOU16AMPA awarded to Dr. Kaddurah-Daouk at Duke University in partnership with a large number of academic institutions. As such, the investigators within the ADMC, not listed specifically in this publication authors list, provided data along with its pre-processing and prepared it for analysis, but did not participate in analysis or writing of this manuscript. A complete listing of ADMC investigators can be found at: https://sites.duke.edu/adnimetab/team/.
In order to accurately acknowledge data gathering and pre-processing by the ADMC, data users must include a laboratory specific acknowledgement statement in the methods section of manuscripts. These study/dataset-specific acknowledgements can be found in the Study Pages. Depending upon the length and focus of the article, it may be appropriate to include more or less of these statements. The Nightingale datasets have been generated at Nightingale Health.

### Author Declarations

UK Biobank has approval from the Northwest Multi-centre Research Ethics Committee, the Patient Information Advisory Group, and the Community Health Index Advisory Group.

