## Supplementary Text for "Interplay of Human Metabolome and Gut Microbiome in Major Depression"

### *PReDICT Study Design and Participants*

This study examined serum samples from the Predictors of Remission in Depression to Individual and Combined Treatments (PReDICT) study; the design and clinical outcomes of PReDICT have been detailed previously (PMID: 22776534; 28335624; 30764648). Briefly, the PReDICT study aimed to identify predictors and moderators of response to 12 weeks of randomly-assigned treatment with duloxetine (30-60 mg/day), escitalopram (10-20 mg/day) or cognitive behavior therapy (CBT, 16 one-hour individual sessions). Eligible participants were adults aged 18-65 with an active major depressive episode without psychotic features as part of MDD who had never previously been treated for depression. Severity of depression at the randomization visit was assessed with the 17-item Hamilton Depression Rating Scale ($HRSD_{17}$)(PMID: 14399272). Eligibility required a $HRSD_{17}$ score ≥18 at the screening visit and ≥15 at the randomization visit, indicative of moderate-to-severe depression. Active significant suicide risk, current illicit drug use (assessed with urine drug screen) or a history of substance abuse in the three months prior to randomization, pregnancy, lactation, and uncontrolled general medical conditions were all exclusionary.

### *Metabolomic Profiling*

At the randomization visit, antecubital phlebotomy was performed without concern for time of day or fasting status to obtain the serum samples used in the current analysis. Blood samples were allowed to clot for 20 minutes, then centrifuged at 4C for 10 minutes. The serum was pipetted into Eppendorf tubes and immediately frozen at -80C until ready for metabolomic analysis.

Using targeted metabolomics protocols and profiling protocols established in previous studies (PMID: 25581415, 28437060, 19678709), citric acid and fatty acids were quantified by ultra-performance liquid chromatography triple quadrupole mass spectrometry (UPLC-TQMS) (Waters XEVO TQ-S, Milford, USA).

A targeted, liquid chromatography–electrochemical coulometric array (LCECA) metabolomics platform (PMID= 6147209) was used to assay metabolites in serum samples. This platform was used to identify and quantify 35 metabolites including pyruvate and primarily neurotransmitter-related to tryptophan, tyrosine, and tocopherol pathways.

### *Statistical Analysis*

Linear regression models were used to assess the cross-sectional association of selected metabolites with the 17-item Hamilton Rating Scale for Depression ($HRSD_{17}$) and 14-tems Hamilton Rating Scale for Anxiety (HRSA). The outcome variable was square root of the $HRSD_{17}$ and HRSA and covariates include age, sex and BMI.

### *Results*

Lower levels of pyruvate and higher levels of citric acid and EPA were associated with less-severe depression and/or anxiety. DHA along with other PUFAs were showing similar trends as EPA (i.e., higher levels of PUFAs associated or trended with less severe depression and/or anxiety).
