## Supplementary Tables for "Interplay of Human Metabolome and Gut Microbiome in Major Depression"

**Supplementary Table 1**

**Results for all models for MDD**

| | M1 | | | | | | M2 | | | | | | M3 | | | | | | M4 | | | | | |
|---|---|---|---|---|---|---|---|---|---|---|---|---|---|---|---|---|---|---|---|---|---|---|---|---|
| Metabolite | beta | se | z | pvalue | n | fdr | beta | se | z | pvalue | n | fdr | beta | se | z | pvalue | n | fdr | beta | se | z | pvalue | n | fdr |
| Total_C | -0.0399 | 0.0153 | -2.6140 | 8.95E-03 | 6810/51445 | 1.34E-02 | -0.0278 | 0.0153 | -1.8140 | 6.97E-02 | 6810/51445 | 1.02E-01 | -0.0387 | 0.0166 | -2.3286 | 1.99E-02 | 6810/51445 | 3.78E-02 | -0.0399 | 0.0185 | -2.1599 | 3.08E-02 | 6810/51445 | 5.81E-02 |
| non_HDL_C | 0.0019 | 0.0148 | 0.1274 | 8.99E-01 | 6810/51445 | 9.10E-01 | 0.0000 | 0.0148 | -0.0003 | 1.00E+00 | 6810/51445 | 1.00E+00 | -0.0133 | 0.0160 | -0.8333 | 4.05E-01 | 6810/51445 | 4.96E-01 | -0.0121 | 0.0177 | -0.6832 | 4.95E-01 | 6810/51445 | 6.23E-01 |
| Remnant_C | 0.0158 | 0.0147 | 1.0751 | 2.82E-01 | 6810/51445 | 3.36E-01 | 0.0108 | 0.0147 | 0.7320 | 4.64E-01 | 6810/51445 | 5.61E-01 | -0.0075 | 0.0159 | -0.4714 | 6.37E-01 | 6810/51445 | 7.35E-01 | -0.0063 | 0.0178 | -0.3560 | 7.22E-01 | 6810/51445 | 8.24E-01 |
| VLDL_C | 0.0690 | 0.0146 | 4.7353 | 2.19E-06 | 6810/51445 | 4.39E-06 | 0.0508 | 0.0147 | 3.4422 | 5.77E-04 | 6810/51445 | 1.19E-03 | 0.0268 | 0.0159 | 1.6818 | 9.26E-02 | 6810/51445 | 1.50E-01 | 0.0300 | 0.0172 | 1.7471 | 8.06E-02 | 6810/51445 | 1.40E-01 |
| Clinical_LDL_C | -0.0237 | 0.0148 | -1.5972 | 1.10E-01 | 6810/51445 | 1.48E-01 | -0.0206 | 0.0148 | -1.3915 | 1.64E-01 | 6810/51445 | 2.28E-01 | -0.0253 | 0.0160 | -1.5765 | 1.15E-01 | 6810/51445 | 1.78E-01 | -0.0237 | 0.0177 | -1.3381 | 1.81E-01 | 6810/51445 | 2.79E-01 |
| LDL_C | -0.0130 | 0.0120 | -1.0781 | 2.81E-01 | 6810/51445 | 3.36E-01 | -0.0119 | 0.0120 | -0.9866 | 3.24E-01 | 6810/51445 | 4.09E-01 | -0.0164 | 0.0125 | -1.3149 | 1.89E-01 | 6810/51445 | 2.71E-01 | -0.0126 | 0.0133 | -0.9500 | 3.42E-01 | 6810/51445 | 4.60E-01 |
| HDL_C | -0.1362 | 0.0164 | -8.2847 | 1.18E-16 | 6810/51445 | 5.46E-16 | -0.1012 | 0.0174 | -5.8123 | 6.16E-09 | 6810/51445 | 2.44E-08 | -0.0923 | 0.0189 | -4.8769 | 1.08E-06 | 6810/51445 | 7.89E-06 | -0.0891 | 0.0200 | -4.4624 | 8.11E-06 | 6810/51445 | 5.48E-05 |
| Total_TG | 0.1437 | 0.0151 | 9.4964 | 2.17E-21 | 6810/51445 | 4.51E-20 | 0.1164 | 0.0158 | 7.3692 | 1.72E-13 | 6810/51445 | 3.75E-12 | 0.0806 | 0.0171 | 4.7034 | 2.56E-06 | 6810/51445 | 1.45E-05 | 0.0745 | 0.0174 | 4.2728 | 1.93E-05 | 6810/51445 | 9.81E-05 |
| VLDL_TG | 0.1452 | 0.0152 | 9.5290 | 1.59E-21 | 6810/51445 | 3.59E-20 | 0.1171 | 0.0160 | 7.3364 | 2.19E-13 | 6810/51445 | 3.96E-12 | 0.0850 | 0.0173 | 4.9082 | 9.19E-07 | 6810/51445 | 7.38E-06 | 0.0801 | 0.0176 | 4.5475 | 5.43E-06 | 6810/51445 | 4.51E-05 |
| LDL_TG | 0.1084 | 0.0149 | 7.2557 | 4.00E-13 | 6810/51445 | 1.31E-12 | 0.0850 | 0.0153 | 5.5571 | 2.74E-08 | 6810/51445 | 9.23E-08 | 0.0477 | 0.0167 | 2.8622 | 4.21E-03 | 6810/51445 | 9.44E-03 | 0.0368 | 0.0170 | 2.1684 | 3.01E-02 | 6810/51445 | 5.77E-02 |
| HDL_TG | 0.1169 | 0.0148 | 7.8773 | 3.34E-15 | 6810/51445 | 1.28E-14 | 0.0974 | 0.0151 | 6.4521 | 1.10E-10 | 6810/51445 | 6.39E-10 | 0.0589 | 0.0164 | 3.6035 | 3.14E-04 | 6810/51445 | 9.09E-04 | 0.0540 | 0.0166 | 3.2633 | 1.10E-03 | 6810/51445 | 3.16E-03 |
| Total_PL | -0.0126 | 0.0156 | -0.8071 | 4.20E-01 | 6810/51445 | 4.75E-01 | -0.0036 | 0.0156 | -0.2281 | 8.20E-01 | 6810/51445 | 8.61E-01 | -0.0259 | 0.0169 | -1.5315 | 1.26E-01 | 6810/51445 | 1.93E-01 | -0.0262 | 0.0182 | -1.4370 | 1.51E-01 | 6810/51445 | 2.43E-01 |
| VLDL_PL | 0.1061 | 0.0148 | 7.1681 | 7.60E-13 | 6810/51445 | 2.43E-12 | 0.0814 | 0.0152 | 5.3594 | 8.35E-08 | 6810/51445 | 2.67E-07 | 0.0519 | 0.0164 | 3.1580 | 1.59E-03 | 6810/51445 | 3.92E-03 | 0.0515 | 0.0172 | 2.9980 | 2.72E-03 | 6810/51445 | 7.00E-03 |
| LDL_PL | -0.0063 | 0.0148 | -0.4300 | 6.67E-01 | 6810/51445 | 7.05E-01 | -0.0086 | 0.0147 | -0.5809 | 5.61E-01 | 6810/51445 | 6.44E-01 | -0.0169 | 0.0160 | -1.0572 | 2.90E-01 | 6810/51445 | 3.88E-01 | -0.0168 | 0.0175 | -0.9614 | 3.36E-01 | 6810/51445 | 4.55E-01 |
| HDL_PL | -0.0785 | 0.0163 | -4.8166 | 1.46E-06 | 6810/51445 | 2.98E-06 | -0.0492 | 0.0168 | -2.9368 | 3.32E-03 | 6810/51445 | 6.21E-03 | -0.0563 | 0.0182 | -3.0915 | 1.99E-03 | 6810/51445 | 4.81E-03 | -0.0549 | 0.0192 | -2.8594 | 4.25E-03 | 6810/51445 | 1.04E-02 |
| Total_CE | -0.0481 | 0.0153 | -3.1400 | 1.69E-03 | 6810/51445 | 2.66E-03 | -0.0340 | 0.0154 | -2.2062 | 2.74E-02 | 6810/51445 | 4.26E-02 | -0.0433 | 0.0167 | -2.5893 | 9.62E-03 | 6810/51445 | 2.05E-02 | -0.0447 | 0.0185 | -2.4087 | 1.60E-02 | 6810/51445 | 3.47E-02 |
| VLDL_CE | 0.0520 | 0.0145 | 3.5809 | 3.42E-04 | 6810/51445 | 5.92E-04 | 0.0378 | 0.0146 | 2.5873 | 9.67E-03 | 6810/51445 | 1.67E-02 | 0.0164 | 0.0158 | 1.0396 | 2.99E-01 | 6810/51445 | 3.95E-01 | 0.0204 | 0.0173 | 1.1804 | 2.38E-01 | 6810/51445 | 3.42E-01 |
| LDL_CE | -0.0059 | 0.0123 | -0.4792 | 6.32E-01 | 6810/51445 | 6.75E-01 | -0.0065 | 0.0122 | -0.5368 | 5.91E-01 | 6810/51445 | 6.63E-01 | -0.0127 | 0.0123 | -1.0334 | 3.01E-01 | 6810/51445 | 3.95E-01 | -0.0089 | 0.0131 | -0.6816 | 4.95E-01 | 6810/51445 | 6.23E-01 |
| HDL_CE | -0.1403 | 0.0164 | -8.5583 | 1.15E-17 | 6810/51445 | 6.96E-17 | -0.1057 | 0.0174 | -6.0773 | 1.22E-09 | 6810/51445 | 5.43E-09 | -0.0933 | 0.0189 | -4.9338 | 8.06E-07 | 6810/51445 | 7.28E-06 | -0.0892 | 0.0199 | -4.4766 | 7.59E-06 | 6810/51445 | 5.40E-05 |
| Total_FC | -0.0190 | 0.0151 | -1.2593 | 2.08E-01 | 6810/51445 | 2.63E-01 | -0.0123 | 0.0151 | -0.8150 | 4.15E-01 | 6810/51445 | 5.09E-01 | -0.0269 | 0.0164 | -1.6419 | 1.01E-01 | 6810/51445 | 1.61E-01 | -0.0272 | 0.0182 | -1.4961 | 1.35E-01 | 6810/51445 | 2.22E-01 |
| VLDL_FC | 0.0909 | 0.0147 | 6.1779 | 6.50E-10 | 6810/51445 | 1.60E-09 | 0.0682 | 0.0150 | 4.5401 | 5.62E-06 | 6810/51445 | 1.37E-05 | 0.0414 | 0.0162 | 2.5463 | 1.09E-02 | 6810/51445 | 2.24E-02 | 0.0428 | 0.0172 | 2.4925 | 1.27E-02 | 6810/51445 | 2.85E-02 |
| LDL_FC | -0.0354 | 0.0141 | -2.5056 | 1.22E-02 | 6810/51445 | 1.80E-02 | -0.0283 | 0.0133 | -2.1248 | 3.36E-02 | 6810/51445 | 5.10E-02 | -0.0269 | 0.0136 | -1.9833 | 4.73E-02 | 6810/51445 | 8.42E-02 | -0.0231 | 0.0145 | -1.5980 | 1.10E-01 | 6810/51445 | 1.86E-01 |
| HDL_FC | -0.1157 | 0.0164 | -7.0348 | 2.00E-12 | 6810/51445 | 6.14E-12 | -0.0807 | 0.0173 | -4.6751 | 2.94E-06 | 6810/51445 | 7.47E-06 | -0.0841 | 0.0188 | -4.4774 | 7.56E-06 | 6810/51445 | 3.62E-05 | -0.0835 | 0.0198 | -4.2133 | 2.52E-05 | 6810/51445 | 1.21E-04 |
| Total_L | 0.0190 | 0.0151 | 1.2569 | 2.09E-01 | 6810/51445 | 2.63E-01 | 0.0166 | 0.0151 | 1.0990 | 2.72E-01 | 6810/51445 | 3.51E-01 | -0.0074 | 0.0164 | -0.4504 | 6.52E-01 | 6810/51445 | 7.44E-01 | -0.0067 | 0.0176 | -0.3825 | 7.02E-01 | 6810/51445 | 8.13E-01 |
| VLDL_L | 0.1186 | 0.0149 | 7.9365 | 2.08E-15 | 6810/51445 | 8.35E-15 | 0.0922 | 0.0154 | 5.9725 | 2.34E-09 | 6810/51445 | 9.86E-09 | 0.0614 | 0.0167 | 3.6681 | 2.44E-04 | 6810/51445 | 7.33E-04 | 0.0600 | 0.0173 | 3.4629 | 5.34E-04 | 6810/51445 | 1.75E-03 |
| LDL_L | -0.0033 | 0.0148 | -0.2199 | 8.26E-01 | 6810/51445 | 8.46E-01 | -0.0046 | 0.0148 | -0.3123 | 7.55E-01 | 6810/51445 | 8.03E-01 | -0.0149 | 0.0160 | -0.9287 | 3.53E-01 | 6810/51445 | 4.46E-01 | -0.0140 | 0.0175 | -0.8005 | 4.23E-01 | 6810/51445 | 5.46E-01 |
| HDL_L | -0.0976 | 0.0163 | -5.9719 | 2.35E-09 | 6810/51445 | 5.56E-09 | -0.0652 | 0.0170 | -3.8385 | 1.24E-04 | 6810/51445 | 2.73E-04 | -0.0688 | 0.0185 | -3.7285 | 1.93E-04 | 6810/51445 | 5.92E-04 | -0.0669 | 0.0195 | -3.4353 | 5.92E-04 | 6810/51445 | 1.91E-03 |
| Total_P | -0.0595 | 0.0159 | -3.7468 | 1.79E-04 | 6810/51445 | 3.25E-04 | -0.0401 | 0.0161 | -2.4936 | 1.26E-02 | 6810/51445 | 2.14E-02 | -0.0449 | 0.0175 | -2.5699 | 1.02E-02 | 6810/51445 | 2.11E-02 | -0.0418 | 0.0186 | -2.2516 | 2.43E-02 | 6810/51445 | 4.89E-02 |
| VLDL_P | 0.0859 | 0.0147 | 5.8594 | 4.65E-09 | 6810/51445 | 1.08E-08 | 0.0654 | 0.0149 | 4.3835 | 1.17E-05 | 6810/51445 | 2.82E-05 | 0.0377 | 0.0161 | 2.3339 | 1.96E-02 | 6810/51445 | 3.75E-02 | 0.0377 | 0.0170 | 2.2149 | 2.68E-02 | 6810/51445 | 5.29E-02 |
| LDL_P | 0.0138 | 0.0147 | 0.9419 | 3.46E-01 | 6810/51445 | 4.01E-01 | 0.0083 | 0.0147 | 0.5662 | 5.71E-01 | 6810/51445 | 6.47E-01 | -0.0048 | 0.0159 | -0.3006 | 7.64E-01 | 6810/51445 | 8.26E-01 | -0.0034 | 0.0175 | -0.1938 | 8.46E-01 | 6810/51445 | 9.08E-01 |
| HDL_P | -0.0665 | 0.0159 | -4.1729 | 3.01E-05 | 6810/51445 | 5.81E-05 | -0.0451 | 0.0162 | -2.7890 | 5.29E-03 | 6810/51445 | 9.47E-03 | -0.0470 | 0.0176 | -2.6762 | 7.45E-03 | 6810/51445 | 1.63E-02 | -0.0437 | 0.0186 | -2.3435 | 1.91E-02 | 6810/51445 | 4.03E-02 |
| VLDL_size | 0.1478 | 0.0157 | 9.4029 | 5.31E-21 | 6810/51445 | 9.44E-20 | 0.1170 | 0.0166 | 7.0486 | 1.81E-12 | 6810/51445 | 2.14E-11 | 0.0938 | 0.0180 | 5.1974 | 2.02E-07 | 6810/51445 | 3.12E-06 | 0.0898 | 0.0183 | 4.9015 | 9.51E-07 | 6810/51445 | 2.37E-05 |

| | | | | | | | | | | | | | | | | | | | | | | | | |
|---|---|---|---|---|---|---|---|---|---|---|---|---|---|---|---|---|---|---|---|---|---|---|---|---|
| LDL_size | -0.0453 | 0.0153 | -2.9646 | 3.03E-03 | 6810/51445 | 4.69E-03 | -0.0206 | 0.0156 | -1.3232 | 1.86E-01 | 6810/51445 | 2.53E-01 | -0.0026 | 0.0170 | -0.1549 | 8.77E-01 | 6810/51445 | 9.02E-01 | -0.0019 | 0.0173 | -0.1084 | 9.14E-01 | 6810/51445 | 9.40E-01 |
| HDL_size | -0.1389 | 0.0162 | -8.5716 | 1.02E-17 | 6810/51445 | 6.35E-17 | -0.1015 | 0.0175 | -5.8065 | 6.38E-09 | 6810/51445 | 2.48E-08 | -0.0985 | 0.0190 | -5.1851 | 2.16E-07 | 6810/51445 | 3.12E-06 | -0.0931 | 0.0195 | -4.7770 | 1.78E-06 | 6810/51445 | 2.95E-05 |
| Phosphoglyc | -0.0063 | 0.0158 | -0.3980 | 6.91E-01 | 6806/51398 | 7.26E-01 | 0.0030 | 0.0158 | 0.1886 | 8.50E-01 | 6806/51398 | 8.78E-01 | -0.0201 | 0.0171 | -1.1708 | 2.42E-01 | 6806/51398 | 3.38E-01 | -0.0192 | 0.0181 | -1.0574 | 2.90E-01 | 6806/51398 | 4.04E-01 |
| TG_by_PG | 0.1683 | 0.0158 | 10.6864 | 1.18E-26 | 6806/51398 | 7.34E-25 | 0.1389 | 0.0168 | 8.2680 | 1.36E-16 | 6806/51398 | 8.48E-15 | 0.1071 | 0.0182 | 5.8759 | 4.20E-09 | 6806/51398 | 3.49E-07 | 0.0995 | 0.0186 | 5.3420 | 9.19E-08 | 6806/51398 | 5.48E-06 |
| Cholines | -0.0232 | 0.0158 | -1.4701 | 1.42E-01 | 6806/51398 | 1.86E-01 | -0.0092 | 0.0159 | -0.5762 | 5.64E-01 | 6806/51398 | 6.45E-01 | -0.0293 | 0.0173 | -1.6991 | 8.93E-02 | 6806/51398 | 1.46E-01 | -0.0285 | 0.0184 | -1.5484 | 1.22E-01 | 6806/51398 | 2.04E-01 |
| Phosphatidylc | -0.0118 | 0.0158 | -0.7464 | 4.55E-01 | 6806/51398 | 5.08E-01 | 0.0033 | 0.0159 | 0.2100 | 8.34E-01 | 6806/51398 | 8.72E-01 | -0.0189 | 0.0173 | -1.0911 | 2.75E-01 | 6806/51398 | 3.72E-01 | -0.0188 | 0.0184 | -1.0253 | 3.05E-01 | 6806/51398 | 4.18E-01 |
| Sphingomyelins | -0.0694 | 0.0158 | -4.3827 | 1.17E-05 | 6806/51398 | 2.31E-05 | -0.0516 | 0.0160 | -3.2251 | 1.26E-03 | 6806/51398 | 2.49E-03 | -0.0592 | 0.0174 | -3.4092 | 6.51E-04 | 6806/51398 | 1.73E-03 | -0.0629 | 0.0189 | -3.3342 | 8.55E-04 | 6806/51398 | 2.60E-03 |
| ApoB | 0.0177 | 0.0147 | 1.2048 | 2.28E-01 | 6810/51445 | 2.84E-01 | 0.0109 | 0.0147 | 0.7407 | 4.59E-01 | 6810/51445 | 5.57E-01 | -0.0039 | 0.0159 | -0.2476 | 8.04E-01 | 6810/51445 | 8.52E-01 | -0.0031 | 0.0175 | -0.1775 | 8.59E-01 | 6810/51445 | 9.18E-01 |
| ApoA1 | -0.0800 | 0.0162 | -4.9490 | 7.46E-07 | 6810/51445 | 1.57E-06 | -0.0520 | 0.0166 | -3.1343 | 1.72E-03 | 6810/51445 | 3.32E-03 | -0.0571 | 0.0180 | -3.1668 | 1.54E-03 | 6810/51445 | 3.84E-03 | -0.0548 | 0.0191 | -2.8719 | 4.08E-03 | 6810/51445 | 1.02E-02 |
| ApoB_by_ApoA1 | 0.0549 | 0.0148 | 3.7076 | 2.09E-04 | 6810/51445 | 3.72E-04 | 0.0348 | 0.0150 | 2.3173 | 2.05E-02 | 6810/51445 | 3.36E-02 | 0.0239 | 0.0163 | 1.4686 | 1.42E-01 | 6810/51445 | 2.14E-01 | 0.0250 | 0.0177 | 1.4149 | 1.57E-01 | 6810/51445 | 2.49E-01 |
| Total_FA | 0.0741 | 0.0151 | 4.9024 | 9.47E-07 | 6806/51398 | 1.96E-06 | 0.0568 | 0.0153 | 3.7197 | 1.99E-04 | 6806/51398 | 4.32E-04 | 0.0237 | 0.0166 | 1.4326 | 1.52E-01 | 6806/51398 | 2.25E-01 | 0.0229 | 0.0172 | 1.3333 | 1.82E-01 | 6806/51398 | 2.79E-01 |
| Unsaturation | -0.1485 | 0.0159 | -9.3146 | 1.22E-20 | 6806/51398 | 1.90E-19 | -0.1211 | 0.0165 | -7.3236 | 2.41E-13 | 6806/51398 | 4.01E-12 | -0.0803 | 0.0180 | -4.4529 | 8.47E-06 | 6806/51398 | 3.91E-05 | -0.0599 | 0.0184 | -3.2611 | 1.11E-03 | 6806/51398 | 3.16E-03 |
| Omega_3 | -0.0037 | 0.0153 | -0.2383 | 8.12E-01 | 6806/51398 | 8.35E-01 | -0.0016 | 0.0154 | -0.1059 | 9.16E-01 | 6806/51398 | 9.32E-01 | -0.0033 | 0.0167 | -0.1994 | 8.42E-01 | 6806/51398 | 8.77E-01 | 0.0034 | 0.0170 | 0.2029 | 8.39E-01 | 6806/51398 | 9.05E-01 |
| Omega_6 | 0.0086 | 0.0152 | 0.5704 | 5.68E-01 | 6806/51398 | 6.18E-01 | 0.0092 | 0.0152 | 0.6062 | 5.44E-01 | 6806/51398 | 6.28E-01 | -0.0099 | 0.0165 | -0.6016 | 5.47E-01 | 6806/51398 | 6.55E-01 | -0.0048 | 0.0174 | -0.2759 | 7.83E-01 | 6806/51398 | 8.74E-01 |
| PUFA | 0.0071 | 0.0152 | 0.4689 | 6.39E-01 | 6806/51398 | 6.80E-01 | 0.0084 | 0.0152 | 0.5543 | 5.79E-01 | 6806/51398 | 6.53E-01 | -0.0085 | 0.0165 | -0.5160 | 6.06E-01 | 6806/51398 | 7.08E-01 | -0.0025 | 0.0172 | -0.1451 | 8.85E-01 | 6806/51398 | 9.31E-01 |
| MUFA | 0.1232 | 0.0151 | 8.1376 | 4.03E-16 | 6806/51398 | 1.70E-15 | 0.0968 | 0.0157 | 6.1808 | 6.38E-10 | 6806/51398 | 3.00E-09 | 0.0585 | 0.0170 | 3.4373 | 5.88E-04 | 6806/51398 | 1.63E-03 | 0.0525 | 0.0174 | 3.0200 | 2.53E-03 | 6806/51398 | 6.77E-03 |
| SFA | 0.0790 | 0.0152 | 5.1883 | 2.12E-07 | 6806/51398 | 4.64E-07 | 0.0595 | 0.0154 | 3.8617 | 1.13E-04 | 6806/51398 | 2.50E-04 | 0.0221 | 0.0168 | 1.3194 | 1.87E-01 | 6806/51398 | 2.71E-01 | 0.0188 | 0.0174 | 1.0805 | 2.80E-01 | 6806/51398 | 3.92E-01 |
| LA | 0.0115 | 0.0151 | 0.7629 | 4.46E-01 | 6806/51398 | 5.02E-01 | 0.0164 | 0.0151 | 1.0835 | 2.79E-01 | 6806/51398 | 3.58E-01 | -0.0035 | 0.0164 | -0.2134 | 8.31E-01 | 6806/51398 | 8.73E-01 | 0.0012 | 0.0175 | 0.0660 | 9.47E-01 | 6806/51398 | 9.60E-01 |
| DHA | -0.0652 | 0.0157 | -4.1492 | 3.34E-05 | 6806/51398 | 6.39E-05 | -0.0445 | 0.0159 | -2.7949 | 5.19E-03 | 6806/51398 | 9.44E-03 | -0.0282 | 0.0173 | -1.6300 | 1.03E-01 | 6806/51398 | 1.64E-01 | -0.0208 | 0.0178 | -1.1705 | 2.42E-01 | 6806/51398 | 3.46E-01 |
| Omega_3_pct | -0.0482 | 0.0155 | -3.1130 | 1.85E-03 | 6806/51398 | 2.88E-03 | -0.0354 | 0.0156 | -2.2693 | 2.32E-02 | 6806/51398 | 3.73E-02 | -0.0180 | 0.0169 | -1.0652 | 2.87E-01 | 6806/51398 | 3.86E-01 | -0.0088 | 0.0173 | -0.5084 | 6.11E-01 | 6806/51398 | 7.39E-01 |
| Omega_6_pct | -0.1425 | 0.0154 | -9.2321 | 2.65E-20 | 6806/51398 | 3.00E-19 | -0.1131 | 0.0162 | -6.9642 | 3.30E-12 | 6806/51398 | 3.58E-11 | -0.0730 | 0.0178 | -4.0970 | 4.19E-05 | 6806/51398 | 1.51E-04 | -0.0595 | 0.0182 | -3.2696 | 1.08E-03 | 6806/51398 | 3.15E-03 |
| PUFA_pct | -0.1589 | 0.0156 | -10.1990 | 2.00E-24 | 6806/51398 | 6.23E-23 | -0.1287 | 0.0166 | -7.7452 | 9.54E-15 | 6806/51398 | 2.97E-13 | -0.0810 | 0.0183 | -4.4307 | 9.39E-06 | 6806/51398 | 4.18E-05 | -0.0637 | 0.0187 | -3.4085 | 6.53E-04 | 6806/51398 | 2.08E-03 |
| MUFA_pct | 0.1800 | 0.0156 | 11.5487 | 7.49E-31 | 6806/51398 | 9.33E-29 | 0.1519 | 0.0168 | 9.0245 | 1.80E-19 | 6806/51398 | 2.25E-17 | 0.1144 | 0.0183 | 6.2502 | 4.10E-10 | 6806/51398 | 5.10E-08 | 0.1010 | 0.0187 | 5.3906 | 7.02E-08 | 6806/51398 | 5.48E-06 |
| SFA_pct | 0.0536 | 0.0154 | 3.4863 | 4.90E-04 | 6806/51398 | 8.35E-04 | 0.0359 | 0.0156 | 2.3091 | 2.09E-02 | 6806/51398 | 3.41E-02 | 0.0036 | 0.0169 | 0.2145 | 8.30E-01 | 6806/51398 | 8.73E-01 | -0.0069 | 0.0174 | -0.3980 | 6.91E-01 | 6806/51398 | 8.04E-01 |
| LA_pct | -0.1084 | 0.0156 | -6.9309 | 4.18E-12 | 6806/51398 | 1.22E-11 | -0.0748 | 0.0165 | -4.5449 | 5.50E-06 | 6806/51398 | 1.36E-05 | -0.0519 | 0.0180 | -2.8915 | 3.83E-03 | 6806/51398 | 8.68E-03 | -0.0428 | 0.0190 | -2.2577 | 2.40E-02 | 6806/51398 | 4.89E-02 |
| DHA_pct | -0.1123 | 0.0157 | -7.1593 | 8.11E-13 | 6806/51398 | 2.56E-12 | -0.0841 | 0.0162 | -5.1781 | 2.24E-07 | 6806/51398 | 6.57E-07 | -0.0457 | 0.0177 | -2.5772 | 9.96E-03 | 6806/51398 | 2.10E-02 | -0.0371 | 0.0182 | -2.0332 | 4.20E-02 | 6806/51398 | 7.70E-02 |
| PUFA_by_MUFA | -0.1785 | 0.0156 | -11.4096 | 3.75E-30 | 6806/51398 | 3.11E-28 | -0.1497 | 0.0169 | -8.8603 | 7.98E-19 | 6806/51398 | 6.62E-17 | -0.1053 | 0.0185 | -5.7017 | 1.19E-08 | 6806/51398 | 5.91E-07 | -0.0891 | 0.0189 | -4.7178 | 2.38E-06 | 6806/51398 | 3.30E-05 |
| Omega_6_by_Omega_3 | 0.0077 | 0.0154 | 0.5023 | 6.15E-01 | 6806/51398 | 6.63E-01 | 0.0057 | 0.0154 | 0.3692 | 7.12E-01 | 6806/51398 | 7.71E-01 | -0.0004 | 0.0167 | -0.0242 | 9.81E-01 | 6806/51398 | 9.86E-01 | -0.0058 | 0.0171 | -0.3391 | 7.35E-01 | 6806/51398 | 8.28E-01 |
| Ala | 0.0640 | 0.0177 | 3.6078 | 3.09E-04 | 6810/51443 | 5.38E-04 | 0.0441 | 0.0179 | 2.4557 | 1.41E-02 | 6810/51443 | 2.37E-02 | 0.0517 | 0.0194 | 2.6590 | 7.84E-03 | 6810/51443 | 1.70E-02 | 0.0542 | 0.0199 | 2.7183 | 6.56E-03 | 6810/51443 | 1.54E-02 |
| Gln | -0.0099 | 0.0152 | -0.6531 | 5.14E-01 | 6790/51308 | 5.64E-01 | 0.0097 | 0.0154 | 0.6283 | 5.30E-01 | 6790/51308 | 6.16E-01 | 0.0147 | 0.0167 | 0.8789 | 3.79E-01 | 6790/51308 | 4.72E-01 | 0.0133 | 0.0169 | 0.7842 | 4.33E-01 | 6790/51308 | 5.56E-01 |
| Gly | -0.0152 | 0.0140 | -1.0808 | 2.80E-01 | 6805/51396 | 3.36E-01 | -0.0081 | 0.0143 | -0.5680 | 5.70E-01 | 6805/51396 | 6.47E-01 | -0.0171 | 0.0150 | -1.1354 | 2.56E-01 | 6805/51396 | 3.51E-01 | -0.0169 | 0.0152 | -1.1140 | 2.65E-01 | 6805/51396 | 3.77E-01 |
| His | -0.0140 | 0.0153 | -0.9168 | 3.59E-01 | 6802/51346 | 4.14E-01 | -0.0105 | 0.0153 | -0.6884 | 4.91E-01 | 6802/51346 | 5.88E-01 | -0.0071 | 0.0166 | -0.4278 | 6.69E-01 | 6802/51346 | 7.57E-01 | -0.0041 | 0.0168 | -0.2470 | 8.05E-01 | 6802/51346 | 8.95E-01 |
| Total_BCAA | 0.0362 | 0.0156 | 2.3146 | 2.06E-02 | 6809/51433 | 2.99E-02 | 0.0032 | 0.0162 | 0.2002 | 8.41E-01 | 6809/51433 | 8.76E-01 | 0.0194 | 0.0176 | 1.0998 | 2.71E-01 | 6809/51433 | 3.69E-01 | 0.0165 | 0.0178 | 0.9231 | 3.56E-01 | 6809/51433 | 4.74E-01 |
| Ile | 0.0348 | 0.0152 | 2.2975 | 2.16E-02 | 6810/51444 | 3.09E-02 | 0.0101 | 0.0155 | 0.6546 | 5.13E-01 | 6810/51444 | 5.99E-01 | 0.0202 | 0.0169 | 1.1957 | 2.32E-01 | 6810/51444 | 3.28E-01 | 0.0176 | 0.0171 | 1.0316 | 3.02E-01 | 6810/51444 | 4.18E-01 |
| Leu | 0.0315 | 0.0157 | 2.0084 | 4.46E-02 | 6810/51443 | 6.17E-02 | 0.0037 | 0.0161 | 0.2282 | 8.20E-01 | 6810/51443 | 8.61E-01 | 0.0148 | 0.0175 | 0.8409 | 4.00E-01 | 6810/51443 | 4.94E-01 | 0.0108 | 0.0177 | 0.6110 | 5.41E-01 | 6810/51443 | 6.67E-01 |
| Val | 0.0367 | 0.0157 | 2.3306 | 1.98E-02 | 6809/51434 | 2.88E-02 | -0.0001 | 0.0164 | -0.0069 | 9.95E-01 | 6809/51434 | 9.99E-01 | 0.0207 | 0.0179 | 1.1573 | 2.47E-01 | 6809/51434 | 3.42E-01 | 0.0185 | 0.0181 | 1.0239 | 3.06E-01 | 6809/51434 | 4.18E-01 |

| | | | | | | | | | | | | | | | | | | | | | | | | |
|---|---|---|---|---|---|---|---|---|---|---|---|---|---|---|---|---|---|---|---|---|---|---|---|---|
| Phe | -0.0101 | 0.0149 | -0.6792 | 4.97E-01 | 6809/51411 | 5.48E-01 | -0.0293 | 0.0151 | -1.9367 | 5.28E-02 | 6809/51411 | 7.82E-02 | -0.0345 | 0.0165 | -2.0965 | 3.60E-02 | 6809/51411 | 6.46E-02 | -0.0361 | 0.0166 | -2.1726 | 2.98E-02 | 6809/51411 | 5.75E-02 |
| Tyr | 0.0156 | 0.0151 | 1.0379 | 2.99E-01 | 6801/51357 | 3.52E-01 | -0.0102 | 0.0154 | -0.6626 | 5.08E-01 | 6801/51357 | 5.96E-01 | -0.0070 | 0.0167 | -0.4157 | 6.78E-01 | 6801/51357 | 7.64E-01 | -0.0063 | 0.0169 | -0.3745 | 7.08E-01 | 6801/51357 | 8.16E-01 |
| Glucose | -0.0100 | 0.0160 | -0.6214 | 5.34E-01 | 6800/51337 | 5.84E-01 | -0.0247 | 0.0161 | -1.5375 | 1.24E-01 | 6800/51337 | 1.76E-01 | -0.0378 | 0.0176 | -2.1499 | 3.16E-02 | 6800/51337 | 5.82E-02 | -0.0395 | 0.0183 | -2.1636 | 3.05E-02 | 6800/51337 | 5.80E-02 |
| Lactate | 0.0167 | 0.0148 | 1.1285 | 2.59E-01 | 6795/51308 | 3.16E-01 | 0.0064 | 0.0148 | 0.4324 | 6.65E-01 | 6795/51308 | 7.33E-01 | 0.0130 | 0.0160 | 0.8126 | 4.16E-01 | 6795/51308 | 5.08E-01 | 0.0133 | 0.0162 | 0.8173 | 4.14E-01 | 6795/51308 | 5.38E-01 |
| Pyruvate | 0.0439 | 0.0135 | 3.2437 | 1.18E-03 | 6790/51309 | 1.90E-03 | 0.0380 | 0.0136 | 2.7903 | 5.27E-03 | 6790/51309 | 9.47E-03 | 0.0399 | 0.0146 | 2.7224 | 6.48E-03 | 6790/51309 | 1.43E-02 | 0.0403 | 0.0148 | 2.7337 | 6.26E-03 | 6790/51309 | 1.50E-02 |
| Citrate | -0.0878 | 0.0153 | -5.7451 | 9.19E-09 | 6810/51435 | 2.08E-08 | -0.0820 | 0.0153 | -5.3500 | 8.80E-08 | 6810/51435 | 2.77E-07 | -0.0744 | 0.0166 | -4.4687 | 7.87E-06 | 6810/51435 | 3.70E-05 | -0.0661 | 0.0170 | -3.8896 | 1.00E-04 | 6810/51435 | 4.03E-04 |
| bOHbutyrate | -0.0157 | 0.0149 | -1.0555 | 2.91E-01 | 6735/50863 | 3.45E-01 | -0.0166 | 0.0150 | -1.1118 | 2.66E-01 | 6735/50863 | 3.45E-01 | -0.0066 | 0.0166 | -0.4009 | 6.89E-01 | 6735/50863 | 7.72E-01 | -0.0079 | 0.0167 | -0.4724 | 6.37E-01 | 6735/50863 | 7.59E-01 |
| Acetate | -0.0142 | 0.0162 | -0.8723 | 3.83E-01 | 6807/51433 | 4.38E-01 | -0.0032 | 0.0166 | -0.1929 | 8.47E-01 | 6807/51433 | 8.78E-01 | -0.0098 | 0.0178 | -0.5482 | 5.84E-01 | 6807/51433 | 6.85E-01 | -0.0061 | 0.0179 | -0.3389 | 7.35E-01 | 6807/51433 | 8.28E-01 |
| Acetoacetate | 0.0168 | 0.0144 | 1.1671 | 2.43E-01 | 6810/51444 | 3.01E-01 | 0.0105 | 0.0144 | 0.7259 | 4.68E-01 | 6810/51444 | 5.63E-01 | -0.0056 | 0.0152 | -0.3709 | 7.11E-01 | 6810/51444 | 7.83E-01 | -0.0069 | 0.0153 | -0.4529 | 6.51E-01 | 6810/51444 | 7.68E-01 |
| Acetone | 0.0075 | 0.0150 | 0.4992 | 6.18E-01 | 6810/51444 | 6.63E-01 | 0.0169 | 0.0150 | 1.1244 | 2.61E-01 | 6810/51444 | 3.40E-01 | 0.0188 | 0.0162 | 1.1577 | 2.47E-01 | 6810/51444 | 3.42E-01 | 0.0148 | 0.0164 | 0.9039 | 3.66E-01 | 6810/51444 | 4.82E-01 |
| Creatinine | 0.0456 | 0.0181 | 2.5181 | 1.18E-02 | 6399/49287 | 1.75E-02 | 0.0236 | 0.0184 | 1.2871 | 1.98E-01 | 6399/49287 | 2.68E-01 | 0.0036 | 0.0201 | 0.1797 | 8.57E-01 | 6399/49287 | 8.90E-01 | 0.0043 | 0.0203 | 0.2130 | 8.31E-01 | 6399/49287 | 9.04E-01 |
| Albumin | -0.0576 | 0.0151 | -3.8105 | 1.39E-04 | 6810/51429 | 2.54E-04 | -0.0371 | 0.0154 | -2.4097 | 1.60E-02 | 6810/51429 | 2.67E-02 | -0.0293 | 0.0167 | -1.7561 | 7.91E-02 | 6810/51429 | 1.34E-01 | -0.0212 | 0.0169 | -1.2549 | 2.10E-01 | 6810/51429 | 3.11E-01 |
| GlycA | 0.1133 | 0.0151 | 7.5068 | 6.06E-14 | 6810/51445 | 2.04E-13 | 0.0772 | 0.0162 | 4.7662 | 1.88E-06 | 6810/51445 | 4.87E-06 | 0.0458 | 0.0176 | 2.6070 | 9.14E-03 | 6810/51445 | 1.96E-02 | 0.0390 | 0.0179 | 2.1757 | 2.96E-02 | 6810/51445 | 5.75E-02 |
| XXL_VLDL_P | 0.1388 | 0.0154 | 8.9955 | 2.35E-19 | 6810/51445 | 2.02E-18 | 0.1099 | 0.0161 | 6.8326 | 8.34E-12 | 6810/51445 | 7.69E-11 | 0.0802 | 0.0173 | 4.6281 | 3.69E-06 | 6810/51445 | 1.96E-05 | 0.0761 | 0.0176 | 4.3297 | 1.49E-05 | 6810/51445 | 8.26E-05 |
| XXL_VLDL_L | 0.1348 | 0.0155 | 8.6705 | 4.30E-18 | 6810/51445 | 2.75E-17 | 0.1048 | 0.0162 | 6.4572 | 1.07E-10 | 6810/51445 | 6.32E-10 | 0.0746 | 0.0175 | 4.2547 | 2.09E-05 | 6810/51445 | 8.27E-05 | 0.0716 | 0.0178 | 4.0213 | 5.79E-05 | 6810/51445 | 2.44E-04 |
| XXL_VLDL_PL | 0.1283 | 0.0155 | 8.2723 | 1.31E-16 | 6810/51445 | 5.84E-16 | 0.1004 | 0.0160 | 6.2707 | 3.59E-10 | 6810/51445 | 1.81E-09 | 0.0739 | 0.0172 | 4.3080 | 1.65E-05 | 6810/51445 | 6.84E-05 | 0.0708 | 0.0174 | 4.0786 | 4.53E-05 | 6810/51445 | 2.01E-04 |
| XXL_VLDL_C | 0.1359 | 0.0154 | 8.8361 | 9.92E-19 | 6810/51445 | 7.72E-18 | 0.1065 | 0.0161 | 6.6235 | 3.51E-11 | 6810/51445 | 2.43E-10 | 0.0770 | 0.0174 | 4.4359 | 9.17E-06 | 6810/51445 | 4.15E-05 | 0.0749 | 0.0177 | 4.2348 | 2.29E-05 | 6810/51445 | 1.12E-04 |
| XXL_VLDL_CE | 0.1308 | 0.0153 | 8.5231 | 1.55E-17 | 6810/51445 | 8.79E-17 | 0.1014 | 0.0160 | 6.3363 | 2.35E-10 | 6810/51445 | 1.22E-09 | 0.0746 | 0.0173 | 4.3132 | 1.61E-05 | 6810/51445 | 6.84E-05 | 0.0735 | 0.0177 | 4.1573 | 3.22E-05 | 6810/51445 | 1.49E-04 |
| XXL_VLDL_FC | 0.1399 | 0.0155 | 9.0458 | 1.49E-19 | 6810/51445 | 1.42E-18 | 0.1108 | 0.0161 | 6.8702 | 6.41E-12 | 6810/51445 | 6.38E-11 | 0.0799 | 0.0174 | 4.5880 | 4.48E-06 | 6810/51445 | 2.32E-05 | 0.0766 | 0.0177 | 4.3358 | 1.45E-05 | 6810/51445 | 8.22E-05 |
| XXL_VLDL_TG | 0.0921 | 0.0160 | 5.7477 | 9.05E-09 | 6810/51445 | 2.07E-08 | 0.0659 | 0.0161 | 4.0817 | 4.47E-05 | 6810/51445 | 1.02E-04 | 0.0422 | 0.0172 | 2.4467 | 1.44E-02 | 6810/51445 | 2.85E-02 | 0.0410 | 0.0175 | 2.3478 | 1.89E-02 | 6810/51445 | 4.02E-02 |
| XL_VLDL_P | 0.1416 | 0.0153 | 9.2715 | 1.84E-20 | 6810/51445 | 2.54E-19 | 0.1129 | 0.0160 | 7.0770 | 1.47E-12 | 6810/51445 | 1.93E-11 | 0.0846 | 0.0172 | 4.9281 | 8.30E-07 | 6810/51445 | 7.28E-06 | 0.0806 | 0.0175 | 4.5996 | 4.23E-06 | 6810/51445 | 4.07E-05 |
| XL_VLDL_L | 0.1414 | 0.0153 | 9.2390 | 2.49E-20 | 6810/51445 | 2.95E-19 | 0.1123 | 0.0160 | 7.0063 | 2.45E-12 | 6810/51445 | 2.77E-11 | 0.0837 | 0.0173 | 4.8351 | 1.33E-06 | 6810/51445 | 8.96E-06 | 0.0804 | 0.0177 | 4.5472 | 5.44E-06 | 6810/51445 | 4.51E-05 |
| XL_VLDL_PL | 0.1207 | 0.0155 | 7.7778 | 7.38E-15 | 6810/51445 | 2.74E-14 | 0.0933 | 0.0158 | 5.8928 | 3.80E-09 | 6810/51445 | 1.52E-08 | 0.0707 | 0.0169 | 4.1875 | 2.82E-05 | 6810/51445 | 1.10E-04 | 0.0685 | 0.0172 | 3.9766 | 6.99E-05 | 6810/51445 | 2.90E-04 |
| XL_VLDL_C | 0.1153 | 0.0150 | 7.6796 | 1.60E-14 | 6810/51445 | 5.76E-14 | 0.0877 | 0.0155 | 5.6515 | 1.59E-08 | 6810/51445 | 5.66E-08 | 0.0628 | 0.0167 | 3.7628 | 1.68E-04 | 6810/51445 | 5.23E-04 | 0.0635 | 0.0173 | 3.6622 | 2.50E-04 | 6810/51445 | 9.03E-04 |
| XL_VLDL_CE | 0.1008 | 0.0149 | 6.7670 | 1.32E-11 | 6810/51445 | 3.72E-11 | 0.0746 | 0.0153 | 4.8834 | 1.04E-06 | 6810/51445 | 2.82E-06 | 0.0533 | 0.0164 | 3.2405 | 1.19E-03 | 6810/51445 | 3.06E-03 | 0.0559 | 0.0172 | 3.2499 | 1.15E-03 | 6810/51445 | 3.23E-03 |
| XL_VLDL_FC | 0.1288 | 0.0152 | 8.4911 | 2.05E-17 | 6810/51445 | 1.06E-16 | 0.1006 | 0.0158 | 6.3826 | 1.74E-10 | 6810/51445 | 9.22E-10 | 0.0734 | 0.0170 | 4.3255 | 1.52E-05 | 6810/51445 | 6.65E-05 | 0.0716 | 0.0175 | 4.1027 | 4.08E-05 | 6810/51445 | 1.85E-04 |
| XL_VLDL_TG | 0.1427 | 0.0158 | 9.0022 | 2.21E-19 | 6810/51445 | 1.97E-18 | 0.1126 | 0.0164 | 6.8536 | 7.20E-12 | 6810/51445 | 6.90E-11 | 0.0911 | 0.0178 | 5.1290 | 2.91E-07 | 6810/51445 | 3.65E-06 | 0.0871 | 0.0180 | 4.8497 | 1.24E-06 | 6810/51445 | 2.39E-05 |
| L_VLDL_P | 0.1374 | 0.0152 | 9.0420 | 1.54E-19 | 6810/51445 | 1.42E-18 | 0.1092 | 0.0158 | 6.8961 | 5.34E-12 | 6810/51445 | 5.54E-11 | 0.0821 | 0.0171 | 4.7903 | 1.67E-06 | 6810/51445 | 1.06E-05 | 0.0793 | 0.0176 | 4.5151 | 6.33E-06 | 6810/51445 | 5.08E-05 |
| L_VLDL_L | 0.1337 | 0.0152 | 8.8117 | 1.23E-18 | 6810/51445 | 9.30E-18 | 0.1056 | 0.0158 | 6.6862 | 2.29E-11 | 6810/51445 | 1.78E-10 | 0.0800 | 0.0171 | 4.6747 | 2.94E-06 | 6810/51445 | 1.63E-05 | 0.0778 | 0.0175 | 4.4366 | 9.14E-06 | 6810/51445 | 5.69E-05 |
| L_VLDL_PL | 0.1087 | 0.0156 | 6.9560 | 3.50E-12 | 6810/51445 | 1.05E-11 | 0.0825 | 0.0157 | 5.2641 | 1.41E-07 | 6810/51445 | 4.24E-07 | 0.0656 | 0.0167 | 3.9218 | 8.79E-05 | 6810/51445 | 2.96E-04 | 0.0626 | 0.0170 | 3.6896 | 2.25E-04 | 6810/51445 | 8.22E-04 |
| L_VLDL_C | 0.1172 | 0.0149 | 7.8537 | 4.04E-15 | 6810/51445 | 1.52E-14 | 0.0908 | 0.0154 | 5.8995 | 3.65E-09 | 6810/51445 | 1.49E-08 | 0.0662 | 0.0166 | 3.9902 | 6.60E-05 | 6810/51445 | 2.28E-04 | 0.0662 | 0.0172 | 3.8522 | 1.17E-04 | 6810/51445 | 4.55E-04 |
| L_VLDL_CE | 0.1014 | 0.0148 | 6.8692 | 6.46E-12 | 6810/51445 | 1.87E-11 | 0.0779 | 0.0151 | 5.1681 | 2.36E-07 | 6810/51445 | 6.77E-07 | 0.0557 | 0.0162 | 3.4287 | 6.07E-04 | 6810/51445 | 1.64E-03 | 0.0576 | 0.0170 | 3.3965 | 6.83E-04 | 6810/51445 | 2.15E-03 |
| L_VLDL_FC | 0.1317 | 0.0151 | 8.7221 | 2.73E-18 | 6810/51445 | 1.84E-17 | 0.1034 | 0.0157 | 6.5687 | 5.08E-11 | 6810/51445 | 3.33E-10 | 0.0771 | 0.0170 | 4.5415 | 5.59E-06 | 6810/51445 | 2.83E-05 | 0.0750 | 0.0174 | 4.3035 | 1.68E-05 | 6810/51445 | 9.10E-05 |
| L_VLDL_TG | 0.1325 | 0.0156 | 8.4975 | 1.94E-17 | 6810/51445 | 1.03E-16 | 0.1034 | 0.0162 | 6.3945 | 1.61E-10 | 6810/51445 | 8.72E-10 | 0.0851 | 0.0176 | 4.8288 | 1.37E-06 | 6810/51445 | 9.00E-06 | 0.0825 | 0.0179 | 4.5989 | 4.25E-06 | 6810/51445 | 4.07E-05 |
| M_VLDL_P | 0.0643 | 0.0147 | 4.3814 | 1.18E-05 | 6810/51445 | 2.31E-05 | 0.0474 | 0.0148 | 3.2027 | 1.36E-03 | 6810/51445 | 2.67E-03 | 0.0284 | 0.0160 | 1.7712 | 7.65E-02 | 6810/51445 | 1.31E-01 | 0.0320 | 0.0172 | 1.8653 | 6.21E-02 | 6810/51445 | 1.12E-01 |
| M_VLDL_L | 0.0816 | 0.0148 | 5.5232 | 3.33E-08 | 6810/51445 | 7.40E-08 | 0.0617 | 0.0150 | 4.1188 | 3.81E-05 | 6810/51445 | 8.86E-05 | 0.0404 | 0.0162 | 2.4913 | 1.27E-02 | 6810/51445 | 2.58E-02 | 0.0429 | 0.0171 | 2.5077 | 1.22E-02 | 6810/51445 | 2.78E-02 |

| | | | | | | | | | | | | | | | | | | | | | | | | |
|---|---|---|---|---|---|---|---|---|---|---|---|---|---|---|---|---|---|---|---|---|---|---|---|---|
| M_VLDL_PL | 0.0618 | 0.0148 | 4.1812 | 2.90E-05 | 6810/51445 | 5.64E-05 | 0.0465 | 0.0148 | 3.1356 | 1.71E-03 | 6810/51445 | 3.32E-03 | 0.0273 | 0.0160 | 1.7070 | 8.78E-02 | 6810/51445 | 1.45E-01 | 0.0309 | 0.0173 | 1.7882 | 7.37E-02 | 6810/51445 | 1.29E-01 |
| M_VLDL_C | 0.0036 | 0.0148 | 0.2452 | 8.06E-01 | 6810/51445 | 8.33E-01 | 0.0027 | 0.0147 | 0.1844 | 8.54E-01 | 6810/51445 | 8.78E-01 | -0.0063 | 0.0160 | -0.3930 | 6.94E-01 | 6810/51445 | 7.75E-01 | -0.0022 | 0.0179 | -0.1238 | 9.01E-01 | 6810/51445 | 9.39E-01 |
| M_VLDL_CE | -0.0253 | 0.0150 | -1.6841 | 9.22E-02 | 6810/51445 | 1.24E-01 | -0.0171 | 0.0151 | -1.1331 | 2.57E-01 | 6810/51445 | 3.37E-01 | -0.0165 | 0.0164 | -1.0083 | 3.13E-01 | 6810/51445 | 4.04E-01 | -0.0113 | 0.0183 | -0.6183 | 5.36E-01 | 6810/51445 | 6.64E-01 |
| M_VLDL_FC | 0.0420 | 0.0147 | 2.8685 | 4.12E-03 | 6810/51445 | 6.34E-03 | 0.0309 | 0.0147 | 2.1017 | 3.56E-02 | 6810/51445 | 5.37E-02 | 0.0148 | 0.0159 | 0.9327 | 3.51E-01 | 6810/51445 | 4.46E-01 | 0.0190 | 0.0173 | 1.0989 | 2.72E-01 | 6810/51445 | 3.82E-01 |
| M_VLDL_TG | 0.1221 | 0.0152 | 8.0431 | 8.76E-16 | 6810/51445 | 3.58E-15 | 0.0954 | 0.0157 | 6.0911 | 1.12E-09 | 6810/51445 | 5.11E-09 | 0.0764 | 0.0170 | 4.4882 | 7.18E-06 | 6810/51445 | 3.51E-05 | 0.0738 | 0.0174 | 4.2416 | 2.22E-05 | 6810/51445 | 1.11E-04 |
| S_VLDL_P | 0.1023 | 0.0147 | 6.9458 | 3.76E-12 | 6810/51445 | 1.12E-11 | 0.0793 | 0.0151 | 5.2634 | 1.41E-07 | 6810/51445 | 4.24E-07 | 0.0524 | 0.0163 | 3.2133 | 1.31E-03 | 6810/51445 | 3.30E-03 | 0.0506 | 0.0169 | 2.9970 | 2.73E-03 | 6810/51445 | 7.00E-03 |
| S_VLDL_L | 0.0986 | 0.0147 | 6.7068 | 1.99E-11 | 6810/51445 | 5.44E-11 | 0.0760 | 0.0150 | 5.0542 | 4.32E-07 | 6810/51445 | 1.21E-06 | 0.0495 | 0.0163 | 3.0414 | 2.35E-03 | 6810/51445 | 5.53E-03 | 0.0484 | 0.0169 | 2.8653 | 4.17E-03 | 6810/51445 | 1.03E-02 |
| S_VLDL_PL | 0.0656 | 0.0146 | 4.4967 | 6.90E-06 | 6810/51445 | 1.37E-05 | 0.0481 | 0.0147 | 3.2602 | 1.11E-03 | 6810/51445 | 2.22E-03 | 0.0279 | 0.0160 | 1.7457 | 8.09E-02 | 6810/51445 | 1.36E-01 | 0.0294 | 0.0170 | 1.7326 | 8.32E-02 | 6810/51445 | 1.44E-01 |
| S_VLDL_C | 0.0578 | 0.0145 | 3.9940 | 6.50E-05 | 6810/51445 | 1.23E-04 | 0.0412 | 0.0146 | 2.8181 | 4.83E-03 | 6810/51445 | 8.91E-03 | 0.0228 | 0.0158 | 1.4439 | 1.49E-01 | 6810/51445 | 2.22E-01 | 0.0250 | 0.0169 | 1.4745 | 1.40E-01 | 6810/51445 | 2.30E-01 |
| S_VLDL_CE | 0.0686 | 0.0144 | 4.7495 | 2.04E-06 | 6810/51445 | 4.13E-06 | 0.0494 | 0.0146 | 3.3718 | 7.47E-04 | 6810/51445 | 1.52E-03 | 0.0295 | 0.0158 | 1.8617 | 6.26E-02 | 6810/51445 | 1.09E-01 | 0.0311 | 0.0168 | 1.8472 | 6.47E-02 | 6810/51445 | 1.16E-01 |
| S_VLDL_FC | 0.0373 | 0.0146 | 2.5581 | 1.05E-02 | 6810/51445 | 1.57E-02 | 0.0262 | 0.0146 | 1.7904 | 7.34E-02 | 6810/51445 | 1.07E-01 | 0.0113 | 0.0159 | 0.7101 | 4.78E-01 | 6810/51445 | 5.77E-01 | 0.0140 | 0.0172 | 0.8155 | 4.15E-01 | 6810/51445 | 5.38E-01 |
| S_VLDL_TG | 0.1397 | 0.0150 | 9.2893 | 1.55E-20 | 6810/51445 | 2.28E-19 | 0.1146 | 0.0156 | 7.3624 | 1.81E-13 | 6810/51445 | 3.75E-12 | 0.0828 | 0.0169 | 4.9133 | 8.96E-07 | 6810/51445 | 7.38E-06 | 0.0766 | 0.0171 | 4.4861 | 7.25E-06 | 6810/51445 | 5.40E-05 |
| XS_VLDL_P | 0.0410 | 0.0148 | 2.7757 | 5.51E-03 | 6810/51445 | 8.41E-03 | 0.0318 | 0.0148 | 2.1478 | 3.17E-02 | 6810/51445 | 4.85E-02 | 0.0049 | 0.0160 | 0.3027 | 7.62E-01 | 6810/51445 | 8.26E-01 | 0.0030 | 0.0173 | 0.1704 | 8.65E-01 | 6810/51445 | 9.20E-01 |
| XS_VLDL_L | 0.0474 | 0.0148 | 3.2009 | 1.37E-03 | 6810/51445 | 2.19E-03 | 0.0386 | 0.0149 | 2.5994 | 9.34E-03 | 6810/51445 | 1.63E-02 | 0.0095 | 0.0161 | 0.5900 | 5.55E-01 | 6810/51445 | 6.61E-01 | 0.0079 | 0.0173 | 0.4559 | 6.48E-01 | 6810/51445 | 7.68E-01 |
| XS_VLDL_PL | 0.0578 | 0.0148 | 3.9035 | 9.48E-05 | 6810/51445 | 1.76E-04 | 0.0462 | 0.0149 | 3.1004 | 1.93E-03 | 6810/51445 | 3.70E-03 | 0.0165 | 0.0161 | 1.0226 | 3.06E-01 | 6810/51445 | 3.97E-01 | 0.0140 | 0.0172 | 0.8156 | 4.15E-01 | 6810/51445 | 5.38E-01 |
| XS_VLDL_C | 0.0013 | 0.0151 | 0.0854 | 9.32E-01 | 6810/51445 | 9.36E-01 | 0.0040 | 0.0150 | 0.2676 | 7.89E-01 | 6810/51445 | 8.36E-01 | -0.0168 | 0.0163 | -1.0283 | 3.04E-01 | 6810/51445 | 3.96E-01 | -0.0182 | 0.0183 | -0.9979 | 3.18E-01 | 6810/51445 | 4.33E-01 |
| XS_VLDL_CE | -0.0123 | 0.0152 | -0.8102 | 4.18E-01 | 6810/51445 | 4.75E-01 | -0.0054 | 0.0152 | -0.3569 | 7.21E-01 | 6810/51445 | 7.77E-01 | -0.0240 | 0.0164 | -1.4615 | 1.44E-01 | 6810/51445 | 2.16E-01 | -0.0260 | 0.0187 | -1.3959 | 1.63E-01 | 6810/51445 | 2.53E-01 |
| XS_VLDL_FC | 0.0323 | 0.0148 | 2.1771 | 2.95E-02 | 6810/51445 | 4.15E-02 | 0.0256 | 0.0149 | 1.7250 | 8.45E-02 | 6810/51445 | 1.22E-01 | 0.0006 | 0.0161 | 0.0369 | 9.71E-01 | 6810/51445 | 9.86E-01 | -0.0007 | 0.0175 | -0.0389 | 9.69E-01 | 6810/51445 | 9.77E-01 |
| XS_VLDL_TG | 0.1269 | 0.0148 | 8.5485 | 1.25E-17 | 6810/51445 | 7.39E-17 | 0.1038 | 0.0152 | 6.8101 | 9.76E-12 | 6810/51445 | 8.68E-11 | 0.0664 | 0.0165 | 4.0161 | 5.92E-05 | 6810/51445 | 2.08E-04 | 0.0568 | 0.0167 | 3.3917 | 6.95E-04 | 6810/51445 | 2.16E-03 |
| IDL_P | -0.0108 | 0.0148 | -0.7292 | 4.66E-01 | 6810/51445 | 5.18E-01 | -0.0117 | 0.0148 | -0.7941 | 4.27E-01 | 6810/51445 | 5.21E-01 | -0.0226 | 0.0160 | -1.4081 | 1.59E-01 | 6810/51445 | 2.33E-01 | -0.0260 | 0.0181 | -1.4321 | 1.52E-01 | 6810/51445 | 2.43E-01 |
| IDL_L | -0.0350 | 0.0152 | -2.3062 | 2.11E-02 | 6810/51445 | 3.04E-02 | -0.0254 | 0.0152 | -1.6682 | 9.53E-02 | 6810/51445 | 1.36E-01 | -0.0375 | 0.0165 | -2.2765 | 2.28E-02 | 6810/51445 | 4.30E-02 | -0.0421 | 0.0187 | -2.2545 | 2.42E-02 | 6810/51445 | 4.89E-02 |
| IDL_PL | -0.0309 | 0.0152 | -2.0330 | 4.20E-02 | 6810/51445 | 5.85E-02 | -0.0209 | 0.0152 | -1.3713 | 1.70E-01 | 6810/51445 | 2.36E-01 | -0.0346 | 0.0165 | -2.0962 | 3.61E-02 | 6810/51445 | 6.46E-02 | -0.0382 | 0.0186 | -2.0482 | 4.05E-02 | 6810/51445 | 7.48E-02 |
| IDL_C | -0.0511 | 0.0153 | -3.3524 | 8.01E-04 | 6810/51445 | 1.33E-03 | -0.0382 | 0.0153 | -2.4937 | 1.26E-02 | 6810/51445 | 2.14E-02 | -0.0453 | 0.0166 | -2.7256 | 6.42E-03 | 6810/51445 | 1.43E-02 | -0.0502 | 0.0189 | -2.6507 | 8.03E-03 | 6810/51445 | 1.85E-02 |
| IDL_CE | -0.0478 | 0.0153 | -3.1345 | 1.72E-03 | 6810/51445 | 2.70E-03 | -0.0349 | 0.0153 | -2.2809 | 2.26E-02 | 6810/51445 | 3.65E-02 | -0.0427 | 0.0166 | -2.5728 | 1.01E-02 | 6810/51445 | 2.11E-02 | -0.0473 | 0.0189 | -2.5023 | 1.23E-02 | 6810/51445 | 2.79E-02 |
| IDL_FC | -0.0596 | 0.0152 | -3.9121 | 9.15E-05 | 6810/51445 | 1.71E-04 | -0.0467 | 0.0153 | -3.0528 | 2.27E-03 | 6810/51445 | 4.28E-03 | -0.0515 | 0.0166 | -3.1050 | 1.90E-03 | 6810/51445 | 4.64E-03 | -0.0566 | 0.0188 | -3.0020 | 2.68E-03 | 6810/51445 | 7.00E-03 |
| IDL_TG | 0.1076 | 0.0148 | 7.2504 | 4.16E-13 | 6810/51445 | 1.34E-12 | 0.0869 | 0.0151 | 5.7439 | 9.25E-09 | 6810/51445 | 3.49E-08 | 0.0489 | 0.0165 | 2.9716 | 2.96E-03 | 6810/51445 | 6.83E-03 | 0.0385 | 0.0168 | 2.2940 | 2.18E-02 | 6810/51445 | 4.56E-02 |
| L_LDL_P | 0.0079 | 0.0147 | 0.5347 | 5.93E-01 | 6810/51445 | 6.42E-01 | 0.0061 | 0.0147 | 0.4176 | 6.76E-01 | 6810/51445 | 7.42E-01 | -0.0049 | 0.0159 | -0.3047 | 7.61E-01 | 6810/51445 | 8.26E-01 | -0.0039 | 0.0175 | -0.2201 | 8.26E-01 | 6810/51445 | 9.04E-01 |
| L_LDL_L | -0.0182 | 0.0149 | -1.2191 | 2.23E-01 | 6810/51445 | 2.79E-01 | -0.0142 | 0.0149 | -0.9517 | 3.41E-01 | 6810/51445 | 4.27E-01 | -0.0228 | 0.0161 | -1.4166 | 1.57E-01 | 6810/51445 | 2.31E-01 | -0.0222 | 0.0177 | -1.2563 | 2.09E-01 | 6810/51445 | 3.11E-01 |
| L_LDL_PL | -0.0201 | 0.0148 | -1.3557 | 1.75E-01 | 6810/51445 | 2.24E-01 | -0.0179 | 0.0148 | -1.2088 | 2.27E-01 | 6810/51445 | 3.04E-01 | -0.0254 | 0.0161 | -1.5775 | 1.15E-01 | 6810/51445 | 1.78E-01 | -0.0248 | 0.0177 | -1.4005 | 1.61E-01 | 6810/51445 | 2.53E-01 |
| L_LDL_C | -0.0237 | 0.0130 | -1.8274 | 6.76E-02 | 6810/51445 | 9.25E-02 | -0.0192 | 0.0127 | -1.5156 | 1.30E-01 | 6810/51445 | 1.82E-01 | -0.0223 | 0.0133 | -1.6802 | 9.29E-02 | 6810/51445 | 1.50E-01 | -0.0185 | 0.0143 | -1.2944 | 1.96E-01 | 6810/51445 | 2.93E-01 |
| L_LDL_CE | -0.0205 | 0.0139 | -1.4763 | 1.40E-01 | 6810/51445 | 1.85E-01 | -0.0165 | 0.0139 | -1.1861 | 2.36E-01 | 6810/51445 | 3.14E-01 | -0.0225 | 0.0147 | -1.5289 | 1.26E-01 | 6810/51445 | 1.93E-01 | -0.0188 | 0.0159 | -1.1840 | 2.36E-01 | 6810/51445 | 3.42E-01 |
| L_LDL_FC | -0.0413 | 0.0155 | -2.6621 | 7.76E-03 | 6810/51445 | 1.17E-02 | -0.0310 | 0.0140 | -2.2153 | 2.67E-02 | 6810/51445 | 4.21E-02 | -0.0301 | 0.0142 | -2.1124 | 3.47E-02 | 6810/51445 | 6.30E-02 | -0.0264 | 0.0153 | -1.7288 | 8.38E-02 | 6810/51445 | 1.44E-01 |
| L_LDL_TG | 0.1005 | 0.0149 | 6.7415 | 1.57E-11 | 6810/51445 | 4.39E-11 | 0.0786 | 0.0152 | 5.1715 | 2.32E-07 | 6810/51445 | 6.72E-07 | 0.0420 | 0.0166 | 2.5387 | 1.11E-02 | 6810/51445 | 2.27E-02 | 0.0307 | 0.0169 | 1.8150 | 6.95E-02 | 6810/51445 | 1.23E-01 |
| M_LDL_P | 0.0201 | 0.0147 | 1.3715 | 1.70E-01 | 6810/51445 | 2.18E-01 | 0.0091 | 0.0147 | 0.6174 | 5.37E-01 | 6810/51445 | 6.22E-01 | -0.0047 | 0.0160 | -0.2970 | 7.67E-01 | 6810/51445 | 8.26E-01 | -0.0019 | 0.0173 | -0.1116 | 9.11E-01 | 6810/51445 | 9.40E-01 |
| M_LDL_L | 0.0231 | 0.0147 | 1.5692 | 1.17E-01 | 6810/51445 | 1.55E-01 | 0.0126 | 0.0148 | 0.8502 | 3.95E-01 | 6810/51445 | 4.90E-01 | 0.0004 | 0.0160 | 0.0227 | 9.82E-01 | 6810/51445 | 9.86E-01 | 0.0021 | 0.0172 | 0.1227 | 9.02E-01 | 6810/51445 | 9.39E-01 |
| M_LDL_PL | 0.0195 | 0.0146 | 1.3367 | 1.81E-01 | 6810/51445 | 2.30E-01 | 0.0098 | 0.0145 | 0.6769 | 4.98E-01 | 6810/51445 | 5.88E-01 | 0.0005 | 0.0156 | 0.0288 | 9.77E-01 | 6810/51445 | 9.86E-01 | 0.0016 | 0.0167 | 0.0964 | 9.23E-01 | 6810/51445 | 9.46E-01 |

| | | | | | | | | | | | | | | | | | | | | | | | | |
|---|---|---|---|---|---|---|---|---|---|---|---|---|---|---|---|---|---|---|---|---|---|---|---|---|
| M_LDL_C | 0.0060 | 0.0140 | 0.4282 | 6.68E-01 | 6810/51445 | 7.05E-01 | -0.0004 | 0.0132 | -0.0272 | 9.78E-01 | 6810/51445 | 9.86E-01 | -0.0083 | 0.0132 | -0.6290 | 5.29E-01 | 6810/51445 | 6.37E-01 | -0.0043 | 0.0139 | -0.3121 | 7.55E-01 | 6810/51445 | 8.47E-01 |
| M_LDL_CE | 0.0164 | 0.0152 | 1.0776 | 2.81E-01 | 6810/51445 | 3.36E-01 | 0.0060 | 0.0137 | 0.4378 | 6.62E-01 | 6810/51445 | 7.32E-01 | -0.0048 | 0.0130 | -0.3656 | 7.15E-01 | 6810/51445 | 7.84E-01 | -0.0009 | 0.0136 | -0.0646 | 9.49E-01 | 6810/51445 | 9.60E-01 |
| M_LDL_FC | -0.0241 | 0.0130 | -1.8486 | 6.45E-02 | 6810/51445 | 8.88E-02 | -0.0220 | 0.0129 | -1.6980 | 8.95E-02 | 6810/51445 | 1.29E-01 | -0.0212 | 0.0134 | -1.5838 | 1.13E-01 | 6810/51445 | 1.77E-01 | -0.0170 | 0.0142 | -1.1911 | 2.34E-01 | 6810/51445 | 3.40E-01 |
| M_LDL_TG | 0.1125 | 0.0150 | 7.4919 | 6.79E-14 | 6810/51445 | 2.25E-13 | 0.0879 | 0.0154 | 5.6938 | 1.24E-08 | 6810/51445 | 4.55E-08 | 0.0513 | 0.0168 | 3.0536 | 2.26E-03 | 6810/51445 | 5.36E-03 | 0.0409 | 0.0171 | 2.3899 | 1.69E-02 | 6810/51445 | 3.62E-02 |
| S_LDL_P | 0.0257 | 0.0146 | 1.7570 | 7.89E-02 | 6810/51445 | 1.07E-01 | 0.0151 | 0.0147 | 1.0262 | 3.05E-01 | 6810/51445 | 3.87E-01 | -0.0040 | 0.0159 | -0.2502 | 8.02E-01 | 6810/51445 | 8.52E-01 | -0.0036 | 0.0174 | -0.2088 | 8.35E-01 | 6810/51445 | 9.04E-01 |
| S_LDL_L | 0.0141 | 0.0147 | 0.9568 | 3.39E-01 | 6810/51445 | 3.94E-01 | 0.0058 | 0.0147 | 0.3934 | 6.94E-01 | 6810/51445 | 7.58E-01 | -0.0062 | 0.0160 | -0.3863 | 6.99E-01 | 6810/51445 | 7.77E-01 | -0.0070 | 0.0173 | -0.4021 | 6.88E-01 | 6810/51445 | 8.04E-01 |
| S_LDL_PL | -0.0038 | 0.0147 | -0.2575 | 7.97E-01 | 6810/51445 | 8.27E-01 | -0.0073 | 0.0147 | -0.4933 | 6.22E-01 | 6810/51445 | 6.91E-01 | -0.0144 | 0.0160 | -0.9040 | 3.66E-01 | 6810/51445 | 4.60E-01 | -0.0192 | 0.0173 | -1.1091 | 2.67E-01 | 6810/51445 | 3.78E-01 |
| S_LDL_C | -0.0007 | 0.0130 | -0.0572 | 9.54E-01 | 6810/51445 | 9.54E-01 | -0.0048 | 0.0125 | -0.3877 | 6.98E-01 | 6810/51445 | 7.59E-01 | -0.0113 | 0.0126 | -0.9008 | 3.68E-01 | 6810/51445 | 4.60E-01 | -0.0081 | 0.0133 | -0.6077 | 5.43E-01 | 6810/51445 | 6.67E-01 |
| S_LDL_CE | 0.0134 | 0.0150 | 0.8938 | 3.71E-01 | 6810/51445 | 4.26E-01 | 0.0046 | 0.0137 | 0.3334 | 7.39E-01 | 6810/51445 | 7.90E-01 | -0.0065 | 0.0130 | -0.5040 | 6.14E-01 | 6810/51445 | 7.11E-01 | -0.0032 | 0.0137 | -0.2355 | 8.14E-01 | 6810/51445 | 9.01E-01 |
| S_LDL_FC | -0.0348 | 0.0140 | -2.4812 | 1.31E-02 | 6810/51445 | 1.92E-02 | -0.0304 | 0.0136 | -2.2304 | 2.57E-02 | 6810/51445 | 4.08E-02 | -0.0236 | 0.0136 | -1.7371 | 8.24E-02 | 6810/51445 | 1.38E-01 | -0.0201 | 0.0144 | -1.3965 | 1.63E-01 | 6810/51445 | 2.53E-01 |
| S_LDL_TG | 0.1282 | 0.0151 | 8.4806 | 2.24E-17 | 6810/51445 | 1.14E-16 | 0.1020 | 0.0156 | 6.5278 | 6.67E-11 | 6810/51445 | 4.26E-10 | 0.0641 | 0.0170 | 3.7639 | 1.67E-04 | 6810/51445 | 5.23E-04 | 0.0560 | 0.0173 | 3.2389 | 1.20E-03 | 6810/51445 | 3.32E-03 |
| XL_HDL_P | -0.1300 | 0.0157 | -8.2852 | 1.18E-16 | 6810/51445 | 5.46E-16 | -0.0944 | 0.0168 | -5.6137 | 1.98E-08 | 6810/51445 | 6.85E-08 | -0.0960 | 0.0182 | -5.2621 | 1.42E-07 | 6810/51445 | 2.75E-06 | -0.0972 | 0.0192 | -5.0560 | 4.28E-07 | 6810/51445 | 1.78E-05 |
| XL_HDL_L | -0.1335 | 0.0158 | -8.4440 | 3.07E-17 | 6810/51445 | 1.53E-16 | -0.0975 | 0.0170 | -5.7303 | 1.00E-08 | 6810/51445 | 3.72E-08 | -0.0936 | 0.0185 | -5.0671 | 4.04E-07 | 6810/51445 | 4.37E-06 | -0.0939 | 0.0193 | -4.8587 | 1.18E-06 | 6810/51445 | 2.39E-05 |
| XL_HDL_PL | -0.0835 | 0.0141 | -5.9368 | 2.91E-09 | 6810/51445 | 6.83E-09 | -0.0561 | 0.0149 | -3.7548 | 1.73E-04 | 6810/51445 | 3.79E-04 | -0.0535 | 0.0161 | -3.3259 | 8.81E-04 | 6810/51445 | 2.29E-03 | -0.0514 | 0.0166 | -3.1049 | 1.90E-03 | 6810/51445 | 5.21E-03 |
| XL_HDL_C | -0.1423 | 0.0155 | -9.1498 | 5.70E-20 | 6810/51445 | 5.68E-19 | -0.1086 | 0.0167 | -6.5043 | 7.81E-11 | 6810/51445 | 4.86E-10 | -0.1001 | 0.0181 | -5.5366 | 3.08E-08 | 6810/51445 | 1.28E-06 | -0.1006 | 0.0189 | -5.3094 | 1.10E-07 | 6810/51445 | 5.48E-06 |
| XL_HDL_CE | -0.1211 | 0.0142 | -8.5095 | 1.75E-17 | 6810/51445 | 9.67E-17 | -0.0923 | 0.0152 | -6.0900 | 1.13E-09 | 6810/51445 | 5.11E-09 | -0.0838 | 0.0165 | -5.0830 | 3.71E-07 | 6810/51445 | 4.20E-06 | -0.0808 | 0.0172 | -4.6846 | 2.81E-06 | 6810/51445 | 3.68E-05 |
| XL_HDL_FC | -0.1115 | 0.0148 | -7.5498 | 4.36E-14 | 6810/51445 | 1.49E-13 | -0.0812 | 0.0153 | -5.3046 | 1.13E-07 | 6810/51445 | 3.47E-07 | -0.0804 | 0.0164 | -4.8940 | 9.88E-07 | 6810/51445 | 7.63E-06 | -0.0830 | 0.0171 | -4.8481 | 1.25E-06 | 6810/51445 | 2.39E-05 |
| XL_HDL_TG | 0.0585 | 0.0150 | 3.8910 | 9.99E-05 | 6810/51445 | 1.84E-04 | 0.0558 | 0.0150 | 3.7088 | 2.08E-04 | 6810/51445 | 4.47E-04 | 0.0192 | 0.0163 | 1.1779 | 2.39E-01 | 6810/51445 | 3.36E-01 | 0.0157 | 0.0166 | 0.9430 | 3.46E-01 | 6810/51445 | 4.63E-01 |
| L_HDL_P | -0.1421 | 0.0162 | -8.7680 | 1.82E-18 | 6810/51445 | 1.33E-17 | -0.1055 | 0.0176 | -6.0043 | 1.92E-09 | 6810/51445 | 8.25E-09 | -0.0977 | 0.0191 | -5.1186 | 3.08E-07 | 6810/51445 | 3.65E-06 | -0.0917 | 0.0198 | -4.6199 | 3.84E-06 | 6810/51445 | 4.07E-05 |
| L_HDL_L | -0.1440 | 0.0166 | -8.6797 | 3.97E-18 | 6810/51445 | 2.60E-17 | -0.1063 | 0.0179 | -5.9452 | 2.76E-09 | 6810/51445 | 1.15E-08 | -0.0980 | 0.0194 | -5.0413 | 4.62E-07 | 6810/51445 | 4.80E-06 | -0.0932 | 0.0202 | -4.6145 | 3.94E-06 | 6810/51445 | 4.07E-05 |
| L_HDL_PL | -0.1300 | 0.0161 | -8.0817 | 6.39E-16 | 6810/51445 | 2.65E-15 | -0.0944 | 0.0174 | -5.4158 | 6.10E-08 | 6810/51445 | 2.03E-07 | -0.0893 | 0.0192 | -4.6626 | 3.12E-06 | 6810/51445 | 1.69E-05 | -0.0853 | 0.0199 | -4.2753 | 1.91E-05 | 6810/51445 | 9.81E-05 |
| L_HDL_C | -0.1450 | 0.0157 | -9.2476 | 2.30E-20 | 6810/51445 | 2.86E-19 | -0.1109 | 0.0171 | -6.4993 | 8.07E-11 | 6810/51445 | 4.90E-10 | -0.0969 | 0.0187 | -5.1936 | 2.06E-07 | 6810/51445 | 3.12E-06 | -0.0904 | 0.0194 | -4.6641 | 3.10E-06 | 6810/51445 | 3.86E-05 |
| L_HDL_CE | -0.1476 | 0.0157 | -9.4096 | 4.98E-21 | 6810/51445 | 9.44E-20 | -0.1136 | 0.0171 | -6.6423 | 3.09E-11 | 6810/51445 | 2.26E-10 | -0.0970 | 0.0187 | -5.1768 | 2.26E-07 | 6810/51445 | 3.12E-06 | -0.0896 | 0.0194 | -4.6069 | 4.09E-06 | 6810/51445 | 4.07E-05 |
| L_HDL_FC | -0.1266 | 0.0154 | -8.1997 | 2.41E-16 | 6810/51445 | 1.05E-15 | -0.0938 | 0.0166 | -5.6441 | 1.66E-08 | 6810/51445 | 5.82E-08 | -0.0880 | 0.0181 | -4.8680 | 1.13E-06 | 6810/51445 | 8.02E-06 | -0.0838 | 0.0188 | -4.4613 | 8.15E-06 | 6810/51445 | 5.48E-05 |
| L_HDL_TG | 0.0322 | 0.0152 | 2.1207 | 3.39E-02 | 6810/51445 | 4.75E-02 | 0.0399 | 0.0153 | 2.6181 | 8.84E-03 | 6810/51445 | 1.55E-02 | 0.0074 | 0.0165 | 0.4482 | 6.54E-01 | 6810/51445 | 7.44E-01 | 0.0074 | 0.0167 | 0.4434 | 6.57E-01 | 6810/51445 | 7.72E-01 |
| M_HDL_P | -0.0790 | 0.0153 | -5.1762 | 2.26E-07 | 6810/51445 | 4.90E-07 | -0.0566 | 0.0157 | -3.5989 | 3.20E-04 | 6810/51445 | 6.74E-04 | -0.0609 | 0.0169 | -3.6097 | 3.07E-04 | 6810/51445 | 9.06E-04 | -0.0605 | 0.0181 | -3.3483 | 8.13E-04 | 6810/51445 | 2.50E-03 |
| M_HDL_L | -0.0584 | 0.0161 | -3.6294 | 2.84E-04 | 6810/51445 | 4.98E-04 | -0.0361 | 0.0164 | -2.2036 | 2.76E-02 | 6810/51445 | 4.26E-02 | -0.0424 | 0.0178 | -2.3832 | 1.72E-02 | 6810/51445 | 3.34E-02 | -0.0403 | 0.0188 | -2.1395 | 3.24E-02 | 6810/51445 | 6.06E-02 |
| M_HDL_PL | -0.0368 | 0.0162 | -2.2801 | 2.26E-02 | 6810/51445 | 3.20E-02 | -0.0192 | 0.0163 | -1.1764 | 2.39E-01 | 6810/51445 | 3.17E-01 | -0.0297 | 0.0177 | -1.6754 | 9.39E-02 | 6810/51445 | 1.51E-01 | -0.0281 | 0.0187 | -1.4982 | 1.34E-01 | 6810/51445 | 2.22E-01 |
| M_HDL_C | -0.0934 | 0.0148 | -6.2906 | 3.16E-10 | 6810/51445 | 7.87E-10 | -0.0702 | 0.0151 | -4.6404 | 3.48E-06 | 6810/51445 | 8.75E-06 | -0.0680 | 0.0164 | -4.1347 | 3.55E-05 | 6810/51445 | 1.32E-04 | -0.0672 | 0.0175 | -3.8326 | 1.27E-04 | 6810/51445 | 4.86E-04 |
| M_HDL_CE | -0.0939 | 0.0149 | -6.3156 | 2.69E-10 | 6810/51445 | 6.84E-10 | -0.0710 | 0.0149 | -4.7633 | 1.90E-06 | 6810/51445 | 4.89E-06 | -0.0673 | 0.0161 | -4.1700 | 3.05E-05 | 6810/51445 | 1.15E-04 | -0.0664 | 0.0172 | -3.8594 | 1.14E-04 | 6810/51445 | 4.49E-04 |
| M_HDL_FC | -0.0869 | 0.0152 | -5.7173 | 1.08E-08 | 6810/51445 | 2.43E-08 | -0.0622 | 0.0160 | -3.8933 | 9.89E-05 | 6810/51445 | 2.22E-04 | -0.0670 | 0.0173 | -3.8718 | 1.08E-04 | 6810/51445 | 3.54E-04 | -0.0666 | 0.0185 | -3.6101 | 3.06E-04 | 6810/51445 | 1.07E-03 |
| M_HDL_TG | 0.1212 | 0.0149 | 8.1511 | 3.61E-16 | 6810/51445 | 1.55E-15 | 0.1005 | 0.0152 | 6.6305 | 3.34E-11 | 6810/51445 | 2.38E-10 | 0.0636 | 0.0164 | 3.8830 | 1.03E-04 | 6810/51445 | 3.43E-04 | 0.0594 | 0.0166 | 3.5756 | 3.49E-04 | 6810/51445 | 1.21E-03 |
| S_HDL_P | 0.0210 | 0.0150 | 1.4001 | 1.61E-01 | 6810/51445 | 2.09E-01 | 0.0102 | 0.0151 | 0.6766 | 4.99E-01 | 6810/51445 | 5.88E-01 | 0.0075 | 0.0164 | 0.4563 | 6.48E-01 | 6810/51445 | 7.44E-01 | 0.0110 | 0.0170 | 0.6440 | 5.20E-01 | 6810/51445 | 6.50E-01 |
| S_HDL_L | 0.0480 | 0.0151 | 3.1801 | 1.47E-03 | 6810/51445 | 2.34E-03 | 0.0337 | 0.0152 | 2.2131 | 2.69E-02 | 6810/51445 | 4.21E-02 | 0.0208 | 0.0165 | 1.2569 | 2.09E-01 | 6810/51445 | 2.97E-01 | 0.0226 | 0.0172 | 1.3108 | 1.90E-01 | 6810/51445 | 2.88E-01 |
| S_HDL_PL | 0.0498 | 0.0152 | 3.2834 | 1.03E-03 | 6810/51445 | 1.68E-03 | 0.0365 | 0.0153 | 2.3912 | 1.68E-02 | 6810/51445 | 2.77E-02 | 0.0209 | 0.0166 | 1.2604 | 2.08E-01 | 6810/51445 | 2.97E-01 | 0.0225 | 0.0174 | 1.2971 | 1.95E-01 | 6810/51445 | 2.93E-01 |
| S_HDL_C | 0.0049 | 0.0151 | 0.3249 | 7.45E-01 | 6810/51445 | 7.76E-01 | -0.0013 | 0.0152 | -0.0830 | 9.34E-01 | 6810/51445 | 9.45E-01 | -0.0001 | 0.0165 | -0.0070 | 9.94E-01 | 6810/51445 | 9.94E-01 | 0.0036 | 0.0172 | 0.2126 | 8.32E-01 | 6810/51445 | 9.04E-01 |

| | | | | | | | | | | | | | | | | | | | | | | | | |
|---|---|---|---|---|---|---|---|---|---|---|---|---|---|---|---|---|---|---|---|---|---|---|---|---|
| S_HDL_CE | 0.0023 | 0.0156 | 0.1456 | 8.84E-01 | 6810/51445 | 8.99E-01 | -0.0055 | 0.0157 | -0.3524 | 7.25E-01 | 6810/51445 | 7.78E-01 | 0.0014 | 0.0171 | 0.0847 | 9.33E-01 | 6810/51445 | 9.56E-01 | 0.0062 | 0.0177 | 0.3513 | 7.25E-01 | 6810/51445 | 8.25E-01 |
| S_HDL_FC | 0.0117 | 0.0154 | 0.7585 | 4.48E-01 | 6810/51445 | 5.03E-01 | 0.0105 | 0.0154 | 0.6789 | 4.97E-01 | 6810/51445 | 5.88E-01 | -0.0043 | 0.0167 | -0.2580 | 7.96E-01 | 6810/51445 | 8.51E-01 | -0.0036 | 0.0175 | -0.2082 | 8.35E-01 | 6810/51445 | 9.04E-01 |
| S_HDL_TG | 0.1540 | 0.0151 | 10.2254 | 1.53E-24 | 6810/51445 | 5.43E-23 | 0.1266 | 0.0158 | 7.9875 | 1.38E-15 | 6810/51445 | 4.90E-14 | 0.0900 | 0.0171 | 5.2562 | 1.47E-07 | 6810/51445 | 2.75E-06 | 0.0822 | 0.0173 | 4.7421 | 2.12E-06 | 6810/51445 | 3.10E-05 |
| XXL_VLDL_PL_pct | 0.0777 | 0.0149 | 5.2171 | 1.82E-07 | 6610/49808 | 4.00E-07 | 0.0613 | 0.0150 | 4.0893 | 4.33E-05 | 6610/49808 | 9.97E-05 | 0.0482 | 0.0160 | 3.0177 | 2.55E-03 | 6610/49808 | 5.93E-03 | 0.0446 | 0.0160 | 2.7822 | 5.40E-03 | 6610/49808 | 1.31E-02 |
| XXL_VLDL_C_pct | -0.0540 | 0.0147 | -3.6673 | 2.45E-04 | 6610/49808 | 4.33E-04 | -0.0396 | 0.0149 | -2.6630 | 7.75E-03 | 6610/49808 | 1.37E-02 | -0.0244 | 0.0160 | -1.5247 | 1.27E-01 | 6610/49808 | 1.93E-01 | -0.0199 | 0.0161 | -1.2319 | 2.18E-01 | 6610/49808 | 3.19E-01 |
| XXL_VLDL_CE_pct | -0.0402 | 0.0147 | -2.7419 | 6.11E-03 | 6610/49808 | 9.27E-03 | -0.0306 | 0.0148 | -2.0669 | 3.87E-02 | 6610/49808 | 5.81E-02 | -0.0168 | 0.0160 | -1.0548 | 2.91E-01 | 6610/49808 | 3.88E-01 | -0.0119 | 0.0161 | -0.7355 | 4.62E-01 | 6610/49808 | 5.87E-01 |
| XXL_VLDL_FC_pct | -0.0470 | 0.0145 | -3.2452 | 1.17E-03 | 6610/49808 | 1.90E-03 | -0.0304 | 0.0147 | -2.0634 | 3.91E-02 | 6610/49808 | 5.83E-02 | -0.0165 | 0.0159 | -1.0353 | 3.01E-01 | 6610/49808 | 3.95E-01 | -0.0147 | 0.0161 | -0.9143 | 3.61E-01 | 6610/49808 | 4.78E-01 |
| XXL_VLDL_TG_pct | 0.0224 | 0.0153 | 1.4611 | 1.44E-01 | 6610/49808 | 1.89E-01 | 0.0173 | 0.0153 | 1.1362 | 2.56E-01 | 6610/49808 | 3.37E-01 | 0.0091 | 0.0163 | 0.5539 | 5.80E-01 | 6610/49808 | 6.84E-01 | 0.0095 | 0.0164 | 0.5765 | 5.64E-01 | 6610/49808 | 6.85E-01 |
| XL_VLDL_PL_pct | 0.0536 | 0.0150 | 3.5790 | 3.45E-04 | 6749/51000 | 5.92E-04 | 0.0418 | 0.0149 | 2.8135 | 4.90E-03 | 6749/51000 | 8.97E-03 | 0.0333 | 0.0157 | 2.1190 | 3.41E-02 | 6749/51000 | 6.24E-02 | 0.0332 | 0.0159 | 2.0940 | 3.63E-02 | 6749/51000 | 6.74E-02 |
| XL_VLDL_C_pct | -0.1419 | 0.0153 | -9.2582 | 2.08E-20 | 6749/51000 | 2.73E-19 | -0.1156 | 0.0159 | -7.2571 | 3.95E-13 | 6749/51000 | 5.47E-12 | -0.0916 | 0.0172 | -5.3319 | 9.72E-08 | 6749/51000 | 2.42E-06 | -0.0836 | 0.0176 | -4.7431 | 2.10E-06 | 6749/51000 | 3.10E-05 |
| XL_VLDL_CE_pct | -0.1434 | 0.0153 | -9.3434 | 9.33E-21 | 6749/51000 | 1.55E-19 | -0.1170 | 0.0160 | -7.3344 | 2.23E-13 | 6749/51000 | 3.96E-12 | -0.0908 | 0.0172 | -5.2665 | 1.39E-07 | 6749/51000 | 2.75E-06 | -0.0819 | 0.0177 | -4.6290 | 3.67E-06 | 6749/51000 | 4.07E-05 |
| XL_VLDL_FC_pct | -0.0937 | 0.0145 | -6.4621 | 1.03E-10 | 6749/51000 | 2.68E-10 | -0.0719 | 0.0149 | -4.8231 | 1.41E-06 | 6749/51000 | 3.74E-06 | -0.0618 | 0.0160 | -3.8523 | 1.17E-04 | 6749/51000 | 3.78E-04 | -0.0577 | 0.0165 | -3.4972 | 4.70E-04 | 6749/51000 | 1.58E-03 |
| XL_VLDL_TG_pct | 0.0885 | 0.0182 | 4.8723 | 1.10E-06 | 6749/51000 | 2.27E-06 | 0.0623 | 0.0176 | 3.5384 | 4.03E-04 | 6749/51000 | 8.35E-04 | 0.0782 | 0.0184 | 4.2564 | 2.08E-05 | 6749/51000 | 8.27E-05 | 0.0744 | 0.0187 | 3.9728 | 7.10E-05 | 6749/51000 | 2.90E-04 |
| L_VLDL_PL_pct | 0.0599 | 0.0146 | 4.0970 | 4.19E-05 | 6809/51438 | 7.96E-05 | 0.0443 | 0.0144 | 3.0746 | 2.11E-03 | 6809/51438 | 4.01E-03 | 0.0378 | 0.0155 | 2.4413 | 1.46E-02 | 6809/51438 | 2.87E-02 | 0.0351 | 0.0156 | 2.2547 | 2.42E-02 | 6809/51438 | 4.89E-02 |
| L_VLDL_C_pct | -0.0482 | 0.0140 | -3.4397 | 5.82E-04 | 6809/51438 | 9.80E-04 | -0.0366 | 0.0142 | -2.5802 | 9.87E-03 | 6809/51438 | 1.70E-02 | -0.0337 | 0.0153 | -2.2032 | 2.76E-02 | 6809/51438 | 5.16E-02 | -0.0309 | 0.0158 | -1.9639 | 4.95E-02 | 6809/51438 | 9.00E-02 |
| L_VLDL_CE_pct | -0.0878 | 0.0147 | -5.9812 | 2.21E-09 | 6809/51438 | 5.30E-09 | -0.0650 | 0.0151 | -4.3151 | 1.60E-05 | 6809/51438 | 3.78E-05 | -0.0559 | 0.0163 | -3.4322 | 5.99E-04 | 6809/51438 | 1.64E-03 | -0.0516 | 0.0169 | -3.0529 | 2.27E-03 | 6809/51438 | 6.13E-03 |
| L_VLDL_FC_pct | 0.0472 | 0.0139 | 3.3980 | 6.79E-04 | 6809/51438 | 1.13E-03 | 0.0302 | 0.0140 | 2.1500 | 3.16E-02 | 6809/51438 | 4.85E-02 | 0.0206 | 0.0150 | 1.3714 | 1.70E-01 | 6809/51438 | 2.48E-01 | 0.0188 | 0.0152 | 1.2336 | 2.17E-01 | 6809/51438 | 3.19E-01 |
| L_VLDL_TG_pct | -0.0245 | 0.0143 | -1.7077 | 8.77E-02 | 6809/51438 | 1.19E-01 | -0.0188 | 0.0142 | -1.3224 | 1.86E-01 | 6809/51438 | 2.53E-01 | -0.0030 | 0.0186 | -0.1635 | 8.70E-01 | 6809/51438 | 8.99E-01 | -0.0005 | 0.0190 | -0.0239 | 9.81E-01 | 6809/51438 | 9.85E-01 |
| M_VLDL_PL_pct | -0.0540 | 0.0156 | -3.4555 | 5.49E-04 | 6810/51445 | 9.30E-04 | -0.0359 | 0.0150 | -2.3945 | 1.66E-02 | 6810/51445 | 2.76E-02 | -0.0338 | 0.0154 | -2.1989 | 2.79E-02 | 6810/51445 | 5.18E-02 | -0.0350 | 0.0160 | -2.1926 | 2.83E-02 | 6810/51445 | 5.56E-02 |
| M_VLDL_C_pct | -0.1268 | 0.0153 | -8.3003 | 1.04E-16 | 6810/51445 | 4.97E-16 | -0.0995 | 0.0159 | -6.2648 | 3.73E-10 | 6810/51445 | 1.82E-09 | -0.0823 | 0.0173 | -4.7492 | 2.04E-06 | 6810/51445 | 1.24E-05 | -0.0816 | 0.0188 | -4.3406 | 1.42E-05 | 6810/51445 | 8.22E-05 |
| M_VLDL_CE_pct | -0.0963 | 0.0144 | -6.6979 | 2.11E-11 | 6810/51445 | 5.66E-11 | -0.0731 | 0.0149 | -4.9047 | 9.36E-07 | 6810/51445 | 2.56E-06 | -0.0560 | 0.0164 | -3.4086 | 6.53E-04 | 6810/51445 | 1.73E-03 | -0.0506 | 0.0176 | -2.8792 | 3.99E-03 | 6810/51445 | 1.00E-02 |
| M_VLDL_FC_pct | -0.1059 | 0.0150 | -7.0499 | 1.79E-12 | 6810/51445 | 5.57E-12 | -0.0819 | 0.0154 | -5.3092 | 1.10E-07 | 6810/51445 | 3.43E-07 | -0.0722 | 0.0167 | -4.3102 | 1.63E-05 | 6810/51445 | 6.84E-05 | -0.0736 | 0.0182 | -4.0342 | 5.48E-05 | 6810/51445 | 2.35E-04 |
| M_VLDL_TG_pct | 0.1220 | 0.0159 | 7.6698 | 1.72E-14 | 6810/51445 | 6.13E-14 | 0.0935 | 0.0164 | 5.6842 | 1.31E-08 | 6810/51445 | 4.74E-08 | 0.0871 | 0.0152 | 5.7196 | 1.07E-08 | 6810/51445 | 5.91E-07 | 0.0883 | 0.0160 | 5.5348 | 3.12E-08 | 6810/51445 | 3.88E-06 |
| S_VLDL_PL_pct | -0.1287 | 0.0151 | -8.5012 | 1.88E-17 | 6810/51445 | 1.02E-16 | -0.1050 | 0.0155 | -6.7508 | 1.47E-11 | 6810/51445 | 1.26E-10 | -0.0825 | 0.0169 | -4.8894 | 1.01E-06 | 6810/51445 | 7.63E-06 | -0.0825 | 0.0181 | -4.5585 | 5.15E-06 | 6810/51445 | 4.51E-05 |
| S_VLDL_C_pct | -0.0948 | 0.0149 | -6.3539 | 2.10E-10 | 6810/51445 | 5.39E-10 | -0.0778 | 0.0151 | -5.1509 | 2.59E-07 | 6810/51445 | 7.34E-07 | -0.0626 | 0.0164 | -3.8134 | 1.37E-04 | 6810/51445 | 4.38E-04 | -0.0625 | 0.0177 | -3.5214 | 4.29E-04 | 6810/51445 | 1.46E-03 |
| S_VLDL_CE_pct | -0.0546 | 0.0146 | -3.7432 | 1.82E-04 | 6810/51445 | 3.28E-04 | -0.0482 | 0.0146 | -3.2997 | 9.68E-04 | 6810/51445 | 1.94E-03 | -0.0395 | 0.0159 | -2.4844 | 1.30E-02 | 6810/51445 | 2.61E-02 | -0.0379 | 0.0169 | -2.2384 | 2.52E-02 | 6810/51445 | 5.02E-02 |
| S_VLDL_FC_pct | -0.1297 | 0.0152 | -8.5406 | 1.33E-17 | 6810/51445 | 7.73E-17 | -0.1047 | 0.0157 | -6.6778 | 2.43E-11 | 6810/51445 | 1.83E-10 | -0.0827 | 0.0171 | -4.8475 | 1.25E-06 | 6810/51445 | 8.65E-06 | -0.0826 | 0.0184 | -4.4791 | 7.50E-06 | 6810/51445 | 5.40E-05 |
| S_VLDL_TG_pct | 0.1142 | 0.0150 | 7.6085 | 2.77E-14 | 6810/51445 | 9.59E-14 | 0.0954 | 0.0152 | 6.2693 | 3.63E-10 | 6810/51445 | 1.81E-09 | 0.0787 | 0.0165 | 4.7842 | 1.72E-06 | 6810/51445 | 1.07E-05 | 0.0799 | 0.0175 | 4.5551 | 5.24E-06 | 6810/51445 | 4.51E-05 |
| XS_VLDL_PL_pct | 0.1035 | 0.0157 | 6.5784 | 4.75E-11 | 6810/51445 | 1.26E-10 | 0.0772 | 0.0162 | 4.7799 | 1.75E-06 | 6810/51445 | 4.60E-06 | 0.0694 | 0.0175 | 3.9606 | 7.48E-05 | 6810/51445 | 2.55E-04 | 0.0597 | 0.0183 | 3.2590 | 1.12E-03 | 6810/51445 | 3.16E-03 |
| XS_VLDL_C_pct | -0.1354 | 0.0155 | -8.7627 | 1.91E-18 | 6810/51445 | 1.36E-17 | -0.1077 | 0.0161 | -6.7004 | 2.08E-11 | 6810/51445 | 1.67E-10 | -0.0876 | 0.0175 | -5.0018 | 5.68E-07 | 6810/51445 | 5.66E-06 | -0.0866 | 0.0193 | -4.4803 | 7.45E-06 | 6810/51445 | 5.40E-05 |
| XS_VLDL_CE_pct | -0.1352 | 0.0155 | -8.7279 | 2.59E-18 | 6810/51445 | 1.79E-17 | -0.1065 | 0.0162 | -6.5811 | 4.67E-11 | 6810/51445 | 3.14E-10 | -0.0874 | 0.0176 | -4.9571 | 7.16E-07 | 6810/51445 | 6.85E-06 | -0.0861 | 0.0195 | -4.4158 | 1.01E-05 | 6810/51445 | 6.11E-05 |
| XS_VLDL_FC_pct | -0.0946 | 0.0150 | -6.3066 | 2.85E-10 | 6810/51445 | 7.17E-10 | -0.0814 | 0.0151 | -5.3945 | 6.87E-08 | 6810/51445 | 2.25E-07 | -0.0613 | 0.0165 | -3.7179 | 2.01E-04 | 6810/51445 | 6.10E-04 | -0.0578 | 0.0175 | -3.2983 | 9.73E-04 | 6810/51445 | 2.92E-03 |
| XS_VLDL_TG_pct | 0.1385 | 0.0154 | 8.9876 | 2.53E-19 | 6810/51445 | 2.10E-18 | 0.1124 | 0.0159 | 7.0565 | 1.71E-12 | 6810/51445 | 2.13E-11 | 0.0925 | 0.0172 | 5.3628 | 8.20E-08 | 6810/51445 | 2.27E-06 | 0.0927 | 0.0187 | 4.9455 | 7.60E-07 | 6810/51445 | 2.37E-05 |
| IDL_PL_pct | 0.0341 | 0.0149 | 2.2928 | 2.19E-02 | 6810/51445 | 3.11E-02 | 0.0333 | 0.0149 | 2.2432 | 2.49E-02 | 6810/51445 | 3.97E-02 | 0.0279 | 0.0161 | 1.7340 | 8.29E-02 | 6810/51445 | 1.38E-01 | 0.0280 | 0.0165 | 1.6901 | 9.10E-02 | 6810/51445 | 1.55E-01 |
| IDL_C_pct | -0.1167 | 0.0151 | -7.7078 | 1.28E-14 | 6810/51445 | 4.69E-14 | -0.0938 | 0.0155 | -6.0573 | 1.38E-09 | 6810/51445 | 6.05E-09 | -0.0698 | 0.0170 | -4.1062 | 4.02E-05 | 6810/51445 | 1.47E-04 | -0.0632 | 0.0182 | -3.4662 | 5.28E-04 | 6810/51445 | 1.75E-03 |
| IDL_CE_pct | -0.0892 | 0.0152 | -5.8556 | 4.75E-09 | 6810/51445 | 1.10E-08 | -0.0673 | 0.0155 | -4.3392 | 1.43E-05 | 6810/51445 | 3.42E-05 | -0.0499 | 0.0170 | -2.9403 | 3.28E-03 | 6810/51445 | 7.49E-03 | -0.0438 | 0.0180 | -2.4284 | 1.52E-02 | 6810/51445 | 3.31E-02 |

| | | | | | | | | | | | | | | | | | | | | | | | | |
|---|---|---|---|---|---|---|---|---|---|---|---|---|---|---|---|---|---|---|---|---|---|---|---|---|
| IDL_FC_pct | -0.1180 | 0.0149 | -7.9194 | 2.39E-15 | 6810/51445 | 9.44E-15 | -0.1013 | 0.0151 | -6.7207 | 1.81E-11 | 6810/51445 | 1.50E-10 | -0.0747 | 0.0165 | -4.5376 | 5.69E-06 | 6810/51445 | 2.83E-05 | -0.0644 | 0.0170 | -3.7787 | 1.58E-04 | 6810/51445 | 5.95E-04 |
| IDL_TG_pct | 0.1404 | 0.0152 | 9.2137 | 3.15E-20 | 6810/51445 | 3.41E-19 | 0.1147 | 0.0158 | 7.2732 | 3.51E-13 | 6810/51445 | 5.14E-12 | 0.0880 | 0.0172 | 5.1275 | 2.94E-07 | 6810/51445 | 3.65E-06 | 0.0817 | 0.0184 | 4.4407 | 8.97E-06 | 6810/51445 | 5.69E-05 |
| L_LDL_PL_pct | -0.0108 | 0.0150 | -0.7193 | 4.72E-01 | 6810/51445 | 5.22E-01 | -0.0246 | 0.0151 | -1.6303 | 1.03E-01 | 6810/51445 | 1.47E-01 | -0.0139 | 0.0163 | -0.8505 | 3.95E-01 | 6810/51445 | 4.89E-01 | -0.0105 | 0.0164 | -0.6387 | 5.23E-01 | 6810/51445 | 6.51E-01 |
| L_LDL_C_pct | -0.0164 | 0.0114 | -1.4327 | 1.52E-01 | 6810/51445 | 1.98E-01 | -0.0133 | 0.0106 | -1.2553 | 2.09E-01 | 6810/51445 | 2.82E-01 | -0.0116 | 0.0102 | -1.1408 | 2.54E-01 | 6810/51445 | 3.49E-01 | -0.0079 | 0.0106 | -0.7490 | 4.54E-01 | 6810/51445 | 5.80E-01 |
| L_LDL_CE_pct | -0.0109 | 0.0112 | -0.9675 | 3.33E-01 | 6810/51445 | 3.90E-01 | -0.0091 | 0.0112 | -0.8154 | 4.15E-01 | 6810/51445 | 5.09E-01 | -0.0087 | 0.0114 | -0.7620 | 4.46E-01 | 6810/51445 | 5.42E-01 | -0.0044 | 0.0120 | -0.3665 | 7.14E-01 | 6810/51445 | 8.19E-01 |
| L_LDL_FC_pct | -0.0817 | 0.0252 | -3.2452 | 1.17E-03 | 6810/51445 | 1.90E-03 | -0.0469 | 0.0259 | -1.8150 | 6.95E-02 | 6810/51445 | 1.02E-01 | -0.0222 | 0.0137 | -1.6152 | 1.06E-01 | 6810/51445 | 1.67E-01 | -0.0170 | 0.0128 | -1.3337 | 1.82E-01 | 6810/51445 | 2.79E-01 |
| L_LDL_TG_pct | 0.1252 | 0.0151 | 8.2788 | 1.24E-16 | 6810/51445 | 5.63E-16 | 0.1003 | 0.0156 | 6.4358 | 1.23E-10 | 6810/51445 | 6.95E-10 | 0.0710 | 0.0170 | 4.1800 | 2.92E-05 | 6810/51445 | 1.12E-04 | 0.0591 | 0.0180 | 3.2774 | 1.05E-03 | 6810/51445 | 3.11E-03 |
| M_LDL_PL_pct | -0.0104 | 0.0091 | -1.1394 | 2.55E-01 | 6810/51445 | 3.12E-01 | -0.0077 | 0.0089 | -0.8664 | 3.86E-01 | 6810/51445 | 4.81E-01 | 0.0004 | 0.0114 | 0.0312 | 9.75E-01 | 6810/51445 | 9.86E-01 | -0.0013 | 0.0105 | -0.1197 | 9.05E-01 | 6810/51445 | 9.39E-01 |
| M_LDL_C_pct | -0.0107 | 0.0092 | -1.1575 | 2.47E-01 | 6810/51445 | 3.05E-01 | -0.0094 | 0.0091 | -1.0370 | 3.00E-01 | 6810/51445 | 3.83E-01 | -0.0091 | 0.0090 | -1.0022 | 3.16E-01 | 6810/51445 | 4.06E-01 | -0.0056 | 0.0094 | -0.5965 | 5.51E-01 | 6810/51445 | 6.72E-01 |
| M_LDL_CE_pct | 0.0010 | 0.0107 | 0.0886 | 9.29E-01 | 6810/51445 | 9.36E-01 | -0.0014 | 0.0097 | -0.1451 | 8.85E-01 | 6810/51445 | 9.06E-01 | -0.0046 | 0.0092 | -0.5051 | 6.13E-01 | 6810/51445 | 7.11E-01 | -0.0014 | 0.0096 | -0.1428 | 8.86E-01 | 6810/51445 | 9.31E-01 |
| M_LDL_FC_pct | -0.1197 | 0.0241 | -4.9734 | 6.58E-07 | 6810/51445 | 1.40E-06 | -0.0792 | 0.0215 | -3.6898 | 2.24E-04 | 6810/51445 | 4.78E-04 | -0.0312 | 0.0176 | -1.7761 | 7.57E-02 | 6810/51445 | 1.30E-01 | -0.0233 | 0.0150 | -1.5458 | 1.22E-01 | 6810/51445 | 2.04E-01 |
| M_LDL_TG_pct | 0.1042 | 0.0149 | 6.9697 | 3.18E-12 | 6810/51445 | 9.65E-12 | 0.0850 | 0.0152 | 5.5956 | 2.20E-08 | 6810/51445 | 7.50E-08 | 0.0580 | 0.0166 | 3.5018 | 4.62E-04 | 6810/51445 | 1.29E-03 | 0.0476 | 0.0175 | 2.7222 | 6.48E-03 | 6810/51445 | 1.54E-02 |
| S_LDL_PL_pct | -0.0723 | 0.0147 | -4.9047 | 9.36E-07 | 6810/51445 | 1.96E-06 | -0.0530 | 0.0150 | -3.5430 | 3.96E-04 | 6810/51445 | 8.28E-04 | -0.0303 | 0.0162 | -1.8683 | 6.17E-02 | 6810/51445 | 1.08E-01 | -0.0406 | 0.0166 | -2.4475 | 1.44E-02 | 6810/51445 | 3.17E-02 |
| S_LDL_C_pct | -0.0094 | 0.0090 | -1.0468 | 2.95E-01 | 6810/51445 | 3.48E-01 | -0.0086 | 0.0090 | -0.9609 | 3.37E-01 | 6810/51445 | 4.23E-01 | -0.0083 | 0.0089 | -0.9313 | 3.52E-01 | 6810/51445 | 4.46E-01 | -0.0047 | 0.0094 | -0.4974 | 6.19E-01 | 6810/51445 | 7.45E-01 |
| S_LDL_CE_pct | 0.0051 | 0.0136 | 0.3702 | 7.11E-01 | 6810/51445 | 7.44E-01 | 0.0012 | 0.0111 | 0.1038 | 9.17E-01 | 6810/51445 | 9.32E-01 | -0.0036 | 0.0097 | -0.3725 | 7.09E-01 | 6810/51445 | 7.83E-01 | 0.0002 | 0.0104 | 0.0179 | 9.86E-01 | 6810/51445 | 9.86E-01 |
| S_LDL_FC_pct | -0.1032 | 0.0203 | -5.0972 | 3.45E-07 | 6810/51445 | 7.40E-07 | -0.0722 | 0.0217 | -3.3351 | 8.53E-04 | 6810/51445 | 1.73E-03 | -0.0250 | 0.0139 | -1.7986 | 7.21E-02 | 6810/51445 | 1.25E-01 | -0.0188 | 0.0131 | -1.4341 | 1.52E-01 | 6810/51445 | 2.43E-01 |
| S_LDL_TG_pct | 0.1384 | 0.0151 | 9.1505 | 5.67E-20 | 6810/51445 | 5.68E-19 | 0.1138 | 0.0156 | 7.2936 | 3.02E-13 | 6810/51445 | 4.70E-12 | 0.0805 | 0.0170 | 4.7317 | 2.23E-06 | 6810/51445 | 1.29E-05 | 0.0733 | 0.0176 | 4.1736 | 3.00E-05 | 6810/51445 | 1.41E-04 |
| XL_HDL_PL_pct | -0.0190 | 0.0136 | -1.3959 | 1.63E-01 | 6808/51433 | 2.10E-01 | -0.0071 | 0.0144 | -0.4970 | 6.19E-01 | 6808/51433 | 6.91E-01 | -0.0086 | 0.0152 | -0.5663 | 5.71E-01 | 6808/51433 | 6.77E-01 | -0.0074 | 0.0154 | -0.4835 | 6.29E-01 | 6808/51433 | 7.53E-01 |
| XL_HDL_C_pct | -0.0026 | 0.0159 | -0.1615 | 8.72E-01 | 6808/51433 | 8.90E-01 | -0.0216 | 0.0158 | -1.3646 | 1.72E-01 | 6808/51433 | 2.37E-01 | -0.0048 | 0.0174 | -0.2752 | 7.83E-01 | 6808/51433 | 8.41E-01 | -0.0025 | 0.0175 | -0.1448 | 8.85E-01 | 6808/51433 | 9.31E-01 |
| XL_HDL_CE_pct | -0.0390 | 0.0117 | -3.3221 | 8.93E-04 | 6808/51433 | 1.47E-03 | -0.0336 | 0.0119 | -2.8238 | 4.75E-03 | 6808/51433 | 8.82E-03 | -0.0253 | 0.0133 | -1.9064 | 5.66E-02 | 6808/51433 | 9.99E-02 | -0.0199 | 0.0136 | -1.4589 | 1.45E-01 | 6808/51433 | 2.35E-01 |
| XL_HDL_FC_pct | 0.1117 | 0.0163 | 6.8576 | 7.00E-12 | 6808/51433 | 2.00E-11 | 0.0722 | 0.0174 | 4.1486 | 3.34E-05 | 6808/51433 | 7.86E-05 | 0.0631 | 0.0189 | 3.3448 | 8.24E-04 | 6808/51433 | 2.16E-03 | 0.0585 | 0.0195 | 3.0057 | 2.65E-03 | 6808/51433 | 7.00E-03 |
| XL_HDL_TG_pct | 0.1562 | 0.0154 | 10.1215 | 4.44E-24 | 6808/51433 | 1.23E-22 | 0.1266 | 0.0164 | 7.7289 | 1.09E-14 | 6808/51433 | 3.00E-13 | 0.0933 | 0.0178 | 5.2471 | 1.55E-07 | 6808/51433 | 2.75E-06 | 0.0868 | 0.0181 | 4.7843 | 1.72E-06 | 6808/51433 | 2.95E-05 |
| L_HDL_PL_pct | 0.2159 | 0.0243 | 8.8859 | 6.34E-19 | 6810/51445 | 5.09E-18 | 0.1620 | 0.0261 | 6.2074 | 5.39E-10 | 6810/51445 | 2.58E-09 | 0.1347 | 0.0285 | 4.7313 | 2.23E-06 | 6810/51445 | 1.29E-05 | 0.1309 | 0.0305 | 4.2874 | 1.81E-05 | 6810/51445 | 9.58E-05 |
| L_HDL_C_pct | -0.0757 | 0.0117 | -6.4959 | 8.25E-11 | 6810/51445 | 2.16E-10 | -0.0589 | 0.0121 | -4.8589 | 1.18E-06 | 6810/51445 | 3.16E-06 | -0.0455 | 0.0141 | -3.2311 | 1.23E-03 | 6810/51445 | 3.13E-03 | -0.0390 | 0.0145 | -2.6887 | 7.17E-03 | 6810/51445 | 1.67E-02 |
| L_HDL_CE_pct | -0.0827 | 0.0123 | -6.7052 | 2.01E-11 | 6810/51445 | 5.44E-11 | -0.0642 | 0.0128 | -5.0281 | 4.95E-07 | 6810/51445 | 1.37E-06 | -0.0460 | 0.0149 | -3.0801 | 2.07E-03 | 6810/51445 | 4.95E-03 | -0.0380 | 0.0154 | -2.4605 | 1.39E-02 | 6810/51445 | 3.08E-02 |
| L_HDL_FC_pct | -0.0482 | 0.0130 | -3.7197 | 1.99E-04 | 6810/51445 | 3.57E-04 | -0.0363 | 0.0136 | -2.6727 | 7.53E-03 | 6810/51445 | 1.34E-02 | -0.0363 | 0.0146 | -2.4773 | 1.32E-02 | 6810/51445 | 2.64E-02 | -0.0343 | 0.0150 | -2.2861 | 2.22E-02 | 6810/51445 | 4.62E-02 |
| L_HDL_TG_pct | 0.1523 | 0.0155 | 9.8064 | 1.06E-22 | 6810/51445 | 2.63E-21 | 0.1238 | 0.0163 | 7.6133 | 2.67E-14 | 6810/51445 | 6.65E-13 | 0.0868 | 0.0176 | 4.9239 | 8.48E-07 | 6810/51445 | 7.28E-06 | 0.0799 | 0.0179 | 4.4500 | 8.59E-06 | 6810/51445 | 5.63E-05 |
| M_HDL_PL_pct | 0.2273 | 0.0194 | 11.6895 | 1.44E-31 | 6810/51445 | 3.59E-29 | 0.1922 | 0.0210 | 9.1614 | 5.12E-20 | 6810/51445 | 1.28E-17 | 0.1520 | 0.0229 | 6.6414 | 3.11E-11 | 6810/51445 | 7.74E-09 | 0.1366 | 0.0235 | 5.8070 | 6.36E-09 | 6810/51445 | 1.58E-06 |
| M_HDL_C_pct | -0.1652 | 0.0199 | -8.3184 | 8.91E-17 | 6810/51445 | 4.35E-16 | -0.1222 | 0.0211 | -5.7923 | 6.94E-09 | 6810/51445 | 2.66E-08 | -0.0649 | 0.0183 | -3.5409 | 3.99E-04 | 6810/51445 | 1.13E-03 | -0.0585 | 0.0157 | -3.7219 | 1.98E-04 | 6810/51445 | 7.35E-04 |
| M_HDL_CE_pct | -0.1493 | 0.0195 | -7.6487 | 2.03E-14 | 6810/51445 | 7.12E-14 | -0.1097 | 0.0204 | -5.3827 | 7.34E-08 | 6810/51445 | 2.37E-07 | -0.0572 | 0.0158 | -3.6074 | 3.09E-04 | 6810/51445 | 9.06E-04 | -0.0527 | 0.0145 | -3.6326 | 2.81E-04 | 6810/51445 | 9.98E-04 |
| M_HDL_FC_pct | -0.1242 | 0.0185 | -6.7066 | 1.99E-11 | 6810/51445 | 5.44E-11 | -0.0835 | 0.0183 | -4.5727 | 4.82E-06 | 6810/51445 | 1.20E-05 | -0.0747 | 0.0175 | -4.2693 | 1.96E-05 | 6810/51445 | 8.00E-05 | -0.0731 | 0.0168 | -4.3532 | 1.34E-05 | 6810/51445 | 7.95E-05 |
| M_HDL_TG_pct | 0.1565 | 0.0153 | 10.2516 | 1.16E-24 | 6810/51445 | 4.83E-23 | 0.1299 | 0.0159 | 8.1483 | 3.69E-16 | 6810/51445 | 1.53E-14 | 0.0931 | 0.0172 | 5.4059 | 6.45E-08 | 6810/51445 | 2.01E-06 | 0.0859 | 0.0175 | 4.9109 | 9.07E-07 | 6810/51445 | 2.37E-05 |
| S_HDL_PL_pct | 0.0173 | 0.0154 | 1.1238 | 2.61E-01 | 6810/51445 | 3.17E-01 | 0.0230 | 0.0154 | 1.4918 | 1.36E-01 | 6810/51445 | 1.90E-01 | 0.0035 | 0.0167 | 0.2091 | 8.34E-01 | 6810/51445 | 8.73E-01 | 0.0013 | 0.0171 | 0.0782 | 9.38E-01 | 6810/51445 | 9.57E-01 |
| S_HDL_C_pct | -0.1181 | 0.0149 | -7.9056 | 2.67E-15 | 6810/51445 | 1.04E-14 | -0.0977 | 0.0152 | -6.4180 | 1.38E-10 | 6810/51445 | 7.64E-10 | -0.0591 | 0.0167 | -3.5452 | 3.92E-04 | 6810/51445 | 1.12E-03 | -0.0503 | 0.0169 | -2.9784 | 2.90E-03 | 6810/51445 | 7.36E-03 |
| S_HDL_CE_pct | -0.1037 | 0.0170 | -6.0903 | 1.13E-09 | 6810/51445 | 2.73E-09 | -0.0892 | 0.0171 | -5.2036 | 1.95E-07 | 6810/51445 | 5.79E-07 | -0.0444 | 0.0189 | -2.3568 | 1.84E-02 | 6810/51445 | 3.56E-02 | -0.0346 | 0.0190 | -1.8174 | 6.91E-02 | 6810/51445 | 1.23E-01 |
| S_HDL_FC_pct | -0.0947 | 0.0153 | -6.1710 | 6.79E-10 | 6810/51445 | 1.66E-09 | -0.0640 | 0.0159 | -4.0235 | 5.73E-05 | 6810/51445 | 1.30E-04 | -0.0704 | 0.0174 | -4.0476 | 5.17E-05 | 6810/51445 | 1.84E-04 | -0.0728 | 0.0180 | -4.0341 | 5.48E-05 | 6810/51445 | 2.35E-04 |

| | | | | | | | | | | | | | | | | | | | | | | | | |
|---|---|---|---|---|---|---|---|---|---|---|---|---|---|---|---|---|---|---|---|---|---|---|---|---|
| S_HDL_TG_pct | 0.1594 | 0.0153 | 10.4061 | 2.33E-25 | 6810/51445 | 1.16E-23 | 0.1319 | 0.0161 | 8.1933 | 2.54E-16 | 6810/51445 | 1.27E-14 | 0.0954 | 0.0174 | 5.4853 | 4.13E-08 | 6810/51445 | 1.47E-06 | 0.0871 | 0.0177 | 4.9156 | 8.85E-07 | 6810/51445 | 2.37E-05 |

**Supplementary Table 2**
**Results of sensitivity analysis**

| | mdd | | | | | | recurrent | | | | | |
|---|---|---|---|---|---|---|---|---|---|---|---|---|
| metabolite | beta | se | z | pvalue | n | fdr | beta | se | z | pvalue | n | fdr |
| Total_C | -0.0399 | 0.0185 | -2.1550 | 3.12E-02 | 5498/51404 | 5.88E-02 | -0.0444 | 0.0219 | -2.0252 | 4.28E-02 | 3491/61782 | 8.75E-02 |
| non_HDL_C | -0.0110 | 0.0178 | -0.6167 | 5.37E-01 | 5498/51404 | 6.66E-01 | -0.0160 | 0.0211 | -0.7579 | 4.49E-01 | 3491/61782 | 5.76E-01 |
| Remnant_C | -0.0047 | 0.0179 | -0.2633 | 7.92E-01 | 5498/51404 | 8.77E-01 | -0.0077 | 0.0212 | -0.3649 | 7.15E-01 | 3491/61782 | 7.85E-01 |
| VLDL_C | 0.0325 | 0.0172 | 1.8847 | 5.95E-02 | 5498/51404 | 1.05E-01 | 0.0317 | 0.0204 | 1.5497 | 1.21E-01 | 3491/61782 | 2.10E-01 |
| Clinical_LDL_C | -0.0230 | 0.0178 | -1.2916 | 1.97E-01 | 5498/51404 | 2.95E-01 | -0.0302 | 0.0210 | -1.4414 | 1.49E-01 | 3491/61782 | 2.40E-01 |
| LDL_C | -0.0122 | 0.0133 | -0.9194 | 3.58E-01 | 5498/51404 | 4.77E-01 | -0.0180 | 0.0140 | -1.2801 | 2.00E-01 | 3491/61782 | 2.99E-01 |
| HDL_C | -0.0926 | 0.0200 | -4.6165 | 3.90E-06 | 5498/51404 | 2.94E-05 | -0.0974 | 0.0240 | -4.0644 | 4.82E-05 | 3491/61782 | 7.07E-04 |
| Total_TG | 0.0766 | 0.0175 | 4.3792 | 1.19E-05 | 5498/51404 | 6.31E-05 | 0.0744 | 0.0207 | 3.5906 | 3.30E-04 | 3491/61782 | 1.61E-03 |
| VLDL_TG | 0.0824 | 0.0177 | 4.6599 | 3.16E-06 | 5498/51404 | 2.89E-05 | 0.0797 | 0.0210 | 3.7989 | 1.45E-04 | 3491/61782 | 1.09E-03 |
| LDL_TG | 0.0382 | 0.0171 | 2.2415 | 2.50E-02 | 5498/51404 | 4.94E-02 | 0.0406 | 0.0201 | 2.0160 | 4.38E-02 | 3491/61782 | 8.87E-02 |
| HDL_TG | 0.0556 | 0.0166 | 3.3430 | 8.29E-04 | 5498/51404 | 2.43E-03 | 0.0513 | 0.0197 | 2.6098 | 9.06E-03 | 3491/61782 | 2.48E-02 |
| Total_PL | -0.0266 | 0.0183 | -1.4508 | 1.47E-01 | 5498/51404 | 2.36E-01 | -0.0301 | 0.0217 | -1.3882 | 1.65E-01 | 3491/61782 | 2.60E-01 |
| VLDL_PL | 0.0541 | 0.0172 | 3.1359 | 1.71E-03 | 5498/51404 | 4.59E-03 | 0.0524 | 0.0204 | 2.5670 | 1.03E-02 | 3491/61782 | 2.72E-02 |
| LDL_PL | -0.0160 | 0.0175 | -0.9112 | 3.62E-01 | 5498/51404 | 4.77E-01 | -0.0221 | 0.0208 | -1.0640 | 2.87E-01 | 3491/61782 | 4.07E-01 |
| HDL_PL | -0.0575 | 0.0193 | -2.9822 | 2.86E-03 | 5498/51404 | 6.99E-03 | -0.0627 | 0.0230 | -2.7228 | 6.47E-03 | 3491/61782 | 1.83E-02 |
| Total_CE | -0.0450 | 0.0186 | -2.4158 | 1.57E-02 | 5498/51404 | 3.31E-02 | -0.0501 | 0.0220 | -2.2732 | 2.30E-02 | 3491/61782 | 5.21E-02 |
| VLDL_CE | 0.0228 | 0.0173 | 1.3139 | 1.89E-01 | 5498/51404 | 2.85E-01 | 0.0222 | 0.0206 | 1.0814 | 2.80E-01 | 3491/61782 | 4.07E-01 |
| LDL_CE | -0.0085 | 0.0131 | -0.6485 | 5.17E-01 | 5498/51404 | 6.50E-01 | -0.0148 | 0.0138 | -1.0710 | 2.84E-01 | 3491/61782 | 4.07E-01 |
| HDL_CE | -0.0929 | 0.0200 | -4.6400 | 3.48E-06 | 5498/51404 | 2.89E-05 | -0.0987 | 0.0239 | -4.1224 | 3.75E-05 | 3491/61782 | 6.67E-04 |
| Total_FC | -0.0267 | 0.0183 | -1.4615 | 1.44E-01 | 5498/51404 | 2.33E-01 | -0.0298 | 0.0217 | -1.3738 | 1.70E-01 | 3491/61782 | 2.65E-01 |
| VLDL_FC | 0.0453 | 0.0172 | 2.6292 | 8.56E-03 | 5498/51404 | 1.95E-02 | 0.0443 | 0.0204 | 2.1682 | 3.01E-02 | 3491/61782 | 6.42E-02 |
| LDL_FC | -0.0228 | 0.0145 | -1.5733 | 1.16E-01 | 5498/51404 | 1.92E-01 | -0.0268 | 0.0152 | -1.7692 | 7.69E-02 | 3491/61782 | 1.47E-01 |
| HDL_FC | -0.0862 | 0.0199 | -4.3334 | 1.47E-05 | 5498/51404 | 7.03E-05 | -0.0885 | 0.0237 | -3.7284 | 1.93E-04 | 3491/61782 | 1.16E-03 |
| Total_L | -0.0061 | 0.0177 | -0.3454 | 7.30E-01 | 5498/51404 | 8.37E-01 | -0.0101 | 0.0209 | -0.4811 | 6.30E-01 | 3491/61782 | 7.32E-01 |
| VLDL_L | 0.0625 | 0.0174 | 3.5965 | 3.23E-04 | 5498/51404 | 1.10E-03 | 0.0601 | 0.0206 | 2.9194 | 3.51E-03 | 3491/61782 | 1.15E-02 |
| LDL_L | -0.0132 | 0.0175 | -0.7532 | 4.51E-01 | 5498/51404 | 5.79E-01 | -0.0195 | 0.0207 | -0.9383 | 3.48E-01 | 3491/61782 | 4.66E-01 |
| HDL_L | -0.0697 | 0.0195 | -3.5655 | 3.63E-04 | 5498/51404 | 1.22E-03 | -0.0745 | 0.0233 | -3.1936 | 1.41E-03 | 3491/61782 | 4.94E-03 |
| Total_P | -0.0438 | 0.0186 | -2.3523 | 1.87E-02 | 5498/51404 | 3.81E-02 | -0.0528 | 0.0221 | -2.3869 | 1.70E-02 | 3491/61782 | 4.03E-02 |
| VLDL_P | 0.0402 | 0.0171 | 2.3537 | 1.86E-02 | 5498/51404 | 3.81E-02 | 0.0398 | 0.0202 | 1.9665 | 4.92E-02 | 3491/61782 | 9.81E-02 |
| LDL_P | -0.0021 | 0.0175 | -0.1187 | 9.05E-01 | 5498/51404 | 9.45E-01 | -0.0045 | 0.0208 | -0.2140 | 8.31E-01 | 3491/61782 | 8.65E-01 |
| HDL_P | -0.0461 | 0.0187 | -2.4631 | 1.38E-02 | 5498/51404 | 2.98E-02 | -0.0561 | 0.0222 | -2.5211 | 1.17E-02 | 3491/61782 | 3.03E-02 |
| VLDL_size | 0.0921 | 0.0184 | 5.0037 | 5.62E-07 | 5498/51404 | 1.50E-05 | 0.0866 | 0.0219 | 3.9588 | 7.53E-05 | 3491/61782 | 8.96E-04 |
| LDL_size | -0.0032 | 0.0173 | -0.1858 | 8.53E-01 | 5498/51404 | 9.19E-01 | -0.0123 | 0.0206 | -0.5982 | 5.50E-01 | 3491/61782 | 6.64E-01 |
| HDL_size | -0.0956 | 0.0196 | -4.8834 | 1.04E-06 | 5498/51404 | 1.73E-05 | -0.0892 | 0.0234 | -3.8128 | 1.37E-04 | 3491/61782 | 1.09E-03 |
| Phosphoglyc | -0.0195 | 0.0182 | -1.0709 | 2.84E-01 | 5495/51357 | 3.94E-01 | -0.0251 | 0.0215 | -1.1662 | 2.44E-01 | 3489/61726 | 3.59E-01 |
| TG_by_PG | 0.1022 | 0.0187 | 5.4688 | 4.53E-08 | 5495/51357 | 4.43E-06 | 0.1029 | 0.0223 | 4.6224 | 3.79E-06 | 3489/61726 | 3.80E-04 |
| Cholines | -0.0291 | 0.0185 | -1.5745 | 1.15E-01 | 5495/51357 | 1.92E-01 | -0.0348 | 0.0219 | -1.5880 | 1.12E-01 | 3489/61726 | 1.97E-01 |
| Phosphatidylc | -0.0194 | 0.0184 | -1.0540 | 2.92E-01 | 5495/51357 | 3.99E-01 | -0.0246 | 0.0218 | -1.1263 | 2.60E-01 | 3489/61726 | 3.81E-01 |
| Sphingomyelins | -0.0636 | 0.0189 | -3.3583 | 7.84E-04 | 5495/51357 | 2.32E-03 | -0.0720 | 0.0225 | -3.2056 | 1.35E-03 | 3489/61726 | 4.86E-03 |
| ApoB | -0.0016 | 0.0176 | -0.0890 | 9.29E-01 | 5498/51404 | 9.56E-01 | -0.0041 | 0.0209 | -0.1962 | 8.44E-01 | 3491/61782 | 8.76E-01 |
| ApoA1 | -0.0575 | 0.0191 | -3.0026 | 2.68E-03 | 5498/51404 | 6.67E-03 | -0.0645 | 0.0228 | -2.8255 | 4.72E-03 | 3491/61782 | 1.45E-02 |
| ApoB_by_ApoA1 | 0.0278 | 0.0178 | 1.5631 | 1.18E-01 | 5498/51404 | 1.95E-01 | 0.0290 | 0.0212 | 1.3718 | 1.70E-01 | 3491/61782 | 2.65E-01 |
| Total_FA | 0.0238 | 0.0172 | 1.3832 | 1.67E-01 | 5495/51357 | 2.64E-01 | 0.0191 | 0.0204 | 0.9352 | 3.50E-01 | 3489/61726 | 4.66E-01 |
| Unsaturation | -0.0580 | 0.0185 | -3.1438 | 1.67E-03 | 5495/51357 | 4.51E-03 | -0.0506 | 0.0220 | -2.3011 | 2.14E-02 | 3489/61726 | 4.93E-02 |
| Omega_3 | 0.0074 | 0.0171 | 0.4347 | 6.64E-01 | 5495/51357 | 7.76E-01 | 0.0134 | 0.0203 | 0.6590 | 5.10E-01 | 3489/61726 | 6.25E-01 |
| Omega_6 | -0.0055 | 0.0175 | -0.3154 | 7.52E-01 | 5495/51357 | 8.52E-01 | -0.0113 | 0.0207 | -0.5481 | 5.84E-01 | 3489/61726 | 6.95E-01 |
| PUFA | -0.0018 | 0.0173 | -0.1012 | 9.19E-01 | 5495/51357 | 9.50E-01 | -0.0056 | 0.0205 | -0.2731 | 7.85E-01 | 3489/61726 | 8.32E-01 |
| MUFA | 0.0534 | 0.0175 | 3.0556 | 2.25E-03 | 5495/51357 | 5.77E-03 | 0.0468 | 0.0207 | 2.2663 | 2.34E-02 | 3489/61726 | 5.26E-02 |
| SFA | 0.0197 | 0.0175 | 1.1280 | 2.59E-01 | 5495/51357 | 3.63E-01 | 0.0148 | 0.0207 | 0.7136 | 4.75E-01 | 3489/61726 | 5.98E-01 |
| LA | 0.0003 | 0.0176 | 0.0167 | 9.87E-01 | 5495/51357 | 9.87E-01 | -0.0063 | 0.0208 | -0.3010 | 7.63E-01 | 3489/61726 | 8.19E-01 |
| DHA | -0.0168 | 0.0179 | -0.9416 | 3.46E-01 | 5495/51357 | 4.69E-01 | -0.0158 | 0.0213 | -0.7401 | 4.59E-01 | 3489/61726 | 5.86E-01 |
| Omega_3_pct | -0.0044 | 0.0174 | -0.2528 | 8.00E-01 | 5495/51357 | 8.82E-01 | 0.0057 | 0.0207 | 0.2748 | 7.83E-01 | 3489/61726 | 8.32E-01 |
| Omega_6_pct | -0.0629 | 0.0183 | -3.4420 | 5.77E-04 | 5495/51357 | 1.75E-03 | -0.0623 | 0.0216 | -2.8797 | 3.98E-03 | 3489/61726 | 1.27E-02 |
| PUFA_pct | -0.0647 | 0.0188 | -3.4456 | 5.70E-04 | 5495/51357 | 1.75E-03 | -0.0608 | 0.0223 | -2.7302 | 6.33E-03 | 3489/61726 | 1.81E-02 |
| MUFA_pct | 0.1014 | 0.0188 | 5.3881 | 7.12E-08 | 5495/51357 | 4.43E-06 | 0.0943 | 0.0224 | 4.2016 | 2.65E-05 | 3489/61726 | 5.32E-04 |
| SFA_pct | -0.0068 | 0.0175 | -0.3862 | 6.99E-01 | 5495/51357 | 8.14E-01 | -0.0095 | 0.0208 | -0.4584 | 6.47E-01 | 3489/61726 | 7.39E-01 |
| LA_pct | -0.0464 | 0.0190 | -2.4355 | 1.49E-02 | 5495/51357 | 3.16E-02 | -0.0500 | 0.0226 | -2.2142 | 2.68E-02 | 3489/61726 | 5.86E-02 |
| DHA_pct | -0.0335 | 0.0184 | -1.8267 | 6.78E-02 | 5495/51357 | 1.17E-01 | -0.0295 | 0.0219 | -1.3466 | 1.78E-01 | 3489/61726 | 2.72E-01 |
| PUFA_by_MUFA | -0.0898 | 0.0190 | -4.7345 | 2.20E-06 | 5495/51357 | 2.39E-05 | -0.0836 | 0.0225 | -3.7095 | 2.08E-04 | 3489/61726 | 1.16E-03 |
| Omega_6_by_Omega_3 | -0.0106 | 0.0172 | -0.6168 | 5.37E-01 | 5495/51357 | 6.66E-01 | -0.0197 | 0.0205 | -0.9634 | 3.35E-01 | 3489/61726 | 4.56E-01 |
| Ala | 0.0554 | 0.0200 | 2.7671 | 5.66E-03 | 5498/51402 | 1.35E-02 | 0.0386 | 0.0238 | 1.6215 | 1.05E-01 | 3491/61779 | 1.87E-01 |
| Gln | 0.0118 | 0.0170 | 0.6974 | 4.86E-01 | 5483/51267 | 6.17E-01 | 0.0133 | 0.0202 | 0.6618 | 5.08E-01 | 3481/61613 | 6.25E-01 |
| Gly | -0.0162 | 0.0153 | -1.0633 | 2.88E-01 | 5495/51355 | 3.96E-01 | -0.0246 | 0.0181 | -1.3649 | 1.72E-01 | 3491/61723 | 2.66E-01 |
| His | -0.0047 | 0.0168 | -0.2764 | 7.82E-01 | 5492/51305 | 8.73E-01 | -0.0269 | 0.0202 | -1.3282 | 1.84E-01 | 3489/61670 | 2.78E-01 |
| Total_BCAA | 0.0215 | 0.0179 | 1.1994 | 2.30E-01 | 5497/51392 | 3.32E-01 | -0.0005 | 0.0214 | -0.0255 | 9.80E-01 | 3490/61768 | 9.88E-01 |
| Ile | 0.0207 | 0.0171 | 1.2080 | 2.27E-01 | 5498/51403 | 3.29E-01 | -0.0091 | 0.0205 | -0.4428 | 6.58E-01 | 3491/61780 | 7.48E-01 |
| Leu | 0.0154 | 0.0178 | 0.8631 | 3.88E-01 | 5498/51402 | 5.03E-01 | -0.0040 | 0.0213 | -0.1887 | 8.50E-01 | 3491/61779 | 8.79E-01 |
| Val | 0.0242 | 0.0182 | 1.3309 | 1.83E-01 | 5497/51393 | 2.80E-01 | 0.0058 | 0.0216 | 0.2679 | 7.89E-01 | 3490/61769 | 8.32E-01 |
| Phe | -0.0349 | 0.0167 | -2.0911 | 3.65E-02 | 5497/51370 | 6.79E-02 | -0.0483 | 0.0198 | -2.4314 | 1.50E-02 | 3490/61741 | 3.64E-02 |
| Tyr | -0.0056 | 0.0170 | -0.3294 | 7.42E-01 | 5491/51316 | 8.44E-01 | -0.0203 | 0.0203 | -1.0012 | 3.17E-01 | 3488/61682 | 4.36E-01 |
| Glucose | -0.0397 | 0.0183 | -2.1688 | 3.01E-02 | 5490/51296 | 5.76E-02 | -0.0341 | 0.0214 | -1.5918 | 1.11E-01 | 3488/61656 | 1.97E-01 |

| | | | | | | | | | | | | |
|---|---|---|---|---|---|---|---|---|---|---|---|---|
| Lactate | 0.0137 | 0.0163 | 0.8446 | 3.98E-01 | 5486/51267 | 5.14E-01 | 0.0046 | 0.0192 | 0.2414 | 8.09E-01 | 3485/61616 | 8.49E-01 |
| Pyruvate | 0.0388 | 0.0148 | 2.6233 | 8.71E-03 | 5483/51268 | 1.97E-02 | 0.0292 | 0.0174 | 1.6759 | 9.37E-02 | 3481/61614 | 1.69E-01 |
| Citrate | -0.0682 | 0.0171 | -3.9982 | 6.38E-05 | 5498/51394 | 2.52E-04 | -0.0697 | 0.0202 | -3.4508 | 5.59E-04 | 3491/61769 | 2.25E-03 |
| bOHbutyrate | -0.0090 | 0.0167 | -0.5386 | 5.90E-01 | 5433/50823 | 7.03E-01 | -0.0144 | 0.0200 | -0.7186 | 4.72E-01 | 3452/61088 | 5.97E-01 |
| Acetate | -0.0062 | 0.0180 | -0.3476 | 7.28E-01 | 5496/51392 | 8.37E-01 | -0.0091 | 0.0218 | -0.4188 | 6.75E-01 | 3490/61765 | 7.54E-01 |
| Acetoacetate | -0.0085 | 0.0153 | -0.5580 | 5.77E-01 | 5498/51403 | 6.94E-01 | -0.0271 | 0.0176 | -1.5374 | 1.24E-01 | 3491/61780 | 2.10E-01 |
| Acetone | 0.0149 | 0.0165 | 0.9025 | 3.67E-01 | 5498/51403 | 4.79E-01 | 0.0094 | 0.0196 | 0.4793 | 6.32E-01 | 3491/61780 | 7.32E-01 |
| Creatinine | 0.0042 | 0.0204 | 0.2053 | 8.37E-01 | 5159/49247 | 9.14E-01 | 0.0011 | 0.0243 | 0.0454 | 9.64E-01 | 3266/59170 | 9.76E-01 |
| Albumin | -0.0196 | 0.0170 | -1.1545 | 2.48E-01 | 5498/51388 | 3.54E-01 | -0.0351 | 0.0200 | -1.7518 | 7.98E-02 | 3491/61766 | 1.49E-01 |
| GlycA | 0.0382 | 0.0180 | 2.1270 | 3.34E-02 | 5498/51404 | 6.26E-02 | 0.0214 | 0.0213 | 1.0059 | 3.14E-01 | 3491/61782 | 4.36E-01 |
| XXL_VLDL_P | 0.0779 | 0.0176 | 4.4135 | 1.02E-05 | 5498/51404 | 5.87E-05 | 0.0804 | 0.0211 | 3.8193 | 1.34E-04 | 3491/61782 | 1.09E-03 |
| XXL_VLDL_L | 0.0733 | 0.0179 | 4.1005 | 4.12E-05 | 5498/51404 | 1.77E-04 | 0.0769 | 0.0214 | 3.6008 | 3.17E-04 | 3491/61782 | 1.58E-03 |
| XXL_VLDL_PL | 0.0724 | 0.0174 | 4.1556 | 3.24E-05 | 5498/51404 | 1.44E-04 | 0.0795 | 0.0209 | 3.7931 | 1.49E-04 | 3491/61782 | 1.09E-03 |
| XXL_VLDL_C | 0.0771 | 0.0178 | 4.3408 | 1.42E-05 | 5498/51404 | 6.93E-05 | 0.0791 | 0.0212 | 3.7267 | 1.94E-04 | 3491/61782 | 1.16E-03 |
| XXL_VLDL_CE | 0.0761 | 0.0177 | 4.2853 | 1.82E-05 | 5498/51404 | 8.57E-05 | 0.0758 | 0.0212 | 3.5755 | 3.50E-04 | 3491/61782 | 1.67E-03 |
| XXL_VLDL_FC | 0.0782 | 0.0177 | 4.4094 | 1.04E-05 | 5498/51404 | 5.87E-05 | 0.0829 | 0.0212 | 3.9134 | 9.10E-05 | 3491/61782 | 9.44E-04 |
| XXL_VLDL_TG | 0.0423 | 0.0175 | 2.4127 | 1.58E-02 | 5498/51404 | 3.31E-02 | 0.0471 | 0.0209 | 2.2575 | 2.40E-02 | 3491/61782 | 5.33E-02 |
| XL_VLDL_P | 0.0832 | 0.0176 | 4.7286 | 2.26E-06 | 5498/51404 | 2.39E-05 | 0.0783 | 0.0209 | 3.7448 | 1.81E-04 | 3491/61782 | 1.15E-03 |
| XL_VLDL_L | 0.0831 | 0.0178 | 4.6815 | 2.85E-06 | 5498/51404 | 2.73E-05 | 0.0783 | 0.0211 | 3.7100 | 2.07E-04 | 3491/61782 | 1.16E-03 |
| XL_VLDL_PL | 0.0710 | 0.0173 | 4.1055 | 4.03E-05 | 5498/51404 | 1.76E-04 | 0.0639 | 0.0205 | 3.1111 | 1.86E-03 | 3491/61782 | 6.27E-03 |
| XL_VLDL_C | 0.0663 | 0.0174 | 3.8141 | 1.37E-04 | 5498/51404 | 5.08E-04 | 0.0591 | 0.0206 | 2.8629 | 4.20E-03 | 3491/61782 | 1.31E-02 |
| XL_VLDL_CE | 0.0588 | 0.0173 | 3.4053 | 6.61E-04 | 5498/51404 | 1.98E-03 | 0.0491 | 0.0204 | 2.4055 | 1.62E-02 | 3491/61782 | 3.87E-02 |
| XL_VLDL_FC | 0.0744 | 0.0175 | 4.2450 | 2.19E-05 | 5498/51404 | 1.01E-04 | 0.0705 | 0.0208 | 3.3864 | 7.08E-04 | 3491/61782 | 2.71E-03 |
| XL_VLDL_TG | 0.0896 | 0.0180 | 4.9650 | 6.87E-07 | 5498/51404 | 1.53E-05 | 0.0852 | 0.0216 | 3.9469 | 7.92E-05 | 3491/61782 | 8.96E-04 |
| L_VLDL_P | 0.0819 | 0.0176 | 4.6473 | 3.36E-06 | 5498/51404 | 2.89E-05 | 0.0775 | 0.0209 | 3.7080 | 2.09E-04 | 3491/61782 | 1.16E-03 |
| L_VLDL_L | 0.0804 | 0.0176 | 4.5705 | 4.86E-06 | 5498/51404 | 3.36E-05 | 0.0754 | 0.0209 | 3.6066 | 3.10E-04 | 3491/61782 | 1.58E-03 |
| L_VLDL_PL | 0.0649 | 0.0171 | 3.8066 | 1.41E-04 | 5498/51404 | 5.16E-04 | 0.0510 | 0.0199 | 2.5683 | 1.02E-02 | 3491/61782 | 2.72E-02 |
| L_VLDL_C | 0.0690 | 0.0172 | 4.0045 | 6.22E-05 | 5498/51404 | 2.50E-04 | 0.0635 | 0.0204 | 3.1060 | 1.90E-03 | 3491/61782 | 6.30E-03 |
| L_VLDL_CE | 0.0605 | 0.0170 | 3.5518 | 3.83E-04 | 5498/51404 | 1.27E-03 | 0.0545 | 0.0202 | 2.7036 | 6.86E-03 | 3491/61782 | 1.92E-02 |
| L_VLDL_FC | 0.0778 | 0.0175 | 4.4478 | 8.68E-06 | 5498/51404 | 5.14E-05 | 0.0729 | 0.0208 | 3.5071 | 4.53E-04 | 3491/61782 | 1.96E-03 |
| L_VLDL_TG | 0.0849 | 0.0180 | 4.7088 | 2.49E-06 | 5498/51404 | 2.48E-05 | 0.0785 | 0.0214 | 3.6667 | 2.46E-04 | 3491/61782 | 1.30E-03 |
| M_VLDL_P | 0.0344 | 0.0172 | 1.9939 | 4.62E-02 | 5498/51404 | 8.45E-02 | 0.0315 | 0.0204 | 1.5407 | 1.23E-01 | 3491/61782 | 2.10E-01 |
| M_VLDL_L | 0.0454 | 0.0172 | 2.6416 | 8.25E-03 | 5498/51404 | 1.92E-02 | 0.0413 | 0.0204 | 2.0284 | 4.25E-02 | 3491/61782 | 8.75E-02 |
| M_VLDL_PL | 0.0333 | 0.0173 | 1.9187 | 5.50E-02 | 5498/51404 | 9.93E-02 | 0.0296 | 0.0205 | 1.4436 | 1.49E-01 | 3491/61782 | 2.40E-01 |
| M_VLDL_C | -0.0006 | 0.0180 | -0.0322 | 9.74E-01 | 5498/51404 | 9.85E-01 | -0.0018 | 0.0213 | -0.0864 | 9.31E-01 | 3491/61782 | 9.46E-01 |
| M_VLDL_CE | -0.0126 | 0.0184 | -0.6847 | 4.94E-01 | 5498/51404 | 6.24E-01 | -0.0143 | 0.0214 | -0.6659 | 5.05E-01 | 3491/61782 | 6.25E-01 |
| M_VLDL_FC | 0.0212 | 0.0174 | 1.2205 | 2.22E-01 | 5498/51404 | 3.24E-01 | 0.0187 | 0.0206 | 0.9088 | 3.63E-01 | 3491/61782 | 4.77E-01 |
| M_VLDL_TG | 0.0764 | 0.0175 | 4.3635 | 1.28E-05 | 5498/51404 | 6.52E-05 | 0.0713 | 0.0208 | 3.4340 | 5.95E-04 | 3491/61782 | 2.35E-03 |
| S_VLDL_P | 0.0534 | 0.0169 | 3.1504 | 1.63E-03 | 5498/51404 | 4.49E-03 | 0.0528 | 0.0201 | 2.6322 | 8.48E-03 | 3491/61782 | 2.35E-02 |
| S_VLDL_L | 0.0511 | 0.0169 | 3.0154 | 2.57E-03 | 5498/51404 | 6.52E-03 | 0.0500 | 0.0201 | 2.4937 | 1.26E-02 | 3491/61782 | 3.12E-02 |
| S_VLDL_PL | 0.0318 | 0.0170 | 1.8703 | 6.14E-02 | 5498/51404 | 1.07E-01 | 0.0300 | 0.0202 | 1.4883 | 1.37E-01 | 3491/61782 | 2.27E-01 |
| S_VLDL_C | 0.0274 | 0.0170 | 1.6093 | 1.08E-01 | 5498/51404 | 1.82E-01 | 0.0272 | 0.0201 | 1.3491 | 1.77E-01 | 3491/61782 | 2.72E-01 |
| S_VLDL_CE | 0.0336 | 0.0169 | 1.9881 | 4.68E-02 | 5498/51404 | 8.51E-02 | 0.0337 | 0.0200 | 1.6870 | 9.16E-02 | 3491/61782 | 1.66E-01 |
| S_VLDL_FC | 0.0161 | 0.0173 | 0.9348 | 3.50E-01 | 5498/51404 | 4.71E-01 | 0.0149 | 0.0205 | 0.7301 | 4.65E-01 | 3491/61782 | 5.91E-01 |
| S_VLDL_TG | 0.0792 | 0.0171 | 4.6226 | 3.79E-06 | 5498/51404 | 2.94E-05 | 0.0787 | 0.0203 | 3.8778 | 1.05E-04 | 3491/61782 | 9.61E-04 |
| XS_VLDL_P | 0.0049 | 0.0174 | 0.2835 | 7.77E-01 | 5498/51404 | 8.73E-01 | 0.0090 | 0.0207 | 0.4337 | 6.64E-01 | 3491/61782 | 7.51E-01 |
| XS_VLDL_L | 0.0099 | 0.0174 | 0.5688 | 5.69E-01 | 5498/51404 | 6.92E-01 | 0.0133 | 0.0206 | 0.6471 | 5.18E-01 | 3491/61782 | 6.32E-01 |
| XS_VLDL_PL | 0.0161 | 0.0173 | 0.9318 | 3.51E-01 | 5498/51404 | 4.71E-01 | 0.0206 | 0.0204 | 1.0071 | 3.14E-01 | 3491/61782 | 4.36E-01 |
| XS_VLDL_C | -0.0168 | 0.0183 | -0.9137 | 3.61E-01 | 5498/51404 | 4.77E-01 | -0.0136 | 0.0218 | -0.6250 | 5.32E-01 | 3491/61782 | 6.46E-01 |
| XS_VLDL_CE | -0.0248 | 0.0187 | -1.3261 | 1.85E-01 | 5498/51404 | 2.81E-01 | -0.0223 | 0.0223 | -1.0010 | 3.17E-01 | 3491/61782 | 4.36E-01 |
| XS_VLDL_FC | 0.0012 | 0.0176 | 0.0704 | 9.44E-01 | 5498/51404 | 9.67E-01 | 0.0061 | 0.0209 | 0.2901 | 7.72E-01 | 3491/61782 | 8.25E-01 |
| XS_VLDL_TG | 0.0589 | 0.0168 | 3.5071 | 4.53E-04 | 5498/51404 | 1.45E-03 | 0.0634 | 0.0199 | 3.1929 | 1.41E-03 | 3491/61782 | 4.94E-03 |
| IDL_P | -0.0245 | 0.0182 | -1.3445 | 1.79E-01 | 5498/51404 | 2.76E-01 | -0.0286 | 0.0216 | -1.3236 | 1.86E-01 | 3491/61782 | 2.78E-01 |
| IDL_L | -0.0416 | 0.0187 | -2.2203 | 2.64E-02 | 5498/51404 | 5.14E-02 | -0.0464 | 0.0222 | -2.0871 | 3.69E-02 | 3491/61782 | 7.65E-02 |
| IDL_PL | -0.0377 | 0.0187 | -2.0132 | 4.41E-02 | 5498/51404 | 8.13E-02 | -0.0390 | 0.0222 | -1.7567 | 7.90E-02 | 3491/61782 | 1.49E-01 |
| IDL_C | -0.0500 | 0.0190 | -2.6328 | 8.47E-03 | 5498/51404 | 1.95E-02 | -0.0565 | 0.0225 | -2.5041 | 1.23E-02 | 3491/61782 | 3.12E-02 |
| IDL_CE | -0.0472 | 0.0190 | -2.4846 | 1.30E-02 | 5498/51404 | 2.83E-02 | -0.0561 | 0.0225 | -2.4942 | 1.26E-02 | 3491/61782 | 3.12E-02 |
| IDL_FC | -0.0565 | 0.0189 | -2.9848 | 2.84E-03 | 5498/51404 | 6.99E-03 | -0.0552 | 0.0225 | -2.4547 | 1.41E-02 | 3491/61782 | 3.44E-02 |
| IDL_TG | 0.0401 | 0.0168 | 2.3841 | 1.71E-02 | 5498/51404 | 3.55E-02 | 0.0441 | 0.0199 | 2.2163 | 2.67E-02 | 3491/61782 | 5.86E-02 |
| L_LDL_P | -0.0028 | 0.0176 | -0.1578 | 8.75E-01 | 5498/51404 | 9.31E-01 | -0.0066 | 0.0208 | -0.3170 | 7.51E-01 | 3491/61782 | 8.13E-01 |
| L_LDL_L | -0.0216 | 0.0177 | -1.2195 | 2.23E-01 | 5498/51404 | 3.24E-01 | -0.0282 | 0.0210 | -1.3428 | 1.79E-01 | 3491/61782 | 2.72E-01 |
| L_LDL_PL | -0.0242 | 0.0178 | -1.3585 | 1.74E-01 | 5498/51404 | 2.75E-01 | -0.0306 | 0.0211 | -1.4500 | 1.47E-01 | 3491/61782 | 2.39E-01 |
| L_LDL_C | -0.0181 | 0.0143 | -1.2687 | 2.05E-01 | 5498/51404 | 3.03E-01 | -0.0233 | 0.0151 | -1.5482 | 1.22E-01 | 3491/61782 | 2.10E-01 |
| L_LDL_CE | -0.0183 | 0.0159 | -1.1488 | 2.51E-01 | 5498/51404 | 3.55E-01 | -0.0260 | 0.0176 | -1.4772 | 1.40E-01 | 3491/61782 | 2.30E-01 |
| L_LDL_FC | -0.0262 | 0.0153 | -1.7114 | 8.70E-02 | 5498/51404 | 1.49E-01 | -0.0267 | 0.0148 | -1.8043 | 7.12E-02 | 3491/61782 | 1.37E-01 |
| L_LDL_TG | 0.0320 | 0.0170 | 1.8854 | 5.94E-02 | 5498/51404 | 1.05E-01 | 0.0347 | 0.0201 | 1.7294 | 8.37E-02 | 3491/61782 | 1.56E-01 |
| M_LDL_P | -0.0005 | 0.0174 | -0.0295 | 9.77E-01 | 5498/51404 | 9.85E-01 | -0.0004 | 0.0207 | -0.0174 | 9.86E-01 | 3491/61782 | 9.90E-01 |
| M_LDL_L | 0.0033 | 0.0172 | 0.1899 | 8.49E-01 | 5498/51404 | 9.19E-01 | -0.0031 | 0.0204 | -0.1531 | 8.78E-01 | 3491/61782 | 9.04E-01 |
| M_LDL_PL | 0.0027 | 0.0168 | 0.1638 | 8.70E-01 | 5498/51404 | 9.30E-01 | -0.0047 | 0.0197 | -0.2382 | 8.12E-01 | 3491/61782 | 8.49E-01 |
| M_LDL_C | -0.0037 | 0.0140 | -0.2655 | 7.91E-01 | 5498/51404 | 8.77E-01 | -0.0114 | 0.0149 | -0.7638 | 4.45E-01 | 3491/61782 | 5.74E-01 |
| M_LDL_CE | -0.0002 | 0.0136 | -0.0174 | 9.86E-01 | 5498/51404 | 9.87E-01 | -0.0083 | 0.0145 | -0.5713 | 5.68E-01 | 3491/61782 | 6.80E-01 |
| M_LDL_FC | -0.0165 | 0.0143 | -1.1538 | 2.49E-01 | 5498/51404 | 3.54E-01 | -0.0230 | 0.0153 | -1.5060 | 1.32E-01 | 3491/61782 | 2.21E-01 |
| M_LDL_TG | 0.0422 | 0.0172 | 2.4596 | 1.39E-02 | 5498/51404 | 2.99E-02 | 0.0444 | 0.0203 | 2.1887 | 2.86E-02 | 3491/61782 | 6.20E-02 |
| S_LDL_P | -0.0020 | 0.0174 | -0.1122 | 9.11E-01 | 5498/51404 | 9.45E-01 | -0.0025 | 0.0207 | -0.1215 | 9.03E-01 | 3491/61782 | 9.26E-01 |
| S_LDL_L | -0.0059 | 0.0174 | -0.3409 | 7.33E-01 | 5498/51404 | 8.37E-01 | -0.0096 | 0.0206 | -0.4657 | 6.41E-01 | 3491/61782 | 7.36E-01 |

| | | | | | | | | | | | | |
|---|---|---|---|---|---|---|---|---|---|---|---|---|
| S_LDL_PL | -0.0186 | 0.0174 | -1.0688 | 2.85E-01 | 5498/51404 | 3.94E-01 | -0.0187 | 0.0206 | -0.9082 | 3.64E-01 | 3491/61782 | 4.77E-01 |
| S_LDL_C | -0.0076 | 0.0134 | -0.5646 | 5.72E-01 | 5498/51404 | 6.92E-01 | -0.0137 | 0.0142 | -0.9657 | 3.34E-01 | 3491/61782 | 4.56E-01 |
| S_LDL_CE | -0.0026 | 0.0137 | -0.1893 | 8.50E-01 | 5498/51404 | 9.19E-01 | -0.0098 | 0.0146 | -0.6763 | 4.99E-01 | 3491/61782 | 6.24E-01 |
| S_LDL_FC | -0.0195 | 0.0144 | -1.3490 | 1.77E-01 | 5498/51404 | 2.76E-01 | -0.0235 | 0.0153 | -1.5303 | 1.26E-01 | 3491/61782 | 2.12E-01 |
| S_LDL_TG | 0.0578 | 0.0174 | 3.3242 | 8.87E-04 | 5498/51404 | 2.57E-03 | 0.0591 | 0.0205 | 2.8757 | 4.03E-03 | 3491/61782 | 1.27E-02 |
| XL_HDL_P | -0.0991 | 0.0193 | -5.1356 | 2.81E-07 | 5498/51404 | 1.17E-05 | -0.0870 | 0.0232 | -3.7509 | 1.76E-04 | 3491/61782 | 1.15E-03 |
| XL_HDL_L | -0.0960 | 0.0194 | -4.9519 | 7.35E-07 | 5498/51404 | 1.53E-05 | -0.0820 | 0.0231 | -3.5445 | 3.93E-04 | 3491/61782 | 1.75E-03 |
| XL_HDL_PL | -0.0522 | 0.0166 | -3.1483 | 1.64E-03 | 5498/51404 | 4.49E-03 | -0.0374 | 0.0205 | -1.8261 | 6.78E-02 | 3491/61782 | 1.32E-01 |
| XL_HDL_C | -0.1028 | 0.0190 | -5.4135 | 6.18E-08 | 5498/51404 | 4.43E-06 | -0.0862 | 0.0226 | -3.8125 | 1.38E-04 | 3491/61782 | 1.09E-03 |
| XL_HDL_CE | -0.0826 | 0.0173 | -4.7823 | 1.73E-06 | 5498/51404 | 2.39E-05 | -0.0710 | 0.0209 | -3.4026 | 6.67E-04 | 3491/61782 | 2.60E-03 |
| XL_HDL_FC | -0.0839 | 0.0171 | -4.8953 | 9.82E-07 | 5498/51404 | 1.73E-05 | -0.0700 | 0.0202 | -3.4709 | 5.19E-04 | 3491/61782 | 2.15E-03 |
| XL_HDL_TG | 0.0167 | 0.0167 | 1.0022 | 3.16E-01 | 5498/51404 | 4.30E-01 | 0.0187 | 0.0197 | 0.9509 | 3.42E-01 | 3491/61782 | 4.62E-01 |
| L_HDL_P | -0.0941 | 0.0199 | -4.7252 | 2.30E-06 | 5498/51404 | 2.39E-05 | -0.0847 | 0.0238 | -3.5544 | 3.79E-04 | 3491/61782 | 1.72E-03 |
| L_HDL_L | -0.0960 | 0.0203 | -4.7333 | 2.21E-06 | 5498/51404 | 2.39E-05 | -0.0907 | 0.0242 | -3.7452 | 1.80E-04 | 3491/61782 | 1.15E-03 |
| L_HDL_PL | -0.0880 | 0.0200 | -4.3929 | 1.12E-05 | 5498/51404 | 6.06E-05 | -0.0867 | 0.0243 | -3.5622 | 3.68E-04 | 3491/61782 | 1.72E-03 |
| L_HDL_C | -0.0930 | 0.0194 | -4.7878 | 1.69E-06 | 5498/51404 | 2.39E-05 | -0.0883 | 0.0235 | -3.7566 | 1.72E-04 | 3491/61782 | 1.15E-03 |
| L_HDL_CE | -0.0924 | 0.0195 | -4.7426 | 2.11E-06 | 5498/51404 | 2.39E-05 | -0.0884 | 0.0236 | -3.7493 | 1.77E-04 | 3491/61782 | 1.15E-03 |
| L_HDL_FC | -0.0856 | 0.0188 | -4.5455 | 5.48E-06 | 5498/51404 | 3.50E-05 | -0.0790 | 0.0229 | -3.4503 | 5.60E-04 | 3491/61782 | 2.25E-03 |
| L_HDL_TG | 0.0079 | 0.0168 | 0.4724 | 6.37E-01 | 5498/51404 | 7.48E-01 | 0.0060 | 0.0199 | 0.3032 | 7.62E-01 | 3491/61782 | 8.19E-01 |
| M_HDL_P | -0.0629 | 0.0181 | -3.4851 | 4.92E-04 | 5498/51404 | 1.55E-03 | -0.0761 | 0.0210 | -3.6231 | 2.91E-04 | 3491/61782 | 1.51E-03 |
| M_HDL_L | -0.0427 | 0.0189 | -2.2608 | 2.38E-02 | 5498/51404 | 4.73E-02 | -0.0524 | 0.0225 | -2.3251 | 2.01E-02 | 3491/61782 | 4.67E-02 |
| M_HDL_PL | -0.0302 | 0.0188 | -1.6069 | 1.08E-01 | 5498/51404 | 1.82E-01 | -0.0396 | 0.0224 | -1.7627 | 7.79E-02 | 3491/61782 | 1.48E-01 |
| M_HDL_C | -0.0699 | 0.0175 | -3.9948 | 6.48E-05 | 5498/51404 | 2.52E-04 | -0.0887 | 0.0219 | -4.0440 | 5.25E-05 | 3491/61782 | 7.27E-04 |
| M_HDL_CE | -0.0690 | 0.0171 | -4.0270 | 5.65E-05 | 5498/51404 | 2.31E-04 | -0.0895 | 0.0218 | -4.1010 | 4.11E-05 | 3491/61782 | 6.83E-04 |
| M_HDL_FC | -0.0690 | 0.0185 | -3.7382 | 1.85E-04 | 5498/51404 | 6.69E-04 | -0.0831 | 0.0225 | -3.7021 | 2.14E-04 | 3491/61782 | 1.16E-03 |
| M_HDL_TG | 0.0608 | 0.0167 | 3.6482 | 2.64E-04 | 5498/51404 | 9.13E-04 | 0.0549 | 0.0197 | 2.7807 | 5.42E-03 | 3491/61782 | 1.61E-02 |
| S_HDL_P | 0.0102 | 0.0171 | 0.5954 | 5.52E-01 | 5498/51404 | 6.77E-01 | -0.0022 | 0.0203 | -0.1087 | 9.13E-01 | 3491/61782 | 9.32E-01 |
| S_HDL_L | 0.0221 | 0.0173 | 1.2748 | 2.02E-01 | 5498/51404 | 3.02E-01 | 0.0102 | 0.0205 | 0.4980 | 6.18E-01 | 3491/61782 | 7.26E-01 |
| S_HDL_PL | 0.0219 | 0.0174 | 1.2545 | 2.10E-01 | 5498/51404 | 3.09E-01 | 0.0101 | 0.0207 | 0.4867 | 6.26E-01 | 3491/61782 | 7.32E-01 |
| S_HDL_C | 0.0025 | 0.0172 | 0.1475 | 8.83E-01 | 5498/51404 | 9.31E-01 | -0.0103 | 0.0205 | -0.5018 | 6.16E-01 | 3491/61782 | 7.26E-01 |
| S_HDL_CE | 0.0049 | 0.0178 | 0.2774 | 7.82E-01 | 5498/51404 | 8.73E-01 | -0.0090 | 0.0211 | -0.4267 | 6.70E-01 | 3491/61782 | 7.51E-01 |
| S_HDL_FC | -0.0043 | 0.0176 | -0.2422 | 8.09E-01 | 5498/51404 | 8.87E-01 | -0.0139 | 0.0208 | -0.6663 | 5.05E-01 | 3491/61782 | 6.25E-01 |
| S_HDL_TG | 0.0845 | 0.0174 | 4.8548 | 1.21E-06 | 5498/51404 | 1.88E-05 | 0.0817 | 0.0207 | 3.9519 | 7.75E-05 | 3491/61782 | 8.96E-04 |
| XXL_VLDL_PL_pct | 0.0449 | 0.0161 | 2.7979 | 5.14E-03 | 5335/49767 | 1.24E-02 | 0.0532 | 0.0194 | 2.7382 | 6.18E-03 | 3391/59815 | 1.79E-02 |
| XXL_VLDL_C_pct | -0.0183 | 0.0162 | -1.1312 | 2.58E-01 | 5335/49767 | 3.63E-01 | -0.0200 | 0.0191 | -1.0431 | 2.97E-01 | 3391/59815 | 4.18E-01 |
| XXL_VLDL_CE_pct | -0.0096 | 0.0162 | -0.5913 | 5.54E-01 | 5335/49767 | 6.77E-01 | -0.0148 | 0.0191 | -0.7771 | 4.37E-01 | 3391/59815 | 5.67E-01 |
| XXL_VLDL_FC_pct | -0.0146 | 0.0161 | -0.9013 | 3.67E-01 | 5335/49767 | 4.79E-01 | -0.0072 | 0.0192 | -0.3747 | 7.08E-01 | 3391/59815 | 7.80E-01 |
| XXL_VLDL_TG_pct | 0.0091 | 0.0164 | 0.5534 | 5.80E-01 | 5335/49767 | 6.94E-01 | 0.0089 | 0.0189 | 0.4713 | 6.37E-01 | 3391/59815 | 7.35E-01 |
| XL_VLDL_PL_pct | 0.0343 | 0.0159 | 2.1580 | 3.09E-02 | 5447/50959 | 5.88E-02 | 0.0419 | 0.0196 | 2.1421 | 3.22E-02 | 3455/61253 | 6.79E-02 |
| XL_VLDL_C_pct | -0.0840 | 0.0177 | -4.7497 | 2.04E-06 | 5447/50959 | 2.39E-05 | -0.0878 | 0.0209 | -4.1909 | 2.78E-05 | 3455/61253 | 5.32E-04 |
| XL_VLDL_CE_pct | -0.0824 | 0.0178 | -4.6412 | 3.46E-06 | 5447/50959 | 2.89E-05 | -0.0884 | 0.0210 | -4.2056 | 2.60E-05 | 3455/61253 | 5.32E-04 |
| XL_VLDL_FC_pct | -0.0576 | 0.0166 | -3.4811 | 4.99E-04 | 5447/50959 | 1.55E-03 | -0.0492 | 0.0195 | -2.5289 | 1.14E-02 | 3455/61253 | 3.00E-02 |
| XL_VLDL_TG_pct | 0.0790 | 0.0206 | 3.8370 | 1.25E-04 | 5447/50959 | 4.76E-04 | 0.1054 | 0.0264 | 3.9942 | 6.49E-05 | 3455/61253 | 8.51E-04 |
| L_VLDL_PL_pct | 0.0365 | 0.0156 | 2.3368 | 1.94E-02 | 5498/51397 | 3.94E-02 | 0.0216 | 0.0176 | 1.2244 | 2.21E-01 | 3491/61775 | 3.27E-01 |
| L_VLDL_C_pct | -0.0297 | 0.0158 | -1.8817 | 5.99E-02 | 5498/51397 | 1.05E-01 | -0.0305 | 0.0186 | -1.6440 | 1.00E-01 | 3491/61775 | 1.79E-01 |
| L_VLDL_CE_pct | -0.0510 | 0.0170 | -3.0037 | 2.67E-03 | 5498/51397 | 6.67E-03 | -0.0519 | 0.0200 | -2.5965 | 9.42E-03 | 3491/61775 | 2.55E-02 |
| L_VLDL_FC_pct | 0.0205 | 0.0153 | 1.3457 | 1.78E-01 | 5498/51397 | 2.76E-01 | 0.0194 | 0.0181 | 1.0729 | 2.83E-01 | 3491/61775 | 4.07E-01 |
| L_VLDL_TG_pct | -0.0029 | 0.0191 | -0.1498 | 8.81E-01 | 5498/51397 | 9.31E-01 | -0.0093 | 0.0231 | -0.4022 | 6.88E-01 | 3491/61775 | 7.62E-01 |
| M_VLDL_PL_pct | -0.0347 | 0.0160 | -2.1733 | 2.98E-02 | 5498/51404 | 5.74E-02 | -0.0325 | 0.0173 | -1.8802 | 6.01E-02 | 3491/61782 | 1.19E-01 |
| M_VLDL_C_pct | -0.0830 | 0.0189 | -4.3986 | 1.09E-05 | 5498/51404 | 6.03E-05 | -0.0782 | 0.0223 | -3.5054 | 4.56E-04 | 3491/61782 | 1.96E-03 |
| M_VLDL_CE_pct | -0.0537 | 0.0175 | -3.0673 | 2.16E-03 | 5498/51404 | 5.64E-03 | -0.0497 | 0.0199 | -2.4992 | 1.24E-02 | 3491/61782 | 3.12E-02 |
| M_VLDL_FC_pct | -0.0741 | 0.0183 | -4.0494 | 5.13E-05 | 5498/51404 | 2.17E-04 | -0.0692 | 0.0215 | -3.2105 | 1.33E-03 | 3491/61782 | 4.85E-03 |
| M_VLDL_TG_pct | 0.1012 | 0.0192 | 5.2658 | 1.40E-07 | 5498/51404 | 6.95E-06 | 0.0982 | 0.0229 | 4.2813 | 1.86E-05 | 3491/61782 | 5.32E-04 |
| S_VLDL_PL_pct | -0.0840 | 0.0182 | -4.6211 | 3.82E-06 | 5498/51404 | 2.94E-05 | -0.0875 | 0.0215 | -4.0639 | 4.83E-05 | 3491/61782 | 7.07E-04 |
| S_VLDL_C_pct | -0.0632 | 0.0178 | -3.5460 | 3.91E-04 | 5498/51404 | 1.27E-03 | -0.0609 | 0.0211 | -2.8877 | 3.88E-03 | 3491/61782 | 1.25E-02 |
| S_VLDL_CE_pct | -0.0378 | 0.0170 | -2.2244 | 2.61E-02 | 5498/51404 | 5.12E-02 | -0.0341 | 0.0200 | -1.7024 | 8.87E-02 | 3491/61782 | 1.64E-01 |
| S_VLDL_FC_pct | -0.0842 | 0.0185 | -4.5480 | 5.42E-06 | 5498/51404 | 3.50E-05 | -0.0847 | 0.0219 | -3.8716 | 1.08E-04 | 3491/61782 | 9.61E-04 |
| S_VLDL_TG_pct | 0.0806 | 0.0176 | 4.5759 | 4.74E-06 | 5498/51404 | 3.36E-05 | 0.0823 | 0.0210 | 3.9190 | 8.89E-05 | 3491/61782 | 9.44E-04 |
| XS_VLDL_PL_pct | 0.0607 | 0.0184 | 3.3038 | 9.54E-04 | 5498/51404 | 2.67E-03 | 0.0722 | 0.0218 | 3.3055 | 9.48E-04 | 3491/61782 | 3.58E-03 |
| XS_VLDL_C_pct | -0.0884 | 0.0194 | -4.5557 | 5.22E-06 | 5498/51404 | 3.50E-05 | -0.0891 | 0.0229 | -3.8882 | 1.01E-04 | 3491/61782 | 9.61E-04 |
| XS_VLDL_CE_pct | -0.0880 | 0.0196 | -4.5008 | 6.77E-06 | 5498/51404 | 4.11E-05 | -0.0893 | 0.0230 | -3.8788 | 1.05E-04 | 3491/61782 | 9.61E-04 |
| XS_VLDL_FC_pct | -0.0584 | 0.0176 | -3.3091 | 9.36E-04 | 5498/51404 | 2.65E-03 | -0.0492 | 0.0210 | -2.3396 | 1.93E-02 | 3491/61782 | 4.53E-02 |
| XS_VLDL_TG_pct | 0.0939 | 0.0188 | 4.9899 | 6.04E-07 | 5498/51404 | 1.50E-05 | 0.0989 | 0.0224 | 4.4108 | 1.03E-05 | 3491/61782 | 4.98E-04 |
| IDL_PL_pct | 0.0281 | 0.0166 | 1.6887 | 9.13E-02 | 5498/51404 | 1.56E-01 | 0.0451 | 0.0197 | 2.2882 | 2.21E-02 | 3491/61782 | 5.05E-02 |
| IDL_C_pct | -0.0649 | 0.0183 | -3.5455 | 3.92E-04 | 5498/51404 | 1.27E-03 | -0.0752 | 0.0215 | -3.4959 | 4.72E-04 | 3491/61782 | 1.99E-03 |
| IDL_CE_pct | -0.0453 | 0.0181 | -2.4991 | 1.25E-02 | 5498/51404 | 2.74E-02 | -0.0674 | 0.0213 | -3.1695 | 1.53E-03 | 3491/61782 | 5.28E-03 |
| IDL_FC_pct | -0.0656 | 0.0171 | -3.8338 | 1.26E-04 | 5498/51404 | 4.76E-04 | -0.0443 | 0.0203 | -2.1850 | 2.89E-02 | 3491/61782 | 6.20E-02 |
| IDL_TG_pct | 0.0833 | 0.0185 | 4.5059 | 6.61E-06 | 5498/51404 | 4.11E-05 | 0.0925 | 0.0219 | 4.2246 | 2.39E-05 | 3491/61782 | 5.32E-04 |
| L_LDL_PL_pct | -0.0102 | 0.0166 | -0.6161 | 5.38E-01 | 5498/51404 | 6.66E-01 | -0.0069 | 0.0195 | -0.3571 | 7.21E-01 | 3491/61782 | 7.87E-01 |
| L_LDL_C_pct | -0.0079 | 0.0106 | -0.7427 | 4.58E-01 | 5498/51404 | 5.84E-01 | -0.0113 | 0.0106 | -1.0682 | 2.85E-01 | 3491/61782 | 4.07E-01 |
| L_LDL_CE_pct | -0.0042 | 0.0120 | -0.3528 | 7.24E-01 | 5498/51404 | 8.37E-01 | -0.0098 | 0.0120 | -0.8157 | 4.15E-01 | 3491/61782 | 5.41E-01 |
| L_LDL_FC_pct | -0.0171 | 0.0128 | -1.3316 | 1.83E-01 | 5498/51404 | 2.80E-01 | -0.0155 | 0.0108 | -1.4387 | 1.50E-01 | 3491/61782 | 2.40E-01 |
| L_LDL_TG_pct | 0.0601 | 0.0181 | 3.3196 | 9.01E-04 | 5498/51404 | 2.58E-03 | 0.0702 | 0.0214 | 3.2872 | 1.01E-03 | 3491/61782 | 3.76E-03 |
| M_LDL_PL_pct | -0.0012 | 0.0105 | -0.1168 | 9.07E-01 | 5498/51404 | 9.45E-01 | -0.0043 | 0.0108 | -0.4003 | 6.89E-01 | 3491/61782 | 7.62E-01 |
| M_LDL_C_pct | -0.0056 | 0.0094 | -0.5937 | 5.53E-01 | 5498/51404 | 6.77E-01 | -0.0088 | 0.0094 | -0.9379 | 3.48E-01 | 3491/61782 | 4.66E-01 |

| | | | | | | | | | | | | |
|---|---|---|---|---|---|---|---|---|---|---|---|---|
| M_LDL_CE_pct | -0.0013 | 0.0096 | -0.1342 | 8.93E-01 | 5498/51404 | 9.38E-01 | -0.0056 | 0.0095 | -0.5938 | 5.53E-01 | 3491/61782 | 6.65E-01 |
| M_LDL_FC_pct | -0.0235 | 0.0152 | -1.5432 | 1.23E-01 | 5498/51404 | 2.01E-01 | -0.0230 | 0.0136 | -1.6944 | 9.02E-02 | 3491/61782 | 1.65E-01 |
| M_LDL_TG_pct | 0.0480 | 0.0176 | 2.7305 | 6.32E-03 | 5498/51404 | 1.49E-02 | 0.0582 | 0.0207 | 2.8065 | 5.01E-03 | 3491/61782 | 1.52E-02 |
| S_LDL_PL_pct | -0.0423 | 0.0167 | -2.5397 | 1.11E-02 | 5498/51404 | 2.47E-02 | -0.0290 | 0.0197 | -1.4678 | 1.42E-01 | 3491/61782 | 2.33E-01 |
| S_LDL_C_pct | -0.0046 | 0.0094 | -0.4898 | 6.24E-01 | 5498/51404 | 7.40E-01 | -0.0085 | 0.0093 | -0.9132 | 3.61E-01 | 3491/61782 | 4.77E-01 |
| S_LDL_CE_pct | 0.0003 | 0.0104 | 0.0287 | 9.77E-01 | 5498/51404 | 9.85E-01 | -0.0053 | 0.0099 | -0.5411 | 5.88E-01 | 3491/61782 | 6.98E-01 |
| S_LDL_FC_pct | -0.0186 | 0.0132 | -1.4171 | 1.56E-01 | 5498/51404 | 2.50E-01 | -0.0197 | 0.0128 | -1.5456 | 1.22E-01 | 3491/61782 | 2.10E-01 |
| S_LDL_TG_pct | 0.0746 | 0.0177 | 4.2273 | 2.37E-05 | 5498/51404 | 1.07E-04 | 0.0793 | 0.0209 | 3.8008 | 1.44E-04 | 3491/61782 | 1.09E-03 |
| XL_HDL_PL_pct | -0.0074 | 0.0154 | -0.4787 | 6.32E-01 | 5497/51392 | 7.46E-01 | 0.0068 | 0.0205 | 0.3298 | 7.42E-01 | 3491/61768 | 8.06E-01 |
| XL_HDL_C_pct | -0.0030 | 0.0176 | -0.1711 | 8.64E-01 | 5497/51392 | 9.27E-01 | 0.0090 | 0.0212 | 0.4271 | 6.69E-01 | 3491/61768 | 7.51E-01 |
| XL_HDL_CE_pct | -0.0208 | 0.0136 | -1.5357 | 1.25E-01 | 5497/51392 | 2.03E-01 | -0.0175 | 0.0164 | -1.0634 | 2.88E-01 | 3491/61768 | 4.07E-01 |
| XL_HDL_FC_pct | 0.0612 | 0.0195 | 3.1304 | 1.75E-03 | 5497/51392 | 4.62E-03 | 0.0580 | 0.0232 | 2.4965 | 1.25E-02 | 3491/61768 | 3.12E-02 |
| XL_HDL_TG_pct | 0.0893 | 0.0182 | 4.9092 | 9.14E-07 | 5497/51392 | 1.73E-05 | 0.0803 | 0.0216 | 3.7139 | 2.04E-04 | 3491/61768 | 1.16E-03 |
| L_HDL_PL_pct | 0.1335 | 0.0307 | 4.3539 | 1.34E-05 | 5498/51404 | 6.66E-05 | 0.1021 | 0.0367 | 2.7815 | 5.41E-03 | 3491/61782 | 1.61E-02 |
| L_HDL_C_pct | -0.0400 | 0.0145 | -2.7656 | 5.68E-03 | 5498/51404 | 1.35E-02 | -0.0388 | 0.0185 | -2.0973 | 3.60E-02 | 3491/61782 | 7.53E-02 |
| L_HDL_CE_pct | -0.0395 | 0.0154 | -2.5676 | 1.02E-02 | 5498/51404 | 2.30E-02 | -0.0388 | 0.0193 | -2.0101 | 4.44E-02 | 3491/61782 | 8.92E-02 |
| L_HDL_FC_pct | -0.0342 | 0.0151 | -2.2722 | 2.31E-02 | 5498/51404 | 4.63E-02 | -0.0277 | 0.0198 | -1.3949 | 1.63E-01 | 3491/61782 | 2.59E-01 |
| L_HDL_TG_pct | 0.0825 | 0.0180 | 4.5805 | 4.64E-06 | 5498/51404 | 3.36E-05 | 0.0764 | 0.0215 | 3.5573 | 3.75E-04 | 3491/61782 | 1.72E-03 |
| M_HDL_PL_pct | 0.1405 | 0.0236 | 5.9506 | 2.67E-09 | 5498/51404 | 6.65E-07 | 0.1469 | 0.0281 | 5.2335 | 1.66E-07 | 3491/61782 | 4.14E-05 |
| M_HDL_C_pct | -0.0603 | 0.0161 | -3.7342 | 1.88E-04 | 5498/51404 | 6.70E-04 | -0.1259 | 0.0278 | -4.5228 | 6.10E-06 | 3491/61782 | 3.80E-04 |
| M_HDL_CE_pct | -0.0543 | 0.0148 | -3.6665 | 2.46E-04 | 5498/51404 | 8.62E-04 | -0.1080 | 0.0257 | -4.2011 | 2.66E-05 | 3491/61782 | 5.32E-04 |
| M_HDL_FC_pct | -0.0744 | 0.0170 | -4.3631 | 1.28E-05 | 5498/51404 | 6.52E-05 | -0.1168 | 0.0257 | -4.5470 | 5.44E-06 | 3491/61782 | 3.80E-04 |
| M_HDL_TG_pct | 0.0887 | 0.0176 | 5.0514 | 4.39E-07 | 5498/51404 | 1.37E-05 | 0.0878 | 0.0209 | 4.1940 | 2.74E-05 | 3491/61782 | 5.32E-04 |
| S_HDL_PL_pct | 0.0005 | 0.0172 | 0.0295 | 9.76E-01 | 5498/51404 | 9.85E-01 | -0.0002 | 0.0205 | -0.0117 | 9.91E-01 | 3491/61782 | 9.91E-01 |
| S_HDL_C_pct | -0.0520 | 0.0170 | -3.0651 | 2.18E-03 | 5498/51404 | 5.64E-03 | -0.0552 | 0.0201 | -2.7512 | 5.94E-03 | 3491/61782 | 1.74E-02 |
| S_HDL_CE_pct | -0.0365 | 0.0191 | -1.9057 | 5.67E-02 | 5498/51404 | 1.02E-01 | -0.0422 | 0.0225 | -1.8745 | 6.09E-02 | 3491/61782 | 1.19E-01 |
| S_HDL_FC_pct | -0.0730 | 0.0181 | -4.0303 | 5.57E-05 | 5498/51404 | 2.31E-04 | -0.0672 | 0.0215 | -3.1201 | 1.81E-03 | 3491/61782 | 6.17E-03 |
| S_HDL_TG_pct | 0.0900 | 0.0178 | 5.0629 | 4.13E-07 | 5498/51404 | 1.37E-05 | 0.0928 | 0.0212 | 4.3776 | 1.20E-05 | 3491/61782 | 4.98E-04 |

**Supplementary Table 3**

**Significant Findings and replication in BBMRI-NL**

| | UK Biobank | | | | BBMRI-NL | | | | |
|---|---|---|---|---|---|---|---|---|---|
| metabolite | Z_MDD | FDR_MDD | Z_RECURRENT | FDR_RECURRENT | Effect | SE | Z | pval | FDR |
| Ala | 2.7183 | 1.54E-02 | 1.7793 | 1.56E-01 | 0.0237 | 0.0307 | 0.7722 | 2.20E-01 | 3.35E-01 |
| Gln | 0.7842 | 5.56E-01 | 0.7698 | 5.85E-01 | -0.0153 | 0.0620 | -0.2470 | 5.98E-01 | 7.16E-01 |
| Gly | -1.1140 | 3.77E-01 | -1.3762 | 2.82E-01 | NA | NA | NA | NA | NA |
| His | -0.2470 | 8.95E-01 | -1.3818 | 2.82E-01 | -0.0316 | 0.1335 | -0.2366 | 4.06E-01 | 5.37E-01 |
| ApoA1 | -2.8719 | 1.02E-02 | -2.3243 | 5.16E-02 | -0.1102 | 0.0220 | -5.0106 | 2.71E-07 | 2.50E-06 |
| ApoB | -0.1775 | 9.18E-01 | 0.2600 | 8.62E-01 | 0.0736 | 0.0211 | 3.4916 | 2.40E-04 | 6.90E-04 |
| ApoB_by_ApoA1 | 1.4149 | 2.49E-01 | 1.4950 | 2.37E-01 | 0.1022 | 0.0246 | 4.1601 | 1.59E-05 | 6.42E-05 |
| Phe | -2.1726 | 5.75E-02 | -1.9435 | 1.16E-01 | 0.0128 | 0.0216 | 0.5923 | 7.23E-01 | 8.05E-01 |
| Tyr | -0.3745 | 8.16E-01 | -0.5873 | 6.97E-01 | 0.0697 | 0.0313 | 2.2245 | 1.31E-02 | 2.78E-02 |
| Ile | 1.0316 | 4.18E-01 | -0.2802 | 8.60E-01 | 0.1269 | 0.0295 | 4.3074 | 8.26E-06 | 3.71E-05 |
| Leu | 0.6110 | 6.67E-01 | 0.0929 | 9.53E-01 | 0.0101 | 0.0121 | 0.8354 | 7.98E-01 | 8.66E-01 |
| Val | 1.0239 | 4.18E-01 | 0.3719 | 8.11E-01 | 0.0058 | 0.0051 | 1.1382 | 8.72E-01 | 9.08E-01 |
| Total_BCAA | 0.9231 | 4.74E-01 | 0.1560 | 9.14E-01 | NA | NA | NA | NA | NA |
| Clinical_LDL_C | -1.3381 | 2.79E-01 | -0.8001 | 5.70E-01 | NA | NA | NA | NA | NA |
| non_HDL_C | -0.6832 | 6.23E-01 | -0.1803 | 9.12E-01 | NA | NA | NA | NA | NA |
| HDL_C | -4.4624 | 5.48E-05 | -3.5233 | 2.87E-03 | -0.1523 | 0.0217 | -7.0049 | 1.24E-12 | 9.47E-11 |
| LDL_C | -0.9500 | 4.60E-01 | -0.9194 | 5.18E-01 | -0.0181 | 0.0620 | -0.2911 | 3.85E-01 | 5.15E-01 |
| Remnant_C | -0.3560 | 8.24E-01 | 0.1239 | 9.31E-01 | 0.0702 | 0.0251 | 2.8003 | 2.55E-03 | 6.29E-03 |
| Total_C | -2.1599 | 5.81E-02 | -1.3300 | 3.01E-01 | -0.0265 | 0.0330 | -0.8039 | 2.11E-01 | 3.23E-01 |
| VLDL_C | 1.7471 | 1.40E-01 | 1.7538 | 1.60E-01 | 0.0970 | 0.0270 | 3.5852 | 1.68E-04 | 5.03E-04 |
| HDL_CE | -4.4766 | 5.40E-05 | -3.5867 | 2.69E-03 | NA | NA | NA | NA | NA |
| LDL_CE | -0.6816 | 6.23E-01 | -0.7215 | 6.14E-01 | NA | NA | NA | NA | NA |
| VLDL_CE | 1.1804 | 3.42E-01 | 1.3288 | 3.01E-01 | NA | NA | NA | NA | NA |
| Total_CE | -2.4087 | 3.47E-02 | -1.5525 | 2.21E-01 | -0.0358 | 0.0269 | -1.3296 | 9.18E-02 | 1.63E-01 |
| XXL_VLDL_C | 4.2348 | 1.12E-04 | 3.6937 | 2.20E-03 | 0.1121 | 0.0273 | 4.1030 | 2.04E-05 | 7.95E-05 |
| XXL_VLDL_CE | 4.1573 | 1.49E-04 | 3.5021 | 3.00E-03 | 0.1103 | 0.0295 | 3.7414 | 9.15E-05 | 2.88E-04 |
| XXL_VLDL_FC | 4.3358 | 8.22E-05 | 3.9354 | 1.48E-03 | 0.1087 | 0.0333 | 3.2666 | 5.44E-04 | 1.49E-03 |
| XXL_VLDL_L | 4.0213 | 2.44E-04 | 3.6268 | 2.65E-03 | 0.1226 | 0.0279 | 4.3861 | 5.77E-06 | 2.79E-05 |
| XXL_VLDL_P | 4.3297 | 8.26E-05 | 3.8712 | 1.67E-03 | 0.1089 | 0.0313 | 3.4740 | 2.56E-04 | 7.28E-04 |
| XXL_VLDL_PL | 4.0786 | 2.01E-04 | 3.8535 | 1.67E-03 | 0.1112 | 0.0306 | 3.6376 | 1.38E-04 | 4.16E-04 |
| XXL_VLDL_TG | 2.3478 | 4.02E-02 | 2.6259 | 2.53E-02 | 0.1021 | 0.0347 | 2.9405 | 1.64E-03 | 4.19E-03 |
| XXL_VLDL_CE_pct | -0.7355 | 5.87E-01 | -0.8555 | 5.49E-01 | -0.0010 | 0.0005 | -1.9741 | 9.76E-01 | 9.76E-01 |
| XXL_VLDL_C_pct | -1.2319 | 3.19E-01 | -1.0722 | 4.33E-01 | 0.0166 | 0.0530 | 0.3139 | 6.23E-01 | 7.35E-01 |
| XXL_VLDL_FC_pct | -0.9143 | 4.78E-01 | -0.3612 | 8.13E-01 | 0.0319 | 0.0775 | 0.4117 | 3.40E-01 | 4.69E-01 |
| XXL_VLDL_PL_pct | 2.7822 | 1.31E-02 | 2.7237 | 2.12E-02 | -0.0095 | 0.0134 | -0.7132 | 7.62E-01 | 8.31E-01 |
| XXL_VLDL_TG_pct | 0.5765 | 6.85E-01 | 0.7020 | 6.23E-01 | -0.0216 | 0.0591 | -0.3644 | 6.42E-01 | 7.42E-01 |
| DHA_pct | -2.0332 | 7.70E-02 | -1.4457 | 2.58E-01 | -0.0241 | 0.1427 | -0.1689 | 5.67E-01 | 7.01E-01 |
| LA_pct | -2.2577 | 4.89E-02 | -1.5382 | 2.24E-01 | -0.0247 | 0.1030 | -0.2398 | 5.95E-01 | 7.16E-01 |
| MUFA_pct | 5.3906 | 5.48E-06 | 3.9769 | 1.44E-03 | 0.0876 | 0.0438 | 1.9989 | 2.28E-02 | 4.68E-02 |
| Omega_3_pct | -0.5084 | 7.39E-01 | 0.1897 | 9.08E-01 | -0.0334 | 0.0961 | -0.3475 | 3.64E-01 | 4.99E-01 |
| Omega_6_pct | -3.2696 | 3.15E-03 | -2.5373 | 3.13E-02 | -0.0752 | 0.0469 | -1.6027 | 5.45E-02 | 1.04E-01 |
| PUFA_pct | -3.4085 | 2.08E-03 | -2.4545 | 3.86E-02 | -0.0763 | 0.0397 | -1.9211 | 2.74E-02 | 5.57E-02 |
| SFA_pct | -0.3980 | 8.04E-01 | -0.5584 | 7.04E-01 | -0.0110 | 0.0611 | -0.1803 | 5.72E-01 | 7.03E-01 |
| Omega_6_by_Omega_3 | -0.3391 | 8.28E-01 | -0.8055 | 5.70E-01 | NA | NA | NA | NA | NA |
| PUFA_by_MUFA | -4.7178 | 3.30E-05 | -3.4471 | 3.44E-03 | NA | NA | NA | NA | NA |

| | | | | | | | | | |
|---|---|---|---|---|---|---|---|---|---|
| DHA | -1.1705 | 3.46E-01 | -0.6512 | 6.58E-01 | -0.0179 | 0.0728 | -0.2466 | 5.97E-01 | 7.16E-01 |
| LA | 0.0660 | 9.60E-01 | 0.3959 | 8.02E-01 | 0.0091 | 0.0094 | 0.9659 | 8.33E-01 | 8.83E-01 |
| MUFA | 3.0200 | 6.77E-03 | 2.4146 | 4.13E-02 | 0.0886 | 0.0204 | 4.3397 | 7.13E-06 | 3.35E-05 |
| Omega_3 | 0.2029 | 9.05E-01 | 0.7401 | 6.02E-01 | 0.0044 | 0.0035 | 1.2455 | 8.94E-01 | 9.15E-01 |
| Omega_6 | -0.2759 | 8.74E-01 | 0.0751 | 9.63E-01 | -0.0133 | 0.0266 | -0.4996 | 6.91E-01 | 7.83E-01 |
| PUFA | -0.1451 | 9.31E-01 | 0.2590 | 8.62E-01 | -0.0120 | 0.0253 | -0.4758 | 6.83E-01 | 7.79E-01 |
| SFA | 1.0805 | 3.92E-01 | 0.9719 | 4.82E-01 | 0.0319 | 0.0363 | 0.8769 | 1.90E-01 | 2.96E-01 |
| Total_FA | 1.3333 | 2.79E-01 | 1.2561 | 3.39E-01 | 0.0502 | 0.0225 | 2.2339 | 1.27E-02 | 2.74E-02 |
| Unsaturation | -3.2611 | 3.16E-03 | -2.1188 | 7.87E-02 | -0.0172 | 0.0255 | -0.6769 | 7.51E-01 | 8.26E-01 |
| Albumin | -1.2549 | 3.11E-01 | -1.3582 | 2.90E-01 | -0.0411 | 0.0310 | -1.3248 | 9.26E-02 | 1.63E-01 |
| Creatinine | 0.2130 | 9.04E-01 | 0.1663 | 9.14E-01 | 0.0038 | 0.0034 | 1.1210 | 8.69E-01 | 9.08E-01 |
| HDL_FC | -4.2133 | 1.21E-04 | -3.1932 | 6.26E-03 | NA | NA | NA | NA | NA |
| LDL_FC | -1.5980 | 1.86E-01 | -1.4208 | 2.69E-01 | NA | NA | NA | NA | NA |
| VLDL_FC | 2.4925 | 2.85E-02 | 2.3186 | 5.19E-02 | NA | NA | NA | NA | NA |
| Total_FC | -1.4961 | 2.22E-01 | -0.7563 | 5.92E-01 | -0.0014 | 0.0009 | -1.6152 | 9.47E-01 | 9.55E-01 |
| Citrate | -3.8896 | 4.03E-04 | -3.1055 | 7.75E-03 | -0.0291 | 2.6604 | -0.0109 | 5.04E-01 | 6.44E-01 |
| Glucose | -2.1636 | 5.80E-02 | -1.0048 | 4.65E-01 | 0.0560 | 0.0368 | 1.5224 | 6.40E-02 | 1.20E-01 |
| Lactate | 0.8173 | 5.38E-01 | -0.0496 | 9.68E-01 | 0.0356 | 0.0397 | 0.8952 | 1.85E-01 | 2.92E-01 |
| Pyruvate | 2.7337 | 1.50E-02 | 1.8743 | 1.32E-01 | NA | NA | NA | NA | NA |
| IDL_C | -2.6507 | 1.85E-02 | -1.7656 | 1.58E-01 | -0.0100 | 0.0297 | -0.3371 | 6.32E-01 | 7.42E-01 |
| IDL_CE | -2.5023 | 2.79E-02 | -1.7277 | 1.65E-01 | 0.0073 | 0.0123 | 0.5969 | 7.25E-01 | 8.05E-01 |
| IDL_FC | -3.0020 | 7.00E-03 | -1.8112 | 1.48E-01 | -0.0383 | 0.0258 | -1.4843 | 6.89E-02 | 1.26E-01 |
| IDL_L | -2.2545 | 4.89E-02 | -1.4153 | 2.70E-01 | -0.0015 | 0.0009 | -1.5865 | 9.44E-01 | 9.55E-01 |
| IDL_P | -1.4321 | 2.43E-01 | -0.7903 | 5.72E-01 | 0.0027 | 0.0022 | 1.2539 | 8.95E-01 | 9.15E-01 |
| IDL_PL | -2.0482 | 7.48E-02 | -1.1598 | 3.88E-01 | -0.0210 | 0.0435 | -0.4821 | 3.15E-01 | 4.44E-01 |
| IDL_TG | 2.2940 | 4.56E-02 | 2.1735 | 7.12E-02 | 0.0681 | 0.0208 | 3.2693 | 5.39E-04 | 1.49E-03 |
| IDL_CE_pct | -2.4284 | 3.31E-02 | -2.3249 | 5.16E-02 | 0.0027 | 0.0017 | 1.5633 | 9.41E-01 | 9.55E-01 |
| IDL_C_pct | -3.4662 | 1.75E-03 | -2.7452 | 2.01E-02 | -0.0526 | 0.0492 | -1.0691 | 1.43E-01 | 2.31E-01 |
| IDL_FC_pct | -3.7787 | 5.95E-04 | -2.0621 | 8.95E-02 | -0.1017 | 0.0349 | -2.9182 | 1.76E-03 | 4.45E-03 |
| IDL_PL_pct | 1.6901 | 1.55E-01 | 1.7177 | 1.66E-01 | -0.1121 | 0.0210 | -5.3431 | 4.57E-08 | 5.90E-07 |
| IDL_TG_pct | 4.4407 | 5.69E-05 | 3.5606 | 2.72E-03 | 0.0854 | 0.0356 | 2.4033 | 8.12E-03 | 1.81E-02 |
| GlycA | 2.1757 | 5.75E-02 | 0.9104 | 5.20E-01 | NA | NA | NA | NA | NA |
| Acetate | -0.3389 | 8.28E-01 | -0.1545 | 9.14E-01 | -0.0893 | 0.0319 | -2.7991 | 2.56E-03 | 6.29E-03 |
| Acetoacetate | -0.4529 | 7.68E-01 | -1.2055 | 3.64E-01 | 0.0728 | 0.0741 | 0.9817 | 1.63E-01 | 2.59E-01 |
| bOHbutyrate | -0.4724 | 7.59E-01 | -0.5928 | 6.96E-01 | -0.0093 | 0.0261 | -0.3552 | 6.39E-01 | 7.42E-01 |
| Acetone | 0.9039 | 4.82E-01 | 0.8811 | 5.35E-01 | NA | NA | NA | NA | NA |
| L_HDL_C | -4.6641 | 3.86E-05 | -3.3331 | 4.44E-03 | -0.1253 | 0.0235 | -5.3315 | 4.87E-08 | 5.90E-07 |
| L_HDL_CE | -4.6069 | 4.07E-05 | -3.3287 | 4.44E-03 | -0.1128 | 0.0266 | -4.2461 | 1.09E-05 | 4.55E-05 |
| L_HDL_FC | -4.4613 | 5.48E-05 | -3.0502 | 8.63E-03 | -0.1307 | 0.0224 | -5.8460 | 2.52E-09 | 4.76E-08 |
| L_HDL_L | -4.6145 | 4.07E-05 | -3.3516 | 4.35E-03 | -0.1256 | 0.0241 | -5.2076 | 9.56E-08 | 1.05E-06 |
| L_HDL_P | -4.6199 | 4.07E-05 | -3.1530 | 6.94E-03 | -0.1252 | 0.0234 | -5.3403 | 4.64E-08 | 5.90E-07 |
| L_HDL_PL | -4.2753 | 9.81E-05 | -3.1930 | 6.26E-03 | -0.1260 | 0.0243 | -5.1903 | 1.05E-07 | 1.10E-06 |
| L_HDL_TG | 0.4434 | 7.72E-01 | 0.3981 | 8.02E-01 | -0.0446 | 0.0301 | -1.4818 | 6.92E-02 | 1.26E-01 |
| L_HDL_CE_pct | -2.4605 | 3.08E-02 | -1.7345 | 1.64E-01 | -0.0920 | 0.0229 | -4.0129 | 3.00E-05 | 1.10E-04 |
| L_HDL_C_pct | -2.6887 | 1.67E-02 | -1.8108 | 1.48E-01 | -0.1034 | 0.0225 | -4.5997 | 2.12E-06 | 1.25E-05 |
| L_HDL_FC_pct | -2.2861 | 4.62E-02 | -1.1390 | 3.99E-01 | -0.0836 | 0.0316 | -2.6429 | 4.11E-03 | 9.45E-03 |
| L_HDL_PL_pct | 4.2874 | 9.58E-05 | 2.4316 | 4.02E-02 | 0.0552 | 0.0361 | 1.5306 | 6.29E-02 | 1.19E-01 |
| L_HDL_TG_pct | 4.4500 | 5.63E-05 | 3.2902 | 4.79E-03 | 0.0936 | 0.0247 | 3.7931 | 7.44E-05 | 2.48E-04 |
| L_LDL_C | -1.2944 | 2.93E-01 | -1.2201 | 3.57E-01 | -0.0158 | 0.1199 | -0.1317 | 4.48E-01 | 5.85E-01 |

| | | | | | | | | | |
|---|---|---|---|---|---|---|---|---|---|
| L_LDL_CE | -1.1840 | 3.42E-01 | -1.0036 | 4.65E-01 | -0.0045 | 0.0047 | -0.9495 | 8.29E-01 | 8.83E-01 |
| L_LDL_FC | -1.7288 | 1.44E-01 | -1.5057 | 2.37E-01 | -0.0444 | 0.0245 | -1.8149 | 3.48E-02 | 6.89E-02 |
| L_LDL_L | -1.2563 | 3.11E-01 | -0.7073 | 6.22E-01 | -0.0106 | 0.0373 | -0.2830 | 6.11E-01 | 7.25E-01 |
| L_LDL_P | -0.2201 | 9.04E-01 | 0.1290 | 9.31E-01 | -0.0077 | 0.0140 | -0.5542 | 7.10E-01 | 7.97E-01 |
| L_LDL_PL | -1.4005 | 2.53E-01 | -0.8094 | 5.70E-01 | -0.0206 | 0.0456 | -0.4506 | 3.26E-01 | 4.55E-01 |
| L_LDL_TG | 1.8150 | 1.23E-01 | 1.7666 | 1.58E-01 | 0.0384 | 0.0240 | 1.5986 | 5.50E-02 | 1.04E-01 |
| L_LDL_CE_pct | -0.3665 | 8.19E-01 | -0.7927 | 5.72E-01 | 0.0040 | 0.0032 | 1.2504 | 8.94E-01 | 9.15E-01 |
| L_LDL_C_pct | -0.7490 | 5.80E-01 | -1.0499 | 4.43E-01 | -0.0572 | 0.0513 | -1.1151 | 1.32E-01 | 2.21E-01 |
| L_LDL_FC_pct | -1.3337 | 2.79E-01 | -1.3969 | 2.77E-01 | -0.0895 | 0.0322 | -2.7814 | 2.71E-03 | 6.55E-03 |
| L_LDL_PL_pct | -0.6387 | 6.51E-01 | -0.4836 | 7.50E-01 | 0.0065 | 0.0093 | 0.6933 | 7.56E-01 | 8.28E-01 |
| L_LDL_TG_pct | 3.2774 | 3.11E-03 | 2.6638 | 2.33E-02 | 0.0651 | 0.0384 | 1.6941 | 4.51E-02 | 8.87E-02 |
| L_VLDL_C | 3.8522 | 4.55E-04 | 3.1432 | 7.05E-03 | 0.1261 | 0.0255 | 4.9338 | 4.03E-07 | 3.31E-06 |
| L_VLDL_CE | 3.3965 | 2.15E-03 | 2.7484 | 2.01E-02 | 0.1392 | 0.0208 | 6.6906 | 1.11E-11 | 5.11E-10 |
| L_VLDL_FC | 4.3035 | 9.10E-05 | 3.5459 | 2.78E-03 | 0.1110 | 0.0301 | 3.6891 | 1.13E-04 | 3.50E-04 |
| L_VLDL_L | 4.4366 | 5.69E-05 | 3.6490 | 2.52E-03 | 0.1251 | 0.0246 | 5.0808 | 1.88E-07 | 1.80E-06 |
| L_VLDL_P | 4.5151 | 5.08E-05 | 3.7254 | 2.02E-03 | 0.1167 | 0.0282 | 4.1409 | 1.73E-05 | 6.86E-05 |
| L_VLDL_PL | 3.6896 | 8.22E-04 | 2.5176 | 3.27E-02 | 0.1226 | 0.0270 | 4.5357 | 2.87E-06 | 1.52E-05 |
| L_VLDL_TG | 4.5989 | 4.07E-05 | 3.7258 | 2.02E-03 | 0.1226 | 0.0253 | 4.8558 | 5.99E-07 | 4.60E-06 |
| L_VLDL_CE_pct | -3.0529 | 6.13E-03 | -2.6298 | 2.53E-02 | 0.0009 | 0.0004 | 1.9306 | 9.73E-01 | 9.76E-01 |
| L_VLDL_C_pct | -1.9639 | 9.00E-02 | -1.7166 | 1.66E-01 | 0.0048 | 0.0039 | 1.2170 | 8.88E-01 | 9.15E-01 |
| L_VLDL_FC_pct | 1.2336 | 3.19E-01 | 1.0108 | 4.65E-01 | 0.0149 | 0.0310 | 0.4792 | 6.84E-01 | 7.79E-01 |
| L_VLDL_PL_pct | 2.2547 | 4.89E-02 | 1.1187 | 4.05E-01 | 0.0041 | 0.0040 | 1.0085 | 8.43E-01 | 8.90E-01 |
| L_VLDL_TG_pct | -0.0239 | 9.85E-01 | -0.2702 | 8.62E-01 | -0.0198 | 0.0870 | -0.2276 | 5.90E-01 | 7.16E-01 |
| HDL_P | -2.3435 | 4.03E-02 | -1.9361 | 1.16E-01 | NA | NA | NA | NA | NA |
| LDL_P | -0.1938 | 9.08E-01 | 0.2534 | 8.62E-01 | NA | NA | NA | NA | NA |
| Total_P | -2.2516 | 4.89E-02 | -1.7578 | 1.59E-01 | NA | NA | NA | NA | NA |
| VLDL_P | 2.2149 | 5.29E-02 | 2.1186 | 7.87E-02 | NA | NA | NA | NA | NA |
| HDL_size | -4.7770 | 2.95E-05 | -3.5915 | 2.69E-03 | -0.0958 | 0.0272 | -3.5277 | 2.10E-04 | 6.10E-04 |
| LDL_size | -0.1084 | 9.40E-01 | -0.5648 | 7.02E-01 | 0.0242 | 0.0488 | 0.4948 | 3.10E-01 | 4.42E-01 |
| VLDL_size | 4.9015 | 2.37E-05 | 3.8887 | 1.67E-03 | 0.1195 | 0.0254 | 4.6995 | 1.30E-06 | 8.82E-06 |
| M_HDL_C | -3.8326 | 4.86E-04 | -3.4975 | 3.00E-03 | -0.1173 | 0.0211 | -5.5587 | 1.36E-08 | 2.08E-07 |
| M_HDL_CE | -3.8594 | 4.49E-04 | -3.5656 | 2.72E-03 | -0.1121 | 0.0211 | -5.3190 | 5.22E-08 | 6.00E-07 |
| M_HDL_FC | -3.6101 | 1.07E-03 | -3.1385 | 7.05E-03 | -0.1045 | 0.0225 | -4.6405 | 1.74E-06 | 1.11E-05 |
| M_HDL_L | -2.1395 | 6.06E-02 | -1.8991 | 1.26E-01 | -0.0939 | 0.0214 | -4.3838 | 5.83E-06 | 2.79E-05 |
| M_HDL_P | -3.3483 | 2.50E-03 | -3.0790 | 8.08E-03 | -0.0850 | 0.0225 | -3.7823 | 7.77E-05 | 2.55E-04 |
| M_HDL_PL | -1.4982 | 2.22E-01 | -1.3769 | 2.82E-01 | -0.0972 | 0.0217 | -4.4710 | 3.89E-06 | 1.99E-05 |
| M_HDL_TG | 3.5756 | 1.21E-03 | 2.7780 | 1.89E-02 | 0.0583 | 0.0462 | 1.2608 | 1.04E-01 | 1.77E-01 |
| M_HDL_CE_pct | -3.6326 | 9.98E-04 | -4.1917 | 1.07E-03 | -0.0964 | 0.0272 | -3.5392 | 2.01E-04 | 5.92E-04 |
| M_HDL_C_pct | -3.7219 | 7.35E-04 | -4.2357 | 1.07E-03 | -0.1195 | 0.0258 | -4.6301 | 1.83E-06 | 1.14E-05 |
| M_HDL_FC_pct | -4.3532 | 7.95E-05 | -4.3714 | 1.02E-03 | -0.0713 | 0.0436 | -1.6362 | 5.09E-02 | 9.84E-02 |
| M_HDL_PL_pct | 5.8070 | 1.58E-06 | 4.7630 | 4.75E-04 | 0.0411 | 0.0365 | 1.1277 | 1.30E-01 | 2.18E-01 |
| M_HDL_TG_pct | 4.9109 | 2.37E-05 | 3.9611 | 1.44E-03 | 0.1273 | 0.0269 | 4.7362 | 1.09E-06 | 7.83E-06 |
| M_LDL_C | -0.3121 | 8.47E-01 | -0.3789 | 8.09E-01 | -0.0111 | 0.0486 | -0.2282 | 5.90E-01 | 7.16E-01 |
| M_LDL_CE | -0.0646 | 9.60E-01 | -0.2350 | 8.74E-01 | -0.0056 | 0.0058 | -0.9640 | 8.32E-01 | 8.83E-01 |
| M_LDL_FC | -1.1911 | 3.40E-01 | -1.1226 | 4.05E-01 | -0.0212 | 0.0422 | -0.5022 | 3.08E-01 | 4.42E-01 |
| M_LDL_L | 0.1227 | 9.39E-01 | 0.3829 | 8.09E-01 | -0.0106 | 0.0385 | -0.2758 | 6.09E-01 | 7.25E-01 |
| M_LDL_P | -0.1116 | 9.40E-01 | 0.4743 | 7.50E-01 | -0.0090 | 0.0211 | -0.4264 | 6.65E-01 | 7.65E-01 |
| M_LDL_PL | 0.0964 | 9.46E-01 | 0.2790 | 8.60E-01 | 0.0071 | 0.0116 | 0.6127 | 7.30E-01 | 8.07E-01 |
| M_LDL_TG | 2.3899 | 3.62E-02 | 2.2282 | 6.31E-02 | 0.0303 | 0.0273 | 1.1079 | 1.34E-01 | 2.22E-01 |

| | | | | | | | | | |
|---|---|---|---|---|---|---|---|---|---|
| M_LDL_CE_pct | -0.1428 | 9.31E-01 | -0.5788 | 6.97E-01 | -0.0292 | 0.0285 | -1.0242 | 1.53E-01 | 2.44E-01 |
| M_LDL_C_pct | -0.5965 | 6.72E-01 | -0.9095 | 5.20E-01 | -0.0455 | 0.0305 | -1.4909 | 6.80E-02 | 1.26E-01 |
| M_LDL_FC_pct | -1.5458 | 2.04E-01 | -1.6711 | 1.80E-01 | -0.0197 | 0.0446 | -0.4415 | 3.29E-01 | 4.56E-01 |
| M_LDL_PL_pct | -0.1197 | 9.39E-01 | -0.4815 | 7.50E-01 | 0.0298 | 0.0339 | 0.8806 | 1.89E-01 | 2.96E-01 |
| M_LDL_TG_pct | 2.7222 | 1.54E-02 | 2.2444 | 6.12E-02 | 0.0600 | 0.0364 | 1.6456 | 4.99E-02 | 9.73E-02 |
| M_VLDL_C | -0.1238 | 9.39E-01 | 0.3950 | 8.02E-01 | 0.1251 | 0.0214 | 5.8349 | 2.69E-09 | 4.76E-08 |
| M_VLDL_CE | -0.6183 | 6.64E-01 | -0.0688 | 9.65E-01 | 0.1058 | 0.0263 | 4.0231 | 2.87E-05 | 1.07E-04 |
| M_VLDL_FC | 1.0989 | 3.82E-01 | 1.2541 | 3.39E-01 | 0.1189 | 0.0255 | 4.6588 | 1.59E-06 | 1.04E-05 |
| M_VLDL_L | 2.5077 | 2.78E-02 | 2.2671 | 5.82E-02 | 0.1165 | 0.0257 | 4.5333 | 2.90E-06 | 1.52E-05 |
| M_VLDL_P | 1.8653 | 1.12E-01 | 1.8224 | 1.47E-01 | 0.1152 | 0.0271 | 4.2463 | 1.09E-05 | 4.55E-05 |
| M_VLDL_PL | 1.7882 | 1.29E-01 | 1.7423 | 1.62E-01 | 0.1128 | 0.0282 | 3.9966 | 3.21E-05 | 1.14E-04 |
| M_VLDL_TG | 4.2416 | 1.11E-04 | 3.5243 | 2.87E-03 | 0.1346 | 0.0226 | 5.9462 | 1.37E-09 | 2.87E-08 |
| M_VLDL_CE_pct | -2.8792 | 1.00E-02 | -1.9792 | 1.07E-01 | -0.0184 | 0.0540 | -0.3405 | 3.67E-01 | 4.99E-01 |
| M_VLDL_C_pct | -4.3406 | 8.22E-05 | -3.0113 | 9.67E-03 | -0.0398 | 0.0369 | -1.0785 | 1.40E-01 | 2.29E-01 |
| M_VLDL_FC_pct | -4.0342 | 2.35E-04 | -2.8138 | 1.72E-02 | 0.0423 | 0.0318 | 1.3274 | 9.22E-02 | 1.63E-01 |
| M_VLDL_PL_pct | -2.1926 | 5.56E-02 | -1.5991 | 2.06E-01 | -0.0398 | 0.0639 | -0.6230 | 2.67E-01 | 3.91E-01 |
| M_VLDL_TG_pct | 5.5348 | 3.88E-06 | 3.9598 | 1.44E-03 | 0.0449 | 0.0241 | 1.8609 | 3.14E-02 | 6.28E-02 |
| Cholines | -1.5484 | 2.04E-01 | -1.0080 | 4.65E-01 | -0.0046 | 0.0050 | -0.9220 | 8.22E-01 | 8.83E-01 |
| Phosphatidylc | -1.0253 | 4.18E-01 | -0.5477 | 7.09E-01 | -0.0163 | 0.1036 | -0.1571 | 4.38E-01 | 5.75E-01 |
| Sphingomyelins | -3.3342 | 2.60E-03 | -2.6687 | 2.33E-02 | -0.0369 | 0.0271 | -1.3607 | 8.68E-02 | 1.56E-01 |
| TG_by_PG | 5.3420 | 5.48E-06 | 4.3874 | 1.02E-03 | 0.1409 | 0.0228 | 6.1839 | 3.13E-10 | 8.99E-09 |
| Phosphoglyc | -1.0574 | 4.04E-01 | -0.6341 | 6.68E-01 | NA | NA | NA | NA | NA |
| HDL_PL | -2.8594 | 1.04E-02 | -2.2885 | 5.56E-02 | NA | NA | NA | NA | NA |
| LDL_PL | -0.9614 | 4.55E-01 | -0.4558 | 7.62E-01 | NA | NA | NA | NA | NA |
| Total_PL | -1.4370 | 2.43E-01 | -0.8040 | 5.70E-01 | NA | NA | NA | NA | NA |
| VLDL_PL | 2.9980 | 7.00E-03 | 2.6647 | 2.33E-02 | NA | NA | NA | NA | NA |
| S_HDL_C | 0.2126 | 9.04E-01 | -0.0169 | 9.86E-01 | -0.1265 | 0.0207 | -6.0976 | 5.38E-10 | 1.24E-08 |
| S_HDL_CE | 0.3513 | 8.25E-01 | 0.0229 | 9.86E-01 | -0.1048 | 0.0211 | -4.9579 | 3.56E-07 | 3.15E-06 |
| S_HDL_FC | -0.2082 | 9.04E-01 | -0.1612 | 9.14E-01 | -0.0579 | 0.0214 | -2.7072 | 3.39E-03 | 7.88E-03 |
| S_HDL_L | 1.3108 | 2.88E-01 | 0.8739 | 5.38E-01 | -0.0601 | 0.0215 | -2.7979 | 2.57E-03 | 6.29E-03 |
| S_HDL_P | 0.6440 | 6.50E-01 | 0.3642 | 8.13E-01 | -0.0426 | 0.0227 | -1.8778 | 3.02E-02 | 6.09E-02 |
| S_HDL_PL | 1.2971 | 2.93E-01 | 0.8304 | 5.62E-01 | -0.0127 | 0.3597 | -0.0352 | 5.14E-01 | 6.50E-01 |
| S_HDL_TG | 4.7421 | 3.10E-05 | 3.8453 | 1.67E-03 | 0.1231 | 0.0252 | 4.8740 | 5.47E-07 | 4.34E-06 |
| S_HDL_CE_pct | -1.8174 | 1.23E-01 | -1.6496 | 1.87E-01 | -0.0944 | 0.0378 | -2.4946 | 6.30E-03 | 1.44E-02 |
| S_HDL_C_pct | -2.9784 | 7.36E-03 | -2.4175 | 4.13E-02 | -0.0644 | 0.0506 | -1.2743 | 1.01E-01 | 1.75E-01 |
| S_HDL_FC_pct | -4.0341 | 2.35E-04 | -2.7479 | 2.01E-02 | -0.0518 | 0.0409 | -1.2658 | 1.03E-01 | 1.76E-01 |
| S_HDL_PL_pct | 0.0782 | 9.57E-01 | -0.1692 | 9.14E-01 | 0.0261 | 0.0794 | 0.3288 | 3.71E-01 | 4.99E-01 |
| S_HDL_TG_pct | 4.9156 | 2.37E-05 | 4.0802 | 1.40E-03 | 0.1600 | 0.0204 | 7.8590 | 1.94E-15 | 4.45E-13 |
| S_LDL_C | -0.6077 | 6.67E-01 | -0.6279 | 6.70E-01 | -0.0245 | 0.0343 | -0.7128 | 2.38E-01 | 3.56E-01 |
| S_LDL_CE | -0.2355 | 9.01E-01 | -0.3542 | 8.15E-01 | -0.0229 | 0.0375 | -0.6121 | 2.70E-01 | 3.93E-01 |
| S_LDL_FC | -1.3965 | 2.53E-01 | -1.1996 | 3.65E-01 | -0.0236 | 0.0408 | -0.5783 | 2.82E-01 | 4.07E-01 |
| S_LDL_L | -0.4021 | 8.04E-01 | 0.0590 | 9.68E-01 | -0.0128 | 0.1418 | -0.0901 | 5.36E-01 | 6.74E-01 |
| S_LDL_P | -0.2088 | 9.04E-01 | 0.3196 | 8.33E-01 | -0.0078 | 0.0148 | -0.5318 | 7.03E-01 | 7.92E-01 |
| S_LDL_PL | -1.1091 | 3.78E-01 | -0.3498 | 8.15E-01 | -0.0146 | 0.2965 | -0.0493 | 4.80E-01 | 6.17E-01 |
| S_LDL_TG | 3.2389 | 3.32E-03 | 2.8762 | 1.43E-02 | 0.0667 | 0.0212 | 3.1508 | 8.14E-04 | 2.18E-03 |
| S_LDL_CE_pct | 0.0179 | 9.86E-01 | -0.5293 | 7.21E-01 | -0.0439 | 0.0334 | -1.3149 | 9.43E-02 | 1.64E-01 |
| S_LDL_C_pct | -0.4974 | 7.45E-01 | -0.8845 | 5.35E-01 | -0.0208 | 0.1240 | -0.1677 | 5.67E-01 | 7.01E-01 |
| S_LDL_FC_pct | -1.4341 | 2.43E-01 | -1.4985 | 2.37E-01 | -0.0187 | 0.1709 | -0.1094 | 4.56E-01 | 5.90E-01 |
| S_LDL_PL_pct | -2.4475 | 3.17E-02 | -1.5020 | 2.37E-01 | 0.0208 | 0.0423 | 0.4914 | 3.12E-01 | 4.42E-01 |

| | | | | | | | | | |
|---|---|---|---|---|---|---|---|---|---|
| S_LDL_TG_pct | 4.1736 | 1.41E-04 | 3.3719 | 4.13E-03 | 0.0926 | 0.0308 | 3.0114 | 1.30E-03 | 3.36E-03 |
| S_VLDL_C | 1.4745 | 2.30E-01 | 1.5448 | 2.22E-01 | 0.0894 | 0.0243 | 3.6717 | 1.20E-04 | 3.69E-04 |
| S_VLDL_CE | 1.8472 | 1.16E-01 | 1.8060 | 1.48E-01 | 0.0899 | 0.0209 | 4.3041 | 8.38E-06 | 3.71E-05 |
| S_VLDL_FC | 0.8155 | 5.38E-01 | 1.0566 | 4.41E-01 | 0.1041 | 0.0221 | 4.7106 | 1.23E-06 | 8.61E-06 |
| S_VLDL_L | 2.8653 | 1.03E-02 | 2.5831 | 2.80E-02 | 0.1118 | 0.0244 | 4.5844 | 2.28E-06 | 1.28E-05 |
| S_VLDL_P | 2.9970 | 7.00E-03 | 2.6887 | 2.29E-02 | 0.1202 | 0.0221 | 5.4382 | 2.69E-08 | 3.87E-07 |
| S_VLDL_PL | 1.7326 | 1.44E-01 | 1.7082 | 1.68E-01 | 0.1068 | 0.0233 | 4.5906 | 2.21E-06 | 1.27E-05 |
| S_VLDL_TG | 4.4861 | 5.40E-05 | 3.8150 | 1.72E-03 | 0.1141 | 0.0274 | 4.1601 | 1.59E-05 | 6.42E-05 |
| S_VLDL_CE_pct | -2.2384 | 5.02E-02 | -1.5907 | 2.06E-01 | -0.0368 | 0.0354 | -1.0407 | 1.49E-01 | 2.40E-01 |
| S_VLDL_C_pct | -3.5214 | 1.46E-03 | -2.5651 | 2.92E-02 | -0.0562 | 0.0250 | -2.2496 | 1.22E-02 | 2.66E-02 |
| S_VLDL_FC_pct | -4.4791 | 5.40E-05 | -3.3286 | 4.44E-03 | -0.1384 | 0.0211 | -6.5704 | 2.51E-11 | 9.62E-10 |
| S_VLDL_PL_pct | -4.5585 | 4.51E-05 | -3.5595 | 2.72E-03 | -0.1144 | 0.0205 | -5.5684 | 1.29E-08 | 2.08E-07 |
| S_VLDL_TG_pct | 4.5551 | 4.51E-05 | 3.6146 | 2.67E-03 | 0.1041 | 0.0211 | 4.9444 | 3.82E-07 | 3.25E-06 |
| HDL_L | -3.4353 | 1.91E-03 | -2.7125 | 2.16E-02 | NA | NA | NA | NA | NA |
| LDL_L | -0.8005 | 5.46E-01 | -0.3326 | 8.26E-01 | NA | NA | NA | NA | NA |
| Total_L | -0.3825 | 8.13E-01 | 0.0516 | 9.68E-01 | NA | NA | NA | NA | NA |
| VLDL_L | 3.4629 | 1.75E-03 | 2.9928 | 1.01E-02 | NA | NA | NA | NA | NA |
| HDL_TG | 3.2633 | 3.16E-03 | 2.5847 | 2.80E-02 | 0.0836 | 0.0339 | 2.4652 | 6.85E-03 | 1.54E-02 |
| LDL_TG | 2.1684 | 5.77E-02 | 2.0483 | 9.17E-02 | 0.0484 | 0.0224 | 2.1626 | 1.53E-02 | 3.20E-02 |
| Total_TG | 4.2728 | 9.81E-05 | 3.5941 | 2.69E-03 | 0.1082 | 0.0271 | 3.9913 | 3.29E-05 | 1.15E-04 |
| VLDL_TG | 4.5475 | 4.51E-05 | 3.8116 | 1.72E-03 | 0.1086 | 0.0289 | 3.7548 | 8.68E-05 | 2.77E-04 |
| XL_HDL_C | -5.3094 | 5.48E-06 | -3.4107 | 3.84E-03 | -0.0689 | 0.0252 | -2.7318 | 3.15E-03 | 7.42E-03 |
| XL_HDL_CE | -4.6846 | 3.68E-05 | -3.0651 | 8.34E-03 | -0.0598 | 0.0275 | -2.1778 | 1.47E-02 | 3.10E-02 |
| XL_HDL_FC | -4.8481 | 2.39E-05 | -3.1873 | 6.27E-03 | -0.0924 | 0.0214 | -4.3076 | 8.25E-06 | 3.71E-05 |
| XL_HDL_L | -4.8587 | 2.39E-05 | -3.2191 | 5.93E-03 | -0.1005 | 0.0218 | -4.6060 | 2.05E-06 | 1.24E-05 |
| XL_HDL_P | -5.0560 | 1.78E-05 | -3.3852 | 4.05E-03 | -0.0933 | 0.0218 | -4.2779 | 9.43E-06 | 4.09E-05 |
| XL_HDL_PL | -3.1049 | 5.21E-03 | -1.5936 | 2.06E-01 | -0.1122 | 0.0218 | -5.1541 | 1.27E-07 | 1.27E-06 |
| XL_HDL_TG | 0.9430 | 4.63E-01 | 0.9951 | 4.68E-01 | 0.0668 | 0.0215 | 3.1087 | 9.39E-04 | 2.48E-03 |
| XL_HDL_CE_pct | -1.4589 | 2.35E-01 | -0.8158 | 5.70E-01 | 0.0320 | 0.1130 | 0.2832 | 3.88E-01 | 5.16E-01 |
| XL_HDL_C_pct | -0.1448 | 9.31E-01 | 0.5825 | 6.97E-01 | 0.0306 | 0.2671 | 0.1147 | 4.54E-01 | 5.90E-01 |
| XL_HDL_FC_pct | 3.0057 | 7.00E-03 | 2.1918 | 6.86E-02 | -0.0230 | 0.9432 | -0.0244 | 5.10E-01 | 6.48E-01 |
| XL_HDL_PL_pct | -0.4835 | 7.53E-01 | 0.4768 | 7.50E-01 | -0.0613 | 0.0258 | -2.3739 | 8.80E-03 | 1.95E-02 |
| XL_HDL_TG_pct | 4.7843 | 2.95E-05 | 3.4595 | 3.37E-03 | 0.1273 | 0.0208 | 6.1084 | 5.03E-10 | 1.24E-08 |
| XL_VLDL_C | 3.6622 | 9.03E-04 | 2.8831 | 1.42E-02 | 0.1437 | 0.0205 | 7.0112 | 1.18E-12 | 9.47E-11 |
| XL_VLDL_CE | 3.2499 | 3.23E-03 | 2.4510 | 3.86E-02 | 0.1407 | 0.0205 | 6.8559 | 3.54E-12 | 2.04E-10 |
| XL_VLDL_FC | 4.1027 | 1.85E-04 | 3.3834 | 4.05E-03 | 0.1249 | 0.0263 | 4.7509 | 1.01E-06 | 7.51E-06 |
| XL_VLDL_L | 4.5472 | 4.51E-05 | 3.7304 | 2.02E-03 | 0.1214 | 0.0268 | 4.5383 | 2.83E-06 | 1.52E-05 |
| XL_VLDL_P | 4.5996 | 4.07E-05 | 3.7629 | 1.99E-03 | 0.1078 | 0.0325 | 3.3142 | 4.59E-04 | 1.29E-03 |
| XL_VLDL_PL | 3.9766 | 2.90E-04 | 3.0973 | 7.84E-03 | 0.1154 | 0.0292 | 3.9492 | 3.92E-05 | 1.35E-04 |
| XL_VLDL_TG | 4.8497 | 2.39E-05 | 3.9727 | 1.44E-03 | 0.1134 | 0.0295 | 3.8436 | 6.06E-05 | 2.05E-04 |
| XL_VLDL_CE_pct | -4.6290 | 4.07E-05 | -4.1725 | 1.07E-03 | 0.0298 | 0.0273 | 1.0909 | 1.38E-01 | 2.26E-01 |
| XL_VLDL_C_pct | -4.7431 | 3.10E-05 | -4.2180 | 1.07E-03 | 0.0236 | 0.0718 | 0.3292 | 3.71E-01 | 4.99E-01 |
| XL_VLDL_FC_pct | -3.4972 | 1.58E-03 | -2.6617 | 2.33E-02 | 0.0284 | 0.0604 | 0.4698 | 3.19E-01 | 4.48E-01 |
| XL_VLDL_PL_pct | 2.0940 | 6.74E-02 | 2.1350 | 7.76E-02 | 0.0537 | 0.0228 | 2.3544 | 9.28E-03 | 2.03E-02 |
| XL_VLDL_TG_pct | 3.9728 | 2.90E-04 | 3.3047 | 4.64E-03 | 0.0064 | 0.0057 | 1.1258 | 8.70E-01 | 9.08E-01 |
| XS_VLDL_C | -0.9979 | 4.33E-01 | -0.2562 | 8.62E-01 | 0.0383 | 0.0276 | 1.3876 | 8.26E-02 | 1.50E-01 |
| XS_VLDL_CE | -1.3959 | 2.53E-01 | -0.5703 | 7.01E-01 | 0.0392 | 0.0333 | 1.1757 | 1.20E-01 | 2.03E-01 |
| XS_VLDL_FC | -0.0389 | 9.77E-01 | 0.5103 | 7.34E-01 | 0.0239 | 0.0335 | 0.7126 | 2.38E-01 | 3.56E-01 |
| XS_VLDL_L | 0.4559 | 7.68E-01 | 0.8448 | 5.54E-01 | 0.0694 | 0.0214 | 3.2444 | 5.89E-04 | 1.59E-03 |

| | | | | | | | | | |
|---|---|---|---|---|---|---|---|---|---|
| XS_VLDL_P | 0.1704 | 9.20E-01 | 0.6517 | 6.58E-01 | 0.0789 | 0.0210 | 3.7630 | 8.40E-05 | 2.72E-04 |
| XS_VLDL_PL | 0.8156 | 5.38E-01 | 1.1300 | 4.02E-01 | 0.0261 | 0.0313 | 0.8327 | 2.02E-01 | 3.13E-01 |
| XS_VLDL_TG | 3.3917 | 2.16E-03 | 3.0789 | 8.08E-03 | 0.1005 | 0.0251 | 4.0061 | 3.09E-05 | 1.11E-04 |
| XS_VLDL_CE_pct | -4.4158 | 6.11E-05 | -3.2712 | 5.03E-03 | -0.0391 | 0.0562 | -0.6951 | 2.43E-01 | 3.61E-01 |
| XS_VLDL_C_pct | -4.4803 | 5.40E-05 | -3.3134 | 4.59E-03 | -0.0422 | 0.0651 | -0.6475 | 2.59E-01 | 3.81E-01 |
| XS_VLDL_FC_pct | -3.2983 | 2.92E-03 | -2.1318 | 7.76E-02 | -0.1013 | 0.0367 | -2.7597 | 2.89E-03 | 6.93E-03 |
| XS_VLDL_PL_pct | 3.2590 | 3.16E-03 | 2.6814 | 2.31E-02 | -0.1367 | 0.0214 | -6.3892 | 8.34E-11 | 2.74E-09 |
| XS_VLDL_TG_pct | 4.9455 | 2.37E-05 | 3.9743 | 1.44E-03 | 0.0827 | 0.0272 | 3.0451 | 1.16E-03 | 3.04E-03 |

**Supplementary Table 4**

**Novel Findings**

| | UK Biobank | | | | BBMRI-NL | | | | |
|---|---|---|---|---|---|---|---|---|---|
| metabolite | Z_MDD | FDR_MDD | Z_RECURRENT | FDR_RECURRENT | Effect | SE | Z | Pval | FDR |
| Ala | 2.7183 | 1.54E-02 | 1.7793 | 1.56E-01 | 0.0237 | 0.0307 | 0.7722 | 2.20E-01 | 3.35E-01 |
| HDL_CE | -4.4766 | 5.40E-05 | -3.5867 | 2.69E-03 | NA | NA | NA | NA | NA |
| Total_CE | -2.4087 | 3.47E-02 | -1.5525 | 2.21E-01 | -0.0358 | 0.0269 | -1.3296 | 9.18E-02 | 1.63E-01 |
| XXL_VLDL_PL_pct | 2.7822 | 1.31E-02 | 2.7237 | 2.12E-02 | -0.0095 | 0.0134 | -0.7132 | 7.62E-01 | 8.31E-01 |
| LA_pct | -2.2577 | 4.89E-02 | -1.5382 | 2.24E-01 | -0.0247 | 0.1030 | -0.2398 | 5.95E-01 | 7.16E-01 |
| Omega_6_pct | -3.2696 | 3.15E-03 | -2.5373 | 3.13E-02 | -0.0752 | 0.0469 | -1.6027 | 5.45E-02 | 1.04E-01 |
| PUFA_pct | -3.4085 | 2.08E-03 | -2.4545 | 3.86E-02 | -0.0763 | 0.0397 | -1.9211 | 2.74E-02 | 5.57E-02 |
| PUFA_by_MUFA | -4.7178 | 3.30E-05 | -3.4471 | 3.44E-03 | NA | NA | NA | NA | NA |
| Unsaturation | -3.2611 | 3.16E-03 | -2.1188 | 7.87E-02 | -0.0172 | 0.0255 | -0.6769 | 7.51E-01 | 8.26E-01 |
| HDL_FC | -4.2133 | 1.21E-04 | -3.1932 | 6.26E-03 | NA | NA | NA | NA | NA |
| VLDL_FC | 2.4925 | 2.85E-02 | 2.3186 | 5.19E-02 | NA | NA | NA | NA | NA |
| Citrate | -3.8896 | 4.03E-04 | -3.1055 | 7.75E-03 | -0.0291 | 2.6604 | -0.0109 | 5.04E-01 | 6.44E-01 |
| Pyruvate | 2.7337 | 1.50E-02 | 1.8743 | 1.32E-01 | NA | NA | NA | NA | NA |
| IDL_C | -2.6507 | 1.85E-02 | -1.7656 | 1.58E-01 | -0.0100 | 0.0297 | -0.3371 | 6.32E-01 | 7.42E-01 |
| IDL_CE | -2.5023 | 2.79E-02 | -1.7277 | 1.65E-01 | 0.0073 | 0.0123 | 0.5969 | 7.25E-01 | 8.05E-01 |
| IDL_FC | -3.0020 | 7.00E-03 | -1.8112 | 1.48E-01 | -0.0383 | 0.0258 | -1.4843 | 6.89E-02 | 1.26E-01 |
| IDL_L | -2.2545 | 4.89E-02 | -1.4153 | 2.70E-01 | -0.0015 | 0.0009 | -1.5865 | 9.44E-01 | 9.55E-01 |
| IDL_CE_pct | -2.4284 | 3.31E-02 | -2.3249 | 5.16E-02 | 0.0027 | 0.0017 | 1.5633 | 9.41E-01 | 9.55E-01 |
| IDL_C_pct | -3.4662 | 1.75E-03 | -2.7452 | 2.01E-02 | -0.0526 | 0.0492 | -1.0691 | 1.43E-01 | 2.31E-01 |
| L_HDL_PL_pct | 4.2874 | 9.58E-05 | 2.4316 | 4.02E-02 | 0.0552 | 0.0361 | 1.5306 | 6.29E-02 | 1.19E-01 |
| L_LDL_TG_pct | 3.2774 | 3.11E-03 | 2.6638 | 2.33E-02 | 0.0651 | 0.0384 | 1.6941 | 4.51E-02 | 8.87E-02 |
| L_VLDL_CE_pct | -3.0529 | 6.13E-03 | -2.6298 | 2.53E-02 | 0.0009 | 0.0004 | 1.9306 | 9.73E-01 | 9.76E-01 |
| L_VLDL_PL_pct | 2.2547 | 4.89E-02 | 1.1187 | 4.05E-01 | 0.0041 | 0.0040 | 1.0085 | 8.43E-01 | 8.90E-01 |
| HDL_P | -2.3435 | 4.03E-02 | -1.9361 | 1.16E-01 | NA | NA | NA | NA | NA |
| Total_P | -2.2516 | 4.89E-02 | -1.7578 | 1.59E-01 | NA | NA | NA | NA | NA |
| M_HDL_TG | 3.5756 | 1.21E-03 | 2.7780 | 1.89E-02 | 0.0583 | 0.0462 | 1.2608 | 1.04E-01 | 1.77E-01 |
| M_HDL_FC_pct | -4.3532 | 7.95E-05 | -4.3714 | 1.02E-03 | -0.0713 | 0.0436 | -1.6362 | 5.09E-02 | 9.84E-02 |
| M_HDL_PL_pct | 5.8070 | 1.58E-06 | 4.7630 | 4.75E-04 | 0.0411 | 0.0365 | 1.1277 | 1.30E-01 | 2.18E-01 |
| M_LDL_TG | 2.3899 | 3.62E-02 | 2.2282 | 6.31E-02 | 0.0303 | 0.0273 | 1.1079 | 1.34E-01 | 2.22E-01 |
| M_LDL_TG_pct | 2.7222 | 1.54E-02 | 2.2444 | 6.12E-02 | 0.0600 | 0.0364 | 1.6456 | 4.99E-02 | 9.73E-02 |
| M_VLDL_CE_pct | -2.8792 | 1.00E-02 | -1.9792 | 1.07E-01 | -0.0184 | 0.0540 | -0.3405 | 3.67E-01 | 4.99E-01 |
| M_VLDL_C_pct | -4.3406 | 8.22E-05 | -3.0113 | 9.67E-03 | -0.0398 | 0.0369 | -1.0785 | 1.40E-01 | 2.29E-01 |
| M_VLDL_FC_pct | -4.0342 | 2.35E-04 | -2.8138 | 1.72E-02 | 0.0423 | 0.0318 | 1.3274 | 9.22E-02 | 1.63E-01 |
| M_VLDL_TG_pct | 5.5348 | 3.88E-06 | 3.9598 | 1.44E-03 | 0.0449 | 0.0241 | 1.8609 | 3.14E-02 | 6.28E-02 |
| Sphingomyelins | -3.3342 | 2.60E-03 | -2.6687 | 2.33E-02 | -0.0369 | 0.0271 | -1.3607 | 8.68E-02 | 1.56E-01 |
| HDL_PL | -2.8594 | 1.04E-02 | -2.2885 | 5.56E-02 | NA | NA | NA | NA | NA |
| VLDL_PL | 2.9980 | 7.00E-03 | 2.6647 | 2.33E-02 | NA | NA | NA | NA | NA |
| S_HDL_C_pct | -2.9784 | 7.36E-03 | -2.4175 | 4.13E-02 | -0.0644 | 0.0506 | -1.2743 | 1.01E-01 | 1.75E-01 |
| S_HDL_FC_pct | -4.0341 | 2.35E-04 | -2.7479 | 2.01E-02 | -0.0518 | 0.0409 | -1.2658 | 1.03E-01 | 1.76E-01 |
| S_LDL_PL_pct | -2.4475 | 3.17E-02 | -1.5020 | 2.37E-01 | 0.0208 | 0.0423 | 0.4914 | 3.12E-01 | 4.42E-01 |
| HDL_L | -3.4353 | 1.91E-03 | -2.7125 | 2.16E-02 | NA | NA | NA | NA | NA |
| VLDL_L | 3.4629 | 1.75E-03 | 2.9928 | 1.01E-02 | NA | NA | NA | NA | NA |

| | | | | | | | | | |
|---|---|---|---|---|---|---|---|---|---|
| XL_HDL_FC_pct | 3.0057 | 7.00E-03 | 2.1918 | 6.86E-02 | -0.0230 | 0.9432 | -0.0244 | 5.10E-01 | 6.48E-01 |
| XL_VLDL_CE_pct | -4.6290 | 4.07E-05 | -4.1725 | 1.07E-03 | 0.0298 | 0.0273 | 1.0909 | 1.38E-01 | 2.26E-01 |
| XL_VLDL_C_pct | -4.7431 | 3.10E-05 | -4.2180 | 1.07E-03 | 0.0236 | 0.0718 | 0.3292 | 3.71E-01 | 4.99E-01 |
| XL_VLDL_FC_pct | -3.4972 | 1.58E-03 | -2.6617 | 2.33E-02 | 0.0284 | 0.0604 | 0.4698 | 3.19E-01 | 4.48E-01 |
| XL_VLDL_TG_pct | 3.9728 | 2.90E-04 | 3.3047 | 4.64E-03 | 0.0064 | 0.0057 | 1.1258 | 8.70E-01 | 9.08E-01 |
| XS_VLDL_CE_pct | -4.4158 | 6.11E-05 | -3.2712 | 5.03E-03 | -0.0391 | 0.0562 | -0.6951 | 2.43E-01 | 3.61E-01 |
| XS_VLDL_C_pct | -4.4803 | 5.40E-05 | -3.3134 | 4.59E-03 | -0.0422 | 0.0651 | -0.6475 | 2.59E-01 | 3.81E-01 |

**Supplementary Table 5**

**Results of replication in PReDICT study**

| | | HRSD-17 | | | HRSA | | |
|---|---|---|---|---|---|---|---|
| **Metabolite** | **N** | **Beta** | **Std. Error** | **Pr(>\|t\|)** | **Beta** | **Std. Error** | **Pr(>\|t\|)** |
| Pyruvate | 163 | 0.00006 | 0.00002 | 2.56E-02 | 0.00013 | 0.00004 | 7.18E-04 |
| Citric acid | 208 | -0.00103 | 0.0005 | 4.18E-02 | -0.00258 | 0.00077 | 9.19E-04 |
| L_Alanine | 208 | 0 | 0.00018 | 9.88E-01 | 0.00018 | 0.00028 | 5.17E-01 |
| C18:2(cis_9,12) | 208 | -0.0003 | 0.00017 | 7.35E-02 | -0.00054 | 0.00026 | 3.84E-02 |
| C18:3(cis_6,9,12) | 208 | -0.01687 | 0.00963 | 8.13E-02 | -0.02486 | 0.01494 | 9.78E-02 |
| C18:3(cis_9,12,15) | 208 | -0.00308 | 0.00163 | 6.05E-02 | -0.00539 | 0.00252 | 3.38E-02 |
| C19:2(cis_10,13) | 208 | -0.02472 | 0.01302 | 5.89E-02 | -0.05465 | 0.02 | 6.84E-03 |
| C20:2(cis_11,14) | 208 | -0.00598 | 0.0036 | 9.82E-02 | -0.01091 | 0.00557 | 5.14E-02 |
| C20:3(cis_8,11,14) | 208 | -0.00667 | 0.00649 | 3.05E-01 | -0.00913 | 0.01006 | 3.65E-01 |
| C20:4(cis_5,8,11,14) | 208 | -0.00613 | 0.00302 | 4.39E-02 | -0.01575 | 0.0046 | 7.57E-04 |
| C20:5(cis_5,8,11,14,17) | 208 | -0.02507 | 0.0143 | 8.11E-02 | -0.06345 | 0.02189 | 4.17E-03 |
| C22:4(cis_7,10,13,16) | 208 | -0.01312 | 0.00658 | 4.75E-02 | -0.02352 | 0.01017 | 2.17E-02 |
| C22:5(cis_4,7,10,13,16) | 208 | -0.0188 | 0.01661 | 2.59E-01 | -0.02272 | 0.02579 | 3.79E-01 |
| C22:5(cis_7,10,13,16,19) | 208 | -0.0158 | 0.00764 | 3.98E-02 | -0.03437 | 0.01172 | 3.75E-03 |
| C22:6(cis_4,7,10,13,16,19) | 208 | -0.00258 | 0.00398 | 5.18E-01 | -0.00839 | 0.00615 | 1.74E-01 |

S

**Supplementary Table 6**

**Results of Mendelian Randomization analysis**

| method | nsnp | b | se | pval | analysis | exposure | outcome |
|---|---|---|---|---|---|---|---|
| MR Egger | 57 | 0.0667223 | 0.067091 | 0.3243329 | TwoSample_MR | Ala | MDD |
| Weighted median | 57 | 0.0378879 | 0.0301999 | 0.209635 | TwoSample_MR | Ala | MDD |
| Inverse variance weighted | 57 | 0.0092916 | 0.0281596 | 0.741428 | TwoSample_MR | Ala | MDD |
| Simple mode | 57 | 0.0120272 | 0.0660859 | 0.8562451 | TwoSample_MR | Ala | MDD |
| Weighted mode | 57 | 0.0335892 | 0.053407 | 0.5319529 | TwoSample_MR | Ala | MDD |
| MR Egger | 92 | 0.0077517 | 0.0284373 | 0.7857941 | TwoSample_MR | ApoA1 | MDD |
| Weighted median | 92 | 0.0058253 | 0.0184715 | 0.7524833 | TwoSample_MR | ApoA1 | MDD |
| Inverse variance weighted | 92 | 0.0224671 | 0.0167259 | 0.1791917 | TwoSample_MR | ApoA1 | MDD |
| Simple mode | 92 | -0.0048043 | 0.0333036 | 0.8856166 | TwoSample_MR | ApoA1 | MDD |
| Weighted mode | 92 | 0.0077864 | 0.0190952 | 0.6844018 | TwoSample_MR | ApoA1 | MDD |
| MR Egger | 48 | -0.0189866 | 0.0325635 | 0.5626993 | TwoSample_MR | Citrate | MDD |
| Weighted median | 48 | 0.0137925 | 0.0269352 | 0.6086086 | TwoSample_MR | Citrate | MDD |
| Inverse variance weighted | 48 | 0.0068533 | 0.0191829 | 0.7208962 | TwoSample_MR | Citrate | MDD |
| Simple mode | 48 | 0.0519499 | 0.0535942 | 0.3373479 | TwoSample_MR | Citrate | MDD |
| Weighted mode | 48 | 0.0037736 | 0.0280376 | 0.8935094 | TwoSample_MR | Citrate | MDD |
| MR Egger | 128 | -0.0200796 | 0.0214208 | 0.3503525 | TwoSample_MR | HDL_CE | MDD |
| Weighted median | 128 | -0.0121994 | 0.0173502 | 0.4819766 | TwoSample_MR | HDL_CE | MDD |
| Inverse variance weighted | 128 | 0.0010975 | 0.0132909 | 0.9341871 | TwoSample_MR | HDL_CE | MDD |
| Simple mode | 128 | -0.0475196 | 0.0335391 | 0.1589778 | TwoSample_MR | HDL_CE | MDD |
| Weighted mode | 128 | -0.0108873 | 0.0186774 | 0.560985 | TwoSample_MR | HDL_CE | MDD |
| MR Egger | 122 | -0.0205654 | 0.0215699 | 0.3422883 | TwoSample_MR | HDL_C | MDD |
| Weighted median | 122 | -0.0038288 | 0.017248 | 0.824325 | TwoSample_MR | HDL_C | MDD |
| Inverse variance weighted | 122 | -0.0017461 | 0.013293 | 0.8954955 | TwoSample_MR | HDL_C | MDD |
| Simple mode | 122 | -0.0471621 | 0.0327384 | 0.1522883 | TwoSample_MR | HDL_C | MDD |
| Weighted mode | 122 | -0.00338 | 0.017881 | 0.850386 | TwoSample_MR | HDL_C | MDD |
| MR Egger | 111 | -0.0069945 | 0.0226385 | 0.7579393 | TwoSample_MR | HDL_FC | MDD |
| Weighted median | 111 | -0.0116492 | 0.018655 | 0.5323295 | TwoSample_MR | HDL_FC | MDD |
| Inverse variance weighted | 111 | -0.0094322 | 0.0142742 | 0.5087501 | TwoSample_MR | HDL_FC | MDD |
| Simple mode | 111 | -0.0289881 | 0.0362238 | 0.425291 | TwoSample_MR | HDL_FC | MDD |
| Weighted mode | 111 | -0.0026424 | 0.0170151 | 0.8768724 | TwoSample_MR | HDL_FC | MDD |
| MR Egger | 104 | 0.0041479 | 0.0261135 | 0.8741081 | TwoSample_MR | HDL_L | MDD |
| Weighted median | 104 | 0.0025879 | 0.0183389 | 0.8877784 | TwoSample_MR | HDL_L | MDD |
| Inverse variance weighted | 104 | 0.0105573 | 0.0157385 | 0.502349 | TwoSample_MR | HDL_L | MDD |
| Simple mode | 104 | -0.0072872 | 0.0351068 | 0.8359731 | TwoSample_MR | HDL_L | MDD |
| Weighted mode | 104 | 0.0059389 | 0.0177404 | 0.7384809 | TwoSample_MR | HDL_L | MDD |
| MR Egger | 95 | -0.0083284 | 0.0279203 | 0.7661444 | TwoSample_MR | HDL_PL | MDD |
| Weighted median | 95 | 0.0019159 | 0.0178419 | 0.9144847 | TwoSample_MR | HDL_PL | MDD |
| Inverse variance weighted | 95 | 0.0164131 | 0.0168569 | 0.3302201 | TwoSample_MR | HDL_PL | MDD |
| Simple mode | 95 | -0.009229 | 0.0304663 | 0.7626173 | TwoSample_MR | HDL_PL | MDD |
| Weighted mode | 95 | 0.0036322 | 0.0173544 | 0.8346689 | TwoSample_MR | HDL_PL | MDD |
| MR Egger | 89 | 0.0249119 | 0.0313452 | 0.4289167 | TwoSample_MR | HDL_P | MDD |
| Weighted median | 89 | 0.014967 | 0.0224091 | 0.5041989 | TwoSample_MR | HDL_P | MDD |
| Inverse variance weighted | 89 | 0.0143508 | 0.0177373 | 0.4184715 | TwoSample_MR | HDL_P | MDD |

| | | | | | | | |
|---|---|---|---|---|---|---|---|
| Simple mode | 89 | -0.0071042 | 0.036939 | 0.8479323 | TwoSample_MR | HDL_P | MDD |
| Weighted mode | 89 | 0.0053626 | 0.0218926 | 0.8070671 | TwoSample_MR | HDL_P | MDD |
| MR Egger | 93 | 0.0005753 | 0.0177854 | 0.974265 | TwoSample_MR | HDL_TG | MDD |
| Weighted median | 93 | 0.0054625 | 0.014983 | 0.7154268 | TwoSample_MR | HDL_TG | MDD |
| Inverse variance weighted | 93 | 0.0167021 | 0.0122349 | 0.1722153 | TwoSample_MR | HDL_TG | MDD |
| Simple mode | 93 | -0.0033009 | 0.0263093 | 0.9004283 | TwoSample_MR | HDL_TG | MDD |
| Weighted mode | 93 | 0.0036933 | 0.0125393 | 0.7690122 | TwoSample_MR | HDL_TG | MDD |
| MR Egger | 127 | -0.0081725 | 0.0198247 | 0.68087 | TwoSample_MR | HDL_size | MDD |
| Weighted median | 127 | -0.0057269 | 0.0152923 | 0.7080376 | TwoSample_MR | HDL_size | MDD |
| Inverse variance weighted | 127 | -0.0091093 | 0.0129453 | 0.4816325 | TwoSample_MR | HDL_size | MDD |
| Simple mode | 127 | 0.0054609 | 0.033346 | 0.8701796 | TwoSample_MR | HDL_size | MDD |
| Weighted mode | 127 | -0.0049641 | 0.0132699 | 0.7089714 | TwoSample_MR | HDL_size | MDD |
| MR Egger | 78 | -0.0289804 | 0.0182128 | 0.1157133 | TwoSample_MR | IDL_CE | MDD |
| Weighted median | 78 | -0.0176928 | 0.016629 | 0.287341 | TwoSample_MR | IDL_CE | MDD |
| Inverse variance weighted | 78 | -0.0278218 | 0.0129782 | 0.0320539 | TwoSample_MR | IDL_CE | MDD |
| Simple mode | 78 | -0.027737 | 0.0376587 | 0.4636421 | TwoSample_MR | IDL_CE | MDD |
| Weighted mode | 78 | -0.0163256 | 0.0154278 | 0.2932754 | TwoSample_MR | IDL_CE | MDD |
| MR Egger | 94 | -0.008195 | 0.0164745 | 0.6200682 | TwoSample_MR | IDL_CE_pct | MDD |
| Weighted median | 94 | -0.0080828 | 0.0150541 | 0.5913266 | TwoSample_MR | IDL_CE_pct | MDD |
| Inverse variance weighted | 94 | -0.0114469 | 0.0118357 | 0.3334684 | TwoSample_MR | IDL_CE_pct | MDD |
| Simple mode | 94 | -0.0051527 | 0.0268457 | 0.8482091 | TwoSample_MR | IDL_CE_pct | MDD |
| Weighted mode | 94 | -0.0016703 | 0.0128886 | 0.8971654 | TwoSample_MR | IDL_CE_pct | MDD |
| MR Egger | 76 | -0.0297718 | 0.0181576 | 0.1053275 | TwoSample_MR | IDL_C | MDD |
| Weighted median | 76 | -0.0186075 | 0.0170412 | 0.2748716 | TwoSample_MR | IDL_C | MDD |
| Inverse variance weighted | 76 | -0.0302202 | 0.012855 | 0.0187306 | TwoSample_MR | IDL_C | MDD |
| Simple mode | 76 | -0.033019 | 0.0348359 | 0.346254 | TwoSample_MR | IDL_C | MDD |
| Weighted mode | 76 | -0.01858 | 0.0152302 | 0.2263069 | TwoSample_MR | IDL_C | MDD |
| MR Egger | 104 | -0.0169743 | 0.016344 | 0.3014636 | TwoSample_MR | IDL_C_pct | MDD |
| Weighted median | 104 | -0.0095834 | 0.0146295 | 0.5124206 | TwoSample_MR | IDL_C_pct | MDD |
| Inverse variance weighted | 104 | -0.0188773 | 0.011407 | 0.0979483 | TwoSample_MR | IDL_C_pct | MDD |
| Simple mode | 104 | -0.0125942 | 0.025389 | 0.6209154 | TwoSample_MR | IDL_C_pct | MDD |
| Weighted mode | 104 | -0.0090207 | 0.0126795 | 0.4784209 | TwoSample_MR | IDL_C_pct | MDD |
| MR Egger | 74 | -0.0370013 | 0.0193086 | 0.0592958 | TwoSample_MR | IDL_FC | MDD |
| Weighted median | 74 | -0.021598 | 0.0170439 | 0.2050846 | TwoSample_MR | IDL_FC | MDD |
| Inverse variance weighted | 74 | -0.0295749 | 0.01347 | 0.0281196 | TwoSample_MR | IDL_FC | MDD |
| Simple mode | 74 | -0.0248108 | 0.0364707 | 0.4984707 | TwoSample_MR | IDL_FC | MDD |
| Weighted mode | 74 | -0.019615 | 0.0162453 | 0.2311654 | TwoSample_MR | IDL_FC | MDD |
| MR Egger | 69 | -0.0362612 | 0.0256144 | 0.1615078 | TwoSample_MR | IDL_FC_pct | MDD |
| Weighted median | 69 | -0.0304001 | 0.0225917 | 0.1784218 | TwoSample_MR | IDL_FC_pct | MDD |
| Inverse variance weighted | 69 | -0.0515628 | 0.0155247 | 0.0008959 | TwoSample_MR | IDL_FC_pct | MDD |
| Simple mode | 69 | -0.0302166 | 0.0401056 | 0.4537951 | TwoSample_MR | IDL_FC_pct | MDD |
| Weighted mode | 69 | -0.033304 | 0.0254094 | 0.1943687 | TwoSample_MR | IDL_FC_pct | MDD |
| MR Egger | 75 | -0.0226958 | 0.0193935 | 0.2456964 | TwoSample_MR | IDL_L | MDD |
| Weighted median | 75 | -0.0193819 | 0.0167591 | 0.2474774 | TwoSample_MR | IDL_L | MDD |
| Inverse variance weighted | 75 | -0.026011 | 0.0136397 | 0.0565208 | TwoSample_MR | IDL_L | MDD |
| Simple mode | 75 | -0.0263161 | 0.0358091 | 0.4647224 | TwoSample_MR | IDL_L | MDD |

| | | | | | | | |
|---|---|---|---|---|---|---|---|
| Weighted mode | 75 | -0.0180957 | 0.0159971 | 0.2616283 | TwoSample_MR | IDL_L | MDD |
| MR Egger | 97 | -0.0021349 | 0.020635 | 0.9178162 | TwoSample_MR | IDL_TG | MDD |
| Weighted median | 97 | 0.02102 | 0.0162981 | 0.1971479 | TwoSample_MR | IDL_TG | MDD |
| Inverse variance weighted | 97 | 0.0070399 | 0.0128798 | 0.5846654 | TwoSample_MR | IDL_TG | MDD |
| Simple mode | 97 | -0.0024336 | 0.0297932 | 0.9350684 | TwoSample_MR | IDL_TG | MDD |
| Weighted mode | 97 | 0.0144356 | 0.0139887 | 0.3046883 | TwoSample_MR | IDL_TG | MDD |
| MR Egger | 108 | 0.0224738 | 0.0195563 | 0.2530663 | TwoSample_MR | IDL_TG_pct | MDD |
| Weighted median | 108 | 0.0221403 | 0.015063 | 0.1416016 | TwoSample_MR | IDL_TG_pct | MDD |
| Inverse variance weighted | 108 | 0.0367349 | 0.0133434 | 0.0059044 | TwoSample_MR | IDL_TG_pct | MDD |
| Simple mode | 108 | 0.0217959 | 0.029374 | 0.4597054 | TwoSample_MR | IDL_TG_pct | MDD |
| Weighted mode | 108 | 0.0181482 | 0.0135078 | 0.1819407 | TwoSample_MR | IDL_TG_pct | MDD |
| MR Egger | 67 | 0.0144044 | 0.0317621 | 0.6516932 | TwoSample_MR | LA_pct | MDD |
| Weighted median | 67 | 0.0493091 | 0.0244039 | 0.0433271 | TwoSample_MR | LA_pct | MDD |
| Inverse variance weighted | 67 | 0.0145732 | 0.0197436 | 0.4604415 | TwoSample_MR | LA_pct | MDD |
| Simple mode | 67 | 0.0027054 | 0.0551532 | 0.961025 | TwoSample_MR | LA_pct | MDD |
| Weighted mode | 67 | 0.0443028 | 0.0223115 | 0.0512299 | TwoSample_MR | LA_pct | MDD |
| MR Egger | 147 | -0.0133302 | 0.020707 | 0.5207523 | TwoSample_MR | L_HDL_CE | MDD |
| Weighted median | 147 | -0.014472 | 0.0178187 | 0.416686 | TwoSample_MR | L_HDL_CE | MDD |
| Inverse variance weighted | 147 | -0.0123161 | 0.0131632 | 0.3494526 | TwoSample_MR | L_HDL_CE | MDD |
| Simple mode | 147 | -0.0147702 | 0.0321983 | 0.6471135 | TwoSample_MR | L_HDL_CE | MDD |
| Weighted mode | 147 | -0.0107834 | 0.0137928 | 0.4355901 | TwoSample_MR | L_HDL_CE | MDD |
| MR Egger | 110 | -0.0195947 | 0.0226101 | 0.3880643 | TwoSample_MR | L_HDL_CE_pct | MDD |
| Weighted median | 110 | -0.0306698 | 0.0169305 | 0.0700611 | TwoSample_MR | L_HDL_CE_pct | MDD |
| Inverse variance weighted | 110 | -0.0275854 | 0.0141483 | 0.0512077 | TwoSample_MR | L_HDL_CE_pct | MDD |
| Simple mode | 110 | -0.046432 | 0.0318133 | 0.1472984 | TwoSample_MR | L_HDL_CE_pct | MDD |
| Weighted mode | 110 | -0.0245063 | 0.0163202 | 0.1360931 | TwoSample_MR | L_HDL_CE_pct | MDD |
| MR Egger | 147 | -0.0179553 | 0.0202215 | 0.3760485 | TwoSample_MR | L_HDL_C | MDD |
| Weighted median | 147 | -0.0141883 | 0.016723 | 0.3961987 | TwoSample_MR | L_HDL_C | MDD |
| Inverse variance weighted | 147 | -0.0118204 | 0.0129199 | 0.3602422 | TwoSample_MR | L_HDL_C | MDD |
| Simple mode | 147 | -0.033297 | 0.0323108 | 0.3044685 | TwoSample_MR | L_HDL_C | MDD |
| Weighted mode | 147 | -0.0147807 | 0.0138187 | 0.2865583 | TwoSample_MR | L_HDL_C | MDD |
| MR Egger | 121 | -0.0181402 | 0.0188918 | 0.338894 | TwoSample_MR | L_HDL_C_pct | MDD |
| Weighted median | 121 | -0.0319756 | 0.0171231 | 0.0618459 | TwoSample_MR | L_HDL_C_pct | MDD |
| Inverse variance weighted | 121 | -0.024741 | 0.0121776 | 0.0421867 | TwoSample_MR | L_HDL_C_pct | MDD |
| Simple mode | 121 | -0.037644 | 0.0336221 | 0.2651124 | TwoSample_MR | L_HDL_C_pct | MDD |
| Weighted mode | 121 | -0.0237925 | 0.0157836 | 0.1343301 | TwoSample_MR | L_HDL_C_pct | MDD |
| MR Egger | 138 | -0.0216217 | 0.0193717 | 0.2663274 | TwoSample_MR | L_HDL_FC | MDD |
| Weighted median | 138 | -0.0234829 | 0.0159361 | 0.1405985 | TwoSample_MR | L_HDL_FC | MDD |
| Inverse variance weighted | 138 | -0.0114222 | 0.0127508 | 0.3703564 | TwoSample_MR | L_HDL_FC | MDD |
| Simple mode | 138 | -0.0216931 | 0.0305429 | 0.4787576 | TwoSample_MR | L_HDL_FC | MDD |
| Weighted mode | 138 | -0.0140939 | 0.0144337 | 0.3305588 | TwoSample_MR | L_HDL_FC | MDD |
| MR Egger | 93 | -0.0208117 | 0.0189768 | 0.2756686 | TwoSample_MR | L_HDL_FC_pct | MDD |
| Weighted median | 93 | -0.0138572 | 0.0144451 | 0.3374071 | TwoSample_MR | L_HDL_FC_pct | MDD |
| Inverse variance weighted | 93 | -0.0394307 | 0.0134497 | 0.0033709 | TwoSample_MR | L_HDL_FC_pct | MDD |
| Simple mode | 93 | -0.024212 | 0.0297823 | 0.4183366 | TwoSample_MR | L_HDL_FC_pct | MDD |
| Weighted mode | 93 | -0.0162187 | 0.0133235 | 0.2266013 | TwoSample_MR | L_HDL_FC_pct | MDD |

| | | | | | | | |
|---|---|---|---|---|---|---|---|
| MR Egger | 143 | -0.0152596 | 0.0196828 | 0.4394732 | TwoSample_MR | L_HDL_L | MDD |
| Weighted median | 143 | -0.0150206 | 0.0157814 | 0.3412037 | TwoSample_MR | L_HDL_L | MDD |
| Inverse variance weighted | 143 | -0.0107968 | 0.0128592 | 0.401122 | TwoSample_MR | L_HDL_L | MDD |
| Simple mode | 143 | -0.0246487 | 0.0327604 | 0.4530605 | TwoSample_MR | L_HDL_L | MDD |
| Weighted mode | 143 | -0.0136424 | 0.0139611 | 0.3301453 | TwoSample_MR | L_HDL_L | MDD |
| MR Egger | 132 | -0.0132372 | 0.0195424 | 0.4993831 | TwoSample_MR | L_HDL_PL | MDD |
| Weighted median | 132 | -0.01298 | 0.0167522 | 0.438442 | TwoSample_MR | L_HDL_PL | MDD |
| Inverse variance weighted | 132 | -0.0103831 | 0.0127513 | 0.4154848 | TwoSample_MR | L_HDL_PL | MDD |
| Simple mode | 132 | -0.0368031 | 0.0310574 | 0.2381618 | TwoSample_MR | L_HDL_PL | MDD |
| Weighted mode | 132 | -0.0156784 | 0.014296 | 0.2747831 | TwoSample_MR | L_HDL_PL | MDD |
| MR Egger | 113 | 0.0319482 | 0.0274729 | 0.2473639 | TwoSample_MR | L_HDL_PL_pct | MDD |
| Weighted median | 113 | 0.034956 | 0.0182565 | 0.0555285 | TwoSample_MR | L_HDL_PL_pct | MDD |
| Inverse variance weighted | 113 | 0.0388777 | 0.0156737 | 0.013122 | TwoSample_MR | L_HDL_PL_pct | MDD |
| Simple mode | 113 | 0.0294069 | 0.0343561 | 0.3938563 | TwoSample_MR | L_HDL_PL_pct | MDD |
| Weighted mode | 113 | 0.0258127 | 0.0181764 | 0.15835 | TwoSample_MR | L_HDL_PL_pct | MDD |
| MR Egger | 145 | -0.0175477 | 0.0193964 | 0.3671528 | TwoSample_MR | L_HDL_P | MDD |
| Weighted median | 145 | -0.0145798 | 0.0168053 | 0.3856307 | TwoSample_MR | L_HDL_P | MDD |
| Inverse variance weighted | 145 | -0.0084792 | 0.0126673 | 0.5032579 | TwoSample_MR | L_HDL_P | MDD |
| Simple mode | 145 | -0.0263388 | 0.0329921 | 0.4259887 | TwoSample_MR | L_HDL_P | MDD |
| Weighted mode | 145 | -0.0113731 | 0.0139775 | 0.4171766 | TwoSample_MR | L_HDL_P | MDD |
| MR Egger | 115 | 0.0158246 | 0.0184772 | 0.3935669 | TwoSample_MR | L_HDL_TG_pct | MDD |
| Weighted median | 115 | 0.0247651 | 0.0156719 | 0.1140574 | TwoSample_MR | L_HDL_TG_pct | MDD |
| Inverse variance weighted | 115 | 0.0155953 | 0.0122304 | 0.2022637 | TwoSample_MR | L_HDL_TG_pct | MDD |
| Simple mode | 115 | 0.0111314 | 0.0279463 | 0.6911439 | TwoSample_MR | L_HDL_TG_pct | MDD |
| Weighted mode | 115 | 0.019673 | 0.0145946 | 0.18034 | TwoSample_MR | L_HDL_TG_pct | MDD |
| MR Egger | 96 | 0.0137518 | 0.0160807 | 0.3946299 | TwoSample_MR | L_LDL_TG_pct | MDD |
| Weighted median | 96 | 0.0076081 | 0.0156109 | 0.6260041 | TwoSample_MR | L_LDL_TG_pct | MDD |
| Inverse variance weighted | 96 | 0.0213182 | 0.0113317 | 0.0599313 | TwoSample_MR | L_LDL_TG_pct | MDD |
| Simple mode | 96 | 0.0189319 | 0.029183 | 0.5180791 | TwoSample_MR | L_LDL_TG_pct | MDD |
| Weighted mode | 96 | 0.0035656 | 0.0134379 | 0.7913239 | TwoSample_MR | L_LDL_TG_pct | MDD |
| MR Egger | 90 | 0.0204596 | 0.0225623 | 0.3669856 | TwoSample_MR | L_VLDL_CE | MDD |
| Weighted median | 90 | 0.0133007 | 0.0179707 | 0.4592204 | TwoSample_MR | L_VLDL_CE | MDD |
| Inverse variance weighted | 90 | -0.004315 | 0.0135046 | 0.7493296 | TwoSample_MR | L_VLDL_CE | MDD |
| Simple mode | 90 | 0.015517 | 0.0335452 | 0.6448021 | TwoSample_MR | L_VLDL_CE | MDD |
| Weighted mode | 90 | 0.0126011 | 0.0184024 | 0.4952776 | TwoSample_MR | L_VLDL_CE | MDD |
| MR Egger | 92 | -0.0278373 | 0.0211825 | 0.1921297 | TwoSample_MR | L_VLDL_CE_pct | MDD |
| Weighted median | 92 | -0.0258279 | 0.0167398 | 0.1228542 | TwoSample_MR | L_VLDL_CE_pct | MDD |
| Inverse variance weighted | 92 | -0.0501386 | 0.0135641 | 0.0002187 | TwoSample_MR | L_VLDL_CE_pct | MDD |
| Simple mode | 92 | -0.0042013 | 0.0290767 | 0.885434 | TwoSample_MR | L_VLDL_CE_pct | MDD |
| Weighted mode | 92 | -0.0220466 | 0.0153101 | 0.1532982 | TwoSample_MR | L_VLDL_CE_pct | MDD |
| MR Egger | 90 | 0.0233637 | 0.0233943 | 0.3206823 | TwoSample_MR | L_VLDL_C | MDD |
| Weighted median | 90 | 0.0231563 | 0.0181669 | 0.2024348 | TwoSample_MR | L_VLDL_C | MDD |
| Inverse variance weighted | 90 | 0.0039428 | 0.014225 | 0.7816457 | TwoSample_MR | L_VLDL_C | MDD |
| Simple mode | 90 | 0.007521 | 0.0315463 | 0.8121112 | TwoSample_MR | L_VLDL_C | MDD |
| Weighted mode | 90 | 0.0140711 | 0.017427 | 0.4215727 | TwoSample_MR | L_VLDL_C | MDD |
| MR Egger | 95 | 0.0203543 | 0.0223967 | 0.3657997 | TwoSample_MR | L_VLDL_FC | MDD |

| | | | | | | | |
|---|---|---|---|---|---|---|---|
| Weighted median | 95 | 0.0231628 | 0.0183145 | 0.2059707 | TwoSample_MR | L_VLDL_FC | MDD |
| Inverse variance weighted | 95 | 0.0077144 | 0.0139871 | 0.5812676 | TwoSample_MR | L_VLDL_FC | MDD |
| Simple mode | 95 | 0.0152591 | 0.0301609 | 0.6140953 | TwoSample_MR | L_VLDL_FC | MDD |
| Weighted mode | 95 | 0.0183598 | 0.0174559 | 0.2955958 | TwoSample_MR | L_VLDL_FC | MDD |
| MR Egger | 92 | 0.020775 | 0.021193 | 0.329576 | TwoSample_MR | L_VLDL_L | MDD |
| Weighted median | 92 | 0.0231273 | 0.0181845 | 0.2034397 | TwoSample_MR | L_VLDL_L | MDD |
| Inverse variance weighted | 92 | 0.0100136 | 0.0132463 | 0.449677 | TwoSample_MR | L_VLDL_L | MDD |
| Simple mode | 92 | 0.0043723 | 0.031619 | 0.8903242 | TwoSample_MR | L_VLDL_L | MDD |
| Weighted mode | 92 | 0.019737 | 0.0181801 | 0.2805085 | TwoSample_MR | L_VLDL_L | MDD |
| MR Egger | 98 | 0.0159627 | 0.0221016 | 0.4719005 | TwoSample_MR | L_VLDL_PL | MDD |
| Weighted median | 98 | 0.0239594 | 0.0178235 | 0.1788652 | TwoSample_MR | L_VLDL_PL | MDD |
| Inverse variance weighted | 98 | 0.0107863 | 0.0137873 | 0.4340182 | TwoSample_MR | L_VLDL_PL | MDD |
| Simple mode | 98 | 0.0113303 | 0.0305256 | 0.711318 | TwoSample_MR | L_VLDL_PL | MDD |
| Weighted mode | 98 | 0.0170118 | 0.0169154 | 0.3170601 | TwoSample_MR | L_VLDL_PL | MDD |
| MR Egger | 107 | 0.0159769 | 0.0200418 | 0.4271454 | TwoSample_MR | L_VLDL_PL_pct | MDD |
| Weighted median | 107 | -0.0109689 | 0.0166042 | 0.5088614 | TwoSample_MR | L_VLDL_PL_pct | MDD |
| Inverse variance weighted | 107 | 0.0101405 | 0.0124924 | 0.4169469 | TwoSample_MR | L_VLDL_PL_pct | MDD |
| Simple mode | 107 | 0.0180708 | 0.031828 | 0.5713958 | TwoSample_MR | L_VLDL_PL_pct | MDD |
| Weighted mode | 107 | 0.0042174 | 0.0160973 | 0.7938335 | TwoSample_MR | L_VLDL_PL_pct | MDD |
| MR Egger | 97 | 0.0140828 | 0.0221307 | 0.5260811 | TwoSample_MR | L_VLDL_P | MDD |
| Weighted median | 97 | 0.0232153 | 0.0182028 | 0.2021799 | TwoSample_MR | L_VLDL_P | MDD |
| Inverse variance weighted | 97 | 0.0110675 | 0.0139069 | 0.4261335 | TwoSample_MR | L_VLDL_P | MDD |
| Simple mode | 97 | 0.0055316 | 0.0284187 | 0.8460801 | TwoSample_MR | L_VLDL_P | MDD |
| Weighted mode | 97 | 0.0168303 | 0.0169652 | 0.323667 | TwoSample_MR | L_VLDL_P | MDD |
| MR Egger | 100 | 0.0237174 | 0.0203006 | 0.2455149 | TwoSample_MR | L_VLDL_TG | MDD |
| Weighted median | 100 | 0.0267263 | 0.0179061 | 0.1355464 | TwoSample_MR | L_VLDL_TG | MDD |
| Inverse variance weighted | 100 | 0.0186259 | 0.0129583 | 0.1506108 | TwoSample_MR | L_VLDL_TG | MDD |
| Simple mode | 100 | 0.0093169 | 0.0303414 | 0.7594355 | TwoSample_MR | L_VLDL_TG | MDD |
| Weighted mode | 100 | 0.0254808 | 0.0167269 | 0.1308604 | TwoSample_MR | L_VLDL_TG | MDD |
| MR Egger | 108 | 0.0429152 | 0.0849723 | 0.6145725 | TwoSample_MR | MDD | Ala |
| Weighted median | 108 | -0.0052613 | 0.0233899 | 0.8220255 | TwoSample_MR | MDD | Ala |
| Inverse variance weighted | 108 | 0.0000488 | 0.0186586 | 0.9979142 | TwoSample_MR | MDD | Ala |
| Simple mode | 108 | 0.0341722 | 0.0625182 | 0.5857943 | TwoSample_MR | MDD | Ala |
| Weighted mode | 108 | 0.0278651 | 0.0589238 | 0.6372468 | TwoSample_MR | MDD | Ala |
| MR Egger | 108 | 0.1716817 | 0.0918296 | 0.0643037 | TwoSample_MR | MDD | ApoA1 |
| Weighted median | 108 | 0.0015291 | 0.0238761 | 0.9489366 | TwoSample_MR | MDD | ApoA1 |
| Inverse variance weighted | 108 | -0.009673 | 0.0205248 | 0.6374378 | TwoSample_MR | MDD | ApoA1 |
| Simple mode | 108 | 0.0778783 | 0.0757384 | 0.3061495 | TwoSample_MR | MDD | ApoA1 |
| Weighted mode | 108 | 0.0808434 | 0.0730576 | 0.2709609 | TwoSample_MR | MDD | ApoA1 |
| MR Egger | 108 | 0.0230932 | 0.0805891 | 0.7750122 | TwoSample_MR | MDD | Citrate |
| Weighted median | 108 | -0.0228207 | 0.0240303 | 0.342283 | TwoSample_MR | MDD | Citrate |
| Inverse variance weighted | 108 | -0.0239407 | 0.0177045 | 0.1762994 | TwoSample_MR | MDD | Citrate |
| Simple mode | 108 | -0.0577916 | 0.0681501 | 0.3983287 | TwoSample_MR | MDD | Citrate |
| Weighted mode | 108 | -0.040612 | 0.063424 | 0.5233298 | TwoSample_MR | MDD | Citrate |
| MR Egger | 108 | 0.1660635 | 0.0934113 | 0.0783097 | TwoSample_MR | MDD | HDL_C |
| Weighted median | 108 | -0.047646 | 0.0236424 | 0.0438758 | TwoSample_MR | MDD | HDL_C |

| | | | | | | | |
|---|---|---|---|---|---|---|---|
| Inverse variance weighted | 108 | -0.0482568 | 0.0210142 | 0.0216537 | TwoSample_MR | MDD | HDL_C |
| Simple mode | 108 | -0.0547498 | 0.0763523 | 0.4748943 | TwoSample_MR | MDD | HDL_C |
| Weighted mode | 108 | -0.0488457 | 0.0675989 | 0.4715139 | TwoSample_MR | MDD | HDL_C |
| MR Egger | 108 | 0.1711233 | 0.0932204 | 0.0692074 | TwoSample_MR | MDD | HDL_CE |
| Weighted median | 108 | -0.0524676 | 0.0230607 | 0.0228943 | TwoSample_MR | MDD | HDL_CE |
| Inverse variance weighted | 108 | -0.0497039 | 0.0210058 | 0.0179717 | TwoSample_MR | MDD | HDL_CE |
| Simple mode | 108 | -0.0605757 | 0.0713844 | 0.398007 | TwoSample_MR | MDD | HDL_CE |
| Weighted mode | 108 | -0.0636092 | 0.0731175 | 0.3862704 | TwoSample_MR | MDD | HDL_CE |
| MR Egger | 108 | 0.144876 | 0.0928055 | 0.1214877 | TwoSample_MR | MDD | HDL_FC |
| Weighted median | 108 | -0.0275149 | 0.0227359 | 0.2262036 | TwoSample_MR | MDD | HDL_FC |
| Inverse variance weighted | 108 | -0.042645 | 0.0207606 | 0.0399634 | TwoSample_MR | MDD | HDL_FC |
| Simple mode | 108 | 0.0022606 | 0.0733741 | 0.9754788 | TwoSample_MR | MDD | HDL_FC |
| Weighted mode | 108 | 0.0135555 | 0.0681541 | 0.8427226 | TwoSample_MR | MDD | HDL_FC |
| MR Egger | 108 | 0.1764009 | 0.0913569 | 0.0561667 | TwoSample_MR | MDD | HDL_L |
| Weighted median | 108 | -0.0074167 | 0.0236544 | 0.7538668 | TwoSample_MR | MDD | HDL_L |
| Inverse variance weighted | 108 | -0.0241873 | 0.0205085 | 0.2382479 | TwoSample_MR | MDD | HDL_L |
| Simple mode | 108 | 0.0294969 | 0.0758773 | 0.6982377 | TwoSample_MR | MDD | HDL_L |
| Weighted mode | 108 | 0.0324806 | 0.0728643 | 0.6566656 | TwoSample_MR | MDD | HDL_L |
| MR Egger | 108 | 0.1466165 | 0.0938509 | 0.1212149 | TwoSample_MR | MDD | HDL_P |
| Weighted median | 108 | -0.0019065 | 0.0247073 | 0.9384925 | TwoSample_MR | MDD | HDL_P |
| Inverse variance weighted | 108 | 0.0002559 | 0.0208295 | 0.9901987 | TwoSample_MR | MDD | HDL_P |
| Simple mode | 108 | 0.1356337 | 0.0838858 | 0.1088477 | TwoSample_MR | MDD | HDL_P |
| Weighted mode | 108 | 0.1326871 | 0.0900653 | 0.1436242 | TwoSample_MR | MDD | HDL_P |
| MR Egger | 108 | 0.1800691 | 0.0903589 | 0.0488513 | TwoSample_MR | MDD | HDL_PL |
| Weighted median | 108 | -0.0026711 | 0.0232668 | 0.9086023 | TwoSample_MR | MDD | HDL_PL |
| Inverse variance weighted | 108 | -0.0137773 | 0.0202634 | 0.4965624 | TwoSample_MR | MDD | HDL_PL |
| Simple mode | 108 | 0.0563216 | 0.0690162 | 0.4162768 | TwoSample_MR | MDD | HDL_PL |
| Weighted mode | 108 | 0.0533411 | 0.0715225 | 0.4574266 | TwoSample_MR | MDD | HDL_PL |
| MR Egger | 108 | 0.0501171 | 0.0989441 | 0.6135441 | TwoSample_MR | MDD | HDL_TG |
| Weighted median | 108 | 0.0758253 | 0.0268244 | 0.0047026 | TwoSample_MR | MDD | HDL_TG |
| Inverse variance weighted | 108 | 0.084077 | 0.0217142 | 0.000108 | TwoSample_MR | MDD | HDL_TG |
| Simple mode | 108 | 0.1105927 | 0.079056 | 0.1647299 | TwoSample_MR | MDD | HDL_TG |
| Weighted mode | 108 | 0.1046043 | 0.0799921 | 0.1937836 | TwoSample_MR | MDD | HDL_TG |
| MR Egger | 108 | 0.1470527 | 0.0945139 | 0.1227163 | TwoSample_MR | MDD | HDL_size |
| Weighted median | 108 | -0.0365008 | 0.0238352 | 0.1256751 | TwoSample_MR | MDD | HDL_size |
| Inverse variance weighted | 108 | -0.0740735 | 0.021283 | 0.0005006 | TwoSample_MR | MDD | HDL_size |
| Simple mode | 108 | 0.0218232 | 0.0774008 | 0.7785259 | TwoSample_MR | MDD | HDL_size |
| Weighted mode | 108 | 0.0249414 | 0.0767589 | 0.7458676 | TwoSample_MR | MDD | HDL_size |
| MR Egger | 108 | -0.0983424 | 0.0905414 | 0.2798723 | TwoSample_MR | MDD | IDL_C |
| Weighted median | 108 | -0.0830309 | 0.0248814 | 0.0008467 | TwoSample_MR | MDD | IDL_C |
| Inverse variance weighted | 108 | -0.0624313 | 0.0198737 | 0.0016814 | TwoSample_MR | MDD | IDL_C |
| Simple mode | 108 | -0.1306796 | 0.0787589 | 0.0999968 | TwoSample_MR | MDD | IDL_C |
| Weighted mode | 108 | -0.1244711 | 0.0753567 | 0.1015154 | TwoSample_MR | MDD | IDL_C |
| MR Egger | 108 | -0.0979286 | 0.0892744 | 0.2751535 | TwoSample_MR | MDD | IDL_CE |
| Weighted median | 108 | -0.0724667 | 0.0246902 | 0.0033351 | TwoSample_MR | MDD | IDL_CE |
| Inverse variance weighted | 108 | -0.0597182 | 0.0195981 | 0.0023103 | TwoSample_MR | MDD | IDL_CE |

| | | | | | | | |
|---|---|---|---|---|---|---|---|
| Simple mode | 108 | -0.1309907 | 0.0749019 | 0.083188 | TwoSample_MR | MDD | IDL_CE |
| Weighted mode | 108 | -0.1250071 | 0.068924 | 0.0725274 | TwoSample_MR | MDD | IDL_CE |
| MR Egger | 108 | -0.078013 | 0.0891073 | 0.3832841 | TwoSample_MR | MDD | IDL_CE_pct |
| Weighted median | 108 | -0.0641769 | 0.024792 | 0.0096364 | TwoSample_MR | MDD | IDL_CE_pct |
| Inverse variance weighted | 108 | -0.0675203 | 0.0195448 | 0.000551 | TwoSample_MR | MDD | IDL_CE_pct |
| Simple mode | 108 | -0.0865604 | 0.0744784 | 0.2477307 | TwoSample_MR | MDD | IDL_CE_pct |
| Weighted mode | 108 | -0.0865604 | 0.082063 | 0.2938903 | TwoSample_MR | MDD | IDL_CE_pct |
| MR Egger | 108 | -0.096096 | 0.0972416 | 0.325296 | TwoSample_MR | MDD | IDL_C_pct |
| Weighted median | 108 | -0.0731037 | 0.0246137 | 0.0029776 | TwoSample_MR | MDD | IDL_C_pct |
| Inverse variance weighted | 108 | -0.0884259 | 0.0213292 | 0.0000339 | TwoSample_MR | MDD | IDL_C_pct |
| Simple mode | 108 | -0.0780528 | 0.0748092 | 0.299134 | TwoSample_MR | MDD | IDL_C_pct |
| Weighted mode | 108 | -0.0812766 | 0.0703026 | 0.2502166 | TwoSample_MR | MDD | IDL_C_pct |
| MR Egger | 108 | -0.0979512 | 0.0943394 | 0.3014999 | TwoSample_MR | MDD | IDL_FC |
| Weighted median | 108 | -0.086174 | 0.0249912 | 0.0005644 | TwoSample_MR | MDD | IDL_FC |
| Inverse variance weighted | 108 | -0.0687456 | 0.0207011 | 0.0008974 | TwoSample_MR | MDD | IDL_FC |
| Simple mode | 108 | -0.1351555 | 0.072624 | 0.065485 | TwoSample_MR | MDD | IDL_FC |
| Weighted mode | 108 | -0.1316883 | 0.0685825 | 0.057502 | TwoSample_MR | MDD | IDL_FC |
| MR Egger | 108 | -0.0394218 | 0.1099065 | 0.7205444 | TwoSample_MR | MDD | IDL_FC_pct |
| Weighted median | 108 | -0.0841705 | 0.0258263 | 0.0011177 | TwoSample_MR | MDD | IDL_FC_pct |
| Inverse variance weighted | 108 | -0.0916572 | 0.0241313 | 0.0001457 | TwoSample_MR | MDD | IDL_FC_pct |
| Simple mode | 108 | -0.1028821 | 0.075717 | 0.1770763 | TwoSample_MR | MDD | IDL_FC_pct |
| Weighted mode | 108 | -0.0865288 | 0.0742616 | 0.2465336 | TwoSample_MR | MDD | IDL_FC_pct |
| MR Egger | 108 | -0.0909133 | 0.088515 | 0.306714 | TwoSample_MR | MDD | IDL_L |
| Weighted median | 108 | -0.0573279 | 0.0243301 | 0.0184604 | TwoSample_MR | MDD | IDL_L |
| Inverse variance weighted | 108 | -0.0514176 | 0.0194329 | 0.0081471 | TwoSample_MR | MDD | IDL_L |
| Simple mode | 108 | -0.1160598 | 0.0722296 | 0.1110414 | TwoSample_MR | MDD | IDL_L |
| Weighted mode | 108 | -0.1099308 | 0.0700004 | 0.1192673 | TwoSample_MR | MDD | IDL_L |
| MR Egger | 108 | 0.0120357 | 0.0855689 | 0.8884089 | TwoSample_MR | MDD | IDL_TG |
| Weighted median | 108 | 0.0561034 | 0.0247333 | 0.0233092 | TwoSample_MR | MDD | IDL_TG |
| Inverse variance weighted | 108 | 0.0702384 | 0.0188108 | 0.0001885 | TwoSample_MR | MDD | IDL_TG |
| Simple mode | 108 | 0.0586139 | 0.0701398 | 0.405201 | TwoSample_MR | MDD | IDL_TG |
| Weighted mode | 108 | 0.0613588 | 0.0713622 | 0.3918094 | TwoSample_MR | MDD | IDL_TG |
| MR Egger | 108 | 0.1138111 | 0.1004108 | 0.2595805 | TwoSample_MR | MDD | IDL_TG_pct |
| Weighted median | 108 | 0.1207863 | 0.024937 | 0.0000013 | TwoSample_MR | MDD | IDL_TG_pct |
| Inverse variance weighted | 108 | 0.1095837 | 0.0220248 | 0.0000007 | TwoSample_MR | MDD | IDL_TG_pct |
| Simple mode | 108 | 0.1404439 | 0.0633561 | 0.0287568 | TwoSample_MR | MDD | IDL_TG_pct |
| Weighted mode | 108 | 0.1404439 | 0.0628182 | 0.0274462 | TwoSample_MR | MDD | IDL_TG_pct |
| MR Egger | 108 | 0.1545073 | 0.1382876 | 0.2663965 | TwoSample_MR | MDD | LA_pct |
| Weighted median | 108 | -0.0957583 | 0.023778 | 0.0000564 | TwoSample_MR | MDD | LA_pct |
| Inverse variance weighted | 108 | -0.0772879 | 0.0307493 | 0.0119546 | TwoSample_MR | MDD | LA_pct |
| Simple mode | 108 | -0.1016282 | 0.0640287 | 0.1154114 | TwoSample_MR | MDD | LA_pct |
| Weighted mode | 108 | -0.1081691 | 0.0701199 | 0.1258728 | TwoSample_MR | MDD | LA_pct |
| MR Egger | 108 | 0.1410849 | 0.0964643 | 0.1465462 | TwoSample_MR | MDD | L_HDL_C |
| Weighted median | 108 | -0.0435033 | 0.0233403 | 0.0623397 | TwoSample_MR | MDD | L_HDL_C |
| Inverse variance weighted | 108 | -0.0863337 | 0.0217314 | 0.000071 | TwoSample_MR | MDD | L_HDL_C |
| Simple mode | 108 | -0.0168701 | 0.0740831 | 0.8202993 | TwoSample_MR | MDD | L_HDL_C |

| | | | | | | | |
|---|---|---|---|---|---|---|---|
| Weighted mode | 108 | -0.0135855 | 0.075415 | 0.857381 | TwoSample_MR | MDD | L_HDL_C |
| MR Egger | 108 | 0.1443342 | 0.0968985 | 0.1393146 | TwoSample_MR | MDD | L_HDL_CE |
| Weighted median | 108 | -0.050163 | 0.0227944 | 0.0277592 | TwoSample_MR | MDD | L_HDL_CE |
| Inverse variance weighted | 108 | -0.0880154 | 0.0218492 | 0.0000562 | TwoSample_MR | MDD | L_HDL_CE |
| Simple mode | 108 | -0.0239176 | 0.069866 | 0.7327695 | TwoSample_MR | MDD | L_HDL_CE |
| Weighted mode | 108 | -0.0239176 | 0.077263 | 0.7574966 | TwoSample_MR | MDD | L_HDL_CE |
| MR Egger | 108 | 0.0515869 | 0.1033034 | 0.6185518 | TwoSample_MR | MDD | L_HDL_CE_pct |
| Weighted median | 108 | -0.1071638 | 0.0253817 | 0.0000242 | TwoSample_MR | MDD | L_HDL_CE_pct |
| Inverse variance weighted | 108 | -0.1020077 | 0.0229052 | 0.0000084 | TwoSample_MR | MDD | L_HDL_CE_pct |
| Simple mode | 108 | -0.1784611 | 0.0682303 | 0.0101941 | TwoSample_MR | MDD | L_HDL_CE_pct |
| Weighted mode | 108 | -0.1467247 | 0.0591227 | 0.0146314 | TwoSample_MR | MDD | L_HDL_CE_pct |
| MR Egger | 108 | 0.041952 | 0.1059549 | 0.6929438 | TwoSample_MR | MDD | L_HDL_C_pct |
| Weighted median | 108 | -0.1159401 | 0.0251205 | 0.0000039 | TwoSample_MR | MDD | L_HDL_C_pct |
| Inverse variance weighted | 108 | -0.1106657 | 0.0234781 | 0.0000024 | TwoSample_MR | MDD | L_HDL_C_pct |
| Simple mode | 108 | -0.1748162 | 0.0730761 | 0.0184894 | TwoSample_MR | MDD | L_HDL_C_pct |
| Weighted mode | 108 | -0.1588416 | 0.061951 | 0.0117355 | TwoSample_MR | MDD | L_HDL_C_pct |
| MR Egger | 108 | 0.1285039 | 0.0946317 | 0.1773674 | TwoSample_MR | MDD | L_HDL_FC |
| Weighted median | 108 | -0.0352733 | 0.0227787 | 0.1214976 | TwoSample_MR | MDD | L_HDL_FC |
| Inverse variance weighted | 108 | -0.0793562 | 0.0212444 | 0.0001874 | TwoSample_MR | MDD | L_HDL_FC |
| Simple mode | 108 | 0.0121276 | 0.0803056 | 0.8802463 | TwoSample_MR | MDD | L_HDL_FC |
| Weighted mode | 108 | 0.0152071 | 0.082698 | 0.8544502 | TwoSample_MR | MDD | L_HDL_FC |
| MR Egger | 108 | -0.0544252 | 0.1013622 | 0.5924355 | TwoSample_MR | MDD | L_HDL_FC_pct |
| Weighted median | 108 | -0.0923776 | 0.0236264 | 0.0000923 | TwoSample_MR | MDD | L_HDL_FC_pct |
| Inverse variance weighted | 108 | -0.1017455 | 0.0222548 | 0.0000048 | TwoSample_MR | MDD | L_HDL_FC_pct |
| Simple mode | 108 | -0.0838145 | 0.0673745 | 0.2162143 | TwoSample_MR | MDD | L_HDL_FC_pct |
| Weighted mode | 108 | -0.0995847 | 0.0666348 | 0.1379921 | TwoSample_MR | MDD | L_HDL_FC_pct |
| MR Egger | 108 | 0.1558268 | 0.0937306 | 0.0993676 | TwoSample_MR | MDD | L_HDL_L |
| Weighted median | 108 | -0.0113803 | 0.0231007 | 0.622267 | TwoSample_MR | MDD | L_HDL_L |
| Inverse variance weighted | 108 | -0.0716527 | 0.0211479 | 0.0007036 | TwoSample_MR | MDD | L_HDL_L |
| Simple mode | 108 | 0.0252173 | 0.0778113 | 0.7465081 | TwoSample_MR | MDD | L_HDL_L |
| Weighted mode | 108 | 0.0282564 | 0.0788492 | 0.7207797 | TwoSample_MR | MDD | L_HDL_L |
| MR Egger | 108 | 0.1465677 | 0.0955352 | 0.1279662 | TwoSample_MR | MDD | L_HDL_P |
| Weighted median | 108 | -0.0210933 | 0.0228583 | 0.35612 | TwoSample_MR | MDD | L_HDL_P |
| Inverse variance weighted | 108 | -0.0750282 | 0.0215035 | 0.0004846 | TwoSample_MR | MDD | L_HDL_P |
| Simple mode | 108 | 0.0222688 | 0.0805927 | 0.7828411 | TwoSample_MR | MDD | L_HDL_P |
| Weighted mode | 108 | 0.0253913 | 0.0821908 | 0.7579749 | TwoSample_MR | MDD | L_HDL_P |
| MR Egger | 108 | 0.1644614 | 0.0915849 | 0.0753873 | TwoSample_MR | MDD | L_HDL_PL |
| Weighted median | 108 | -0.0119261 | 0.0235147 | 0.6120322 | TwoSample_MR | MDD | L_HDL_PL |
| Inverse variance weighted | 108 | -0.0622339 | 0.0206865 | 0.002626 | TwoSample_MR | MDD | L_HDL_PL |
| Simple mode | 108 | 0.0409936 | 0.0775018 | 0.5979438 | TwoSample_MR | MDD | L_HDL_PL |
| Weighted mode | 108 | 0.0439161 | 0.0826458 | 0.596259 | TwoSample_MR | MDD | L_HDL_PL |
| MR Egger | 108 | 0.0007278 | 0.105004 | 0.994483 | TwoSample_MR | MDD | L_HDL_PL_pct |
| Weighted median | 108 | 0.1123032 | 0.0251392 | 0.0000079 | TwoSample_MR | MDD | L_HDL_PL_pct |
| Inverse variance weighted | 108 | 0.1050653 | 0.023144 | 0.0000056 | TwoSample_MR | MDD | L_HDL_PL_pct |
| Simple mode | 108 | 0.1263067 | 0.073794 | 0.0898651 | TwoSample_MR | MDD | L_HDL_PL_pct |
| Weighted mode | 108 | 0.1403004 | 0.0606969 | 0.0227215 | TwoSample_MR | MDD | L_HDL_PL_pct |

| | | | | | | | |
|---|---|---|---|---|---|---|---|
| MR Egger | 108 | -0.0746732 | 0.0971104 | 0.4436322 | TwoSample_MR | MDD | L_HDL_TG_pct |
| Weighted median | 108 | 0.0809663 | 0.0242452 | 0.0008394 | TwoSample_MR | MDD | L_HDL_TG_pct |
| Inverse variance weighted | 108 | 0.0936494 | 0.0216137 | 0.0000147 | TwoSample_MR | MDD | L_HDL_TG_pct |
| Simple mode | 108 | 0.0500048 | 0.0669582 | 0.4568181 | TwoSample_MR | MDD | L_HDL_TG_pct |
| Weighted mode | 108 | 0.0647889 | 0.0565783 | 0.254715 | TwoSample_MR | MDD | L_HDL_TG_pct |
| MR Egger | 108 | 0.1145354 | 0.0943719 | 0.2275748 | TwoSample_MR | MDD | L_LDL_TG_pct |
| Weighted median | 108 | 0.1080903 | 0.0249059 | 0.0000143 | TwoSample_MR | MDD | L_LDL_TG_pct |
| Inverse variance weighted | 108 | 0.1013585 | 0.0207008 | 0.000001 | TwoSample_MR | MDD | L_LDL_TG_pct |
| Simple mode | 108 | 0.1260947 | 0.0628856 | 0.0474723 | TwoSample_MR | MDD | L_LDL_TG_pct |
| Weighted mode | 108 | 0.1229554 | 0.0539227 | 0.0245772 | TwoSample_MR | MDD | L_LDL_TG_pct |
| MR Egger | 108 | -0.1227203 | 0.0875531 | 0.1639359 | TwoSample_MR | MDD | L_VLDL_C |
| Weighted median | 108 | 0.0435371 | 0.0244481 | 0.0749453 | TwoSample_MR | MDD | L_VLDL_C |
| Inverse variance weighted | 108 | 0.0681648 | 0.019654 | 0.0005239 | TwoSample_MR | MDD | L_VLDL_C |
| Simple mode | 108 | 0.0556422 | 0.0635899 | 0.3835252 | TwoSample_MR | MDD | L_VLDL_C |
| Weighted mode | 108 | 0.0497386 | 0.0627523 | 0.4297544 | TwoSample_MR | MDD | L_VLDL_C |
| MR Egger | 108 | -0.1265369 | 0.0850667 | 0.139851 | TwoSample_MR | MDD | L_VLDL_CE |
| Weighted median | 108 | 0.0307575 | 0.0240447 | 0.2008335 | TwoSample_MR | MDD | L_VLDL_CE |
| Inverse variance weighted | 108 | 0.0513737 | 0.0190606 | 0.0070328 | TwoSample_MR | MDD | L_VLDL_CE |
| Simple mode | 108 | 0.0047159 | 0.0675466 | 0.9444696 | TwoSample_MR | MDD | L_VLDL_CE |
| Weighted mode | 108 | -0.0006985 | 0.0650572 | 0.991454 | TwoSample_MR | MDD | L_VLDL_CE |
| MR Egger | 108 | 0.0465489 | 0.0979284 | 0.6355258 | TwoSample_MR | MDD | L_VLDL_CE_pct |
| Weighted median | 108 | -0.1139866 | 0.0244245 | 0.0000031 | TwoSample_MR | MDD | L_VLDL_CE_pct |
| Inverse variance weighted | 108 | -0.0944112 | 0.0217034 | 0.0000136 | TwoSample_MR | MDD | L_VLDL_CE_pct |
| Simple mode | 108 | -0.1181823 | 0.0623581 | 0.060762 | TwoSample_MR | MDD | L_VLDL_CE_pct |
| Weighted mode | 108 | -0.1212848 | 0.0584876 | 0.0405097 | TwoSample_MR | MDD | L_VLDL_CE_pct |
| MR Egger | 108 | -0.1182685 | 0.0905663 | 0.1944209 | TwoSample_MR | MDD | L_VLDL_FC |
| Weighted median | 108 | 0.0536379 | 0.0248893 | 0.0311572 | TwoSample_MR | MDD | L_VLDL_FC |
| Inverse variance weighted | 108 | 0.0813884 | 0.0203417 | 0.0000631 | TwoSample_MR | MDD | L_VLDL_FC |
| Simple mode | 108 | 0.0741148 | 0.0699319 | 0.2916154 | TwoSample_MR | MDD | L_VLDL_FC |
| Weighted mode | 108 | 0.0678387 | 0.0648711 | 0.2980351 | TwoSample_MR | MDD | L_VLDL_FC |
| MR Egger | 108 | -0.1234773 | 0.0919666 | 0.1822591 | TwoSample_MR | MDD | L_VLDL_L |
| Weighted median | 108 | 0.0651641 | 0.0234916 | 0.0055384 | TwoSample_MR | MDD | L_VLDL_L |
| Inverse variance weighted | 108 | 0.0832398 | 0.0206756 | 0.0000567 | TwoSample_MR | MDD | L_VLDL_L |
| Simple mode | 108 | 0.0817419 | 0.065952 | 0.217903 | TwoSample_MR | MDD | L_VLDL_L |
| Weighted mode | 108 | 0.0724768 | 0.0638901 | 0.2591623 | TwoSample_MR | MDD | L_VLDL_L |
| MR Egger | 108 | -0.1092083 | 0.0917915 | 0.2368054 | TwoSample_MR | MDD | L_VLDL_P |
| Weighted median | 108 | 0.06391 | 0.0240758 | 0.0079417 | TwoSample_MR | MDD | L_VLDL_P |
| Inverse variance weighted | 108 | 0.0856809 | 0.0205831 | 0.0000315 | TwoSample_MR | MDD | L_VLDL_P |
| Simple mode | 108 | 0.0749635 | 0.0672712 | 0.2676268 | TwoSample_MR | MDD | L_VLDL_P |
| Weighted mode | 108 | 0.0687351 | 0.069396 | 0.3241758 | TwoSample_MR | MDD | L_VLDL_P |
| MR Egger | 108 | -0.1126758 | 0.0928903 | 0.2278267 | TwoSample_MR | MDD | L_VLDL_PL |
| Weighted median | 108 | 0.0706091 | 0.0237978 | 0.0030068 | TwoSample_MR | MDD | L_VLDL_PL |
| Inverse variance weighted | 108 | 0.0876494 | 0.0208434 | 0.0000261 | TwoSample_MR | MDD | L_VLDL_PL |
| Simple mode | 108 | 0.0800075 | 0.0648767 | 0.2201944 | TwoSample_MR | MDD | L_VLDL_PL |
| Weighted mode | 108 | 0.0800075 | 0.0632333 | 0.2085213 | TwoSample_MR | MDD | L_VLDL_PL |
| MR Egger | 108 | -0.0381771 | 0.0947581 | 0.6878406 | TwoSample_MR | MDD | L_VLDL_PL_pct |

| | | | | | | | |
|---|---|---|---|---|---|---|---|
| Weighted median | 108 | 0.0654189 | 0.0242023 | 0.0068715 | TwoSample_MR | MDD | L_VLDL_PL_pct |
| Inverse variance weighted | 108 | 0.0806011 | 0.020945 | 0.000119 | TwoSample_MR | MDD | L_VLDL_PL_pct |
| Simple mode | 108 | 0.0364949 | 0.0676986 | 0.5909526 | TwoSample_MR | MDD | L_VLDL_PL_pct |
| Weighted mode | 108 | 0.0426041 | 0.0684169 | 0.5347984 | TwoSample_MR | MDD | L_VLDL_PL_pct |
| MR Egger | 108 | -0.1293194 | 0.0930533 | 0.1675215 | TwoSample_MR | MDD | L_VLDL_TG |
| Weighted median | 108 | 0.0605391 | 0.0232529 | 0.0092277 | TwoSample_MR | MDD | L_VLDL_TG |
| Inverse variance weighted | 108 | 0.0864613 | 0.0209528 | 0.0000368 | TwoSample_MR | MDD | L_VLDL_TG |
| Simple mode | 108 | 0.0887755 | 0.0675313 | 0.1914608 | TwoSample_MR | MDD | L_VLDL_TG |
| Weighted mode | 108 | 0.0764212 | 0.0639551 | 0.2347601 | TwoSample_MR | MDD | L_VLDL_TG |
| MR Egger | 108 | 0.1881659 | 0.090788 | 0.0406366 | TwoSample_MR | MDD | M_HDL_C |
| Weighted median | 108 | -0.0196553 | 0.0239132 | 0.4111097 | TwoSample_MR | MDD | M_HDL_C |
| Inverse variance weighted | 108 | -0.0169035 | 0.0204084 | 0.4075201 | TwoSample_MR | MDD | M_HDL_C |
| Simple mode | 108 | -0.0026508 | 0.0738528 | 0.9714349 | TwoSample_MR | MDD | M_HDL_C |
| Weighted mode | 108 | 0.0063829 | 0.0691615 | 0.9266408 | TwoSample_MR | MDD | M_HDL_C |
| MR Egger | 108 | 0.1909571 | 0.0904527 | 0.0371127 | TwoSample_MR | MDD | M_HDL_CE |
| Weighted median | 108 | -0.0253394 | 0.022908 | 0.2686678 | TwoSample_MR | MDD | M_HDL_CE |
| Inverse variance weighted | 108 | -0.0172107 | 0.0203518 | 0.3977422 | TwoSample_MR | MDD | M_HDL_CE |
| Simple mode | 108 | -0.0559723 | 0.0712821 | 0.4340579 | TwoSample_MR | MDD | M_HDL_CE |
| Weighted mode | 108 | -0.0438774 | 0.0716685 | 0.5416866 | TwoSample_MR | MDD | M_HDL_CE |
| MR Egger | 108 | 0.0749979 | 0.0985669 | 0.4484164 | TwoSample_MR | MDD | M_HDL_CE_pct |
| Weighted median | 108 | -0.0771762 | 0.025148 | 0.0021487 | TwoSample_MR | MDD | M_HDL_CE_pct |
| Inverse variance weighted | 108 | -0.0748195 | 0.0218655 | 0.0006221 | TwoSample_MR | MDD | M_HDL_CE_pct |
| Simple mode | 108 | -0.1372911 | 0.0723485 | 0.0604394 | TwoSample_MR | MDD | M_HDL_CE_pct |
| Weighted mode | 108 | -0.1300355 | 0.0697604 | 0.0650586 | TwoSample_MR | MDD | M_HDL_CE_pct |
| MR Egger | 108 | 0.085855 | 0.1008401 | 0.3964681 | TwoSample_MR | MDD | M_HDL_C_pct |
| Weighted median | 108 | -0.0673699 | 0.0238899 | 0.0048022 | TwoSample_MR | MDD | M_HDL_C_pct |
| Inverse variance weighted | 108 | -0.082856 | 0.0224232 | 0.0002198 | TwoSample_MR | MDD | M_HDL_C_pct |
| Simple mode | 108 | -0.1619271 | 0.0745709 | 0.032106 | TwoSample_MR | MDD | M_HDL_C_pct |
| Weighted mode | 108 | -0.1367961 | 0.0650553 | 0.0378315 | TwoSample_MR | MDD | M_HDL_C_pct |
| MR Egger | 108 | 0.1734277 | 0.0915816 | 0.0609918 | TwoSample_MR | MDD | M_HDL_FC |
| Weighted median | 108 | -0.0036625 | 0.0233461 | 0.8753404 | TwoSample_MR | MDD | M_HDL_FC |
| Inverse variance weighted | 108 | -0.015694 | 0.0205048 | 0.444046 | TwoSample_MR | MDD | M_HDL_FC |
| Simple mode | 108 | 0.0483581 | 0.0741906 | 0.5159219 | TwoSample_MR | MDD | M_HDL_FC |
| Weighted mode | 108 | 0.0513748 | 0.0747136 | 0.4931781 | TwoSample_MR | MDD | M_HDL_FC |
| MR Egger | 108 | 0.0863751 | 0.0988655 | 0.3842769 | TwoSample_MR | MDD | M_HDL_FC_pct |
| Weighted median | 108 | -0.0406672 | 0.0229151 | 0.0759497 | TwoSample_MR | MDD | M_HDL_FC_pct |
| Inverse variance weighted | 108 | -0.0688896 | 0.0219462 | 0.0016951 | TwoSample_MR | MDD | M_HDL_FC_pct |
| Simple mode | 108 | -0.0339502 | 0.0755006 | 0.6538597 | TwoSample_MR | MDD | M_HDL_FC_pct |
| Weighted mode | 108 | -0.0270182 | 0.0720793 | 0.7085213 | TwoSample_MR | MDD | M_HDL_FC_pct |
| MR Egger | 108 | 0.1873088 | 0.0914509 | 0.0430128 | TwoSample_MR | MDD | M_HDL_P |
| Weighted median | 108 | 0.004825 | 0.0233695 | 0.8364274 | TwoSample_MR | MDD | M_HDL_P |
| Inverse variance weighted | 108 | -0.0048295 | 0.0204904 | 0.8136687 | TwoSample_MR | MDD | M_HDL_P |
| Simple mode | 108 | 0.0704548 | 0.0752707 | 0.3513716 | TwoSample_MR | MDD | M_HDL_P |
| Weighted mode | 108 | 0.0704548 | 0.0757175 | 0.3542096 | TwoSample_MR | MDD | M_HDL_P |
| MR Egger | 108 | -0.0893854 | 0.1027949 | 0.3865108 | TwoSample_MR | MDD | M_HDL_PL_pct |
| Weighted median | 108 | 0.0721815 | 0.0241125 | 0.0027577 | TwoSample_MR | MDD | M_HDL_PL_pct |

| | | | | | | | |
|---|---|---|---|---|---|---|---|
| Inverse variance weighted | 108 | 0.0787633 | 0.0228444 | 0.0005651 | TwoSample_MR | MDD | M_HDL_PL_pct |
| Simple mode | 108 | 0.063235 | 0.0703901 | 0.3710154 | TwoSample_MR | MDD | M_HDL_PL_pct |
| Weighted mode | 108 | 0.0414411 | 0.0622148 | 0.5067833 | TwoSample_MR | MDD | M_HDL_PL_pct |
| MR Egger | 108 | 0.0575156 | 0.1034937 | 0.5795595 | TwoSample_MR | MDD | M_HDL_TG |
| Weighted median | 108 | 0.0854352 | 0.0259983 | 0.0010155 | TwoSample_MR | MDD | M_HDL_TG |
| Inverse variance weighted | 108 | 0.0863929 | 0.0227083 | 0.0001421 | TwoSample_MR | MDD | M_HDL_TG |
| Simple mode | 108 | 0.1305428 | 0.08081 | 0.1091626 | TwoSample_MR | MDD | M_HDL_TG |
| Weighted mode | 108 | 0.1242661 | 0.0788259 | 0.1178712 | TwoSample_MR | MDD | M_HDL_TG |
| MR Egger | 108 | -0.0698796 | 0.0973938 | 0.4746467 | TwoSample_MR | MDD | M_HDL_TG_pct |
| Weighted median | 108 | 0.0839568 | 0.0253043 | 0.000907 | TwoSample_MR | MDD | M_HDL_TG_pct |
| Inverse variance weighted | 108 | 0.0815401 | 0.0216162 | 0.0001618 | TwoSample_MR | MDD | M_HDL_TG_pct |
| Simple mode | 108 | 0.139983 | 0.0693724 | 0.0461099 | TwoSample_MR | MDD | M_HDL_TG_pct |
| Weighted mode | 108 | 0.1331371 | 0.0679042 | 0.0525181 | TwoSample_MR | MDD | M_HDL_TG_pct |
| MR Egger | 108 | -0.0107243 | 0.0870141 | 0.9021438 | TwoSample_MR | MDD | M_LDL_TG |
| Weighted median | 108 | 0.0777509 | 0.0250126 | 0.0018806 | TwoSample_MR | MDD | M_LDL_TG |
| Inverse variance weighted | 108 | 0.0869823 | 0.0192062 | 0.0000059 | TwoSample_MR | MDD | M_LDL_TG |
| Simple mode | 108 | 0.0737938 | 0.0720257 | 0.307887 | TwoSample_MR | MDD | M_LDL_TG |
| Weighted mode | 108 | 0.0620622 | 0.0693323 | 0.3727206 | TwoSample_MR | MDD | M_LDL_TG |
| MR Egger | 108 | 0.1536468 | 0.093331 | 0.1026728 | TwoSample_MR | MDD | M_LDL_TG_pct |
| Weighted median | 108 | 0.1019543 | 0.0243846 | 0.000029 | TwoSample_MR | MDD | M_LDL_TG_pct |
| Inverse variance weighted | 108 | 0.0950515 | 0.0205103 | 0.0000036 | TwoSample_MR | MDD | M_LDL_TG_pct |
| Simple mode | 108 | 0.1058199 | 0.0666779 | 0.1154564 | TwoSample_MR | MDD | M_LDL_TG_pct |
| Weighted mode | 108 | 0.1028082 | 0.0616449 | 0.09829 | TwoSample_MR | MDD | M_LDL_TG_pct |
| MR Egger | 108 | 0.0168086 | 0.1001315 | 0.8670089 | TwoSample_MR | MDD | MUFA |
| Weighted median | 108 | 0.0984043 | 0.0251163 | 0.0000893 | TwoSample_MR | MDD | MUFA |
| Inverse variance weighted | 108 | 0.1058278 | 0.022052 | 0.0000016 | TwoSample_MR | MDD | MUFA |
| Simple mode | 108 | 0.1148532 | 0.0811735 | 0.1599984 | TwoSample_MR | MDD | MUFA |
| Weighted mode | 108 | 0.1117892 | 0.0772001 | 0.1505286 | TwoSample_MR | MDD | MUFA |
| MR Egger | 108 | 0.0307271 | 0.1297817 | 0.8133001 | TwoSample_MR | MDD | MUFA_pct |
| Weighted median | 108 | 0.1156114 | 0.0250831 | 0.000004 | TwoSample_MR | MDD | MUFA_pct |
| Inverse variance weighted | 108 | 0.1451301 | 0.028578 | 0.0000004 | TwoSample_MR | MDD | MUFA_pct |
| Simple mode | 108 | 0.1492265 | 0.0693612 | 0.0336909 | TwoSample_MR | MDD | MUFA_pct |
| Weighted mode | 108 | 0.1159106 | 0.0744271 | 0.1223356 | TwoSample_MR | MDD | MUFA_pct |
| MR Egger | 108 | -0.0131421 | 0.1043882 | 0.9000523 | TwoSample_MR | MDD | M_VLDL_CE_pct |
| Weighted median | 108 | -0.1033709 | 0.0237005 | 0.0000129 | TwoSample_MR | MDD | M_VLDL_CE_pct |
| Inverse variance weighted | 108 | -0.1090868 | 0.0229949 | 0.0000021 | TwoSample_MR | MDD | M_VLDL_CE_pct |
| Simple mode | 108 | -0.1140899 | 0.0597673 | 0.0589541 | TwoSample_MR | MDD | M_VLDL_CE_pct |
| Weighted mode | 108 | -0.1209549 | 0.0525263 | 0.0232274 | TwoSample_MR | MDD | M_VLDL_CE_pct |
| MR Egger | 108 | -0.0244469 | 0.1047959 | 0.8159927 | TwoSample_MR | MDD | M_VLDL_C_pct |
| Weighted median | 108 | -0.1127586 | 0.0238814 | 0.0000023 | TwoSample_MR | MDD | M_VLDL_C_pct |
| Inverse variance weighted | 108 | -0.1090252 | 0.0230624 | 0.0000023 | TwoSample_MR | MDD | M_VLDL_C_pct |
| Simple mode | 108 | -0.1218218 | 0.0644426 | 0.0614111 | TwoSample_MR | MDD | M_VLDL_C_pct |
| Weighted mode | 108 | -0.1285208 | 0.0543099 | 0.0197592 | TwoSample_MR | MDD | M_VLDL_C_pct |
| MR Egger | 108 | -0.0525862 | 0.1049803 | 0.6174696 | TwoSample_MR | MDD | M_VLDL_FC_pct |
| Weighted median | 108 | -0.103706 | 0.0244395 | 0.000022 | TwoSample_MR | MDD | M_VLDL_FC_pct |
| Inverse variance weighted | 108 | -0.1041035 | 0.0230546 | 0.0000063 | TwoSample_MR | MDD | M_VLDL_FC_pct |

| | | | | | | | |
|---|---|---|---|---|---|---|---|
| Simple mode | 108 | -0.1033556 | 0.0649612 | 0.1145516 | TwoSample_MR | MDD | M_VLDL_FC_pct |
| Weighted mode | 108 | -0.1105225 | 0.0547139 | 0.0458812 | TwoSample_MR | MDD | M_VLDL_FC_pct |
| MR Egger | 108 | -0.1246501 | 0.0833473 | 0.137742 | TwoSample_MR | MDD | M_VLDL_L |
| Weighted median | 108 | 0.0325507 | 0.0238738 | 0.1727401 | TwoSample_MR | MDD | M_VLDL_L |
| Inverse variance weighted | 108 | 0.0439476 | 0.0186503 | 0.0184532 | TwoSample_MR | MDD | M_VLDL_L |
| Simple mode | 108 | 0.0195537 | 0.0691544 | 0.7779114 | TwoSample_MR | MDD | M_VLDL_L |
| Weighted mode | 108 | 0.0170744 | 0.0643851 | 0.7913723 | TwoSample_MR | MDD | M_VLDL_L |
| MR Egger | 108 | -0.0930682 | 0.0915826 | 0.3118379 | TwoSample_MR | MDD | M_VLDL_TG |
| Weighted median | 108 | 0.0547751 | 0.0246014 | 0.0259806 | TwoSample_MR | MDD | M_VLDL_TG |
| Inverse variance weighted | 108 | 0.0825865 | 0.0204545 | 0.000054 | TwoSample_MR | MDD | M_VLDL_TG |
| Simple mode | 108 | 0.0440557 | 0.0684974 | 0.5214874 | TwoSample_MR | MDD | M_VLDL_TG |
| Weighted mode | 108 | 0.0348506 | 0.0625724 | 0.5787152 | TwoSample_MR | MDD | M_VLDL_TG |
| MR Egger | 108 | 0.0344883 | 0.1036421 | 0.7399702 | TwoSample_MR | MDD | M_VLDL_TG_pct |
| Weighted median | 108 | 0.1120856 | 0.024167 | 0.0000035 | TwoSample_MR | MDD | M_VLDL_TG_pct |
| Inverse variance weighted | 108 | 0.1059606 | 0.022788 | 0.0000033 | TwoSample_MR | MDD | M_VLDL_TG_pct |
| Simple mode | 108 | 0.1171951 | 0.0568911 | 0.0418262 | TwoSample_MR | MDD | M_VLDL_TG_pct |
| Weighted mode | 108 | 0.1237836 | 0.0538859 | 0.023557 | TwoSample_MR | MDD | M_VLDL_TG_pct |
| MR Egger | 108 | 0.0714435 | 0.0997078 | 0.4752407 | TwoSample_MR | MDD | Omega_6_pct |
| Weighted median | 108 | -0.0810268 | 0.0247871 | 0.0010796 | TwoSample_MR | MDD | Omega_6_pct |
| Inverse variance weighted | 108 | -0.1008933 | 0.022193 | 0.0000055 | TwoSample_MR | MDD | Omega_6_pct |
| Simple mode | 108 | -0.0803998 | 0.064071 | 0.2122656 | TwoSample_MR | MDD | Omega_6_pct |
| Weighted mode | 108 | -0.0803998 | 0.0649831 | 0.2187058 | TwoSample_MR | MDD | Omega_6_pct |
| MR Egger | 108 | 0.0152848 | 0.1236368 | 0.9018453 | TwoSample_MR | MDD | PUFA_by_MUFA |
| Weighted median | 108 | -0.1206485 | 0.0248057 | 0.0000012 | TwoSample_MR | MDD | PUFA_by_MUFA |
| Inverse variance weighted | 108 | -0.1359765 | 0.0273214 | 0.0000006 | TwoSample_MR | MDD | PUFA_by_MUFA |
| Simple mode | 108 | -0.1566297 | 0.0740667 | 0.0367768 | TwoSample_MR | MDD | PUFA_by_MUFA |
| Weighted mode | 108 | -0.146574 | 0.0714334 | 0.042621 | TwoSample_MR | MDD | PUFA_by_MUFA |
| MR Egger | 108 | 0.0784115 | 0.1126044 | 0.4877373 | TwoSample_MR | MDD | PUFA_pct |
| Weighted median | 108 | -0.0877099 | 0.0259054 | 0.0007098 | TwoSample_MR | MDD | PUFA_pct |
| Inverse variance weighted | 108 | -0.1142222 | 0.0250558 | 0.0000051 | TwoSample_MR | MDD | PUFA_pct |
| Simple mode | 108 | -0.1247905 | 0.0754478 | 0.1010588 | TwoSample_MR | MDD | PUFA_pct |
| Weighted mode | 108 | -0.1016634 | 0.0714972 | 0.1579576 | TwoSample_MR | MDD | PUFA_pct |
| MR Egger | 108 | 0.0531089 | 0.0801549 | 0.5090386 | TwoSample_MR | MDD | Pyruvate |
| Weighted median | 108 | 0.0293892 | 0.0250656 | 0.2409997 | TwoSample_MR | MDD | Pyruvate |
| Inverse variance weighted | 108 | 0.0298772 | 0.0175862 | 0.0893376 | TwoSample_MR | MDD | Pyruvate |
| Simple mode | 108 | 0.005308 | 0.0703141 | 0.9399662 | TwoSample_MR | MDD | Pyruvate |
| Weighted mode | 108 | 0.0178188 | 0.0704937 | 0.8009296 | TwoSample_MR | MDD | Pyruvate |
| MR Egger | 108 | -0.0518511 | 0.0972319 | 0.5949614 | TwoSample_MR | MDD | S_HDL_C_pct |
| Weighted median | 108 | -0.0806612 | 0.0255207 | 0.0015743 | TwoSample_MR | MDD | S_HDL_C_pct |
| Inverse variance weighted | 108 | -0.0731061 | 0.0213308 | 0.0006097 | TwoSample_MR | MDD | S_HDL_C_pct |
| Simple mode | 108 | -0.0930532 | 0.0772158 | 0.2308211 | TwoSample_MR | MDD | S_HDL_C_pct |
| Weighted mode | 108 | -0.096331 | 0.0780051 | 0.2195604 | TwoSample_MR | MDD | S_HDL_C_pct |
| MR Egger | 108 | 0.0038405 | 0.096705 | 0.9683959 | TwoSample_MR | MDD | S_HDL_FC_pct |
| Weighted median | 108 | -0.0645623 | 0.0234731 | 0.0059508 | TwoSample_MR | MDD | S_HDL_FC_pct |
| Inverse variance weighted | 108 | -0.0678958 | 0.021267 | 0.0014102 | TwoSample_MR | MDD | S_HDL_FC_pct |
| Simple mode | 108 | -0.0871921 | 0.0677903 | 0.201148 | TwoSample_MR | MDD | S_HDL_FC_pct |

| | | | | | | | |
|---|---|---|---|---|---|---|---|
| Weighted mode | 108 | -0.0871921 | 0.060875 | 0.1549687 | TwoSample_MR | MDD | S_HDL_FC_pct |
| MR Egger | 108 | -0.0122485 | 0.1044066 | 0.9068324 | TwoSample_MR | MDD | S_HDL_TG |
| Weighted median | 108 | 0.0974063 | 0.026381 | 0.0002222 | TwoSample_MR | MDD | S_HDL_TG |
| Inverse variance weighted | 108 | 0.1083879 | 0.0230536 | 0.0000026 | TwoSample_MR | MDD | S_HDL_TG |
| Simple mode | 108 | 0.1743372 | 0.0840084 | 0.0403628 | TwoSample_MR | MDD | S_HDL_TG |
| Weighted mode | 108 | 0.1534657 | 0.0842359 | 0.0712703 | TwoSample_MR | MDD | S_HDL_TG |
| MR Egger | 108 | -0.0569276 | 0.0964989 | 0.5564928 | TwoSample_MR | MDD | S_HDL_TG_pct |
| Weighted median | 108 | 0.1054354 | 0.0238596 | 0.0000099 | TwoSample_MR | MDD | S_HDL_TG_pct |
| Inverse variance weighted | 108 | 0.0901884 | 0.0214091 | 0.0000252 | TwoSample_MR | MDD | S_HDL_TG_pct |
| Simple mode | 108 | 0.1295313 | 0.06991 | 0.0666625 | TwoSample_MR | MDD | S_HDL_TG_pct |
| Weighted mode | 108 | 0.1295313 | 0.0620763 | 0.0392964 | TwoSample_MR | MDD | S_HDL_TG_pct |
| MR Egger | 108 | -0.0591222 | 0.0992667 | 0.5527193 | TwoSample_MR | MDD | S_LDL_PL_pct |
| Weighted median | 108 | -0.0632553 | 0.0251198 | 0.0117976 | TwoSample_MR | MDD | S_LDL_PL_pct |
| Inverse variance weighted | 108 | -0.0705468 | 0.0217752 | 0.0011962 | TwoSample_MR | MDD | S_LDL_PL_pct |
| Simple mode | 108 | -0.0950822 | 0.0799242 | 0.2368153 | TwoSample_MR | MDD | S_LDL_PL_pct |
| Weighted mode | 108 | -0.0856519 | 0.0808879 | 0.2920317 | TwoSample_MR | MDD | S_LDL_PL_pct |
| MR Egger | 108 | -0.0235492 | 0.0910571 | 0.7964299 | TwoSample_MR | MDD | S_LDL_TG |
| Weighted median | 108 | 0.0835167 | 0.0247833 | 0.000752 | TwoSample_MR | MDD | S_LDL_TG |
| Inverse variance weighted | 108 | 0.0933573 | 0.0201398 | 0.0000036 | TwoSample_MR | MDD | S_LDL_TG |
| Simple mode | 108 | 0.0909734 | 0.0757482 | 0.2324044 | TwoSample_MR | MDD | S_LDL_TG |
| Weighted mode | 108 | 0.0848164 | 0.0748969 | 0.2599799 | TwoSample_MR | MDD | S_LDL_TG |
| MR Egger | 108 | 0.1121516 | 0.1029963 | 0.2786719 | TwoSample_MR | MDD | S_LDL_TG_pct |
| Weighted median | 108 | 0.1172553 | 0.0252716 | 0.0000035 | TwoSample_MR | MDD | S_LDL_TG_pct |
| Inverse variance weighted | 108 | 0.1111269 | 0.022595 | 0.0000009 | TwoSample_MR | MDD | S_LDL_TG_pct |
| Simple mode | 108 | 0.1406928 | 0.06535 | 0.033572 | TwoSample_MR | MDD | S_LDL_TG_pct |
| Weighted mode | 108 | 0.1406928 | 0.0624199 | 0.0262356 | TwoSample_MR | MDD | S_LDL_TG_pct |
| MR Egger | 108 | -0.1325769 | 0.09821 | 0.1799132 | TwoSample_MR | MDD | S_VLDL_C_pct |
| Weighted median | 108 | -0.1161921 | 0.0236767 | 0.0000009 | TwoSample_MR | MDD | S_VLDL_C_pct |
| Inverse variance weighted | 108 | -0.0962925 | 0.0215567 | 0.0000079 | TwoSample_MR | MDD | S_VLDL_C_pct |
| Simple mode | 108 | -0.116706 | 0.0641988 | 0.0718797 | TwoSample_MR | MDD | S_VLDL_C_pct |
| Weighted mode | 108 | -0.1321955 | 0.0554334 | 0.01885 | TwoSample_MR | MDD | S_VLDL_C_pct |
| MR Egger | 108 | -0.0969671 | 0.1043085 | 0.3546809 | TwoSample_MR | MDD | S_VLDL_FC_pct |
| Weighted median | 108 | -0.1237079 | 0.0233128 | 0.0000001 | TwoSample_MR | MDD | S_VLDL_FC_pct |
| Inverse variance weighted | 108 | -0.1046791 | 0.0228809 | 0.0000048 | TwoSample_MR | MDD | S_VLDL_FC_pct |
| Simple mode | 108 | -0.1438262 | 0.0646844 | 0.0282834 | TwoSample_MR | MDD | S_VLDL_FC_pct |
| Weighted mode | 108 | -0.1405416 | 0.0570458 | 0.0153458 | TwoSample_MR | MDD | S_VLDL_FC_pct |
| MR Egger | 108 | -0.0827061 | 0.0848566 | 0.3319488 | TwoSample_MR | MDD | S_VLDL_L |
| Weighted median | 108 | 0.0377801 | 0.0247268 | 0.126537 | TwoSample_MR | MDD | S_VLDL_L |
| Inverse variance weighted | 108 | 0.0573384 | 0.0188636 | 0.0023686 | TwoSample_MR | MDD | S_VLDL_L |
| Simple mode | 108 | 0.0272315 | 0.0678233 | 0.688848 | TwoSample_MR | MDD | S_VLDL_L |
| Weighted mode | 108 | 0.0189435 | 0.073028 | 0.7958248 | TwoSample_MR | MDD | S_VLDL_L |
| MR Egger | 108 | -0.0811725 | 0.0844834 | 0.3388338 | TwoSample_MR | MDD | S_VLDL_P |
| Weighted median | 108 | 0.042411 | 0.0241214 | 0.0787081 | TwoSample_MR | MDD | S_VLDL_P |
| Inverse variance weighted | 108 | 0.0608646 | 0.01879 | 0.0011987 | TwoSample_MR | MDD | S_VLDL_P |
| Simple mode | 108 | 0.0217626 | 0.0671997 | 0.7466841 | TwoSample_MR | MDD | S_VLDL_P |
| Weighted mode | 108 | 0.010679 | 0.0665357 | 0.87279 | TwoSample_MR | MDD | S_VLDL_P |

| | | | | | | | |
|---|---|---|---|---|---|---|---|
| MR Egger | 108 | -0.0945186 | 0.1029487 | 0.3606438 | TwoSample_MR | MDD | S_VLDL_PL_pct |
| Weighted median | 108 | -0.1205443 | 0.0246074 | 0.000001 | TwoSample_MR | MDD | S_VLDL_PL_pct |
| Inverse variance weighted | 108 | -0.1013302 | 0.0225823 | 0.0000072 | TwoSample_MR | MDD | S_VLDL_PL_pct |
| Simple mode | 108 | -0.1381097 | 0.0620833 | 0.0282083 | TwoSample_MR | MDD | S_VLDL_PL_pct |
| Weighted mode | 108 | -0.1381097 | 0.0572754 | 0.0175979 | TwoSample_MR | MDD | S_VLDL_PL_pct |
| MR Egger | 108 | -0.0109853 | 0.0947317 | 0.907902 | TwoSample_MR | MDD | S_VLDL_TG |
| Weighted median | 108 | 0.0632181 | 0.0252607 | 0.0123275 | TwoSample_MR | MDD | S_VLDL_TG |
| Inverse variance weighted | 108 | 0.0942995 | 0.0209085 | 0.0000065 | TwoSample_MR | MDD | S_VLDL_TG |
| Simple mode | 108 | 0.0606293 | 0.0734189 | 0.4107555 | TwoSample_MR | MDD | S_VLDL_TG |
| Weighted mode | 108 | 0.0447095 | 0.0730159 | 0.5416214 | TwoSample_MR | MDD | S_VLDL_TG |
| MR Egger | 108 | 0.1228652 | 0.0999325 | 0.2216132 | TwoSample_MR | MDD | S_VLDL_TG_pct |
| Weighted median | 108 | 0.1126045 | 0.0242182 | 0.0000033 | TwoSample_MR | MDD | S_VLDL_TG_pct |
| Inverse variance weighted | 108 | 0.0994497 | 0.0219262 | 0.0000057 | TwoSample_MR | MDD | S_VLDL_TG_pct |
| Simple mode | 108 | 0.1215705 | 0.0602978 | 0.0462882 | TwoSample_MR | MDD | S_VLDL_TG_pct |
| Weighted mode | 108 | 0.1308776 | 0.054461 | 0.0179755 | TwoSample_MR | MDD | S_VLDL_TG_pct |
| MR Egger | 108 | -0.0301046 | 0.0972402 | 0.7574805 | TwoSample_MR | MDD | Sphingomyelins |
| Weighted median | 108 | -0.0215488 | 0.0239087 | 0.3674317 | TwoSample_MR | MDD | Sphingomyelins |
| Inverse variance weighted | 108 | -0.0391216 | 0.0213279 | 0.0666114 | TwoSample_MR | MDD | Sphingomyelins |
| Simple mode | 108 | -0.0392064 | 0.064211 | 0.542768 | TwoSample_MR | MDD | Sphingomyelins |
| Weighted mode | 108 | -0.0352774 | 0.0650357 | 0.5886494 | TwoSample_MR | MDD | Sphingomyelins |
| MR Egger | 108 | -0.1098953 | 0.0949893 | 0.2499041 | TwoSample_MR | MDD | TG_by_PG |
| Weighted median | 108 | 0.0872591 | 0.0235867 | 0.000216 | TwoSample_MR | MDD | TG_by_PG |
| Inverse variance weighted | 108 | 0.0967149 | 0.0213234 | 0.0000057 | TwoSample_MR | MDD | TG_by_PG |
| Simple mode | 108 | 0.1490407 | 0.0636666 | 0.0210874 | TwoSample_MR | MDD | TG_by_PG |
| Weighted mode | 108 | 0.126478 | 0.058155 | 0.0318438 | TwoSample_MR | MDD | TG_by_PG |
| MR Egger | 108 | -0.034466 | 0.089382 | 0.7005629 | TwoSample_MR | MDD | Total_CE |
| Weighted median | 108 | -0.0392492 | 0.0247825 | 0.1132515 | TwoSample_MR | MDD | Total_CE |
| Inverse variance weighted | 108 | -0.0366045 | 0.0196043 | 0.0618783 | TwoSample_MR | MDD | Total_CE |
| Simple mode | 108 | -0.0788884 | 0.077936 | 0.3137165 | TwoSample_MR | MDD | Total_CE |
| Weighted mode | 108 | -0.0701567 | 0.0817044 | 0.3924444 | TwoSample_MR | MDD | Total_CE |
| MR Egger | 108 | 0.1193979 | 0.0939162 | 0.2063964 | TwoSample_MR | MDD | Total_P |
| Weighted median | 108 | 0.0014931 | 0.0240262 | 0.9504474 | TwoSample_MR | MDD | Total_P |
| Inverse variance weighted | 108 | -0.0016036 | 0.0207662 | 0.9384478 | TwoSample_MR | MDD | Total_P |
| Simple mode | 108 | 0.0086752 | 0.0838194 | 0.9177613 | TwoSample_MR | MDD | Total_P |
| Weighted mode | 108 | 0.014436 | 0.0838168 | 0.8635794 | TwoSample_MR | MDD | Total_P |
| MR Egger | 108 | -0.0642302 | 0.0961866 | 0.5057338 | TwoSample_MR | MDD | Total_TG |
| Weighted median | 108 | 0.08367 | 0.0248925 | 0.0007759 | TwoSample_MR | MDD | Total_TG |
| Inverse variance weighted | 108 | 0.0937994 | 0.0213834 | 0.0000115 | TwoSample_MR | MDD | Total_TG |
| Simple mode | 108 | 0.1003354 | 0.0777913 | 0.1999003 | TwoSample_MR | MDD | Total_TG |
| Weighted mode | 108 | 0.0939459 | 0.0758893 | 0.2184504 | TwoSample_MR | MDD | Total_TG |
| MR Egger | 108 | 0.0473825 | 0.1763137 | 0.7886523 | TwoSample_MR | MDD | Unsaturation |
| Weighted median | 108 | -0.0850081 | 0.0251523 | 0.0007256 | TwoSample_MR | MDD | Unsaturation |
| Inverse variance weighted | 108 | -0.1093329 | 0.0388172 | 0.0048534 | TwoSample_MR | MDD | Unsaturation |
| Simple mode | 108 | -0.1394672 | 0.0714158 | 0.0534445 | TwoSample_MR | MDD | Unsaturation |
| Weighted mode | 108 | -0.1394672 | 0.0663919 | 0.0380196 | TwoSample_MR | MDD | Unsaturation |
| MR Egger | 108 | -0.1213888 | 0.0845281 | 0.1539254 | TwoSample_MR | MDD | VLDL_FC |

| | | | | | | | |
|---|---|---|---|---|---|---|---|
| Weighted median | 108 | 0.040934 | 0.0253536 | 0.1064136 | TwoSample_MR | MDD | VLDL_FC |
| Inverse variance weighted | 108 | 0.0493852 | 0.0189138 | 0.0090262 | TwoSample_MR | MDD | VLDL_FC |
| Simple mode | 108 | 0.0288359 | 0.0653654 | 0.6599952 | TwoSample_MR | MDD | VLDL_FC |
| Weighted mode | 108 | 0.0288359 | 0.0671217 | 0.6683461 | TwoSample_MR | MDD | VLDL_FC |
| MR Egger | 108 | -0.1060687 | 0.0900116 | 0.2412802 | TwoSample_MR | MDD | VLDL_L |
| Weighted median | 108 | 0.0615789 | 0.024222 | 0.0110134 | TwoSample_MR | MDD | VLDL_L |
| Inverse variance weighted | 108 | 0.0736427 | 0.0201335 | 0.0002545 | TwoSample_MR | MDD | VLDL_L |
| Simple mode | 108 | 0.0831542 | 0.0698827 | 0.2367148 | TwoSample_MR | MDD | VLDL_L |
| Weighted mode | 108 | 0.0772887 | 0.0667487 | 0.2494811 | TwoSample_MR | MDD | VLDL_L |
| MR Egger | 108 | -0.1112465 | 0.0865755 | 0.2016052 | TwoSample_MR | MDD | VLDL_PL |
| Weighted median | 108 | 0.0528203 | 0.0240844 | 0.0282979 | TwoSample_MR | MDD | VLDL_PL |
| Inverse variance weighted | 108 | 0.0625069 | 0.0193676 | 0.0012492 | TwoSample_MR | MDD | VLDL_PL |
| Simple mode | 108 | 0.0633802 | 0.0705836 | 0.3712315 | TwoSample_MR | MDD | VLDL_PL |
| Weighted mode | 108 | 0.057646 | 0.06715 | 0.3925535 | TwoSample_MR | MDD | VLDL_PL |
| MR Egger | 108 | -0.0862813 | 0.0957993 | 0.3698181 | TwoSample_MR | MDD | VLDL_TG |
| Weighted median | 108 | 0.0875126 | 0.024496 | 0.0003536 | TwoSample_MR | MDD | VLDL_TG |
| Inverse variance weighted | 108 | 0.0945365 | 0.021387 | 0.0000099 | TwoSample_MR | MDD | VLDL_TG |
| Simple mode | 108 | 0.1019205 | 0.0690151 | 0.1426706 | TwoSample_MR | MDD | VLDL_TG |
| Weighted mode | 108 | 0.0894501 | 0.0694775 | 0.2007088 | TwoSample_MR | MDD | VLDL_TG |
| MR Egger | 108 | -0.097012 | 0.0951045 | 0.3100241 | TwoSample_MR | MDD | VLDL_size |
| Weighted median | 108 | 0.0655982 | 0.0236466 | 0.0055354 | TwoSample_MR | MDD | VLDL_size |
| Inverse variance weighted | 108 | 0.1008164 | 0.0213106 | 0.0000022 | TwoSample_MR | MDD | VLDL_size |
| Simple mode | 108 | 0.0527944 | 0.061621 | 0.3934919 | TwoSample_MR | MDD | VLDL_size |
| Weighted mode | 108 | 0.0527944 | 0.0597079 | 0.3785639 | TwoSample_MR | MDD | VLDL_size |
| MR Egger | 108 | 0.0460789 | 0.1026475 | 0.6544173 | TwoSample_MR | MDD | XL_HDL_C |
| Weighted median | 108 | -0.0811916 | 0.0241303 | 0.0007662 | TwoSample_MR | MDD | XL_HDL_C |
| Inverse variance weighted | 108 | -0.1085324 | 0.0227649 | 0.0000019 | TwoSample_MR | MDD | XL_HDL_C |
| Simple mode | 108 | -0.0038446 | 0.0732127 | 0.9582182 | TwoSample_MR | MDD | XL_HDL_C |
| Weighted mode | 108 | -0.0108784 | 0.0670774 | 0.8714728 | TwoSample_MR | MDD | XL_HDL_C |
| MR Egger | 108 | 0.0671016 | 0.1018949 | 0.5116206 | TwoSample_MR | MDD | XL_HDL_CE |
| Weighted median | 108 | -0.0906519 | 0.0239799 | 0.0001566 | TwoSample_MR | MDD | XL_HDL_CE |
| Inverse variance weighted | 108 | -0.1048883 | 0.0226614 | 0.0000037 | TwoSample_MR | MDD | XL_HDL_CE |
| Simple mode | 108 | 0.001317 | 0.0719617 | 0.9854321 | TwoSample_MR | MDD | XL_HDL_CE |
| Weighted mode | 108 | 0.001317 | 0.0677903 | 0.9845358 | TwoSample_MR | MDD | XL_HDL_CE |
| MR Egger | 108 | -0.0307203 | 0.1042087 | 0.768726 | TwoSample_MR | MDD | XL_HDL_FC |
| Weighted median | 108 | -0.0942132 | 0.0249696 | 0.0001612 | TwoSample_MR | MDD | XL_HDL_FC |
| Inverse variance weighted | 108 | -0.114635 | 0.022929 | 0.0000006 | TwoSample_MR | MDD | XL_HDL_FC |
| Simple mode | 108 | -0.0157798 | 0.0760959 | 0.8361173 | TwoSample_MR | MDD | XL_HDL_FC |
| Weighted mode | 108 | -0.0195793 | 0.0734011 | 0.7901802 | TwoSample_MR | MDD | XL_HDL_FC |
| MR Egger | 108 | -0.1882638 | 0.0895752 | 0.0379458 | TwoSample_MR | MDD | XL_HDL_FC_pct |
| Weighted median | 108 | 0.0203043 | 0.0226748 | 0.3705437 | TwoSample_MR | MDD | XL_HDL_FC_pct |
| Inverse variance weighted | 108 | 0.0588756 | 0.020373 | 0.0038539 | TwoSample_MR | MDD | XL_HDL_FC_pct |
| Simple mode | 108 | -0.0144502 | 0.075584 | 0.848746 | TwoSample_MR | MDD | XL_HDL_FC_pct |
| Weighted mode | 108 | -0.0116501 | 0.0705143 | 0.8690859 | TwoSample_MR | MDD | XL_HDL_FC_pct |
| MR Egger | 108 | 0.0621785 | 0.0992543 | 0.5323623 | TwoSample_MR | MDD | XL_HDL_L |
| Weighted median | 108 | -0.0736329 | 0.0240025 | 0.002157 | TwoSample_MR | MDD | XL_HDL_L |

| | | | | | | | |
|---|---|---|---|---|---|---|---|
| Inverse variance weighted | 108 | -0.0993926 | 0.0220523 | 0.0000066 | TwoSample_MR | MDD | XL_HDL_L |
| Simple mode | 108 | 0.0045406 | 0.0738193 | 0.9510684 | TwoSample_MR | MDD | XL_HDL_L |
| Weighted mode | 108 | 0.0045406 | 0.0715059 | 0.9494875 | TwoSample_MR | MDD | XL_HDL_L |
| MR Egger | 108 | 0.0530931 | 0.0986735 | 0.5916572 | TwoSample_MR | MDD | XL_HDL_P |
| Weighted median | 108 | -0.0682041 | 0.0237141 | 0.0040263 | TwoSample_MR | MDD | XL_HDL_P |
| Inverse variance weighted | 108 | -0.089919 | 0.0218648 | 0.0000391 | TwoSample_MR | MDD | XL_HDL_P |
| Simple mode | 108 | -0.0028275 | 0.0794568 | 0.9716788 | TwoSample_MR | MDD | XL_HDL_P |
| Weighted mode | 108 | 0.0077108 | 0.0777911 | 0.9212268 | TwoSample_MR | MDD | XL_HDL_P |
| MR Egger | 108 | 0.0741992 | 0.0973497 | 0.4476379 | TwoSample_MR | MDD | XL_HDL_PL |
| Weighted median | 108 | -0.0662213 | 0.0246644 | 0.0072552 | TwoSample_MR | MDD | XL_HDL_PL |
| Inverse variance weighted | 108 | -0.0963752 | 0.0216729 | 0.0000087 | TwoSample_MR | MDD | XL_HDL_PL |
| Simple mode | 108 | 0.0054136 | 0.074356 | 0.942096 | TwoSample_MR | MDD | XL_HDL_PL |
| Weighted mode | 108 | 0.0087596 | 0.0674177 | 0.8968655 | TwoSample_MR | MDD | XL_HDL_PL |
| MR Egger | 108 | -0.0381645 | 0.09999 | 0.7034609 | TwoSample_MR | MDD | XL_HDL_TG_pct |
| Weighted median | 108 | 0.0719727 | 0.0251728 | 0.0042479 | TwoSample_MR | MDD | XL_HDL_TG_pct |
| Inverse variance weighted | 108 | 0.1154466 | 0.0221886 | 0.0000002 | TwoSample_MR | MDD | XL_HDL_TG_pct |
| Simple mode | 108 | 0.0298359 | 0.0711392 | 0.6757638 | TwoSample_MR | MDD | XL_HDL_TG_pct |
| Weighted mode | 108 | 0.0477081 | 0.0660995 | 0.4720146 | TwoSample_MR | MDD | XL_HDL_TG_pct |
| MR Egger | 108 | -0.1247457 | 0.0885253 | 0.1617168 | TwoSample_MR | MDD | XL_VLDL_C |
| Weighted median | 108 | 0.0691438 | 0.0244516 | 0.0046872 | TwoSample_MR | MDD | XL_VLDL_C |
| Inverse variance weighted | 108 | 0.0734942 | 0.0198977 | 0.0002211 | TwoSample_MR | MDD | XL_VLDL_C |
| Simple mode | 108 | 0.1022705 | 0.0657852 | 0.1229911 | TwoSample_MR | MDD | XL_VLDL_C |
| Weighted mode | 108 | 0.0993496 | 0.0621472 | 0.1128546 | TwoSample_MR | MDD | XL_VLDL_C |
| MR Egger | 108 | -0.1304235 | 0.0847624 | 0.1268585 | TwoSample_MR | MDD | XL_VLDL_CE |
| Weighted median | 108 | 0.0486075 | 0.024507 | 0.0473215 | TwoSample_MR | MDD | XL_VLDL_CE |
| Inverse variance weighted | 108 | 0.0612329 | 0.0190595 | 0.0013148 | TwoSample_MR | MDD | XL_VLDL_CE |
| Simple mode | 108 | 0.0526522 | 0.0606574 | 0.3873224 | TwoSample_MR | MDD | XL_VLDL_CE |
| Weighted mode | 108 | 0.0498703 | 0.061765 | 0.4212154 | TwoSample_MR | MDD | XL_VLDL_CE |
| MR Egger | 108 | 0.0230698 | 0.1010813 | 0.8199072 | TwoSample_MR | MDD | XL_VLDL_CE_pct |
| Weighted median | 108 | -0.0855639 | 0.0254442 | 0.0007715 | TwoSample_MR | MDD | XL_VLDL_CE_pct |
| Inverse variance weighted | 108 | -0.0936713 | 0.0223228 | 0.0000271 | TwoSample_MR | MDD | XL_VLDL_CE_pct |
| Simple mode | 108 | -0.0651047 | 0.0645768 | 0.3156448 | TwoSample_MR | MDD | XL_VLDL_CE_pct |
| Weighted mode | 108 | -0.075279 | 0.0541414 | 0.1672883 | TwoSample_MR | MDD | XL_VLDL_CE_pct |
| MR Egger | 108 | 0.0291698 | 0.1022413 | 0.7759687 | TwoSample_MR | MDD | XL_VLDL_C_pct |
| Weighted median | 108 | -0.0926865 | 0.0246224 | 0.000167 | TwoSample_MR | MDD | XL_VLDL_C_pct |
| Inverse variance weighted | 108 | -0.0958011 | 0.0225959 | 0.0000224 | TwoSample_MR | MDD | XL_VLDL_C_pct |
| Simple mode | 108 | -0.0733215 | 0.0643811 | 0.2573023 | TwoSample_MR | MDD | XL_VLDL_C_pct |
| Weighted mode | 108 | -0.0835576 | 0.0515168 | 0.1077573 | TwoSample_MR | MDD | XL_VLDL_C_pct |
| MR Egger | 108 | -0.1132111 | 0.0918822 | 0.2206258 | TwoSample_MR | MDD | XL_VLDL_FC |
| Weighted median | 108 | 0.0742946 | 0.0244183 | 0.0023456 | TwoSample_MR | MDD | XL_VLDL_FC |
| Inverse variance weighted | 108 | 0.0831878 | 0.0206101 | 0.0000543 | TwoSample_MR | MDD | XL_VLDL_FC |
| Simple mode | 108 | 0.1321275 | 0.0674853 | 0.0528489 | TwoSample_MR | MDD | XL_VLDL_FC |
| Weighted mode | 108 | 0.1195963 | 0.0654941 | 0.0706296 | TwoSample_MR | MDD | XL_VLDL_FC |
| MR Egger | 108 | 0.0403612 | 0.1023857 | 0.6942203 | TwoSample_MR | MDD | XL_VLDL_FC_pct |
| Weighted median | 108 | -0.0747353 | 0.025034 | 0.0028325 | TwoSample_MR | MDD | XL_VLDL_FC_pct |
| Inverse variance weighted | 108 | -0.0893022 | 0.0226345 | 0.0000797 | TwoSample_MR | MDD | XL_VLDL_FC_pct |

| | | | | | | | |
|---|---|---|---|---|---|---|---|
| Simple mode | 108 | -0.1217942 | 0.0680764 | 0.0764311 | TwoSample_MR | MDD | XL_VLDL_FC_pct |
| Weighted mode | 108 | -0.1075447 | 0.0600417 | 0.0760937 | TwoSample_MR | MDD | XL_VLDL_FC_pct |
| MR Egger | 108 | -0.1109747 | 0.095329 | 0.2469875 | TwoSample_MR | MDD | XL_VLDL_L |
| Weighted median | 108 | 0.0886957 | 0.0245143 | 0.0002968 | TwoSample_MR | MDD | XL_VLDL_L |
| Inverse variance weighted | 108 | 0.0928537 | 0.0213843 | 0.0000141 | TwoSample_MR | MDD | XL_VLDL_L |
| Simple mode | 108 | 0.1457912 | 0.070862 | 0.0420793 | TwoSample_MR | MDD | XL_VLDL_L |
| Weighted mode | 108 | 0.130229 | 0.0672 | 0.055267 | TwoSample_MR | MDD | XL_VLDL_L |
| MR Egger | 108 | -0.1046125 | 0.0948724 | 0.2726699 | TwoSample_MR | MDD | XL_VLDL_P |
| Weighted median | 108 | 0.0783325 | 0.0251671 | 0.0018551 | TwoSample_MR | MDD | XL_VLDL_P |
| Inverse variance weighted | 108 | 0.0920552 | 0.0212537 | 0.0000148 | TwoSample_MR | MDD | XL_VLDL_P |
| Simple mode | 108 | 0.1424983 | 0.0695573 | 0.0429441 | TwoSample_MR | MDD | XL_VLDL_P |
| Weighted mode | 108 | 0.1173755 | 0.0653502 | 0.0753006 | TwoSample_MR | MDD | XL_VLDL_P |
| MR Egger | 108 | -0.116293 | 0.0935171 | 0.2164093 | TwoSample_MR | MDD | XL_VLDL_PL |
| Weighted median | 108 | 0.0758504 | 0.0240975 | 0.0016459 | TwoSample_MR | MDD | XL_VLDL_PL |
| Inverse variance weighted | 108 | 0.0884453 | 0.0209995 | 0.0000253 | TwoSample_MR | MDD | XL_VLDL_PL |
| Simple mode | 108 | 0.1416071 | 0.0683361 | 0.040649 | TwoSample_MR | MDD | XL_VLDL_PL |
| Weighted mode | 108 | 0.12604 | 0.0669517 | 0.0624779 | TwoSample_MR | MDD | XL_VLDL_PL |
| MR Egger | 108 | -0.1056947 | 0.0983072 | 0.2847498 | TwoSample_MR | MDD | XL_VLDL_TG |
| Weighted median | 108 | 0.1053273 | 0.0255381 | 0.0000372 | TwoSample_MR | MDD | XL_VLDL_TG |
| Inverse variance weighted | 108 | 0.0992173 | 0.0220296 | 0.0000067 | TwoSample_MR | MDD | XL_VLDL_TG |
| Simple mode | 108 | 0.1576927 | 0.0743293 | 0.0361852 | TwoSample_MR | MDD | XL_VLDL_TG |
| Weighted mode | 108 | 0.1385824 | 0.0653712 | 0.0363239 | TwoSample_MR | MDD | XL_VLDL_TG |
| MR Egger | 108 | -0.0068398 | 0.0996471 | 0.9454054 | TwoSample_MR | MDD | XL_VLDL_TG_pct |
| Weighted median | 108 | 0.0953845 | 0.0240521 | 0.0000732 | TwoSample_MR | MDD | XL_VLDL_TG_pct |
| Inverse variance weighted | 108 | 0.0884344 | 0.0219573 | 0.0000564 | TwoSample_MR | MDD | XL_VLDL_TG_pct |
| Simple mode | 108 | 0.0889529 | 0.0628171 | 0.1596616 | TwoSample_MR | MDD | XL_VLDL_TG_pct |
| Weighted mode | 108 | 0.0954921 | 0.054305 | 0.081532 | TwoSample_MR | MDD | XL_VLDL_TG_pct |
| MR Egger | 108 | -0.0910129 | 0.1013396 | 0.3711687 | TwoSample_MR | MDD | XS_VLDL_CE_pct |
| Weighted median | 108 | -0.1036245 | 0.0236245 | 0.0000115 | TwoSample_MR | MDD | XS_VLDL_CE_pct |
| Inverse variance weighted | 108 | -0.1228486 | 0.0222401 | 0 | TwoSample_MR | MDD | XS_VLDL_CE_pct |
| Simple mode | 108 | -0.1136037 | 0.0663872 | 0.0899363 | TwoSample_MR | MDD | XS_VLDL_CE_pct |
| Weighted mode | 108 | -0.1136037 | 0.0662236 | 0.089156 | TwoSample_MR | MDD | XS_VLDL_CE_pct |
| MR Egger | 108 | -0.1108525 | 0.1032793 | 0.2855617 | TwoSample_MR | MDD | XS_VLDL_C_pct |
| Weighted median | 108 | -0.1153357 | 0.0238168 | 0.0000013 | TwoSample_MR | MDD | XS_VLDL_C_pct |
| Inverse variance weighted | 108 | -0.1240725 | 0.0226566 | 0 | TwoSample_MR | MDD | XS_VLDL_C_pct |
| Simple mode | 108 | -0.133772 | 0.0633004 | 0.0369009 | TwoSample_MR | MDD | XS_VLDL_C_pct |
| Weighted mode | 108 | -0.133772 | 0.0610954 | 0.0307282 | TwoSample_MR | MDD | XS_VLDL_C_pct |
| MR Egger | 108 | -0.200275 | 0.1035628 | 0.055798 | TwoSample_MR | MDD | XS_VLDL_FC_pct |
| Weighted median | 108 | -0.0982301 | 0.0249266 | 0.0000812 | TwoSample_MR | MDD | XS_VLDL_FC_pct |
| Inverse variance weighted | 108 | -0.0960376 | 0.022828 | 0.0000259 | TwoSample_MR | MDD | XS_VLDL_FC_pct |
| Simple mode | 108 | -0.1409695 | 0.0656973 | 0.0341543 | TwoSample_MR | MDD | XS_VLDL_FC_pct |
| Weighted mode | 108 | -0.1409695 | 0.0646024 | 0.0312886 | TwoSample_MR | MDD | XS_VLDL_FC_pct |
| MR Egger | 108 | -0.0488634 | 0.0824945 | 0.5548955 | TwoSample_MR | MDD | XS_VLDL_PL_pct |
| Weighted median | 108 | 0.0619305 | 0.024721 | 0.0122389 | TwoSample_MR | MDD | XS_VLDL_PL_pct |
| Inverse variance weighted | 108 | 0.0727944 | 0.0182868 | 0.0000687 | TwoSample_MR | MDD | XS_VLDL_PL_pct |
| Simple mode | 108 | 0.0512566 | 0.0637991 | 0.423521 | TwoSample_MR | MDD | XS_VLDL_PL_pct |

| | | | | | | | |
|---|---|---|---|---|---|---|---|
| Weighted mode | 108 | 0.0640704 | 0.0615822 | 0.3004965 | TwoSample_MR | MDD | XS_VLDL_PL_pct |
| MR Egger | 108 | 0.0183218 | 0.0910906 | 0.8409757 | TwoSample_MR | MDD | XS_VLDL_TG |
| Weighted median | 108 | 0.0879937 | 0.0246966 | 0.0003667 | TwoSample_MR | MDD | XS_VLDL_TG |
| Inverse variance weighted | 108 | 0.085075 | 0.0200334 | 0.0000217 | TwoSample_MR | MDD | XS_VLDL_TG |
| Simple mode | 108 | 0.0998697 | 0.0735046 | 0.1771033 | TwoSample_MR | MDD | XS_VLDL_TG |
| Weighted mode | 108 | 0.0941448 | 0.0733385 | 0.2020192 | TwoSample_MR | MDD | XS_VLDL_TG |
| MR Egger | 108 | 0.1317897 | 0.1051832 | 0.2129793 | TwoSample_MR | MDD | XS_VLDL_TG_pct |
| Weighted median | 108 | 0.1231057 | 0.0247354 | 0.0000006 | TwoSample_MR | MDD | XS_VLDL_TG_pct |
| Inverse variance weighted | 108 | 0.1235534 | 0.0230734 | 0.0000001 | TwoSample_MR | MDD | XS_VLDL_TG_pct |
| Simple mode | 108 | 0.1666903 | 0.0673644 | 0.0149146 | TwoSample_MR | MDD | XS_VLDL_TG_pct |
| Weighted mode | 108 | 0.1701994 | 0.0686529 | 0.0147313 | TwoSample_MR | MDD | XS_VLDL_TG_pct |
| MR Egger | 108 | -0.0328402 | 0.0983754 | 0.7391711 | TwoSample_MR | MDD | XXL_VLDL_C |
| Weighted median | 108 | 0.0996144 | 0.0248239 | 0.00006 | TwoSample_MR | MDD | XXL_VLDL_C |
| Inverse variance weighted | 108 | 0.1024825 | 0.0217867 | 0.0000026 | TwoSample_MR | MDD | XXL_VLDL_C |
| Simple mode | 108 | 0.1578329 | 0.072162 | 0.0309044 | TwoSample_MR | MDD | XXL_VLDL_C |
| Weighted mode | 108 | 0.1517247 | 0.0728258 | 0.0395981 | TwoSample_MR | MDD | XXL_VLDL_C |
| MR Egger | 108 | -0.0420691 | 0.0979528 | 0.6684432 | TwoSample_MR | MDD | XXL_VLDL_CE |
| Weighted median | 108 | 0.106221 | 0.0242665 | 0.000012 | TwoSample_MR | MDD | XXL_VLDL_CE |
| Inverse variance weighted | 108 | 0.0998791 | 0.0217141 | 0.0000042 | TwoSample_MR | MDD | XXL_VLDL_CE |
| Simple mode | 108 | 0.1474082 | 0.072805 | 0.0453888 | TwoSample_MR | MDD | XXL_VLDL_CE |
| Weighted mode | 108 | 0.138308 | 0.0707569 | 0.0532284 | TwoSample_MR | MDD | XXL_VLDL_CE |
| MR Egger | 108 | -0.0292412 | 0.0985706 | 0.7673126 | TwoSample_MR | MDD | XXL_VLDL_FC |
| Weighted median | 108 | 0.0904131 | 0.0246836 | 0.0002494 | TwoSample_MR | MDD | XXL_VLDL_FC |
| Inverse variance weighted | 108 | 0.1029572 | 0.0218206 | 0.0000024 | TwoSample_MR | MDD | XXL_VLDL_FC |
| Simple mode | 108 | 0.1570263 | 0.0709542 | 0.0290165 | TwoSample_MR | MDD | XXL_VLDL_FC |
| Weighted mode | 108 | 0.1507519 | 0.0685471 | 0.0300123 | TwoSample_MR | MDD | XXL_VLDL_FC |
| MR Egger | 108 | -0.0687228 | 0.0979677 | 0.4845383 | TwoSample_MR | MDD | XXL_VLDL_L |
| Weighted median | 108 | 0.1121153 | 0.0245612 | 0.000005 | TwoSample_MR | MDD | XXL_VLDL_L |
| Inverse variance weighted | 108 | 0.0989111 | 0.0218076 | 0.0000057 | TwoSample_MR | MDD | XXL_VLDL_L |
| Simple mode | 108 | 0.155625 | 0.0681667 | 0.0244063 | TwoSample_MR | MDD | XXL_VLDL_L |
| Weighted mode | 108 | 0.155625 | 0.0649786 | 0.0183575 | TwoSample_MR | MDD | XXL_VLDL_L |
| MR Egger | 108 | -0.0550746 | 0.0978957 | 0.5749057 | TwoSample_MR | MDD | XXL_VLDL_P |
| Weighted median | 108 | 0.1019024 | 0.0246549 | 0.0000358 | TwoSample_MR | MDD | XXL_VLDL_P |
| Inverse variance weighted | 108 | 0.101442 | 0.0217515 | 0.0000031 | TwoSample_MR | MDD | XXL_VLDL_P |
| Simple mode | 108 | 0.1637175 | 0.0713674 | 0.023743 | TwoSample_MR | MDD | XXL_VLDL_P |
| Weighted mode | 108 | 0.1606548 | 0.0724977 | 0.0288089 | TwoSample_MR | MDD | XXL_VLDL_P |
| MR Egger | 108 | -0.0399942 | 0.0978113 | 0.6834444 | TwoSample_MR | MDD | XXL_VLDL_PL |
| Weighted median | 108 | 0.0896769 | 0.0244563 | 0.0002456 | TwoSample_MR | MDD | XXL_VLDL_PL |
| Inverse variance weighted | 108 | 0.1064028 | 0.0216995 | 0.0000009 | TwoSample_MR | MDD | XXL_VLDL_PL |
| Simple mode | 108 | 0.1560608 | 0.0679222 | 0.0235277 | TwoSample_MR | MDD | XXL_VLDL_PL |
| Weighted mode | 108 | 0.1530212 | 0.0699226 | 0.030812 | TwoSample_MR | MDD | XXL_VLDL_PL |
| MR Egger | 108 | 0.178329 | 0.0884501 | 0.0463135 | TwoSample_MR | MDD | XXL_VLDL_PL_pct |
| Weighted median | 108 | 0.0359582 | 0.025281 | 0.1549274 | TwoSample_MR | MDD | XXL_VLDL_PL_pct |
| Inverse variance weighted | 108 | 0.0606765 | 0.0195697 | 0.0019317 | TwoSample_MR | MDD | XXL_VLDL_PL_pct |
| Simple mode | 108 | -0.0547996 | 0.0741743 | 0.4616487 | TwoSample_MR | MDD | XXL_VLDL_PL_pct |
| Weighted mode | 108 | -0.0575055 | 0.0735714 | 0.4361588 | TwoSample_MR | MDD | XXL_VLDL_PL_pct |

| | | | | | | | |
|---|---|---|---|---|---|---|---|
| MR Egger | 108 | -0.0843522 | 0.0979446 | 0.3910587 | TwoSample_MR | MDD | XXL_VLDL_TG |
| Weighted median | 108 | 0.1011882 | 0.024636 | 0.00004 | TwoSample_MR | MDD | XXL_VLDL_TG |
| Inverse variance weighted | 108 | 0.0950513 | 0.0218481 | 0.0000136 | TwoSample_MR | MDD | XXL_VLDL_TG |
| Simple mode | 108 | 0.1302121 | 0.0653307 | 0.0487935 | TwoSample_MR | MDD | XXL_VLDL_TG |
| Weighted mode | 108 | 0.1362733 | 0.0670184 | 0.0444901 | TwoSample_MR | MDD | XXL_VLDL_TG |
| MR Egger | 112 | 0.0035335 | 0.0282478 | 0.9006821 | TwoSample_MR | M_HDL_CE | MDD |
| Weighted median | 112 | 0.0169073 | 0.0185203 | 0.361293 | TwoSample_MR | M_HDL_CE | MDD |
| Inverse variance weighted | 112 | 0.027633 | 0.0165354 | 0.094694 | TwoSample_MR | M_HDL_CE | MDD |
| Simple mode | 112 | -0.0106186 | 0.0304467 | 0.7279303 | TwoSample_MR | M_HDL_CE | MDD |
| Weighted mode | 112 | 0.0126273 | 0.0194481 | 0.5174958 | TwoSample_MR | M_HDL_CE | MDD |
| MR Egger | 117 | -0.0136864 | 0.0216825 | 0.5291492 | TwoSample_MR | M_HDL_CE_pct | MDD |
| Weighted median | 117 | 0.0003032 | 0.0162096 | 0.9850779 | TwoSample_MR | M_HDL_CE_pct | MDD |
| Inverse variance weighted | 117 | -0.011638 | 0.0136917 | 0.3953248 | TwoSample_MR | M_HDL_CE_pct | MDD |
| Simple mode | 117 | -0.0110793 | 0.029334 | 0.7063488 | TwoSample_MR | M_HDL_CE_pct | MDD |
| Weighted mode | 117 | -0.0110793 | 0.0147552 | 0.4542517 | TwoSample_MR | M_HDL_CE_pct | MDD |
| MR Egger | 104 | 0.0005469 | 0.0271863 | 0.9839885 | TwoSample_MR | M_HDL_C | MDD |
| Weighted median | 104 | 0.0149782 | 0.0186233 | 0.421241 | TwoSample_MR | M_HDL_C | MDD |
| Inverse variance weighted | 104 | 0.0227144 | 0.0159981 | 0.1556622 | TwoSample_MR | M_HDL_C | MDD |
| Simple mode | 104 | -0.0101278 | 0.0323375 | 0.7547705 | TwoSample_MR | M_HDL_C | MDD |
| Weighted mode | 104 | 0.0124592 | 0.0201959 | 0.5386509 | TwoSample_MR | M_HDL_C | MDD |
| MR Egger | 116 | -0.0133333 | 0.0201345 | 0.5091722 | TwoSample_MR | M_HDL_C_pct | MDD |
| Weighted median | 116 | -0.0283735 | 0.0180541 | 0.1160484 | TwoSample_MR | M_HDL_C_pct | MDD |
| Inverse variance weighted | 116 | -0.0122743 | 0.0124935 | 0.3258776 | TwoSample_MR | M_HDL_C_pct | MDD |
| Simple mode | 116 | -0.027548 | 0.0323329 | 0.3959789 | TwoSample_MR | M_HDL_C_pct | MDD |
| Weighted mode | 116 | -0.0175472 | 0.0164994 | 0.2897815 | TwoSample_MR | M_HDL_C_pct | MDD |
| MR Egger | 97 | 0.0051733 | 0.027971 | 0.8536612 | TwoSample_MR | M_HDL_FC | MDD |
| Weighted median | 97 | 0.0049625 | 0.0186573 | 0.7902516 | TwoSample_MR | M_HDL_FC | MDD |
| Inverse variance weighted | 97 | 0.0131797 | 0.0163971 | 0.4215219 | TwoSample_MR | M_HDL_FC | MDD |
| Simple mode | 97 | -0.0064359 | 0.0318661 | 0.8403696 | TwoSample_MR | M_HDL_FC | MDD |
| Weighted mode | 97 | 0.0062744 | 0.0181311 | 0.7300593 | TwoSample_MR | M_HDL_FC | MDD |
| MR Egger | 112 | -0.0287081 | 0.0207522 | 0.1693523 | TwoSample_MR | M_HDL_FC_pct | MDD |
| Weighted median | 112 | -0.0204349 | 0.0174733 | 0.2422041 | TwoSample_MR | M_HDL_FC_pct | MDD |
| Inverse variance weighted | 112 | -0.0236948 | 0.0135043 | 0.0793256 | TwoSample_MR | M_HDL_FC_pct | MDD |
| Simple mode | 112 | -0.030728 | 0.0341465 | 0.370128 | TwoSample_MR | M_HDL_FC_pct | MDD |
| Weighted mode | 112 | -0.0208036 | 0.0165156 | 0.2104455 | TwoSample_MR | M_HDL_FC_pct | MDD |
| MR Egger | 122 | 0.0066767 | 0.0251029 | 0.7907158 | TwoSample_MR | M_HDL_PL_pct | MDD |
| Weighted median | 122 | 0.0156366 | 0.0182032 | 0.3903404 | TwoSample_MR | M_HDL_PL_pct | MDD |
| Inverse variance weighted | 122 | 0.0100798 | 0.0147312 | 0.4938208 | TwoSample_MR | M_HDL_PL_pct | MDD |
| Simple mode | 122 | 0.0353852 | 0.0390502 | 0.3666601 | TwoSample_MR | M_HDL_PL_pct | MDD |
| Weighted mode | 122 | 0.0165637 | 0.0208803 | 0.4291721 | TwoSample_MR | M_HDL_PL_pct | MDD |
| MR Egger | 97 | -0.0064347 | 0.027084 | 0.8127166 | TwoSample_MR | M_HDL_P | MDD |
| Weighted median | 97 | 0.0144595 | 0.0191758 | 0.450819 | TwoSample_MR | M_HDL_P | MDD |
| Inverse variance weighted | 97 | 0.0175448 | 0.0162292 | 0.2796699 | TwoSample_MR | M_HDL_P | MDD |
| Simple mode | 97 | -0.0085273 | 0.032427 | 0.7931391 | TwoSample_MR | M_HDL_P | MDD |
| Weighted mode | 97 | -0.0000955 | 0.0182919 | 0.9958458 | TwoSample_MR | M_HDL_P | MDD |
| MR Egger | 86 | 0.0061576 | 0.0176435 | 0.7279636 | TwoSample_MR | M_HDL_TG | MDD |

| | | | | | | | |
|---|---|---|---|---|---|---|---|
| Weighted median | 86 | 0.0130515 | 0.0153422 | 0.3949394 | TwoSample_MR | M_HDL_TG | MDD |
| Inverse variance weighted | 86 | 0.0225557 | 0.011854 | 0.0570679 | TwoSample_MR | M_HDL_TG | MDD |
| Simple mode | 86 | -0.0079949 | 0.0242662 | 0.7426152 | TwoSample_MR | M_HDL_TG | MDD |
| Weighted mode | 86 | 0.005443 | 0.013844 | 0.6951789 | TwoSample_MR | M_HDL_TG | MDD |
| MR Egger | 121 | 0.0204972 | 0.019479 | 0.2948059 | TwoSample_MR | M_HDL_TG_pct | MDD |
| Weighted median | 121 | 0.0183143 | 0.0153221 | 0.231976 | TwoSample_MR | M_HDL_TG_pct | MDD |
| Inverse variance weighted | 121 | 0.0130648 | 0.0128215 | 0.3082119 | TwoSample_MR | M_HDL_TG_pct | MDD |
| Simple mode | 121 | 0.0134439 | 0.0260031 | 0.6061 | TwoSample_MR | M_HDL_TG_pct | MDD |
| Weighted mode | 121 | 0.0134439 | 0.0131914 | 0.3101865 | TwoSample_MR | M_HDL_TG_pct | MDD |
| MR Egger | 102 | 0.003271 | 0.0212068 | 0.877729 | TwoSample_MR | M_LDL_TG | MDD |
| Weighted median | 102 | -0.0023547 | 0.0165723 | 0.8870128 | TwoSample_MR | M_LDL_TG | MDD |
| Inverse variance weighted | 102 | 0.0065387 | 0.0131263 | 0.618389 | TwoSample_MR | M_LDL_TG | MDD |
| Simple mode | 102 | -0.0000128 | 0.0294818 | 0.9996557 | TwoSample_MR | M_LDL_TG | MDD |
| Weighted mode | 102 | 0.0027142 | 0.0148597 | 0.855437 | TwoSample_MR | M_LDL_TG | MDD |
| MR Egger | 86 | 0.0140016 | 0.01442 | 0.3343418 | TwoSample_MR | M_LDL_TG_pct | MDD |
| Weighted median | 86 | 0.0088877 | 0.0147792 | 0.5475968 | TwoSample_MR | M_LDL_TG_pct | MDD |
| Inverse variance weighted | 86 | 0.0240219 | 0.0102961 | 0.0196432 | TwoSample_MR | M_LDL_TG_pct | MDD |
| Simple mode | 86 | 0.0178222 | 0.0297305 | 0.550462 | TwoSample_MR | M_LDL_TG_pct | MDD |
| Weighted mode | 86 | 0.0056889 | 0.0125045 | 0.6503083 | TwoSample_MR | M_LDL_TG_pct | MDD |
| MR Egger | 90 | 0.0137373 | 0.0237401 | 0.5643008 | TwoSample_MR | MUFA | MDD |
| Weighted median | 90 | 0.0279333 | 0.0180549 | 0.1218308 | TwoSample_MR | MUFA | MDD |
| Inverse variance weighted | 90 | 0.0372712 | 0.0144973 | 0.0101435 | TwoSample_MR | MUFA | MDD |
| Simple mode | 90 | 0.0111691 | 0.0330136 | 0.73592 | TwoSample_MR | MUFA | MDD |
| Weighted mode | 90 | 0.0199088 | 0.0180542 | 0.2731193 | TwoSample_MR | MUFA | MDD |
| MR Egger | 98 | 0.0386852 | 0.0206977 | 0.0646637 | TwoSample_MR | MUFA_pct | MDD |
| Weighted median | 98 | 0.0434932 | 0.0180042 | 0.0157037 | TwoSample_MR | MUFA_pct | MDD |
| Inverse variance weighted | 98 | 0.0442173 | 0.0134159 | 0.0009811 | TwoSample_MR | MUFA_pct | MDD |
| Simple mode | 98 | 0.0282811 | 0.0314183 | 0.3702699 | TwoSample_MR | MUFA_pct | MDD |
| Weighted mode | 98 | 0.047032 | 0.0164023 | 0.005077 | TwoSample_MR | MUFA_pct | MDD |
| MR Egger | 104 | -0.0292549 | 0.017953 | 0.1062861 | TwoSample_MR | M_VLDL_CE_pct | MDD |
| Weighted median | 104 | -0.02193 | 0.0155824 | 0.15932 | TwoSample_MR | M_VLDL_CE_pct | MDD |
| Inverse variance weighted | 104 | -0.0465791 | 0.0121939 | 0.0001335 | TwoSample_MR | M_VLDL_CE_pct | MDD |
| Simple mode | 104 | -0.0206317 | 0.0339374 | 0.5445703 | TwoSample_MR | M_VLDL_CE_pct | MDD |
| Weighted mode | 104 | -0.0234393 | 0.0142048 | 0.1019691 | TwoSample_MR | M_VLDL_CE_pct | MDD |
| MR Egger | 103 | -0.0353829 | 0.0184936 | 0.0585471 | TwoSample_MR | M_VLDL_C_pct | MDD |
| Weighted median | 103 | -0.0269461 | 0.0164143 | 0.1006687 | TwoSample_MR | M_VLDL_C_pct | MDD |
| Inverse variance weighted | 103 | -0.0494348 | 0.0124874 | 0.0000753 | TwoSample_MR | M_VLDL_C_pct | MDD |
| Simple mode | 103 | -0.0281186 | 0.0277938 | 0.3140818 | TwoSample_MR | M_VLDL_C_pct | MDD |
| Weighted mode | 103 | -0.0281186 | 0.0158788 | 0.0795765 | TwoSample_MR | M_VLDL_C_pct | MDD |
| MR Egger | 89 | -0.0363956 | 0.0203558 | 0.0772619 | TwoSample_MR | M_VLDL_FC_pct | MDD |
| Weighted median | 89 | -0.0328346 | 0.0173945 | 0.0590734 | TwoSample_MR | M_VLDL_FC_pct | MDD |
| Inverse variance weighted | 89 | -0.0525923 | 0.0135054 | 0.0000985 | TwoSample_MR | M_VLDL_FC_pct | MDD |
| Simple mode | 89 | -0.0225333 | 0.0259813 | 0.3881426 | TwoSample_MR | M_VLDL_FC_pct | MDD |
| Weighted mode | 89 | -0.0288734 | 0.0148846 | 0.0556052 | TwoSample_MR | M_VLDL_FC_pct | MDD |
| MR Egger | 83 | 0.0096575 | 0.0223781 | 0.6672078 | TwoSample_MR | M_VLDL_L | MDD |
| Weighted median | 83 | -0.0004089 | 0.0182264 | 0.9821028 | TwoSample_MR | M_VLDL_L | MDD |

| | | | | | | | |
|---|---|---|---|---|---|---|---|
| Inverse variance weighted | 83 | 0.0000181 | 0.0134439 | 0.9989281 | TwoSample_MR | M_VLDL_L | MDD |
| Simple mode | 83 | -0.000328 | 0.0311319 | 0.9916204 | TwoSample_MR | M_VLDL_L | MDD |
| Weighted mode | 83 | -0.0031127 | 0.0200262 | 0.8768634 | TwoSample_MR | M_VLDL_L | MDD |
| MR Egger | 96 | 0.0139845 | 0.0210721 | 0.5085402 | TwoSample_MR | M_VLDL_TG | MDD |
| Weighted median | 96 | 0.0245449 | 0.0178199 | 0.1683925 | TwoSample_MR | M_VLDL_TG | MDD |
| Inverse variance weighted | 96 | 0.016826 | 0.0133384 | 0.2071377 | TwoSample_MR | M_VLDL_TG | MDD |
| Simple mode | 96 | 0.00256 | 0.0299213 | 0.9319971 | TwoSample_MR | M_VLDL_TG | MDD |
| Weighted mode | 96 | 0.0219053 | 0.0155948 | 0.1633864 | TwoSample_MR | M_VLDL_TG | MDD |
| MR Egger | 99 | 0.0274891 | 0.0187458 | 0.1457689 | TwoSample_MR | M_VLDL_TG_pct | MDD |
| Weighted median | 99 | 0.0280126 | 0.0167095 | 0.0936497 | TwoSample_MR | M_VLDL_TG_pct | MDD |
| Inverse variance weighted | 99 | 0.0558883 | 0.0128663 | 0.000014 | TwoSample_MR | M_VLDL_TG_pct | MDD |
| Simple mode | 99 | 0.0220595 | 0.0288361 | 0.4461117 | TwoSample_MR | M_VLDL_TG_pct | MDD |
| Weighted mode | 99 | 0.0284612 | 0.0150733 | 0.0619591 | TwoSample_MR | M_VLDL_TG_pct | MDD |
| MR Egger | 82 | -0.0319599 | 0.0245779 | 0.1972152 | TwoSample_MR | Omega_6_pct | MDD |
| Weighted median | 82 | -0.0352077 | 0.0201465 | 0.0805364 | TwoSample_MR | Omega_6_pct | MDD |
| Inverse variance weighted | 82 | -0.024641 | 0.0148291 | 0.0965796 | TwoSample_MR | Omega_6_pct | MDD |
| Simple mode | 82 | -0.0231237 | 0.0342334 | 0.5012999 | TwoSample_MR | Omega_6_pct | MDD |
| Weighted mode | 82 | -0.0314019 | 0.020396 | 0.1275528 | TwoSample_MR | Omega_6_pct | MDD |
| MR Egger | 89 | -0.0382504 | 0.0234197 | 0.1060285 | TwoSample_MR | PUFA_by_MUFA | MDD |
| Weighted median | 89 | -0.0417372 | 0.0183828 | 0.02318 | TwoSample_MR | PUFA_by_MUFA | MDD |
| Inverse variance weighted | 89 | -0.0474931 | 0.0149248 | 0.0014618 | TwoSample_MR | PUFA_by_MUFA | MDD |
| Simple mode | 89 | -0.0220535 | 0.0289842 | 0.4487621 | TwoSample_MR | PUFA_by_MUFA | MDD |
| Weighted mode | 89 | -0.0523097 | 0.0169321 | 0.0026845 | TwoSample_MR | PUFA_by_MUFA | MDD |
| MR Egger | 72 | -0.050375 | 0.0301444 | 0.0991626 | TwoSample_MR | PUFA_pct | MDD |
| Weighted median | 72 | -0.0486055 | 0.0222414 | 0.0288627 | TwoSample_MR | PUFA_pct | MDD |
| Inverse variance weighted | 72 | -0.0572537 | 0.0176194 | 0.0011562 | TwoSample_MR | PUFA_pct | MDD |
| Simple mode | 72 | -0.0158541 | 0.0388347 | 0.6843229 | TwoSample_MR | PUFA_pct | MDD |
| Weighted mode | 72 | -0.0562293 | 0.0228009 | 0.0160764 | TwoSample_MR | PUFA_pct | MDD |
| MR Egger | 34 | -0.0149478 | 0.0407908 | 0.7164413 | TwoSample_MR | Pyruvate | MDD |
| Weighted median | 34 | 0.0148562 | 0.0327883 | 0.6504803 | TwoSample_MR | Pyruvate | MDD |
| Inverse variance weighted | 34 | 0.0022197 | 0.0216931 | 0.9184997 | TwoSample_MR | Pyruvate | MDD |
| Simple mode | 34 | 0.0411894 | 0.0625865 | 0.515027 | TwoSample_MR | Pyruvate | MDD |
| Weighted mode | 34 | 0.0256781 | 0.0357439 | 0.4775721 | TwoSample_MR | Pyruvate | MDD |
| MR Egger | 83 | -0.011988 | 0.0150033 | 0.4266141 | TwoSample_MR | S_HDL_C_pct | MDD |
| Weighted median | 83 | -0.0104092 | 0.0165662 | 0.5297838 | TwoSample_MR | S_HDL_C_pct | MDD |
| Inverse variance weighted | 83 | -0.0193174 | 0.0104158 | 0.0636513 | TwoSample_MR | S_HDL_C_pct | MDD |
| Simple mode | 83 | -0.0071713 | 0.0282667 | 0.8003613 | TwoSample_MR | S_HDL_C_pct | MDD |
| Weighted mode | 83 | -0.0071713 | 0.0138072 | 0.604888 | TwoSample_MR | S_HDL_C_pct | MDD |
| MR Egger | 86 | -0.0224947 | 0.015803 | 0.1583109 | TwoSample_MR | S_HDL_FC_pct | MDD |
| Weighted median | 86 | -0.0071355 | 0.0147474 | 0.628496 | TwoSample_MR | S_HDL_FC_pct | MDD |
| Inverse variance weighted | 86 | -0.0291485 | 0.0111263 | 0.0087987 | TwoSample_MR | S_HDL_FC_pct | MDD |
| Simple mode | 86 | -0.0167821 | 0.0268894 | 0.5342242 | TwoSample_MR | S_HDL_FC_pct | MDD |
| Weighted mode | 86 | -0.0095608 | 0.0136382 | 0.4852001 | TwoSample_MR | S_HDL_FC_pct | MDD |
| MR Egger | 118 | 0.0267287 | 0.0193284 | 0.169359 | TwoSample_MR | S_HDL_TG | MDD |
| Weighted median | 118 | 0.0257178 | 0.0160743 | 0.1096149 | TwoSample_MR | S_HDL_TG | MDD |
| Inverse variance weighted | 118 | 0.0179925 | 0.012393 | 0.1465508 | TwoSample_MR | S_HDL_TG | MDD |

| | | | | | | | |
|---|---|---|---|---|---|---|---|
| Simple mode | 118 | 0.0116566 | 0.0282686 | 0.6808371 | TwoSample_MR | S_HDL_TG | MDD |
| Weighted mode | 118 | 0.0214114 | 0.0138817 | 0.1256727 | TwoSample_MR | S_HDL_TG | MDD |
| MR Egger | 122 | 0.0215027 | 0.0187815 | 0.2545326 | TwoSample_MR | S_HDL_TG_pct | MDD |
| Weighted median | 122 | 0.0226386 | 0.0153204 | 0.1394957 | TwoSample_MR | S_HDL_TG_pct | MDD |
| Inverse variance weighted | 122 | 0.0150337 | 0.0121121 | 0.2145255 | TwoSample_MR | S_HDL_TG_pct | MDD |
| Simple mode | 122 | 0.0122534 | 0.0262848 | 0.6419257 | TwoSample_MR | S_HDL_TG_pct | MDD |
| Weighted mode | 122 | 0.0155456 | 0.0136719 | 0.2577643 | TwoSample_MR | S_HDL_TG_pct | MDD |
| MR Egger | 67 | -0.0289549 | 0.0220565 | 0.1938807 | TwoSample_MR | S_LDL_PL_pct | MDD |
| Weighted median | 67 | -0.019506 | 0.0178307 | 0.2739739 | TwoSample_MR | S_LDL_PL_pct | MDD |
| Inverse variance weighted | 67 | -0.0239504 | 0.0144362 | 0.0971051 | TwoSample_MR | S_LDL_PL_pct | MDD |
| Simple mode | 67 | -0.015712 | 0.0326951 | 0.6324178 | TwoSample_MR | S_LDL_PL_pct | MDD |
| Weighted mode | 67 | -0.015712 | 0.0154375 | 0.3124985 | TwoSample_MR | S_LDL_PL_pct | MDD |
| MR Egger | 106 | 0.0080347 | 0.0202459 | 0.6922888 | TwoSample_MR | S_LDL_TG | MDD |
| Weighted median | 106 | 0.0158125 | 0.0161041 | 0.3261543 | TwoSample_MR | S_LDL_TG | MDD |
| Inverse variance weighted | 106 | 0.0126737 | 0.0128369 | 0.3235039 | TwoSample_MR | S_LDL_TG | MDD |
| Simple mode | 106 | 0.0082239 | 0.0273829 | 0.76452 | TwoSample_MR | S_LDL_TG | MDD |
| Weighted mode | 106 | 0.0110907 | 0.0143287 | 0.4406578 | TwoSample_MR | S_LDL_TG | MDD |
| MR Egger | 105 | 0.0190286 | 0.0167899 | 0.259704 | TwoSample_MR | S_LDL_TG_pct | MDD |
| Weighted median | 105 | 0.0212991 | 0.0141736 | 0.132909 | TwoSample_MR | S_LDL_TG_pct | MDD |
| Inverse variance weighted | 105 | 0.0325052 | 0.0119005 | 0.0063064 | TwoSample_MR | S_LDL_TG_pct | MDD |
| Simple mode | 105 | 0.0059349 | 0.0270952 | 0.8270488 | TwoSample_MR | S_LDL_TG_pct | MDD |
| Weighted mode | 105 | 0.0187737 | 0.0130651 | 0.15374 | TwoSample_MR | S_LDL_TG_pct | MDD |
| MR Egger | 82 | -0.0355609 | 0.0164624 | 0.0337557 | TwoSample_MR | S_VLDL_C_pct | MDD |
| Weighted median | 82 | -0.0243364 | 0.0162838 | 0.1350412 | TwoSample_MR | S_VLDL_C_pct | MDD |
| Inverse variance weighted | 82 | -0.0346982 | 0.0112834 | 0.0021039 | TwoSample_MR | S_VLDL_C_pct | MDD |
| Simple mode | 82 | -0.0220758 | 0.0278215 | 0.4298163 | TwoSample_MR | S_VLDL_C_pct | MDD |
| Weighted mode | 82 | -0.0243177 | 0.0153877 | 0.1179286 | TwoSample_MR | S_VLDL_C_pct | MDD |
| MR Egger | 93 | -0.0323258 | 0.0177739 | 0.0722429 | TwoSample_MR | S_VLDL_FC_pct | MDD |
| Weighted median | 93 | -0.0229728 | 0.0162979 | 0.1586692 | TwoSample_MR | S_VLDL_FC_pct | MDD |
| Inverse variance weighted | 93 | -0.0403614 | 0.0123859 | 0.0011193 | TwoSample_MR | S_VLDL_FC_pct | MDD |
| Simple mode | 93 | -0.0092248 | 0.030022 | 0.7593325 | TwoSample_MR | S_VLDL_FC_pct | MDD |
| Weighted mode | 93 | -0.0233141 | 0.0143235 | 0.107012 | TwoSample_MR | S_VLDL_FC_pct | MDD |
| MR Egger | 95 | 0.004342 | 0.0242828 | 0.858477 | TwoSample_MR | S_VLDL_L | MDD |
| Weighted median | 95 | 0.0118603 | 0.0164243 | 0.4702212 | TwoSample_MR | S_VLDL_L | MDD |
| Inverse variance weighted | 95 | -0.0049254 | 0.0138356 | 0.7218425 | TwoSample_MR | S_VLDL_L | MDD |
| Simple mode | 95 | 0.0019504 | 0.0270469 | 0.9426661 | TwoSample_MR | S_VLDL_L | MDD |
| Weighted mode | 95 | 0.0019504 | 0.0156645 | 0.9011763 | TwoSample_MR | S_VLDL_L | MDD |
| MR Egger | 97 | -0.0347647 | 0.0169291 | 0.0427675 | TwoSample_MR | S_VLDL_PL_pct | MDD |
| Weighted median | 97 | -0.0221606 | 0.0155577 | 0.1543272 | TwoSample_MR | S_VLDL_PL_pct | MDD |
| Inverse variance weighted | 97 | -0.0377122 | 0.0118804 | 0.0015018 | TwoSample_MR | S_VLDL_PL_pct | MDD |
| Simple mode | 97 | -0.034759 | 0.0299416 | 0.2485642 | TwoSample_MR | S_VLDL_PL_pct | MDD |
| Weighted mode | 97 | -0.0231759 | 0.0143194 | 0.1088374 | TwoSample_MR | S_VLDL_PL_pct | MDD |
| MR Egger | 100 | 0.0064214 | 0.0221198 | 0.7721979 | TwoSample_MR | S_VLDL_P | MDD |
| Weighted median | 100 | 0.0115647 | 0.0172118 | 0.5016454 | TwoSample_MR | S_VLDL_P | MDD |
| Inverse variance weighted | 100 | -0.0052812 | 0.0128628 | 0.6813797 | TwoSample_MR | S_VLDL_P | MDD |
| Simple mode | 100 | -0.0000177 | 0.0276454 | 0.9994895 | TwoSample_MR | S_VLDL_P | MDD |

| | | | | | | | |
|---|---|---|---|---|---|---|---|
| Weighted mode | 100 | 0.0060504 | 0.0160523 | 0.7070416 | TwoSample_MR | S_VLDL_P | MDD |
| MR Egger | 108 | 0.0107474 | 0.018164 | 0.5553197 | TwoSample_MR | S_VLDL_TG | MDD |
| Weighted median | 108 | 0.0233708 | 0.0165843 | 0.1587739 | TwoSample_MR | S_VLDL_TG | MDD |
| Inverse variance weighted | 108 | 0.0226772 | 0.011752 | 0.0536499 | TwoSample_MR | S_VLDL_TG | MDD |
| Simple mode | 108 | 0.014005 | 0.0280592 | 0.6187164 | TwoSample_MR | S_VLDL_TG | MDD |
| Weighted mode | 108 | 0.0192441 | 0.0147509 | 0.1948261 | TwoSample_MR | S_VLDL_TG | MDD |
| MR Egger | 94 | 0.0316379 | 0.0153259 | 0.0417998 | TwoSample_MR | S_VLDL_TG_pct | MDD |
| Weighted median | 94 | 0.0233712 | 0.0159763 | 0.1435029 | TwoSample_MR | S_VLDL_TG_pct | MDD |
| Inverse variance weighted | 94 | 0.0348882 | 0.0106754 | 0.0010828 | TwoSample_MR | S_VLDL_TG_pct | MDD |
| Simple mode | 94 | 0.0164613 | 0.0305565 | 0.5913696 | TwoSample_MR | S_VLDL_TG_pct | MDD |
| Weighted mode | 94 | 0.0237257 | 0.014532 | 0.105923 | TwoSample_MR | S_VLDL_TG_pct | MDD |
| MR Egger | 83 | -0.0472279 | 0.0212265 | 0.0288656 | TwoSample_MR | Sphingomyelins | MDD |
| Weighted median | 83 | -0.0141809 | 0.0196336 | 0.4701237 | TwoSample_MR | Sphingomyelins | MDD |
| Inverse variance weighted | 83 | -0.02255 | 0.0139601 | 0.1062431 | TwoSample_MR | Sphingomyelins | MDD |
| Simple mode | 83 | -0.0139508 | 0.0367298 | 0.7050598 | TwoSample_MR | Sphingomyelins | MDD |
| Weighted mode | 83 | -0.0166067 | 0.0173182 | 0.3404212 | TwoSample_MR | Sphingomyelins | MDD |
| MR Egger | 125 | 0.0149048 | 0.0209578 | 0.4783167 | TwoSample_MR | TG_by_PG | MDD |
| Weighted median | 125 | 0.0271254 | 0.0165631 | 0.1014828 | TwoSample_MR | TG_by_PG | MDD |
| Inverse variance weighted | 125 | 0.0225461 | 0.0133453 | 0.0911344 | TwoSample_MR | TG_by_PG | MDD |
| Simple mode | 125 | 0.0240462 | 0.0294958 | 0.416497 | TwoSample_MR | TG_by_PG | MDD |
| Weighted mode | 125 | 0.0240462 | 0.016356 | 0.1440466 | TwoSample_MR | TG_by_PG | MDD |
| MR Egger | 77 | -0.0374434 | 0.0231229 | 0.1095767 | TwoSample_MR | Total_CE | MDD |
| Weighted median | 77 | -0.0168026 | 0.0194826 | 0.3884456 | TwoSample_MR | Total_CE | MDD |
| Inverse variance weighted | 77 | -0.0143856 | 0.015189 | 0.3435862 | TwoSample_MR | Total_CE | MDD |
| Simple mode | 77 | -0.0165898 | 0.0361859 | 0.6479302 | TwoSample_MR | Total_CE | MDD |
| Weighted mode | 77 | -0.0165898 | 0.0177059 | 0.3517459 | TwoSample_MR | Total_CE | MDD |
| MR Egger | 85 | 0.0224403 | 0.0319779 | 0.4848025 | TwoSample_MR | Total_P | MDD |
| Weighted median | 85 | 0.0119775 | 0.0231666 | 0.6051446 | TwoSample_MR | Total_P | MDD |
| Inverse variance weighted | 85 | 0.0200544 | 0.017438 | 0.2501265 | TwoSample_MR | Total_P | MDD |
| Simple mode | 85 | -0.0054625 | 0.0395833 | 0.8905703 | TwoSample_MR | Total_P | MDD |
| Weighted mode | 85 | 0.0043885 | 0.0227165 | 0.8472792 | TwoSample_MR | Total_P | MDD |
| MR Egger | 105 | 0.0011506 | 0.0200552 | 0.9543597 | TwoSample_MR | Total_TG | MDD |
| Weighted median | 105 | 0.0248485 | 0.0177665 | 0.1619287 | TwoSample_MR | Total_TG | MDD |
| Inverse variance weighted | 105 | 0.0221601 | 0.0129814 | 0.0878108 | TwoSample_MR | Total_TG | MDD |
| Simple mode | 105 | 0.0027735 | 0.0301512 | 0.9268847 | TwoSample_MR | Total_TG | MDD |
| Weighted mode | 105 | 0.0188574 | 0.0156677 | 0.2314842 | TwoSample_MR | Total_TG | MDD |
| MR Egger | 70 | -0.062081 | 0.0173227 | 0.0006319 | TwoSample_MR | Unsaturation | MDD |
| Weighted median | 70 | -0.0455214 | 0.0146321 | 0.0018641 | TwoSample_MR | Unsaturation | MDD |
| Inverse variance weighted | 70 | -0.0413277 | 0.0131343 | 0.0016521 | TwoSample_MR | Unsaturation | MDD |
| Simple mode | 70 | -0.0745106 | 0.0382092 | 0.0552321 | TwoSample_MR | Unsaturation | MDD |
| Weighted mode | 70 | -0.0505906 | 0.0133715 | 0.0003259 | TwoSample_MR | Unsaturation | MDD |
| MR Egger | 86 | 0.0109256 | 0.02622 | 0.6779688 | TwoSample_MR | VLDL_FC | MDD |
| Weighted median | 86 | 0.0037609 | 0.017913 | 0.8337052 | TwoSample_MR | VLDL_FC | MDD |
| Inverse variance weighted | 86 | -0.0095826 | 0.0148097 | 0.5175994 | TwoSample_MR | VLDL_FC | MDD |
| Simple mode | 86 | -0.0009255 | 0.0349221 | 0.9789192 | TwoSample_MR | VLDL_FC | MDD |
| Weighted mode | 86 | 0.0080622 | 0.0198818 | 0.6861223 | TwoSample_MR | VLDL_FC | MDD |

| | | | | | | | |
|---|---|---|---|---|---|---|---|
| MR Egger | 88 | 0.0148878 | 0.0210775 | 0.4818887 | TwoSample_MR | VLDL_L | MDD |
| Weighted median | 88 | 0.0154725 | 0.0183709 | 0.3996594 | TwoSample_MR | VLDL_L | MDD |
| Inverse variance weighted | 88 | 0.0031596 | 0.0130854 | 0.8091975 | TwoSample_MR | VLDL_L | MDD |
| Simple mode | 88 | 0.0018194 | 0.0345637 | 0.9581403 | TwoSample_MR | VLDL_L | MDD |
| Weighted mode | 88 | 0.0168775 | 0.0181776 | 0.3557285 | TwoSample_MR | VLDL_L | MDD |
| MR Egger | 88 | 0.0109842 | 0.0235726 | 0.6424112 | TwoSample_MR | VLDL_PL | MDD |
| Weighted median | 88 | 0.0154309 | 0.0185464 | 0.4053985 | TwoSample_MR | VLDL_PL | MDD |
| Inverse variance weighted | 88 | -0.0047051 | 0.0136794 | 0.7308777 | TwoSample_MR | VLDL_PL | MDD |
| Simple mode | 88 | 0.0032921 | 0.0328589 | 0.920424 | TwoSample_MR | VLDL_PL | MDD |
| Weighted mode | 88 | 0.0124512 | 0.0188823 | 0.5113739 | TwoSample_MR | VLDL_PL | MDD |
| MR Egger | 104 | 0.0036897 | 0.0196477 | 0.8514134 | TwoSample_MR | VLDL_TG | MDD |
| Weighted median | 104 | 0.0245402 | 0.0169368 | 0.1473566 | TwoSample_MR | VLDL_TG | MDD |
| Inverse variance weighted | 104 | 0.0212492 | 0.0128779 | 0.0989311 | TwoSample_MR | VLDL_TG | MDD |
| Simple mode | 104 | -0.0019564 | 0.0319509 | 0.9512942 | TwoSample_MR | VLDL_TG | MDD |
| Weighted mode | 104 | 0.0173703 | 0.0160012 | 0.2802058 | TwoSample_MR | VLDL_TG | MDD |
| MR Egger | 112 | 0.0233491 | 0.0200274 | 0.2461948 | TwoSample_MR | VLDL_size | MDD |
| Weighted median | 112 | 0.0269206 | 0.0157131 | 0.0866644 | TwoSample_MR | VLDL_size | MDD |
| Inverse variance weighted | 112 | 0.0285317 | 0.0128963 | 0.0269391 | TwoSample_MR | VLDL_size | MDD |
| Simple mode | 112 | 0.0350074 | 0.0282274 | 0.2175197 | TwoSample_MR | VLDL_size | MDD |
| Weighted mode | 112 | 0.0229642 | 0.0140509 | 0.1050162 | TwoSample_MR | VLDL_size | MDD |
| MR Egger | 125 | -0.0258219 | 0.0177102 | 0.1473811 | TwoSample_MR | XL_HDL_CE | MDD |
| Weighted median | 125 | -0.0224752 | 0.0163214 | 0.1685008 | TwoSample_MR | XL_HDL_CE | MDD |
| Inverse variance weighted | 125 | -0.0214417 | 0.0116975 | 0.0667997 | TwoSample_MR | XL_HDL_CE | MDD |
| Simple mode | 125 | -0.0096126 | 0.0324045 | 0.7672352 | TwoSample_MR | XL_HDL_CE | MDD |
| Weighted mode | 125 | -0.0129378 | 0.0138208 | 0.3510387 | TwoSample_MR | XL_HDL_CE | MDD |
| MR Egger | 123 | -0.0216267 | 0.0186156 | 0.2476244 | TwoSample_MR | XL_HDL_C | MDD |
| Weighted median | 123 | -0.0212845 | 0.0158075 | 0.1781467 | TwoSample_MR | XL_HDL_C | MDD |
| Inverse variance weighted | 123 | -0.0250782 | 0.0123511 | 0.0423108 | TwoSample_MR | XL_HDL_C | MDD |
| Simple mode | 123 | -0.0010005 | 0.0344986 | 0.9769118 | TwoSample_MR | XL_HDL_C | MDD |
| Weighted mode | 123 | -0.0150033 | 0.0138207 | 0.279812 | TwoSample_MR | XL_HDL_C | MDD |
| MR Egger | 108 | -0.0372449 | 0.0176394 | 0.037083 | TwoSample_MR | XL_HDL_FC | MDD |
| Weighted median | 108 | -0.0199503 | 0.014842 | 0.1788904 | TwoSample_MR | XL_HDL_FC | MDD |
| Inverse variance weighted | 108 | -0.0270991 | 0.0119252 | 0.0230602 | TwoSample_MR | XL_HDL_FC | MDD |
| Simple mode | 108 | -0.0141622 | 0.0318431 | 0.6573988 | TwoSample_MR | XL_HDL_FC | MDD |
| Weighted mode | 108 | -0.0174225 | 0.0132744 | 0.1921639 | TwoSample_MR | XL_HDL_FC | MDD |
| MR Egger | 124 | 0.0174196 | 0.0200997 | 0.3878297 | TwoSample_MR | XL_HDL_FC_pct | MDD |
| Weighted median | 124 | 0.0106759 | 0.0176023 | 0.5441771 | TwoSample_MR | XL_HDL_FC_pct | MDD |
| Inverse variance weighted | 124 | 0.0015434 | 0.0135594 | 0.9093749 | TwoSample_MR | XL_HDL_FC_pct | MDD |
| Simple mode | 124 | 0.0083835 | 0.034675 | 0.8093589 | TwoSample_MR | XL_HDL_FC_pct | MDD |
| Weighted mode | 124 | 0.0083835 | 0.0127687 | 0.5126893 | TwoSample_MR | XL_HDL_FC_pct | MDD |
| MR Egger | 112 | -0.0313695 | 0.0191386 | 0.1040554 | TwoSample_MR | XL_HDL_L | MDD |
| Weighted median | 112 | -0.0090027 | 0.0163792 | 0.5825655 | TwoSample_MR | XL_HDL_L | MDD |
| Inverse variance weighted | 112 | -0.0183178 | 0.0128211 | 0.1530842 | TwoSample_MR | XL_HDL_L | MDD |
| Simple mode | 112 | -0.0041255 | 0.0336555 | 0.9026606 | TwoSample_MR | XL_HDL_L | MDD |
| Weighted mode | 112 | -0.0079629 | 0.0143154 | 0.579163 | TwoSample_MR | XL_HDL_L | MDD |
| MR Egger | 115 | -0.0274974 | 0.0196746 | 0.1649688 | TwoSample_MR | XL_HDL_PL | MDD |

| | | | | | | | |
|---|---|---|---|---|---|---|---|
| Weighted median | 115 | -0.0078928 | 0.0160883 | 0.623713 | TwoSample_MR | XL_HDL_PL | MDD |
| Inverse variance weighted | 115 | -0.0165251 | 0.0131536 | 0.209 | TwoSample_MR | XL_HDL_PL | MDD |
| Simple mode | 115 | -0.0139491 | 0.0334528 | 0.6774791 | TwoSample_MR | XL_HDL_PL | MDD |
| Weighted mode | 115 | -0.0100441 | 0.0137357 | 0.4661309 | TwoSample_MR | XL_HDL_PL | MDD |
| MR Egger | 104 | -0.0261341 | 0.018954 | 0.1709666 | TwoSample_MR | XL_HDL_P | MDD |
| Weighted median | 104 | -0.0078447 | 0.0153219 | 0.6086566 | TwoSample_MR | XL_HDL_P | MDD |
| Inverse variance weighted | 104 | -0.0135984 | 0.012902 | 0.2918921 | TwoSample_MR | XL_HDL_P | MDD |
| Simple mode | 104 | 0.0010275 | 0.0321419 | 0.9745591 | TwoSample_MR | XL_HDL_P | MDD |
| Weighted mode | 104 | -0.0054466 | 0.0129385 | 0.674662 | TwoSample_MR | XL_HDL_P | MDD |
| MR Egger | 120 | 0.0272115 | 0.0195647 | 0.1668888 | TwoSample_MR | XL_HDL_TG_pct | MDD |
| Weighted median | 120 | 0.0263635 | 0.01535 | 0.0858898 | TwoSample_MR | XL_HDL_TG_pct | MDD |
| Inverse variance weighted | 120 | 0.0294454 | 0.0126257 | 0.0196916 | TwoSample_MR | XL_HDL_TG_pct | MDD |
| Simple mode | 120 | 0.0190348 | 0.0273797 | 0.4882744 | TwoSample_MR | XL_HDL_TG_pct | MDD |
| Weighted mode | 120 | 0.0256673 | 0.0137789 | 0.0649571 | TwoSample_MR | XL_HDL_TG_pct | MDD |
| MR Egger | 89 | 0.0182226 | 0.0249495 | 0.4671201 | TwoSample_MR | XL_VLDL_CE | MDD |
| Weighted median | 89 | 0.0179159 | 0.0180506 | 0.3209356 | TwoSample_MR | XL_VLDL_CE | MDD |
| Inverse variance weighted | 89 | 0.0021999 | 0.0145999 | 0.8802285 | TwoSample_MR | XL_VLDL_CE | MDD |
| Simple mode | 89 | 0.0187992 | 0.030926 | 0.5448353 | TwoSample_MR | XL_VLDL_CE | MDD |
| Weighted mode | 89 | 0.0123278 | 0.0178699 | 0.4920951 | TwoSample_MR | XL_VLDL_CE | MDD |
| MR Egger | 104 | -0.0268953 | 0.0227304 | 0.2394691 | TwoSample_MR | XL_VLDL_CE_pct | MDD |
| Weighted median | 104 | -0.0224657 | 0.0174898 | 0.1989667 | TwoSample_MR | XL_VLDL_CE_pct | MDD |
| Inverse variance weighted | 104 | -0.0520808 | 0.0150888 | 0.0005572 | TwoSample_MR | XL_VLDL_CE_pct | MDD |
| Simple mode | 104 | -0.0223298 | 0.0317615 | 0.4836138 | TwoSample_MR | XL_VLDL_CE_pct | MDD |
| Weighted mode | 104 | -0.0223298 | 0.0160226 | 0.166426 | TwoSample_MR | XL_VLDL_CE_pct | MDD |
| MR Egger | 86 | 0.0162039 | 0.0222618 | 0.4687111 | TwoSample_MR | XL_VLDL_C | MDD |
| Weighted median | 86 | 0.0147959 | 0.0178966 | 0.408381 | TwoSample_MR | XL_VLDL_C | MDD |
| Inverse variance weighted | 86 | 0.0052224 | 0.0136658 | 0.7023494 | TwoSample_MR | XL_VLDL_C | MDD |
| Simple mode | 86 | -0.0001254 | 0.0317951 | 0.9968624 | TwoSample_MR | XL_VLDL_C | MDD |
| Weighted mode | 86 | 0.014207 | 0.0175095 | 0.4194111 | TwoSample_MR | XL_VLDL_C | MDD |
| MR Egger | 104 | -0.0352693 | 0.0226163 | 0.1219843 | TwoSample_MR | XL_VLDL_C_pct | MDD |
| Weighted median | 104 | -0.0230675 | 0.0182748 | 0.2068548 | TwoSample_MR | XL_VLDL_C_pct | MDD |
| Inverse variance weighted | 104 | -0.0509857 | 0.0144565 | 0.0004205 | TwoSample_MR | XL_VLDL_C_pct | MDD |
| Simple mode | 104 | -0.0016744 | 0.0314194 | 0.9576036 | TwoSample_MR | XL_VLDL_C_pct | MDD |
| Weighted mode | 104 | -0.0249079 | 0.0159768 | 0.1220612 | TwoSample_MR | XL_VLDL_C_pct | MDD |
| MR Egger | 92 | 0.0151564 | 0.0205688 | 0.4631227 | TwoSample_MR | XL_VLDL_FC | MDD |
| Weighted median | 92 | 0.0177663 | 0.0178464 | 0.3194875 | TwoSample_MR | XL_VLDL_FC | MDD |
| Inverse variance weighted | 92 | 0.0060255 | 0.0130799 | 0.6450369 | TwoSample_MR | XL_VLDL_FC | MDD |
| Simple mode | 92 | 0.0035345 | 0.0293889 | 0.904538 | TwoSample_MR | XL_VLDL_FC | MDD |
| Weighted mode | 92 | 0.0167799 | 0.0170081 | 0.3264645 | TwoSample_MR | XL_VLDL_FC | MDD |
| MR Egger | 84 | -0.0515638 | 0.0249063 | 0.0415697 | TwoSample_MR | XL_VLDL_FC_pct | MDD |
| Weighted median | 84 | -0.0424769 | 0.0188883 | 0.0245221 | TwoSample_MR | XL_VLDL_FC_pct | MDD |
| Inverse variance weighted | 84 | -0.0641181 | 0.014209 | 0.0000064 | TwoSample_MR | XL_VLDL_FC_pct | MDD |
| Simple mode | 84 | -0.0150902 | 0.0337268 | 0.6557333 | TwoSample_MR | XL_VLDL_FC_pct | MDD |
| Weighted mode | 84 | -0.0363472 | 0.0203285 | 0.0774278 | TwoSample_MR | XL_VLDL_FC_pct | MDD |
| MR Egger | 109 | 0.0203471 | 0.019998 | 0.3112321 | TwoSample_MR | XL_VLDL_L | MDD |
| Weighted median | 109 | 0.0256466 | 0.0174947 | 0.1426569 | TwoSample_MR | XL_VLDL_L | MDD |

| | | | | | | | |
|---|---|---|---|---|---|---|---|
| Inverse variance weighted | 109 | 0.0148945 | 0.0127523 | 0.2428133 | TwoSample_MR | XL_VLDL_L | MDD |
| Simple mode | 109 | 0.0107158 | 0.0304498 | 0.7255878 | TwoSample_MR | XL_VLDL_L | MDD |
| Weighted mode | 109 | 0.0217228 | 0.0166669 | 0.1952275 | TwoSample_MR | XL_VLDL_L | MDD |
| MR Egger | 97 | 0.0211908 | 0.0205129 | 0.3042055 | TwoSample_MR | XL_VLDL_PL | MDD |
| Weighted median | 97 | 0.0201169 | 0.0164257 | 0.2206798 | TwoSample_MR | XL_VLDL_PL | MDD |
| Inverse variance weighted | 97 | 0.0073677 | 0.012999 | 0.5708576 | TwoSample_MR | XL_VLDL_PL | MDD |
| Simple mode | 97 | 0.0077157 | 0.0301372 | 0.7984829 | TwoSample_MR | XL_VLDL_PL | MDD |
| Weighted mode | 97 | 0.0155985 | 0.0159792 | 0.3314323 | TwoSample_MR | XL_VLDL_PL | MDD |
| MR Egger | 107 | 0.0155397 | 0.0206083 | 0.4525086 | TwoSample_MR | XL_VLDL_P | MDD |
| Weighted median | 107 | 0.0261396 | 0.0173263 | 0.1313843 | TwoSample_MR | XL_VLDL_P | MDD |
| Inverse variance weighted | 107 | 0.016159 | 0.0130834 | 0.2168019 | TwoSample_MR | XL_VLDL_P | MDD |
| Simple mode | 107 | 0.0118532 | 0.029975 | 0.6933153 | TwoSample_MR | XL_VLDL_P | MDD |
| Weighted mode | 107 | 0.020091 | 0.0158611 | 0.2080453 | TwoSample_MR | XL_VLDL_P | MDD |
| MR Egger | 110 | 0.0150334 | 0.0201574 | 0.4574067 | TwoSample_MR | XL_VLDL_TG | MDD |
| Weighted median | 110 | 0.0267804 | 0.0175964 | 0.1280273 | TwoSample_MR | XL_VLDL_TG | MDD |
| Inverse variance weighted | 110 | 0.0230817 | 0.012926 | 0.0741511 | TwoSample_MR | XL_VLDL_TG | MDD |
| Simple mode | 110 | 0.0224557 | 0.0309605 | 0.4698211 | TwoSample_MR | XL_VLDL_TG | MDD |
| Weighted mode | 110 | 0.0224557 | 0.0152141 | 0.1428333 | TwoSample_MR | XL_VLDL_TG | MDD |
| MR Egger | 87 | 0.0342148 | 0.0224542 | 0.1312821 | TwoSample_MR | XL_VLDL_TG_pct | MDD |
| Weighted median | 87 | 0.02787 | 0.0183334 | 0.1284676 | TwoSample_MR | XL_VLDL_TG_pct | MDD |
| Inverse variance weighted | 87 | 0.0496514 | 0.0140745 | 0.0004191 | TwoSample_MR | XL_VLDL_TG_pct | MDD |
| Simple mode | 87 | 0.0061838 | 0.0315714 | 0.845175 | TwoSample_MR | XL_VLDL_TG_pct | MDD |
| Weighted mode | 87 | 0.025717 | 0.0171276 | 0.1368916 | TwoSample_MR | XL_VLDL_TG_pct | MDD |
| MR Egger | 107 | -0.0406966 | 0.0188497 | 0.0331269 | TwoSample_MR | XS_VLDL_CE_pct | MDD |
| Weighted median | 107 | -0.0255595 | 0.0150374 | 0.0891805 | TwoSample_MR | XS_VLDL_CE_pct | MDD |
| Inverse variance weighted | 107 | -0.0397608 | 0.0128305 | 0.0019422 | TwoSample_MR | XS_VLDL_CE_pct | MDD |
| Simple mode | 107 | -0.0193852 | 0.0281017 | 0.4918129 | TwoSample_MR | XS_VLDL_CE_pct | MDD |
| Weighted mode | 107 | -0.0286721 | 0.0140637 | 0.0439652 | TwoSample_MR | XS_VLDL_CE_pct | MDD |
| MR Egger | 104 | -0.0393302 | 0.0184812 | 0.035737 | TwoSample_MR | XS_VLDL_C_pct | MDD |
| Weighted median | 104 | -0.026021 | 0.0152782 | 0.0885422 | TwoSample_MR | XS_VLDL_C_pct | MDD |
| Inverse variance weighted | 104 | -0.0429616 | 0.0125419 | 0.0006138 | TwoSample_MR | XS_VLDL_C_pct | MDD |
| Simple mode | 104 | -0.0261104 | 0.0276115 | 0.3465478 | TwoSample_MR | XS_VLDL_C_pct | MDD |
| Weighted mode | 104 | -0.0291609 | 0.013806 | 0.0370873 | TwoSample_MR | XS_VLDL_C_pct | MDD |
| MR Egger | 74 | -0.0399918 | 0.0201761 | 0.0512809 | TwoSample_MR | XS_VLDL_FC_pct | MDD |
| Weighted median | 74 | -0.0300172 | 0.0183401 | 0.1016947 | TwoSample_MR | XS_VLDL_FC_pct | MDD |
| Inverse variance weighted | 74 | -0.0428325 | 0.0128432 | 0.0008529 | TwoSample_MR | XS_VLDL_FC_pct | MDD |
| Simple mode | 74 | -0.0034398 | 0.0376923 | 0.927535 | TwoSample_MR | XS_VLDL_FC_pct | MDD |
| Weighted mode | 74 | -0.0202358 | 0.0191329 | 0.2937057 | TwoSample_MR | XS_VLDL_FC_pct | MDD |
| MR Egger | 87 | 0.0115073 | 0.0232255 | 0.6215537 | TwoSample_MR | XS_VLDL_PL_pct | MDD |
| Weighted median | 87 | 0.016978 | 0.0207707 | 0.4136988 | TwoSample_MR | XS_VLDL_PL_pct | MDD |
| Inverse variance weighted | 87 | 0.0086214 | 0.0137753 | 0.5314067 | TwoSample_MR | XS_VLDL_PL_pct | MDD |
| Simple mode | 87 | 0.0033564 | 0.0351345 | 0.9241168 | TwoSample_MR | XS_VLDL_PL_pct | MDD |
| Weighted mode | 87 | 0.0059667 | 0.0201968 | 0.7683778 | TwoSample_MR | XS_VLDL_PL_pct | MDD |
| MR Egger | 109 | 0.00428 | 0.0163011 | 0.7933935 | TwoSample_MR | XS_VLDL_TG | MDD |
| Weighted median | 109 | -0.00662 | 0.0155188 | 0.6696839 | TwoSample_MR | XS_VLDL_TG | MDD |
| Inverse variance weighted | 109 | 0.0028886 | 0.0110269 | 0.7933538 | TwoSample_MR | XS_VLDL_TG | MDD |

| | | | | | | | |
|---|---|---|---|---|---|---|---|
| Simple mode | 109 | 0.0083506 | 0.0295226 | 0.7778304 | TwoSample_MR | XS_VLDL_TG | MDD |
| Weighted mode | 109 | 0.0022859 | 0.0119744 | 0.848966 | TwoSample_MR | XS_VLDL_TG | MDD |
| MR Egger | 101 | 0.0339355 | 0.0167724 | 0.0457385 | TwoSample_MR | XS_VLDL_TG_pct | MDD |
| Weighted median | 101 | 0.0266839 | 0.0152929 | 0.0810086 | TwoSample_MR | XS_VLDL_TG_pct | MDD |
| Inverse variance weighted | 101 | 0.0422961 | 0.0114302 | 0.0002153 | TwoSample_MR | XS_VLDL_TG_pct | MDD |
| Simple mode | 101 | 0.0126629 | 0.0295227 | 0.6689029 | TwoSample_MR | XS_VLDL_TG_pct | MDD |
| Weighted mode | 101 | 0.0239563 | 0.0139547 | 0.0891261 | TwoSample_MR | XS_VLDL_TG_pct | MDD |
| MR Egger | 96 | 0.0177861 | 0.0204509 | 0.386681 | TwoSample_MR | XXL_VLDL_CE | MDD |
| Weighted median | 96 | 0.025252 | 0.0164475 | 0.124707 | TwoSample_MR | XXL_VLDL_CE | MDD |
| Inverse variance weighted | 96 | 0.0166384 | 0.0134445 | 0.215878 | TwoSample_MR | XXL_VLDL_CE | MDD |
| Simple mode | 96 | 0.007611 | 0.0290326 | 0.7937709 | TwoSample_MR | XXL_VLDL_CE | MDD |
| Weighted mode | 96 | 0.0182457 | 0.015313 | 0.236418 | TwoSample_MR | XXL_VLDL_CE | MDD |
| MR Egger | 101 | 0.0160804 | 0.0188308 | 0.3951987 | TwoSample_MR | XXL_VLDL_C | MDD |
| Weighted median | 101 | 0.0249782 | 0.0164189 | 0.1281833 | TwoSample_MR | XXL_VLDL_C | MDD |
| Inverse variance weighted | 101 | 0.0175507 | 0.0125648 | 0.162468 | TwoSample_MR | XXL_VLDL_C | MDD |
| Simple mode | 101 | 0.0134782 | 0.0290968 | 0.6442133 | TwoSample_MR | XXL_VLDL_C | MDD |
| Weighted mode | 101 | 0.0187012 | 0.0153847 | 0.2270119 | TwoSample_MR | XXL_VLDL_C | MDD |
| MR Egger | 101 | 0.0113039 | 0.0187807 | 0.548625 | TwoSample_MR | XXL_VLDL_FC | MDD |
| Weighted median | 101 | 0.0257062 | 0.0160894 | 0.1101073 | TwoSample_MR | XXL_VLDL_FC | MDD |
| Inverse variance weighted | 101 | 0.0196468 | 0.0126773 | 0.1211985 | TwoSample_MR | XXL_VLDL_FC | MDD |
| Simple mode | 101 | 0.0139467 | 0.030201 | 0.6452292 | TwoSample_MR | XXL_VLDL_FC | MDD |
| Weighted mode | 101 | 0.0197006 | 0.0143766 | 0.1736541 | TwoSample_MR | XXL_VLDL_FC | MDD |
| MR Egger | 99 | 0.0204762 | 0.0196222 | 0.2993009 | TwoSample_MR | XXL_VLDL_L | MDD |
| Weighted median | 99 | 0.0288657 | 0.0167932 | 0.0856338 | TwoSample_MR | XXL_VLDL_L | MDD |
| Inverse variance weighted | 99 | 0.0128918 | 0.013048 | 0.3231397 | TwoSample_MR | XXL_VLDL_L | MDD |
| Simple mode | 99 | 0.0170567 | 0.0299807 | 0.5707099 | TwoSample_MR | XXL_VLDL_L | MDD |
| Weighted mode | 99 | 0.0170567 | 0.0164089 | 0.3011383 | TwoSample_MR | XXL_VLDL_L | MDD |
| MR Egger | 102 | 0.0070633 | 0.0194923 | 0.7178437 | TwoSample_MR | XXL_VLDL_PL | MDD |
| Weighted median | 102 | 0.0257775 | 0.0158504 | 0.1038856 | TwoSample_MR | XXL_VLDL_PL | MDD |
| Inverse variance weighted | 102 | 0.0190974 | 0.0130256 | 0.1426096 | TwoSample_MR | XXL_VLDL_PL | MDD |
| Simple mode | 102 | 0.0175906 | 0.0278363 | 0.5288596 | TwoSample_MR | XXL_VLDL_PL | MDD |
| Weighted mode | 102 | 0.0207415 | 0.0158813 | 0.1945082 | TwoSample_MR | XXL_VLDL_PL | MDD |
| MR Egger | 56 | -0.0065204 | 0.0190132 | 0.7329734 | TwoSample_MR | XXL_VLDL_PL_pct | MDD |
| Weighted median | 56 | -0.0193634 | 0.0169925 | 0.2544824 | TwoSample_MR | XXL_VLDL_PL_pct | MDD |
| Inverse variance weighted | 56 | 0.0067425 | 0.0124161 | 0.5871003 | TwoSample_MR | XXL_VLDL_PL_pct | MDD |
| Simple mode | 56 | -0.0163518 | 0.0309439 | 0.5993237 | TwoSample_MR | XXL_VLDL_PL_pct | MDD |
| Weighted mode | 56 | -0.0047181 | 0.0146063 | 0.7479056 | TwoSample_MR | XXL_VLDL_PL_pct | MDD |
| MR Egger | 98 | 0.0127021 | 0.0194588 | 0.5154655 | TwoSample_MR | XXL_VLDL_P | MDD |
| Weighted median | 98 | 0.02396 | 0.0167838 | 0.1534182 | TwoSample_MR | XXL_VLDL_P | MDD |
| Inverse variance weighted | 98 | 0.0165824 | 0.0129327 | 0.1997706 | TwoSample_MR | XXL_VLDL_P | MDD |
| Simple mode | 98 | 0.0117666 | 0.0294057 | 0.6899292 | TwoSample_MR | XXL_VLDL_P | MDD |
| Weighted mode | 98 | 0.0198659 | 0.0155383 | 0.2041182 | TwoSample_MR | XXL_VLDL_P | MDD |
| MR Egger | 93 | 0.0144875 | 0.0179151 | 0.420812 | TwoSample_MR | XXL_VLDL_TG | MDD |
| Weighted median | 93 | 0.0069284 | 0.0174355 | 0.6910912 | TwoSample_MR | XXL_VLDL_TG | MDD |
| Inverse variance weighted | 93 | 0.0169425 | 0.0118567 | 0.1530216 | TwoSample_MR | XXL_VLDL_TG | MDD |
| Simple mode | 93 | 0.0131409 | 0.0347605 | 0.7062697 | TwoSample_MR | XXL_VLDL_TG | MDD |

| | | | | | | | |
|---|---|---|---|---|---|---|---|
| Weighted mode | 93 | 0.0185089 | 0.0158267 | 0.2452359 | TwoSample_MR | XXL_VLDL_TG | MDD |

**Supplementary Table 7**

**Results of horizontal pleiotropy in MR**

| analysis | egger_intercept | se | pval | exposure | outcome |
|---|---|---|---|---|---|
| Horizontal_pleiotropy | 0.0070856 | 0.002505 | 0.005593 | MDD | XL_HDL_FC_pct |
| Horizontal_pleiotropy | -0.0064994 | 0.0025612 | 0.0126159 | MDD | L_HDL_PL |
| Horizontal_pleiotropy | -0.0065219 | 0.0026212 | 0.0143999 | MDD | L_HDL_L |
| Horizontal_pleiotropy | -0.0066615 | 0.0027098 | 0.0155799 | MDD | L_HDL_CE |
| Horizontal_pleiotropy | -0.0063312 | 0.002607 | 0.0168417 | MDD | HDL_CE |
| Horizontal_pleiotropy | -0.0065201 | 0.0026977 | 0.0173589 | MDD | L_HDL_C |
| Horizontal_pleiotropy | -0.0063398 | 0.0026431 | 0.0182064 | MDD | HDL_size |
| Horizontal_pleiotropy | -0.0063532 | 0.0026717 | 0.0191997 | MDD | L_HDL_P |
| Horizontal_pleiotropy | 0.006187 | 0.0026024 | 0.0192292 | MDD | L_VLDL_TG |
| Horizontal_pleiotropy | -0.0059683 | 0.0025296 | 0.0201362 | MDD | M_HDL_CE |
| Horizontal_pleiotropy | -0.0061446 | 0.0026123 | 0.0205101 | MDD | HDL_C |
| Horizontal_pleiotropy | 0.0054951 | 0.0023705 | 0.0223636 | MDD | XL_VLDL_CE |
| Horizontal_pleiotropy | -0.0058795 | 0.002539 | 0.0225005 | MDD | M_HDL_C |
| Horizontal_pleiotropy | 0.0059271 | 0.002572 | 0.0231485 | MDD | L_VLDL_L |
| Horizontal_pleiotropy | 0.005684 | 0.0024758 | 0.023654 | MDD | XL_VLDL_C |
| Horizontal_pleiotropy | 0.0057246 | 0.0025329 | 0.0258577 | MDD | L_VLDL_FC |
| Horizontal_pleiotropy | -0.0059594 | 0.0026464 | 0.0263927 | MDD | L_HDL_FC |
| Horizontal_pleiotropy | -0.005751 | 0.0025549 | 0.0264506 | MDD | HDL_L |
| Horizontal_pleiotropy | 0.0058704 | 0.0026154 | 0.026876 | MDD | XL_VLDL_PL |
| Horizontal_pleiotropy | 0.005473 | 0.0024486 | 0.027502 | MDD | L_VLDL_C |
| Horizontal_pleiotropy | 0.005924 | 0.0026566 | 0.0278596 | MDD | TG_by_PG |
| Horizontal_pleiotropy | 0.0057438 | 0.0025979 | 0.0291869 | MDD | L_VLDL_PL |
| Horizontal_pleiotropy | -0.0055577 | 0.002527 | 0.0300256 | MDD | HDL_PL |
| Horizontal_pleiotropy | 0.0058443 | 0.0026661 | 0.0305611 | MDD | XL_VLDL_L |
| Horizontal_pleiotropy | 0.0056312 | 0.0025697 | 0.030611 | MDD | XL_VLDL_FC |
| Horizontal_pleiotropy | 0.0055879 | 0.0025671 | 0.0317215 | MDD | L_VLDL_P |
| Horizontal_pleiotropy | -0.0055088 | 0.0025575 | 0.0335107 | MDD | M_HDL_P |
| Horizontal_pleiotropy | 0.005101 | 0.002379 | 0.0343066 | MDD | L_VLDL_CE |
| Horizontal_pleiotropy | 0.0058754 | 0.0027494 | 0.0348998 | MDD | XL_VLDL_TG |
| Horizontal_pleiotropy | 0.0056723 | 0.0026598 | 0.0352678 | MDD | VLDL_size |
| Horizontal_pleiotropy | 0.005639 | 0.0026533 | 0.0358894 | MDD | XL_VLDL_P |
| Horizontal_pleiotropy | -0.0054222 | 0.0025612 | 0.0365914 | MDD | M_HDL_FC |
| Horizontal_pleiotropy | 0.004834 | 0.002331 | 0.0405188 | MDD | M_VLDL_L |
| Horizontal_pleiotropy | -0.0053763 | 0.0025954 | 0.0407408 | MDD | HDL_FC |
| Horizontal_pleiotropy | 0.0048964 | 0.002364 | 0.0407635 | MDD | VLDL_FC |
| Horizontal_pleiotropy | 0.0049819 | 0.0024212 | 0.0420868 | MDD | VLDL_PL |
| Horizontal_pleiotropy | 0.0020594 | 0.0010025 | 0.0426452 | M_VLDL_TG_pct | MDD |
| Horizontal_pleiotropy | 0.0051527 | 0.0025174 | 0.0431442 | MDD | VLDL_L |
| Horizontal_pleiotropy | -0.0051996 | 0.0025681 | 0.0454161 | MDD | ApoA1 |
| Horizontal_pleiotropy | 0.0050364 | 0.0025613 | 0.0518727 | MDD | M_VLDL_TG |
| Horizontal_pleiotropy | 0.0051845 | 0.0026793 | 0.055647 | MDD | VLDL_TG |
| Horizontal_pleiotropy | 0.0051442 | 0.0027393 | 0.063144 | MDD | XXL_VLDL_TG |
| Horizontal_pleiotropy | -0.0048905 | 0.0027225 | 0.0752898 | MDD | XL_HDL_PL |

| | | | | | |
|---|---|---|---|---|---|
| Horizontal_pleiotropy | 0.001889 | 0.0010497 | 0.0763554 | Unsaturation | MDD |
| Horizontal_pleiotropy | 0.0048259 | 0.0027158 | 0.0784382 | MDD | L_HDL_TG_pct |
| Horizontal_pleiotropy | -0.0049412 | 0.0027885 | 0.0792701 | MDD | Omega_6_pct |
| Horizontal_pleiotropy | 0.0048067 | 0.00274 | 0.0822725 | MDD | XXL_VLDL_L |
| Horizontal_pleiotropy | -0.0055231 | 0.0031492 | 0.0823495 | MDD | PUFA_pct |
| Horizontal_pleiotropy | -0.0049311 | 0.0028496 | 0.08646 | MDD | XL_HDL_CE |
| Horizontal_pleiotropy | 0.0040724 | 0.0023627 | 0.0876912 | MDD | S_VLDL_P |
| Horizontal_pleiotropy | -0.0066459 | 0.0038674 | 0.0886392 | MDD | LA_pct |
| Horizontal_pleiotropy | -0.0048371 | 0.0028201 | 0.0892264 | MDD | M_HDL_C_pct |
| Horizontal_pleiotropy | 0.0040153 | 0.0023731 | 0.0935893 | MDD | S_VLDL_L |
| Horizontal_pleiotropy | 0.0045311 | 0.0026901 | 0.0950512 | MDD | Total_TG |
| Horizontal_pleiotropy | 0.004821 | 0.0028748 | 0.0964897 | MDD | M_HDL_PL_pct |
| Horizontal_pleiotropy | -0.0046323 | 0.0027757 | 0.0980945 | MDD | XL_HDL_L |
| Horizontal_pleiotropy | 0.0044878 | 0.0027379 | 0.1041491 | MDD | XXL_VLDL_P |
| Horizontal_pleiotropy | -0.0044515 | 0.0027648 | 0.110364 | MDD | M_HDL_FC_pct |
| Horizontal_pleiotropy | -0.0041963 | 0.0026247 | 0.1128423 | MDD | HDL_P |
| Horizontal_pleiotropy | 0.0043413 | 0.0027237 | 0.1139363 | MDD | M_HDL_TG_pct |
| Horizontal_pleiotropy | 0.0044043 | 0.0027964 | 0.118239 | MDD | XL_HDL_TG_pct |
| Horizontal_pleiotropy | 0.004218 | 0.0026987 | 0.1210422 | MDD | S_HDL_TG_pct |
| Horizontal_pleiotropy | -0.0042954 | 0.0027565 | 0.1221516 | MDD | M_HDL_CE_pct |
| Horizontal_pleiotropy | -0.0044328 | 0.0028706 | 0.125522 | MDD | XL_HDL_C |
| Horizontal_pleiotropy | 0.0041977 | 0.0027356 | 0.1278905 | MDD | XXL_VLDL_PL |
| Horizontal_pleiotropy | 0.0017237 | 0.0011237 | 0.1289551 | Sphingomyelins | MDD |
| Horizontal_pleiotropy | -0.0044037 | 0.002889 | 0.1304113 | MDD | L_HDL_CE_pct |
| Horizontal_pleiotropy | 0.0034881 | 0.0023071 | 0.1335347 | MDD | XS_VLDL_PL_pct |
| Horizontal_pleiotropy | -0.0041002 | 0.0027595 | 0.1402823 | MDD | XL_HDL_P |
| Horizontal_pleiotropy | 0.00407 | 0.0027395 | 0.1403279 | MDD | XXL_VLDL_CE |
| Horizontal_pleiotropy | -0.0043757 | 0.0029632 | 0.1427192 | MDD | L_HDL_C_pct |
| Horizontal_pleiotropy | -0.0040416 | 0.0027388 | 0.1429855 | MDD | L_VLDL_CE_pct |
| Horizontal_pleiotropy | -0.0016755 | 0.001136 | 0.1433043 | XL_VLDL_CE_pct | MDD |
| Horizontal_pleiotropy | 0.0038801 | 0.0027513 | 0.161388 | MDD | XXL_VLDL_C |
| Horizontal_pleiotropy | -0.0015203 | 0.0010986 | 0.1697688 | L_HDL_FC_pct | MDD |
| Horizontal_pleiotropy | 0.0037905 | 0.0027568 | 0.1720362 | MDD | XXL_VLDL_FC |
| Horizontal_pleiotropy | 0.0013819 | 0.0010085 | 0.1736005 | Total_TG | MDD |
| Horizontal_pleiotropy | -0.0015687 | 0.0011475 | 0.1751008 | L_VLDL_CE | MDD |
| Horizontal_pleiotropy | -0.0015482 | 0.0011332 | 0.1752731 | L_VLDL_CE_pct | MDD |
| Horizontal_pleiotropy | -0.0033733 | 0.0024737 | 0.1755576 | MDD | XXL_VLDL_PL_pct |
| Horizontal_pleiotropy | -0.0034692 | 0.0026265 | 0.1893917 | MDD | Total_P |
| Horizontal_pleiotropy | 0.003352 | 0.0025466 | 0.1909271 | MDD | S_LDL_TG |
| Horizontal_pleiotropy | 0.0016427 | 0.0012466 | 0.1916005 | Total_CE | MDD |
| Horizontal_pleiotropy | -0.0012682 | 0.0009674 | 0.1928323 | M_VLDL_CE_pct | MDD |
| Horizontal_pleiotropy | -0.0037177 | 0.0028634 | 0.1969894 | MDD | XL_VLDL_FC_pct |
| Horizontal_pleiotropy | 0.0034055 | 0.00265 | 0.2015654 | MDD | L_VLDL_PL_pct |
| Horizontal_pleiotropy | 0.001217 | 0.0009668 | 0.2104218 | HDL_CE | MDD |
| Horizontal_pleiotropy | -0.0043369 | 0.0034577 | 0.2125016 | MDD | PUFA_by_MUFA |
| Horizontal_pleiotropy | -0.0035832 | 0.0028594 | 0.2129144 | MDD | XL_VLDL_C_pct |

| | | | | | |
|---|---|---|---|---|---|
| Horizontal_pleiotropy | 0.0013492 | 0.0010782 | 0.2142583 | M_HDL_TG | MDD |
| Horizontal_pleiotropy | 0.0014574 | 0.0011663 | 0.2147908 | MUFA | MDD |
| Horizontal_pleiotropy | 0.001345 | 0.0010795 | 0.2159698 | HDL_TG | MDD |
| Horizontal_pleiotropy | 0.0034588 | 0.0029199 | 0.2388347 | MDD | S_HDL_TG |
| Horizontal_pleiotropy | -0.0033473 | 0.002827 | 0.239043 | MDD | XL_VLDL_CE_pct |
| Horizontal_pleiotropy | 0.0011742 | 0.0009937 | 0.2400956 | VLDL_TG | MDD |
| Horizontal_pleiotropy | 0.0028014 | 0.0024335 | 0.2522447 | MDD | M_LDL_TG |
| Horizontal_pleiotropy | 0.0030187 | 0.0026493 | 0.2570961 | MDD | S_VLDL_TG |
| Horizontal_pleiotropy | 0.0011006 | 0.0009686 | 0.2584874 | S_LDL_TG_pct | MDD |
| Horizontal_pleiotropy | 0.0014781 | 0.0013306 | 0.2695084 | HDL_PL | MDD |
| Horizontal_pleiotropy | 0.0010969 | 0.0009907 | 0.2704057 | HDL_C | MDD |
| Horizontal_pleiotropy | 0.0014136 | 0.001279 | 0.2718666 | M_HDL_P | MDD |
| Horizontal_pleiotropy | -0.0010794 | 0.0010092 | 0.2869516 | XL_HDL_FC_pct | MDD |
| Horizontal_pleiotropy | -0.0011928 | 0.0011223 | 0.2907964 | M_VLDL_FC_pct | MDD |
| Horizontal_pleiotropy | 0.0013359 | 0.0012699 | 0.2951026 | M_HDL_CE | MDD |
| Horizontal_pleiotropy | -0.0012764 | 0.001221 | 0.2987232 | L_VLDL_C | MDD |
| Horizontal_pleiotropy | 0.0029887 | 0.0028963 | 0.3044774 | MDD | XS_VLDL_FC_pct |
| Horizontal_pleiotropy | -0.0010121 | 0.0009827 | 0.3055334 | M_VLDL_C_pct | MDD |
| Horizontal_pleiotropy | 0.0029915 | 0.0029366 | 0.3106662 | MDD | L_HDL_PL_pct |
| Horizontal_pleiotropy | 0.0012438 | 0.0012334 | 0.315626 | M_HDL_C | MDD |
| Horizontal_pleiotropy | 0.0011227 | 0.0011255 | 0.3207864 | IDL_TG_pct | MDD |
| Horizontal_pleiotropy | 0.0009414 | 0.0009484 | 0.3237505 | M_LDL_TG_pct | MDD |
| Horizontal_pleiotropy | 0.0027317 | 0.0027868 | 0.3292103 | MDD | XL_VLDL_TG_pct |
| Horizontal_pleiotropy | 0.0015767 | 0.0016053 | 0.3311415 | Citrate | MDD |
| Horizontal_pleiotropy | -0.0012722 | 0.0013418 | 0.3458028 | VLDL_FC | MDD |
| Horizontal_pleiotropy | -0.0027509 | 0.0029194 | 0.3481894 | MDD | M_VLDL_CE_pct |
| Horizontal_pleiotropy | -0.0022397 | 0.0023743 | 0.349651 | Ala | MDD |
| Horizontal_pleiotropy | 0.0008784 | 0.0009558 | 0.3600769 | XL_HDL_L | MDD |
| Horizontal_pleiotropy | 0.0011068 | 0.0012004 | 0.3606209 | XXL_VLDL_PL_pct | MDD |
| Horizontal_pleiotropy | 0.0025524 | 0.0028004 | 0.3641339 | MDD | MUFA |
| Horizontal_pleiotropy | -0.0044933 | 0.004931 | 0.3642372 | MDD | Unsaturation |
| Horizontal_pleiotropy | -0.0010164 | 0.0011241 | 0.3680372 | XL_VLDL_C_pct | MDD |
| Horizontal_pleiotropy | 0.0032802 | 0.0036296 | 0.3681908 | MDD | MUFA_pct |
| Horizontal_pleiotropy | 0.0009043 | 0.0010009 | 0.3683777 | XL_HDL_P | MDD |
| Horizontal_pleiotropy | 0.0010562 | 0.0011961 | 0.3797062 | XL_VLDL_TG_pct | MDD |
| Horizontal_pleiotropy | -0.000917 | 0.0010518 | 0.3854932 | XL_VLDL_PL | MDD |
| Horizontal_pleiotropy | 0.0008227 | 0.0009542 | 0.3905719 | S_VLDL_TG | MDD |
| Horizontal_pleiotropy | 0.0008771 | 0.0010556 | 0.4080093 | XXL_VLDL_PL | MDD |
| Horizontal_pleiotropy | -0.002425 | 0.0029308 | 0.4098569 | MDD | M_VLDL_C_pct |
| Horizontal_pleiotropy | -0.0024059 | 0.0029143 | 0.4109144 | MDD | XL_HDL_FC |
| Horizontal_pleiotropy | -0.0009995 | 0.0012218 | 0.415586 | VLDL_PL | MDD |
| Horizontal_pleiotropy | -0.0010162 | 0.0012818 | 0.4300297 | XL_VLDL_CE | MDD |
| Horizontal_pleiotropy | 0.0007269 | 0.0009298 | 0.4360826 | XL_HDL_FC | MDD |
| Horizontal_pleiotropy | -0.0020568 | 0.0027045 | 0.4486449 | MDD | S_HDL_FC_pct |
| Horizontal_pleiotropy | 0.0019139 | 0.0025475 | 0.4541411 | MDD | XS_VLDL_TG |
| Horizontal_pleiotropy | 0.0007235 | 0.0009632 | 0.4541608 | XL_HDL_PL | MDD |

| | | | | | |
|---|---|---|---|---|---|
| Horizontal_pleiotropy | -0.0009403 | 0.0012496 | 0.4544126 | IDL_FC_pct | MDD |
| Horizontal_pleiotropy | -0.0008208 | 0.001134 | 0.4710113 | L_VLDL_FC | MDD |
| Horizontal_pleiotropy | -0.0007668 | 0.0010784 | 0.4789755 | VLDL_L | MDD |
| Horizontal_pleiotropy | 0.0020492 | 0.0028985 | 0.4811238 | MDD | M_VLDL_TG_pct |
| Horizontal_pleiotropy | 0.0006611 | 0.0009439 | 0.4848866 | L_HDL_FC | MDD |
| Horizontal_pleiotropy | 0.0016687 | 0.002393 | 0.4871249 | MDD | IDL_TG |
| Horizontal_pleiotropy | 0.0006458 | 0.0009459 | 0.496374 | XS_VLDL_TG_pct | MDD |
| Horizontal_pleiotropy | -0.000645 | 0.0009474 | 0.4979237 | S_HDL_C_pct | MDD |
| Horizontal_pleiotropy | 0.0006507 | 0.0009784 | 0.5076204 | L_LDL_TG_pct | MDD |
| Horizontal_pleiotropy | -0.0007124 | 0.0010928 | 0.5161681 | L_VLDL_L | MDD |
| Horizontal_pleiotropy | -0.0007265 | 0.0011155 | 0.516385 | S_VLDL_P | MDD |
| Horizontal_pleiotropy | -0.00168 | 0.0026101 | 0.5211959 | MDD | M_LDL_TG_pct |
| Horizontal_pleiotropy | 0.0008463 | 0.0013204 | 0.5231844 | ApoA1 | MDD |
| Horizontal_pleiotropy | -0.0006471 | 0.0010233 | 0.5287615 | S_VLDL_FC_pct | MDD |
| Horizontal_pleiotropy | -0.0007348 | 0.0011734 | 0.5328487 | XL_VLDL_C | MDD |
| Horizontal_pleiotropy | 0.0005788 | 0.000936 | 0.5373154 | L_HDL_P | MDD |
| Horizontal_pleiotropy | -0.0007568 | 0.0012308 | 0.5403497 | XL_VLDL_FC_pct | MDD |
| Horizontal_pleiotropy | 0.0006038 | 0.0010001 | 0.5474138 | XXL_VLDL_FC | MDD |
| Horizontal_pleiotropy | -0.0013485 | 0.0022538 | 0.5508945 | MDD | Citrate |
| Horizontal_pleiotropy | -0.0006004 | 0.0010088 | 0.5533391 | S_HDL_FC_pct | MDD |
| Horizontal_pleiotropy | -0.0005875 | 0.0009955 | 0.5562346 | S_HDL_TG | MDD |
| Horizontal_pleiotropy | -0.0006171 | 0.0010702 | 0.565616 | XL_VLDL_FC | MDD |
| Horizontal_pleiotropy | 0.0006114 | 0.001072 | 0.5697697 | IDL_TG | MDD |
| Horizontal_pleiotropy | -0.0006166 | 0.0011415 | 0.5905705 | M_VLDL_L | MDD |
| Horizontal_pleiotropy | 0.0006111 | 0.001133 | 0.591322 | IDL_FC | MDD |
| Horizontal_pleiotropy | 0.0005197 | 0.0009965 | 0.6030436 | XL_VLDL_TG | MDD |
| Horizontal_pleiotropy | -0.0005423 | 0.0010447 | 0.6048993 | XXL_VLDL_L | MDD |
| Horizontal_pleiotropy | -0.001229 | 0.0023764 | 0.6061037 | MDD | Ala |
| Horizontal_pleiotropy | -0.0006188 | 0.0012048 | 0.6088339 | PUFA_by_MUFA | MDD |
| Horizontal_pleiotropy | -0.000535 | 0.0010531 | 0.6123612 | M_HDL_TG_pct | MDD |
| Horizontal_pleiotropy | -0.0014771 | 0.002936 | 0.6159365 | MDD | M_VLDL_FC_pct |
| Horizontal_pleiotropy | 0.0009179 | 0.0018385 | 0.6210045 | Pyruvate | MDD |
| Horizontal_pleiotropy | -0.0014977 | 0.0030737 | 0.6270893 | MDD | IDL_FC_pct |
| Horizontal_pleiotropy | -0.0013567 | 0.0028347 | 0.6332035 | MDD | L_HDL_FC_pct |
| Horizontal_pleiotropy | 0.0004795 | 0.0010119 | 0.6364233 | TG_by_PG | MDD |
| Horizontal_pleiotropy | -0.0005758 | 0.0012374 | 0.6427774 | S_VLDL_L | MDD |
| Horizontal_pleiotropy | -0.000415 | 0.000906 | 0.6477171 | L_HDL_C_pct | MDD |
| Horizontal_pleiotropy | 0.0011324 | 0.0024755 | 0.6482856 | MDD | IDL_L |
| Horizontal_pleiotropy | -0.0004898 | 0.0010785 | 0.6506293 | L_HDL_CE_pct | MDD |
| Horizontal_pleiotropy | -0.0004439 | 0.0009825 | 0.6522729 | S_HDL_TG_pct | MDD |
| Horizontal_pleiotropy | 0.0010956 | 0.0024967 | 0.6617018 | MDD | IDL_CE |
| Horizontal_pleiotropy | -0.000572 | 0.0013966 | 0.683129 | HDL_P | MDD |
| Horizontal_pleiotropy | 0.0010296 | 0.0025322 | 0.6851058 | MDD | IDL_C |
| Horizontal_pleiotropy | 0.0003752 | 0.0009496 | 0.6933002 | L_HDL_C | MDD |
| Horizontal_pleiotropy | 0.0010403 | 0.0027466 | 0.7056161 | MDD | S_VLDL_C_pct |
| Horizontal_pleiotropy | 0.0004121 | 0.0011002 | 0.7089826 | Omega_6_pct | MDD |

| | | | | | |
|---|---|---|---|---|---|
| Horizontal_pleiotropy | -0.0003973 | 0.0010641 | 0.7096158 | L_VLDL_PL_pct | MDD |
| Horizontal_pleiotropy | -0.0003498 | 0.0009856 | 0.7233241 | XL_VLDL_L | MDD |
| Horizontal_pleiotropy | 0.000469 | 0.0013243 | 0.7239987 | M_HDL_FC | MDD |
| Horizontal_pleiotropy | 0.0003794 | 0.0010773 | 0.7254876 | MUFA_pct | MDD |
| Horizontal_pleiotropy | 0.0009737 | 0.0027671 | 0.7256321 | MDD | HDL_TG |
| Horizontal_pleiotropy | 0.0003534 | 0.001042 | 0.7351079 | VLDL_size | MDD |
| Horizontal_pleiotropy | 0.0002935 | 0.0008884 | 0.7417109 | XL_HDL_CE | MDD |
| Horizontal_pleiotropy | -0.0003296 | 0.0010086 | 0.7444832 | L_VLDL_TG | MDD |
| Horizontal_pleiotropy | -0.0009128 | 0.0028341 | 0.7480361 | MDD | XS_VLDL_CE_pct |
| Horizontal_pleiotropy | 0.0003455 | 0.0010827 | 0.7502466 | M_HDL_FC_pct | MDD |
| Horizontal_pleiotropy | 0.0008374 | 0.0026384 | 0.7515783 | MDD | IDL_FC |
| Horizontal_pleiotropy | 0.000375 | 0.0012161 | 0.7584327 | HDL_L | MDD |
| Horizontal_pleiotropy | 0.0003684 | 0.0011972 | 0.7588732 | L_HDL_PL_pct | MDD |
| Horizontal_pleiotropy | 0.0004043 | 0.0013402 | 0.7638551 | S_LDL_PL_pct | MDD |
| Horizontal_pleiotropy | -0.000343 | 0.001141 | 0.7643883 | L_VLDL_PL | MDD |
| Horizontal_pleiotropy | 0.0002811 | 0.0009364 | 0.7644586 | L_HDL_L | MDD |
| Horizontal_pleiotropy | 0.0003043 | 0.0010239 | 0.7669171 | S_LDL_TG | MDD |
| Horizontal_pleiotropy | -0.0006661 | 0.0022416 | 0.766943 | MDD | Pyruvate |
| Horizontal_pleiotropy | 0.0002647 | 0.0008913 | 0.7671616 | S_VLDL_TG_pct | MDD |
| Horizontal_pleiotropy | 0.000828 | 0.0028944 | 0.7753929 | MDD | M_HDL_TG |
| Horizontal_pleiotropy | -0.0002958 | 0.0010369 | 0.776061 | IDL_CE_pct | MDD |
| Horizontal_pleiotropy | -0.000398 | 0.0014105 | 0.7786256 | PUFA_pct | MDD |
| Horizontal_pleiotropy | -0.0002804 | 0.001044 | 0.788802 | XS_VLDL_C_pct | MDD |
| Horizontal_pleiotropy | 0.0002771 | 0.0010341 | 0.7893431 | XXL_VLDL_P | MDD |
| Horizontal_pleiotropy | -0.0002327 | 0.0009361 | 0.8041136 | XL_HDL_C | MDD |
| Horizontal_pleiotropy | -0.0002379 | 0.0009685 | 0.8065383 | S_VLDL_PL_pct | MDD |
| Horizontal_pleiotropy | -0.0002759 | 0.00114 | 0.8094601 | IDL_L | MDD |
| Horizontal_pleiotropy | -0.0006714 | 0.0027948 | 0.8106243 | MDD | S_VLDL_TG_pct |
| Horizontal_pleiotropy | -0.0006094 | 0.0027192 | 0.8231028 | MDD | S_HDL_C_pct |
| Horizontal_pleiotropy | 0.000209 | 0.0010622 | 0.8444077 | M_LDL_TG | MDD |
| Horizontal_pleiotropy | 0.0001843 | 0.0009539 | 0.8470625 | L_HDL_PL | MDD |
| Horizontal_pleiotropy | 0.0001786 | 0.0009731 | 0.85477 | XXL_VLDL_TG | MDD |
| Horizontal_pleiotropy | -0.0002093 | 0.0011411 | 0.8550011 | XS_VLDL_FC_pct | MDD |
| Horizontal_pleiotropy | -0.0001988 | 0.001131 | 0.8608733 | L_VLDL_P | MDD |
| Horizontal_pleiotropy | 0.0001907 | 0.0010911 | 0.8616189 | M_VLDL_TG | MDD |
| Horizontal_pleiotropy | 0.0001829 | 0.0010899 | 0.8670396 | M_HDL_PL_pct | MDD |
| Horizontal_pleiotropy | -0.0001685 | 0.0010317 | 0.870589 | IDL_C_pct | MDD |
| Horizontal_pleiotropy | -0.0001788 | 0.0011548 | 0.8773357 | XS_VLDL_PL_pct | MDD |
| Horizontal_pleiotropy | 0.0001484 | 0.0009902 | 0.8810911 | XL_HDL_TG_pct | MDD |
| Horizontal_pleiotropy | -0.0003778 | 0.0026393 | 0.8864468 | MDD | L_LDL_TG_pct |
| Horizontal_pleiotropy | -0.0001516 | 0.0010895 | 0.8895901 | HDL_FC | MDD |
| Horizontal_pleiotropy | -0.000379 | 0.0028884 | 0.8958429 | MDD | XS_VLDL_C_pct |
| Horizontal_pleiotropy | 0.0001332 | 0.0010898 | 0.9029647 | M_HDL_CE_pct | MDD |
| Horizontal_pleiotropy | 0.0003008 | 0.002492 | 0.9041412 | MDD | IDL_CE_pct |
| Horizontal_pleiotropy | -0.0003276 | 0.0027762 | 0.9062976 | MDD | S_LDL_PL_pct |
| Horizontal_pleiotropy | -0.0001066 | 0.0009161 | 0.90759 | XS_VLDL_TG | MDD |

| | | | | | |
|---|---|---|---|---|---|
| Horizontal_pleiotropy | 0.0001052 | 0.0009998 | 0.9163882 | XXL_VLDL_C | MDD |
| Horizontal_pleiotropy | -0.0002585 | 0.0027195 | 0.924442 | MDD | Sphingomyelins |
| Horizontal_pleiotropy | 0.0000985 | 0.0010792 | 0.9274978 | IDL_CE | MDD |
| Horizontal_pleiotropy | -0.0001258 | 0.0014095 | 0.929112 | Total_P | MDD |
| Horizontal_pleiotropy | 0.0002199 | 0.0027195 | 0.9357026 | MDD | IDL_C_pct |
| Horizontal_pleiotropy | -0.0002361 | 0.0029416 | 0.9361674 | MDD | XS_VLDL_TG_pct |
| Horizontal_pleiotropy | -0.0002211 | 0.0029172 | 0.9397223 | MDD | S_VLDL_FC_pct |
| Horizontal_pleiotropy | -0.000083 | 0.0011101 | 0.9405533 | XXL_VLDL_CE | MDD |
| Horizontal_pleiotropy | 0.000071 | 0.0009815 | 0.9424923 | S_VLDL_C_pct | MDD |
| Horizontal_pleiotropy | 0.0000706 | 0.0010377 | 0.9458793 | XS_VLDL_CE_pct | MDD |
| Horizontal_pleiotropy | -0.0001953 | 0.0028791 | 0.9460461 | MDD | S_VLDL_PL_pct |
| Horizontal_pleiotropy | 0.0000699 | 0.0010394 | 0.9464983 | M_HDL_C_pct | MDD |
| Horizontal_pleiotropy | 0.000061 | 0.0009592 | 0.9493856 | L_HDL_CE | MDD |
| Horizontal_pleiotropy | -0.0000634 | 0.0010134 | 0.9502024 | HDL_size | MDD |
| Horizontal_pleiotropy | -0.0001212 | 0.0028082 | 0.9656536 | MDD | IDL_TG_pct |
| Horizontal_pleiotropy | 0.0000399 | 0.0010236 | 0.9689447 | XL_VLDL_P | MDD |
| Horizontal_pleiotropy | -0.0000379 | 0.0010765 | 0.9720063 | IDL_C | MDD |
| Horizontal_pleiotropy | -0.0000613 | 0.0024997 | 0.9804774 | MDD | Total_CE |
| Horizontal_pleiotropy | -0.0000165 | 0.0009904 | 0.9867707 | L_HDL_TG_pct | MDD |
| Horizontal_pleiotropy | -0.0000294 | 0.0028805 | 0.9918812 | MDD | S_LDL_TG_pct |
| Horizontal_pleiotropy | 0.0000096 | 0.0014141 | 0.9945819 | LA_pct | MDD |

**Supplementary Table 8**

**Results of Heterogeneity test in Mendelian Randomization**

| method | Q | Q_df | Q_pval | analysis | exposure | outcome |
|---|---|---|---|---|---|---|
| MR Egger | 201.2925 | 102 | 0 | Heterogeneity_tests | HDL_L | MDD |
| Inverse variance weighted | 201.4802 | 103 | 0 | Heterogeneity_tests | HDL_L | MDD |
| MR Egger | 199.042 | 93 | 0 | Heterogeneity_tests | HDL_PL | MDD |
| Inverse variance weighted | 201.6829 | 94 | 0 | Heterogeneity_tests | HDL_PL | MDD |
| MR Egger | 214.9488 | 106 | 0 | Heterogeneity_tests | IDL_TG_pct | MDD |
| Inverse variance weighted | 216.9665 | 107 | 0 | Heterogeneity_tests | IDL_TG_pct | MDD |
| MR Egger | 262.3141 | 145 | 0 | Heterogeneity_tests | L_HDL_CE | MDD |
| Inverse variance weighted | 262.3214 | 146 | 0 | Heterogeneity_tests | L_HDL_CE | MDD |
| MR Egger | 210.1844 | 106 | 0 | Heterogeneity_tests | MDD | HDL_C |
| Inverse variance weighted | 221.1552 | 107 | 0 | Heterogeneity_tests | MDD | HDL_C |
| MR Egger | 207.2396 | 106 | 0 | Heterogeneity_tests | MDD | HDL_CE |
| Inverse variance weighted | 218.7706 | 107 | 0 | Heterogeneity_tests | MDD | HDL_CE |
| MR Egger | 210.7846 | 106 | 0 | Heterogeneity_tests | MDD | HDL_FC |
| Inverse variance weighted | 219.3175 | 107 | 0 | Heterogeneity_tests | MDD | HDL_FC |
| Inverse variance weighted | 208.3633 | 107 | 0 | Heterogeneity_tests | MDD | HDL_L |
| MR Egger | 218.6184 | 106 | 0 | Heterogeneity_tests | MDD | HDL_size |
| Inverse variance weighted | 230.4838 | 107 | 0 | Heterogeneity_tests | MDD | HDL_size |
| MR Egger | 235.7415 | 106 | 0 | Heterogeneity_tests | MDD | IDL_FC_pct |
| Inverse variance weighted | 236.2695 | 107 | 0 | Heterogeneity_tests | MDD | IDL_FC_pct |
| MR Egger | 207.5323 | 106 | 0 | Heterogeneity_tests | MDD | IDL_TG_pct |
| Inverse variance weighted | 207.5359 | 107 | 0 | Heterogeneity_tests | MDD | IDL_TG_pct |
| MR Egger | 384.1792 | 106 | 0 | Heterogeneity_tests | MDD | LA_pct |
| Inverse variance weighted | 394.8818 | 107 | 0 | Heterogeneity_tests | MDD | LA_pct |
| MR Egger | 230.4705 | 106 | 0 | Heterogeneity_tests | MDD | L_HDL_C |
| Inverse variance weighted | 243.1716 | 107 | 0 | Heterogeneity_tests | MDD | L_HDL_C |
| MR Egger | 231.2864 | 106 | 0 | Heterogeneity_tests | MDD | L_HDL_CE |
| Inverse variance weighted | 244.4723 | 107 | 0 | Heterogeneity_tests | MDD | L_HDL_CE |
| MR Egger | 223.9831 | 106 | 0 | Heterogeneity_tests | MDD | L_HDL_CE_pct |
| Inverse variance weighted | 228.8927 | 107 | 0 | Heterogeneity_tests | MDD | L_HDL_CE_pct |
| MR Egger | 242.7513 | 106 | 0 | Heterogeneity_tests | MDD | L_HDL_C_pct |
| Inverse variance weighted | 247.7452 | 107 | 0 | Heterogeneity_tests | MDD | L_HDL_C_pct |
| MR Egger | 223.9349 | 106 | 0 | Heterogeneity_tests | MDD | L_HDL_FC |
| Inverse variance weighted | 234.6476 | 107 | 0 | Heterogeneity_tests | MDD | L_HDL_FC |
| MR Egger | 228.5701 | 106 | 0 | Heterogeneity_tests | MDD | L_HDL_FC_pct |
| Inverse variance weighted | 229.064 | 107 | 0 | Heterogeneity_tests | MDD | L_HDL_FC_pct |
| MR Egger | 219.1895 | 106 | 0 | Heterogeneity_tests | MDD | L_HDL_L |
| Inverse variance weighted | 231.9907 | 107 | 0 | Heterogeneity_tests | MDD | L_HDL_L |
| MR Egger | 228.5429 | 106 | 0 | Heterogeneity_tests | MDD | L_HDL_P |
| Inverse variance weighted | 240.7348 | 107 | 0 | Heterogeneity_tests | MDD | L_HDL_P |
| MR Egger | 208.0086 | 106 | 0 | Heterogeneity_tests | MDD | L_HDL_PL |
| Inverse variance weighted | 220.6451 | 107 | 0 | Heterogeneity_tests | MDD | L_HDL_PL |
| MR Egger | 226.2845 | 106 | 0 | Heterogeneity_tests | MDD | L_HDL_PL_pct |
| Inverse variance weighted | 228.4998 | 107 | 0 | Heterogeneity_tests | MDD | L_HDL_PL_pct |
| MR Egger | 203.2759 | 106 | 0 | Heterogeneity_tests | MDD | L_HDL_TG_pct |

| | | | | | | |
|---|---|---|---|---|---|---|
| Inverse variance weighted | 209.3314 | 107 | 0 | Heterogeneity_tests | MDD | L_HDL_TG_pct |
| Inverse variance weighted | 205.6039 | 107 | 0 | Heterogeneity_tests | MDD | L_VLDL_CE_pct |
| MR Egger | 215.5218 | 106 | 0 | Heterogeneity_tests | MDD | M_HDL_C_pct |
| Inverse variance weighted | 221.5035 | 107 | 0 | Heterogeneity_tests | MDD | M_HDL_C_pct |
| MR Egger | 238.9084 | 106 | 0 | Heterogeneity_tests | MDD | M_HDL_FC_pct |
| Inverse variance weighted | 244.7509 | 107 | 0 | Heterogeneity_tests | MDD | M_HDL_FC_pct |
| MR Egger | 225.6651 | 106 | 0 | Heterogeneity_tests | MDD | M_HDL_PL_pct |
| Inverse variance weighted | 231.6522 | 107 | 0 | Heterogeneity_tests | MDD | M_HDL_PL_pct |
| MR Egger | 210.9956 | 106 | 0 | Heterogeneity_tests | MDD | M_HDL_TG |
| Inverse variance weighted | 211.1585 | 107 | 0 | Heterogeneity_tests | MDD | M_HDL_TG |
| MR Egger | 352.3428 | 106 | 0 | Heterogeneity_tests | MDD | MUFA_pct |
| Inverse variance weighted | 355.0576 | 107 | 0 | Heterogeneity_tests | MDD | MUFA_pct |
| MR Egger | 237.5876 | 106 | 0 | Heterogeneity_tests | MDD | M_VLDL_CE_pct |
| Inverse variance weighted | 239.5778 | 107 | 0 | Heterogeneity_tests | MDD | M_VLDL_CE_pct |
| MR Egger | 238.1782 | 106 | 0 | Heterogeneity_tests | MDD | M_VLDL_C_pct |
| Inverse variance weighted | 239.7165 | 107 | 0 | Heterogeneity_tests | MDD | M_VLDL_C_pct |
| MR Egger | 234.2096 | 106 | 0 | Heterogeneity_tests | MDD | M_VLDL_FC_pct |
| Inverse variance weighted | 234.7689 | 107 | 0 | Heterogeneity_tests | MDD | M_VLDL_FC_pct |
| MR Egger | 231.4116 | 106 | 0 | Heterogeneity_tests | MDD | M_VLDL_TG_pct |
| Inverse variance weighted | 232.5028 | 107 | 0 | Heterogeneity_tests | MDD | M_VLDL_TG_pct |
| Inverse variance weighted | 207.8016 | 107 | 0 | Heterogeneity_tests | MDD | Omega_6_pct |
| MR Egger | 319.5948 | 106 | 0 | Heterogeneity_tests | MDD | PUFA_by_MUFA |
| Inverse variance weighted | 324.3381 | 107 | 0 | Heterogeneity_tests | MDD | PUFA_by_MUFA |
| MR Egger | 263.2593 | 106 | 0 | Heterogeneity_tests | MDD | PUFA_pct |
| Inverse variance weighted | 270.8986 | 107 | 0 | Heterogeneity_tests | MDD | PUFA_pct |
| MR Egger | 222.7096 | 106 | 0 | Heterogeneity_tests | MDD | S_HDL_TG |
| Inverse variance weighted | 225.6578 | 107 | 0 | Heterogeneity_tests | MDD | S_HDL_TG |
| MR Egger | 217.1116 | 106 | 0 | Heterogeneity_tests | MDD | S_LDL_TG_pct |
| Inverse variance weighted | 217.1118 | 107 | 0 | Heterogeneity_tests | MDD | S_LDL_TG_pct |
| MR Egger | 229.2401 | 106 | 0 | Heterogeneity_tests | MDD | S_VLDL_FC_pct |
| Inverse variance weighted | 229.2525 | 107 | 0 | Heterogeneity_tests | MDD | S_VLDL_FC_pct |
| MR Egger | 222.3953 | 106 | 0 | Heterogeneity_tests | MDD | S_VLDL_PL_pct |
| Inverse variance weighted | 222.405 | 107 | 0 | Heterogeneity_tests | MDD | S_VLDL_PL_pct |
| MR Egger | 205.0128 | 106 | 0 | Heterogeneity_tests | MDD | S_VLDL_TG_pct |
| Inverse variance weighted | 205.1244 | 107 | 0 | Heterogeneity_tests | MDD | S_VLDL_TG_pct |
| MR Egger | 207.9514 | 106 | 0 | Heterogeneity_tests | MDD | Sphingomyelins |
| Inverse variance weighted | 207.9691 | 107 | 0 | Heterogeneity_tests | MDD | Sphingomyelins |
| Inverse variance weighted | 210.8003 | 107 | 0 | Heterogeneity_tests | MDD | TG_by_PG |
| MR Egger | 660.2308 | 106 | 0 | Heterogeneity_tests | MDD | Unsaturation |
| Inverse variance weighted | 665.4027 | 107 | 0 | Heterogeneity_tests | MDD | Unsaturation |
| Inverse variance weighted | 208.7638 | 107 | 0 | Heterogeneity_tests | MDD | VLDL_size |
| MR Egger | 248.8841 | 106 | 0 | Heterogeneity_tests | MDD | XL_HDL_C |
| Inverse variance weighted | 254.4829 | 107 | 0 | Heterogeneity_tests | MDD | XL_HDL_C |
| MR Egger | 248.6384 | 106 | 0 | Heterogeneity_tests | MDD | XL_HDL_CE |
| Inverse variance weighted | 255.6624 | 107 | 0 | Heterogeneity_tests | MDD | XL_HDL_CE |
| MR Egger | 240.3393 | 106 | 0 | Heterogeneity_tests | MDD | XL_HDL_FC |
| Inverse variance weighted | 241.8846 | 107 | 0 | Heterogeneity_tests | MDD | XL_HDL_FC |

| | | | | | | |
|---|---|---|---|---|---|---|
| Inverse variance weighted | 211.0606 | 107 | 0 | Heterogeneity_tests | MDD | XL_HDL_FC_pct |
| MR Egger | 234.3353 | 106 | 0 | Heterogeneity_tests | MDD | XL_HDL_L |
| Inverse variance weighted | 240.4924 | 107 | 0 | Heterogeneity_tests | MDD | XL_HDL_L |
| MR Egger | 230.9444 | 106 | 0 | Heterogeneity_tests | MDD | XL_HDL_P |
| Inverse variance weighted | 235.7546 | 107 | 0 | Heterogeneity_tests | MDD | XL_HDL_P |
| MR Egger | 225.3077 | 106 | 0 | Heterogeneity_tests | MDD | XL_HDL_PL |
| Inverse variance weighted | 232.1664 | 107 | 0 | Heterogeneity_tests | MDD | XL_HDL_PL |
| MR Egger | 216.3487 | 106 | 0 | Heterogeneity_tests | MDD | XL_HDL_TG_pct |
| Inverse variance weighted | 221.4117 | 107 | 0 | Heterogeneity_tests | MDD | XL_HDL_TG_pct |
| MR Egger | 213.6515 | 106 | 0 | Heterogeneity_tests | MDD | XL_VLDL_CE_pct |
| Inverse variance weighted | 216.4773 | 107 | 0 | Heterogeneity_tests | MDD | XL_VLDL_CE_pct |
| MR Egger | 218.7833 | 106 | 0 | Heterogeneity_tests | MDD | XL_VLDL_C_pct |
| Inverse variance weighted | 222.0245 | 107 | 0 | Heterogeneity_tests | MDD | XL_VLDL_C_pct |
| MR Egger | 214.4759 | 106 | 0 | Heterogeneity_tests | MDD | XL_VLDL_FC_pct |
| Inverse variance weighted | 217.8866 | 107 | 0 | Heterogeneity_tests | MDD | XL_VLDL_FC_pct |
| MR Egger | 203.1492 | 106 | 0 | Heterogeneity_tests | MDD | XL_VLDL_TG |
| Inverse variance weighted | 211.9012 | 107 | 0 | Heterogeneity_tests | MDD | XL_VLDL_TG |
| MR Egger | 203.2787 | 106 | 0 | Heterogeneity_tests | MDD | XL_VLDL_TG_pct |
| Inverse variance weighted | 205.1213 | 107 | 0 | Heterogeneity_tests | MDD | XL_VLDL_TG_pct |
| MR Egger | 221.998 | 106 | 0 | Heterogeneity_tests | MDD | XS_VLDL_CE_pct |
| Inverse variance weighted | 222.2152 | 107 | 0 | Heterogeneity_tests | MDD | XS_VLDL_CE_pct |
| MR Egger | 228.1816 | 106 | 0 | Heterogeneity_tests | MDD | XS_VLDL_C_pct |
| Inverse variance weighted | 228.2187 | 107 | 0 | Heterogeneity_tests | MDD | XS_VLDL_C_pct |
| MR Egger | 209.1171 | 106 | 0 | Heterogeneity_tests | MDD | XS_VLDL_FC_pct |
| Inverse variance weighted | 211.2177 | 107 | 0 | Heterogeneity_tests | MDD | XS_VLDL_FC_pct |
| MR Egger | 235.5313 | 106 | 0 | Heterogeneity_tests | MDD | XS_VLDL_TG_pct |
| Inverse variance weighted | 235.5456 | 107 | 0 | Heterogeneity_tests | MDD | XS_VLDL_TG_pct |
| Inverse variance weighted | 204.1011 | 107 | 0 | Heterogeneity_tests | MDD | XXL_VLDL_C |
| Inverse variance weighted | 204.8994 | 107 | 0 | Heterogeneity_tests | MDD | XXL_VLDL_FC |
| Inverse variance weighted | 204.7859 | 107 | 0 | Heterogeneity_tests | MDD | XXL_VLDL_L |
| Inverse variance weighted | 204.2841 | 107 | 0 | Heterogeneity_tests | MDD | XXL_VLDL_P |
| Inverse variance weighted | 204.1907 | 107 | 0 | Heterogeneity_tests | MDD | XXL_VLDL_PL |
| Inverse variance weighted | 204.8323 | 107 | 0 | Heterogeneity_tests | MDD | XXL_VLDL_TG |
| MR Egger | 226.5198 | 110 | 0 | Heterogeneity_tests | M_HDL_CE | MDD |
| Inverse variance weighted | 228.7988 | 111 | 0 | Heterogeneity_tests | M_HDL_CE | MDD |
| MR Egger | 189.9735 | 95 | 0 | Heterogeneity_tests | M_HDL_FC | MDD |
| Inverse variance weighted | 190.2243 | 96 | 0 | Heterogeneity_tests | M_HDL_FC | MDD |
| MR Egger | 188.5988 | 95 | 0 | Heterogeneity_tests | M_HDL_P | MDD |
| Inverse variance weighted | 191.0237 | 96 | 0 | Heterogeneity_tests | M_HDL_P | MDD |
| MR Egger | 179.2346 | 90 | 0.0000001 | Heterogeneity_tests | ApoA1 | MDD |
| Inverse variance weighted | 180.0527 | 91 | 0.0000001 | Heterogeneity_tests | ApoA1 | MDD |
| MR Egger | 224.254 | 125 | 0.0000001 | Heterogeneity_tests | HDL_size | MDD |
| MR Egger | 252.9037 | 145 | 0.0000001 | Heterogeneity_tests | L_HDL_C | MDD |
| Inverse variance weighted | 253.176 | 146 | 0.0000001 | Heterogeneity_tests | L_HDL_C | MDD |
| MR Egger | 248.2768 | 141 | 0.0000001 | Heterogeneity_tests | L_HDL_L | MDD |
| Inverse variance weighted | 248.4355 | 142 | 0.0000001 | Heterogeneity_tests | L_HDL_L | MDD |
| MR Egger | 205.8582 | 111 | 0.0000001 | Heterogeneity_tests | L_HDL_PL_pct | MDD |

| | | | | | | |
|---|---|---|---|---|---|---|
| Inverse variance weighted | 200.9537 | 107 | 0.0000001 | Heterogeneity_tests | MDD | ApoA1 |
| MR Egger | 198.8578 | 106 | 0.0000001 | Heterogeneity_tests | MDD | HDL_L |
| Inverse variance weighted | 200.7227 | 107 | 0.0000001 | Heterogeneity_tests | MDD | HDL_PL |
| MR Egger | 201.4649 | 106 | 0.0000001 | Heterogeneity_tests | MDD | L_VLDL_CE_pct |
| MR Egger | 198.4867 | 106 | 0.0000001 | Heterogeneity_tests | MDD | M_HDL_CE_pct |
| Inverse variance weighted | 203.0335 | 107 | 0.0000001 | Heterogeneity_tests | MDD | M_HDL_CE_pct |
| Inverse variance weighted | 202.8151 | 107 | 0.0000001 | Heterogeneity_tests | MDD | M_HDL_FC |
| Inverse variance weighted | 202.8205 | 107 | 0.0000001 | Heterogeneity_tests | MDD | M_HDL_TG_pct |
| MR Egger | 201.8232 | 106 | 0.0000001 | Heterogeneity_tests | MDD | Omega_6_pct |
| MR Egger | 199.7033 | 106 | 0.0000001 | Heterogeneity_tests | MDD | S_HDL_FC_pct |
| Inverse variance weighted | 200.7929 | 107 | 0.0000001 | Heterogeneity_tests | MDD | S_HDL_FC_pct |
| Inverse variance weighted | 202.252 | 107 | 0.0000001 | Heterogeneity_tests | MDD | S_HDL_TG_pct |
| MR Egger | 201.3544 | 106 | 0.0000001 | Heterogeneity_tests | MDD | TG_by_PG |
| MR Egger | 200.1753 | 106 | 0.0000001 | Heterogeneity_tests | MDD | VLDL_size |
| MR Egger | 200.3421 | 106 | 0.0000001 | Heterogeneity_tests | MDD | XXL_VLDL_C |
| Inverse variance weighted | 201.9046 | 107 | 0.0000001 | Heterogeneity_tests | MDD | XXL_VLDL_CE |
| MR Egger | 201.309 | 106 | 0.0000001 | Heterogeneity_tests | MDD | XXL_VLDL_FC |
| MR Egger | 199.0081 | 106 | 0.0000001 | Heterogeneity_tests | MDD | XXL_VLDL_L |
| MR Egger | 199.2341 | 106 | 0.0000001 | Heterogeneity_tests | MDD | XXL_VLDL_P |
| MR Egger | 199.7534 | 106 | 0.0000001 | Heterogeneity_tests | MDD | XXL_VLDL_PL |
| MR Egger | 198.2372 | 106 | 0.0000001 | Heterogeneity_tests | MDD | XXL_VLDL_TG |
| MR Egger | 193.8073 | 102 | 0.0000001 | Heterogeneity_tests | M_HDL_C | MDD |
| Inverse variance weighted | 195.7396 | 103 | 0.0000001 | Heterogeneity_tests | M_HDL_C | MDD |
| MR Egger | 219.7198 | 119 | 0.0000001 | Heterogeneity_tests | M_HDL_TG_pct | MDD |
| Inverse variance weighted | 220.1964 | 120 | 0.0000001 | Heterogeneity_tests | M_HDL_TG_pct | MDD |
| Inverse variance weighted | 194.8952 | 103 | 0.0000001 | Heterogeneity_tests | XL_VLDL_CE_pct | MDD |
| MR Egger | 170.9156 | 87 | 0.0000002 | Heterogeneity_tests | HDL_P | MDD |
| Inverse variance weighted | 224.261 | 126 | 0.0000002 | Heterogeneity_tests | HDL_size | MDD |
| MR Egger | 236.866 | 136 | 0.0000002 | Heterogeneity_tests | L_HDL_FC | MDD |
| Inverse variance weighted | 237.7203 | 137 | 0.0000002 | Heterogeneity_tests | L_HDL_FC | MDD |
| Inverse variance weighted | 178.9754 | 92 | 0.0000002 | Heterogeneity_tests | L_HDL_FC_pct | MDD |
| Inverse variance weighted | 206.0338 | 112 | 0.0000002 | Heterogeneity_tests | L_HDL_PL_pct | MDD |
| MR Egger | 245.6161 | 143 | 0.0000002 | Heterogeneity_tests | L_HDL_P | MDD |
| Inverse variance weighted | 246.2729 | 144 | 0.0000002 | Heterogeneity_tests | L_HDL_P | MDD |
| Inverse variance weighted | 197.6529 | 107 | 0.0000002 | Heterogeneity_tests | MDD | M_HDL_P |
| MR Egger | 198.0732 | 106 | 0.0000002 | Heterogeneity_tests | MDD | M_HDL_TG_pct |
| MR Egger | 196.7294 | 106 | 0.0000002 | Heterogeneity_tests | MDD | MUFA |
| Inverse variance weighted | 198.2711 | 107 | 0.0000002 | Heterogeneity_tests | MDD | MUFA |
| MR Egger | 197.696 | 106 | 0.0000002 | Heterogeneity_tests | MDD | S_HDL_TG_pct |
| MR Egger | 196.2483 | 106 | 0.0000002 | Heterogeneity_tests | MDD | XL_HDL_FC_pct |
| MR Egger | 197.786 | 106 | 0.0000002 | Heterogeneity_tests | MDD | XXL_VLDL_CE |
| MR Egger | 218.4015 | 122 | 0.0000002 | Heterogeneity_tests | XL_HDL_FC_pct | MDD |
| Inverse variance weighted | 220.4492 | 123 | 0.0000002 | Heterogeneity_tests | XL_HDL_FC_pct | MDD |
| MR Egger | 190.8252 | 102 | 0.0000002 | Heterogeneity_tests | XL_VLDL_CE_pct | MDD |
| Inverse variance weighted | 171.2451 | 88 | 0.0000003 | Heterogeneity_tests | HDL_P | MDD |
| MR Egger | 175.2863 | 91 | 0.0000003 | Heterogeneity_tests | L_HDL_FC_pct | MDD |
| Inverse variance weighted | 196.5948 | 107 | 0.0000003 | Heterogeneity_tests | MDD | HDL_P |

| | | | | | | |
|---|---|---|---|---|---|---|
| Inverse variance weighted | 196.5187 | 107 | 0.0000003 | Heterogeneity_tests | MDD | M_HDL_C |
| MR Egger | 194.5872 | 106 | 0.0000003 | Heterogeneity_tests | MDD | M_HDL_FC |
| MR Egger | 195.8493 | 106 | 0.0000003 | Heterogeneity_tests | MDD | S_VLDL_C_pct |
| Inverse variance weighted | 196.1143 | 107 | 0.0000003 | Heterogeneity_tests | MDD | S_VLDL_C_pct |
| Inverse variance weighted | 197.3065 | 107 | 0.0000003 | Heterogeneity_tests | MDD | VLDL_TG |
| Inverse variance weighted | 197.3122 | 107 | 0.0000003 | Heterogeneity_tests | MDD | XL_VLDL_L |
| MR Egger | 207.8637 | 115 | 0.0000003 | Heterogeneity_tests | M_HDL_CE_pct | MDD |
| Inverse variance weighted | 207.8907 | 116 | 0.0000003 | Heterogeneity_tests | M_HDL_CE_pct | MDD |
| Inverse variance weighted | 194.9265 | 107 | 0.0000004 | Heterogeneity_tests | MDD | Total_TG |
| Inverse variance weighted | 195.3151 | 107 | 0.0000004 | Heterogeneity_tests | MDD | XL_VLDL_P |
| MR Egger | 193.4717 | 106 | 0.0000005 | Heterogeneity_tests | MDD | ApoA1 |
| MR Egger | 193.4594 | 106 | 0.0000005 | Heterogeneity_tests | MDD | HDL_TG |
| Inverse variance weighted | 194.5269 | 107 | 0.0000005 | Heterogeneity_tests | MDD | Total_P |
| MR Egger | 191.9656 | 106 | 0.0000006 | Heterogeneity_tests | MDD | HDL_P |
| MR Egger | 191.9628 | 106 | 0.0000006 | Heterogeneity_tests | MDD | HDL_PL |
| Inverse variance weighted | 193.6854 | 107 | 0.0000006 | Heterogeneity_tests | MDD | HDL_TG |
| Inverse variance weighted | 193.9882 | 107 | 0.0000006 | Heterogeneity_tests | MDD | M_HDL_CE |
| MR Egger | 191.377 | 106 | 0.0000007 | Heterogeneity_tests | MDD | Total_P |
| MR Egger | 190.5892 | 106 | 0.0000009 | Heterogeneity_tests | MDD | IDL_C_pct |
| MR Egger | 190.5744 | 106 | 0.0000009 | Heterogeneity_tests | MDD | VLDL_TG |
| MR Egger | 190.1527 | 106 | 0.000001 | Heterogeneity_tests | MDD | S_LDL_PL_pct |
| MR Egger | 189.8453 | 106 | 0.0000011 | Heterogeneity_tests | MDD | Total_TG |
| Inverse variance weighted | 190.601 | 107 | 0.0000012 | Heterogeneity_tests | MDD | IDL_C_pct |
| MR Egger | 189.3647 | 106 | 0.0000012 | Heterogeneity_tests | MDD | M_HDL_P |
| MR Egger | 194.7631 | 110 | 0.0000012 | Heterogeneity_tests | M_HDL_FC_pct | MDD |
| Inverse variance weighted | 190.1777 | 107 | 0.0000013 | Heterogeneity_tests | MDD | S_LDL_PL_pct |
| MR Egger | 188.7555 | 106 | 0.0000014 | Heterogeneity_tests | MDD | XL_VLDL_L |
| Inverse variance weighted | 194.9434 | 111 | 0.0000015 | Heterogeneity_tests | M_HDL_FC_pct | MDD |
| Inverse variance weighted | 189.462 | 107 | 0.0000016 | Heterogeneity_tests | MDD | L_VLDL_TG |
| Inverse variance weighted | 189.1552 | 107 | 0.0000017 | Heterogeneity_tests | MDD | XL_VLDL_PL |
| MR Egger | 196.6594 | 113 | 0.0000018 | Heterogeneity_tests | XL_HDL_PL | MDD |
| MR Egger | 187.3329 | 106 | 0.0000019 | Heterogeneity_tests | MDD | XL_VLDL_P |
| MR Egger | 187.0558 | 106 | 0.000002 | Heterogeneity_tests | MDD | M_HDL_C |
| Inverse variance weighted | 197.6412 | 114 | 0.000002 | Heterogeneity_tests | XL_HDL_PL | MDD |
| MR Egger | 190.5283 | 109 | 0.0000022 | Heterogeneity_tests | HDL_FC | MDD |
| MR Egger | 185.1815 | 105 | 0.0000023 | Heterogeneity_tests | XS_VLDL_CE_pct | MDD |
| MR Egger | 186.0057 | 106 | 0.0000026 | Heterogeneity_tests | MDD | S_HDL_C_pct |
| MR Egger | 204.1568 | 120 | 0.0000026 | Heterogeneity_tests | M_HDL_PL_pct | MDD |
| MR Egger | 216.8222 | 130 | 0.0000027 | Heterogeneity_tests | L_HDL_PL | MDD |
| MR Egger | 207.7632 | 123 | 0.0000028 | Heterogeneity_tests | TG_by_PG | MDD |
| Inverse variance weighted | 117.4018 | 56 | 0.000003 | Heterogeneity_tests | Ala | MDD |
| Inverse variance weighted | 190.5622 | 110 | 0.000003 | Heterogeneity_tests | HDL_FC | MDD |
| Inverse variance weighted | 186.4075 | 107 | 0.0000031 | Heterogeneity_tests | MDD | L_VLDL_PL |
| Inverse variance weighted | 185.1897 | 106 | 0.0000031 | Heterogeneity_tests | XS_VLDL_CE_pct | MDD |
| Inverse variance weighted | 208.1425 | 124 | 0.0000033 | Heterogeneity_tests | TG_by_PG | MDD |
| MR Egger | 115.5326 | 55 | 0.0000034 | Heterogeneity_tests | Ala | MDD |
| Inverse variance weighted | 186.0938 | 107 | 0.0000034 | Heterogeneity_tests | MDD | S_HDL_C_pct |

| | | | | | | |
|---|---|---|---|---|---|---|
| Inverse variance weighted | 204.2047 | 121 | 0.0000034 | Heterogeneity_tests | M_HDL_PL_pct | MDD |
| Inverse variance weighted | 216.8845 | 131 | 0.0000035 | Heterogeneity_tests | L_HDL_PL | MDD |
| Inverse variance weighted | 185.9072 | 107 | 0.0000035 | Heterogeneity_tests | MDD | L_VLDL_PL_pct |
| MR Egger | 184.309 | 106 | 0.0000038 | Heterogeneity_tests | MDD | M_HDL_CE |
| Inverse variance weighted | 185.3979 | 107 | 0.0000039 | Heterogeneity_tests | MDD | S_VLDL_TG |
| MR Egger | 183.1547 | 106 | 0.0000049 | Heterogeneity_tests | MDD | S_VLDL_TG |
| MR Egger | 183.0552 | 106 | 0.000005 | Heterogeneity_tests | MDD | L_VLDL_PL_pct |
| MR Egger | 182.5227 | 106 | 0.0000056 | Heterogeneity_tests | MDD | IDL_FC |
| Inverse variance weighted | 177.7167 | 103 | 0.0000068 | Heterogeneity_tests | XL_HDL_P | MDD |
| Inverse variance weighted | 182.857 | 107 | 0.0000069 | Heterogeneity_tests | MDD | L_VLDL_L |
| MR Egger | 176.3057 | 102 | 0.000007 | Heterogeneity_tests | XL_HDL_P | MDD |
| Inverse variance weighted | 182.6962 | 107 | 0.0000072 | Heterogeneity_tests | MDD | IDL_FC |
| MR Egger | 183.4453 | 108 | 0.0000081 | Heterogeneity_tests | L_HDL_CE_pct | MDD |
| MR Egger | 180.573 | 106 | 0.0000086 | Heterogeneity_tests | MDD | XL_VLDL_PL |
| Inverse variance weighted | 181.5257 | 107 | 0.0000092 | Heterogeneity_tests | MDD | L_VLDL_P |
| Inverse variance weighted | 186.6848 | 111 | 0.0000092 | Heterogeneity_tests | XL_HDL_L | MDD |
| MR Egger | 185.2622 | 110 | 0.0000095 | Heterogeneity_tests | XL_HDL_L | MDD |
| Inverse variance weighted | 183.7957 | 109 | 0.0000099 | Heterogeneity_tests | L_HDL_CE_pct | MDD |
| MR Egger | 179.8713 | 106 | 0.00001 | Heterogeneity_tests | MDD | L_VLDL_TG |
| Inverse variance weighted | 180.8697 | 107 | 0.0000107 | Heterogeneity_tests | MDD | XL_VLDL_FC |
| MR Egger | 178.19 | 106 | 0.0000144 | Heterogeneity_tests | MDD | L_VLDL_PL |
| MR Egger | 197.2193 | 121 | 0.0000148 | Heterogeneity_tests | XL_HDL_C | MDD |
| MR Egger | 178.0027 | 106 | 0.000015 | Heterogeneity_tests | MDD | L_LDL_TG_pct |
| Inverse variance weighted | 173.9506 | 103 | 0.0000156 | Heterogeneity_tests | XL_VLDL_C_pct | MDD |
| MR Egger | 172.5675 | 102 | 0.0000159 | Heterogeneity_tests | XL_VLDL_C_pct | MDD |
| Inverse variance weighted | 197.32 | 122 | 0.0000187 | Heterogeneity_tests | XL_HDL_C | MDD |
| Inverse variance weighted | 178.0371 | 107 | 0.0000196 | Heterogeneity_tests | MDD | L_LDL_TG_pct |
| MR Egger | 171.5035 | 102 | 0.0000201 | Heterogeneity_tests | XS_VLDL_C_pct | MDD |
| Inverse variance weighted | 177.0602 | 107 | 0.0000241 | Heterogeneity_tests | MDD | M_VLDL_TG |
| MR Egger | 190.8669 | 118 | 0.0000249 | Heterogeneity_tests | XL_HDL_TG_pct | MDD |
| Inverse variance weighted | 171.6248 | 103 | 0.0000258 | Heterogeneity_tests | XS_VLDL_C_pct | MDD |
| Inverse variance weighted | 176.661 | 107 | 0.0000262 | Heterogeneity_tests | MDD | L_VLDL_FC |
| MR Egger | 192.9224 | 120 | 0.0000274 | Heterogeneity_tests | S_HDL_TG_pct | MDD |
| Inverse variance weighted | 190.9032 | 119 | 0.0000319 | Heterogeneity_tests | XL_HDL_TG_pct | MDD |
| Inverse variance weighted | 193.2505 | 121 | 0.000033 | Heterogeneity_tests | S_HDL_TG_pct | MDD |
| MR Egger | 174.1333 | 106 | 0.000034 | Heterogeneity_tests | MDD | L_VLDL_L |
| MR Egger | 173.7589 | 106 | 0.0000368 | Heterogeneity_tests | MDD | L_VLDL_P |
| MR Egger | 143.8299 | 83 | 0.0000395 | Heterogeneity_tests | Total_P | MDD |
| MR Egger | 173.0307 | 106 | 0.0000428 | Heterogeneity_tests | MDD | XL_VLDL_FC |
| Inverse variance weighted | 143.8437 | 84 | 0.0000527 | Heterogeneity_tests | Total_P | MDD |
| MR Egger | 171.7006 | 106 | 0.0000563 | Heterogeneity_tests | MDD | M_LDL_TG_pct |
| MR Egger | 147.2696 | 87 | 0.0000584 | Heterogeneity_tests | XL_VLDL_CE | MDD |
| MR Egger | 176.5544 | 110 | 0.0000586 | Heterogeneity_tests | VLDL_size | MDD |
| Inverse variance weighted | 148.3337 | 88 | 0.0000613 | Heterogeneity_tests | XL_VLDL_CE | MDD |
| MR Egger | 158.5322 | 96 | 0.0000621 | Heterogeneity_tests | L_VLDL_PL | MDD |
| Inverse variance weighted | 172.3716 | 107 | 0.0000636 | Heterogeneity_tests | MDD | M_LDL_TG_pct |
| Inverse variance weighted | 197.0839 | 127 | 0.0000672 | Heterogeneity_tests | HDL_CE | MDD |

| | | | | | | |
|---|---|---|---|---|---|---|
| MR Egger | 170.8289 | 106 | 0.0000673 | Heterogeneity_tests | MDD | M_VLDL_TG |
| MR Egger | 183.334 | 116 | 0.000068 | Heterogeneity_tests | S_HDL_TG | MDD |
| Inverse variance weighted | 176.739 | 111 | 0.0000727 | Heterogeneity_tests | VLDL_size | MDD |
| Inverse variance weighted | 183.8845 | 117 | 0.000078 | Heterogeneity_tests | S_HDL_TG | MDD |
| Inverse variance weighted | 158.6814 | 97 | 0.0000786 | Heterogeneity_tests | L_VLDL_PL | MDD |
| MR Egger | 153.5581 | 93 | 0.0000791 | Heterogeneity_tests | S_VLDL_L | MDD |
| MR Egger | 155.9823 | 95 | 0.0000813 | Heterogeneity_tests | L_VLDL_P | MDD |
| Inverse variance weighted | 148.2652 | 89 | 0.0000822 | Heterogeneity_tests | L_VLDL_C | MDD |
| MR Egger | 164.7877 | 102 | 0.0000824 | Heterogeneity_tests | IDL_C_pct | MDD |
| MR Egger | 194.6361 | 126 | 0.0000845 | Heterogeneity_tests | HDL_CE | MDD |
| MR Egger | 178.3531 | 113 | 0.0000867 | Heterogeneity_tests | L_HDL_TG_pct | MDD |
| MR Egger | 145.2057 | 87 | 0.0000915 | Heterogeneity_tests | PUFA_by_MUFA | MDD |
| MR Egger | 146.4466 | 88 | 0.0000923 | Heterogeneity_tests | L_VLDL_C | MDD |
| Inverse variance weighted | 153.9157 | 94 | 0.000096 | Heterogeneity_tests | S_VLDL_L | MDD |
| Inverse variance weighted | 170.2508 | 107 | 0.0000975 | Heterogeneity_tests | MDD | S_LDL_TG |
| Inverse variance weighted | 166.4637 | 104 | 0.0000979 | Heterogeneity_tests | S_LDL_TG_pct | MDD |
| MR Egger | 168.9391 | 106 | 0.0000986 | Heterogeneity_tests | MDD | IDL_C |
| Inverse variance weighted | 156.033 | 96 | 0.0001048 | Heterogeneity_tests | L_VLDL_P | MDD |
| Inverse variance weighted | 164.8308 | 103 | 0.0001056 | Heterogeneity_tests | IDL_C_pct | MDD |
| MR Egger | 168.5391 | 106 | 0.0001068 | Heterogeneity_tests | MDD | L_VLDL_FC |
| Inverse variance weighted | 145.646 | 88 | 0.0001095 | Heterogeneity_tests | PUFA_by_MUFA | MDD |
| Inverse variance weighted | 150.7398 | 92 | 0.0001096 | Heterogeneity_tests | HDL_TG | MDD |
| Inverse variance weighted | 178.3535 | 114 | 0.0001106 | Heterogeneity_tests | L_HDL_TG_pct | MDD |
| MR Egger | 151.8042 | 93 | 0.0001145 | Heterogeneity_tests | L_VLDL_FC | MDD |
| MR Egger | 164.403 | 103 | 0.0001151 | Heterogeneity_tests | S_LDL_TG_pct | MDD |
| Inverse variance weighted | 169.2026 | 107 | 0.0001201 | Heterogeneity_tests | MDD | IDL_C |
| MR Egger | 115.3412 | 65 | 0.0001214 | Heterogeneity_tests | LA_pct | MDD |
| Inverse variance weighted | 152.6594 | 94 | 0.0001247 | Heterogeneity_tests | L_VLDL_FC | MDD |
| MR Egger | 167.5129 | 106 | 0.000131 | Heterogeneity_tests | MDD | S_LDL_TG |
| MR Egger | 152.2761 | 94 | 0.000135 | Heterogeneity_tests | XXL_VLDL_CE | MDD |
| Inverse variance weighted | 168.4597 | 107 | 0.0001391 | Heterogeneity_tests | MDD | VLDL_L |
| MR Egger | 148.2113 | 91 | 0.0001427 | Heterogeneity_tests | HDL_TG | MDD |
| Inverse variance weighted | 115.3413 | 66 | 0.0001652 | Heterogeneity_tests | LA_pct | MDD |
| Inverse variance weighted | 139.8602 | 85 | 0.0001661 | Heterogeneity_tests | VLDL_FC | MDD |
| MR Egger | 138.3794 | 84 | 0.0001736 | Heterogeneity_tests | VLDL_FC | MDD |
| Inverse variance weighted | 152.2852 | 95 | 0.0001746 | Heterogeneity_tests | XXL_VLDL_CE | MDD |
| Inverse variance weighted | 142.6976 | 88 | 0.0002038 | Heterogeneity_tests | M_VLDL_FC_pct | MDD |
| MR Egger | 165.0257 | 106 | 0.0002134 | Heterogeneity_tests | MDD | Total_CE |
| Inverse variance weighted | 183.3461 | 121 | 0.0002206 | Heterogeneity_tests | HDL_C | MDD |
| MR Egger | 140.8686 | 87 | 0.0002291 | Heterogeneity_tests | M_VLDL_FC_pct | MDD |
| Inverse variance weighted | 165.8071 | 107 | 0.0002331 | Heterogeneity_tests | MDD | XL_VLDL_C |
| MR Egger | 164.4896 | 106 | 0.0002367 | Heterogeneity_tests | MDD | XS_VLDL_TG |
| MR Egger | 164.35 | 106 | 0.0002431 | Heterogeneity_tests | MDD | IDL_CE |
| MR Egger | 181.4919 | 120 | 0.0002477 | Heterogeneity_tests | HDL_C | MDD |
| MR Egger | 156.7372 | 100 | 0.0002508 | Heterogeneity_tests | XXL_VLDL_PL | MDD |
| Inverse variance weighted | 165.3655 | 107 | 0.0002537 | Heterogeneity_tests | MDD | XS_VLDL_TG |
| Inverse variance weighted | 157.8193 | 101 | 0.0002589 | Heterogeneity_tests | XXL_VLDL_PL | MDD |

| | | | | | | |
|---|---|---|---|---|---|---|
| Inverse variance weighted | 165.0267 | 107 | 0.0002707 | Heterogeneity_tests | MDD | Total_CE |
| Inverse variance weighted | 164.6486 | 107 | 0.000291 | Heterogeneity_tests | MDD | IDL_CE |
| MR Egger | 155.583 | 100 | 0.0003142 | Heterogeneity_tests | M_LDL_TG | MDD |
| MR Egger | 162.0544 | 106 | 0.0003766 | Heterogeneity_tests | MDD | VLDL_L |
| Inverse variance weighted | 124.5279 | 76 | 0.0003783 | Heterogeneity_tests | Total_CE | MDD |
| MR Egger | 171.7486 | 114 | 0.000385 | Heterogeneity_tests | M_HDL_C_pct | MDD |
| MR Egger | 149.5455 | 96 | 0.0003871 | Heterogeneity_tests | MUFA_pct | MDD |
| Inverse variance weighted | 155.6433 | 101 | 0.0003945 | Heterogeneity_tests | M_LDL_TG | MDD |
| MR Egger | 161.478 | 106 | 0.0004197 | Heterogeneity_tests | XL_HDL_FC | MDD |
| Inverse variance weighted | 162.409 | 107 | 0.0004437 | Heterogeneity_tests | XL_HDL_FC | MDD |
| MR Egger | 161.0962 | 106 | 0.0004508 | Heterogeneity_tests | MDD | IDL_L |
| Inverse variance weighted | 149.7387 | 97 | 0.0004743 | Heterogeneity_tests | MUFA_pct | MDD |
| Inverse variance weighted | 171.7554 | 115 | 0.0004802 | Heterogeneity_tests | M_HDL_C_pct | MDD |
| MR Egger | 158.1691 | 104 | 0.0004924 | Heterogeneity_tests | S_LDL_TG | MDD |
| MR Egger | 121.71 | 75 | 0.000525 | Heterogeneity_tests | Total_CE | MDD |
| Inverse variance weighted | 161.4142 | 107 | 0.0005336 | Heterogeneity_tests | MDD | IDL_L |
| Inverse variance weighted | 161.3163 | 107 | 0.0005433 | Heterogeneity_tests | MDD | L_VLDL_C |
| Inverse variance weighted | 138.9221 | 89 | 0.0005646 | Heterogeneity_tests | MUFA | MDD |
| MR Egger | 142.5384 | 92 | 0.0005767 | Heterogeneity_tests | IDL_CE_pct | MDD |
| Inverse variance weighted | 158.3034 | 105 | 0.0006032 | Heterogeneity_tests | S_LDL_TG | MDD |
| MR Egger | 136.5003 | 88 | 0.0007118 | Heterogeneity_tests | MUFA | MDD |
| Inverse variance weighted | 142.6645 | 93 | 0.0007145 | Heterogeneity_tests | IDL_CE_pct | MDD |
| MR Egger | 158.4556 | 106 | 0.0007338 | Heterogeneity_tests | MDD | IDL_CE_pct |
| MR Egger | 157.2003 | 105 | 0.0007387 | Heterogeneity_tests | XL_VLDL_P | MDD |
| MR Egger | 144.6359 | 95 | 0.0007846 | Heterogeneity_tests | IDL_TG | MDD |
| MR Egger | 117.1816 | 73 | 0.0007981 | Heterogeneity_tests | IDL_L | MDD |
| MR Egger | 157.953 | 106 | 0.0008039 | Heterogeneity_tests | MDD | XL_VLDL_C |
| Inverse variance weighted | 148.1875 | 98 | 0.0008039 | Heterogeneity_tests | M_VLDL_TG_pct | MDD |
| MR Egger | 142.9286 | 94 | 0.0008588 | Heterogeneity_tests | M_VLDL_TG | MDD |
| MR Egger | 178.0626 | 123 | 0.0008656 | Heterogeneity_tests | XL_HDL_CE | MDD |
| Inverse variance weighted | 145.1312 | 96 | 0.0009002 | Heterogeneity_tests | IDL_TG | MDD |
| Inverse variance weighted | 158.4774 | 107 | 0.0009104 | Heterogeneity_tests | MDD | IDL_CE_pct |
| Inverse variance weighted | 157.2026 | 106 | 0.0009205 | Heterogeneity_tests | XL_VLDL_P | MDD |
| MR Egger | 155.8374 | 105 | 0.0009458 | Heterogeneity_tests | L_VLDL_PL_pct | MDD |
| Inverse variance weighted | 117.2756 | 74 | 0.0010145 | Heterogeneity_tests | IDL_L | MDD |
| Inverse variance weighted | 178.2206 | 124 | 0.0010321 | Heterogeneity_tests | XL_HDL_CE | MDD |
| MR Egger | 147.8664 | 99 | 0.001069 | Heterogeneity_tests | XXL_VLDL_FC | MDD |
| Inverse variance weighted | 142.975 | 95 | 0.001071 | Heterogeneity_tests | M_VLDL_TG | MDD |
| MR Egger | 137.7977 | 91 | 0.0011285 | Heterogeneity_tests | S_VLDL_FC_pct | MDD |
| Inverse variance weighted | 156.0444 | 106 | 0.001132 | Heterogeneity_tests | L_VLDL_PL_pct | MDD |
| Inverse variance weighted | 148.4107 | 100 | 0.0012078 | Heterogeneity_tests | XXL_VLDL_FC | MDD |
| Inverse variance weighted | 137.3051 | 91 | 0.0012378 | Heterogeneity_tests | L_VLDL_CE_pct | MDD |
| Inverse variance weighted | 138.4031 | 92 | 0.0012671 | Heterogeneity_tests | S_VLDL_FC_pct | MDD |
| Inverse variance weighted | 152.3168 | 104 | 0.0014275 | Heterogeneity_tests | Total_TG | MDD |
| MR Egger | 144.6928 | 98 | 0.0015245 | Heterogeneity_tests | S_VLDL_P | MDD |
| Inverse variance weighted | 125.0619 | 82 | 0.0015527 | Heterogeneity_tests | Sphingomyelins | MDD |
| MR Egger | 154.0553 | 106 | 0.0016052 | Heterogeneity_tests | MDD | L_VLDL_C |

| | | | | | | |
|---|---|---|---|---|---|---|
| MR Egger | 134.5153 | 90 | 0.0016566 | Heterogeneity_tests | L_VLDL_CE_pct | MDD |
| Inverse variance weighted | 145.3191 | 99 | 0.001692 | Heterogeneity_tests | S_VLDL_P | MDD |
| MR Egger | 156.0817 | 108 | 0.0017126 | Heterogeneity_tests | XL_VLDL_TG | MDD |
| MR Egger | 109.5437 | 70 | 0.0017675 | Heterogeneity_tests | PUFA_pct | MDD |
| MR Egger | 103.2693 | 65 | 0.0017746 | Heterogeneity_tests | S_LDL_PL_pct | MDD |
| MR Egger | 126.7342 | 84 | 0.0018021 | Heterogeneity_tests | XL_VLDL_C | MDD |
| MR Egger | 142.534 | 97 | 0.0018066 | Heterogeneity_tests | XXL_VLDL_L | MDD |
| MR Egger | 149.5901 | 103 | 0.0018636 | Heterogeneity_tests | Total_TG | MDD |
| MR Egger | 144.7549 | 99 | 0.0018698 | Heterogeneity_tests | XXL_VLDL_C | MDD |
| MR Egger | 127.708 | 85 | 0.0018885 | Heterogeneity_tests | XL_VLDL_TG_pct | MDD |
| Inverse variance weighted | 128.8796 | 86 | 0.0019061 | Heterogeneity_tests | XL_VLDL_TG_pct | MDD |
| Inverse variance weighted | 156.4748 | 109 | 0.0019644 | Heterogeneity_tests | XL_VLDL_TG | MDD |
| MR Egger | 140.8476 | 96 | 0.0019672 | Heterogeneity_tests | XXL_VLDL_P | MDD |
| MR Egger | 142.0097 | 97 | 0.0019831 | Heterogeneity_tests | M_VLDL_TG_pct | MDD |
| Inverse variance weighted | 127.326 | 85 | 0.0020281 | Heterogeneity_tests | XL_VLDL_C | MDD |
| Inverse variance weighted | 142.9299 | 98 | 0.0020857 | Heterogeneity_tests | XXL_VLDL_L | MDD |
| Inverse variance weighted | 153.6003 | 107 | 0.0021325 | Heterogeneity_tests | MDD | VLDL_PL |
| MR Egger | 167.7005 | 119 | 0.0021964 | Heterogeneity_tests | L_HDL_C_pct | MDD |
| Inverse variance weighted | 109.6683 | 71 | 0.0022105 | Heterogeneity_tests | PUFA_pct | MDD |
| Inverse variance weighted | 103.4139 | 66 | 0.0022278 | Heterogeneity_tests | S_LDL_PL_pct | MDD |
| Inverse variance weighted | 144.7711 | 100 | 0.0023015 | Heterogeneity_tests | XXL_VLDL_C | MDD |
| Inverse variance weighted | 148.3309 | 103 | 0.0023161 | Heterogeneity_tests | M_VLDL_CE_pct | MDD |
| Inverse variance weighted | 140.9529 | 97 | 0.0023889 | Heterogeneity_tests | XXL_VLDL_P | MDD |
| MR Egger | 121.5318 | 81 | 0.0024041 | Heterogeneity_tests | Sphingomyelins | MDD |
| Inverse variance weighted | 146.7829 | 102 | 0.0024597 | Heterogeneity_tests | M_VLDL_C_pct | MDD |
| Inverse variance weighted | 167.9963 | 120 | 0.0025345 | Heterogeneity_tests | L_HDL_C_pct | MDD |
| MR Egger | 145.2575 | 101 | 0.0026021 | Heterogeneity_tests | M_VLDL_C_pct | MDD |
| Inverse variance weighted | 147.5161 | 103 | 0.0026614 | Heterogeneity_tests | VLDL_TG | MDD |
| MR Egger | 126.7386 | 86 | 0.0028246 | Heterogeneity_tests | VLDL_PL | MDD |
| MR Egger | 145.8732 | 102 | 0.002872 | Heterogeneity_tests | M_VLDL_CE_pct | MDD |
| Inverse variance weighted | 151.8037 | 107 | 0.0028839 | Heterogeneity_tests | MDD | M_LDL_TG |
| Inverse variance weighted | 127.7249 | 87 | 0.002941 | Heterogeneity_tests | VLDL_PL | MDD |
| MR Egger | 145.524 | 102 | 0.0030467 | Heterogeneity_tests | VLDL_TG | MDD |
| MR Egger | 136.9262 | 95 | 0.0031702 | Heterogeneity_tests | S_VLDL_PL_pct | MDD |
| Inverse variance weighted | 129.6843 | 89 | 0.0031894 | Heterogeneity_tests | L_VLDL_CE | MDD |
| MR Egger | 149.9293 | 106 | 0.0032317 | Heterogeneity_tests | MDD | M_LDL_TG |
| Inverse variance weighted | 151.0752 | 107 | 0.0032535 | Heterogeneity_tests | MDD | XXL_VLDL_PL_pct |
| MR Egger | 150.964 | 107 | 0.0033136 | Heterogeneity_tests | XL_VLDL_L | MDD |
| MR Egger | 108.8109 | 72 | 0.0033224 | Heterogeneity_tests | IDL_FC | MDD |
| Inverse variance weighted | 137.0132 | 96 | 0.0038336 | Heterogeneity_tests | S_VLDL_PL_pct | MDD |
| Inverse variance weighted | 109.2505 | 73 | 0.0038567 | Heterogeneity_tests | IDL_FC | MDD |
| Inverse variance weighted | 151.1418 | 108 | 0.0039046 | Heterogeneity_tests | XL_VLDL_L | MDD |
| MR Egger | 149.919 | 107 | 0.0039309 | Heterogeneity_tests | XS_VLDL_TG | MDD |
| Inverse variance weighted | 149.8125 | 107 | 0.0039994 | Heterogeneity_tests | MDD | XL_VLDL_CE |
| MR Egger | 148.4705 | 106 | 0.004105 | Heterogeneity_tests | MDD | XXL_VLDL_PL_pct |
| MR Egger | 126.9877 | 88 | 0.0041494 | Heterogeneity_tests | L_VLDL_CE | MDD |
| Inverse variance weighted | 122.9055 | 85 | 0.0045161 | Heterogeneity_tests | M_HDL_TG | MDD |

| | | | | | | |
|---|---|---|---|---|---|---|
| MR Egger | 128.7289 | 90 | 0.0046453 | Heterogeneity_tests | L_VLDL_L | MDD |
| MR Egger | 147.7012 | 106 | 0.0046487 | Heterogeneity_tests | MDD | VLDL_PL |
| Inverse variance weighted | 149.938 | 108 | 0.0047378 | Heterogeneity_tests | XS_VLDL_TG | MDD |
| Inverse variance weighted | 148.7033 | 107 | 0.0047817 | Heterogeneity_tests | MDD | L_VLDL_CE |
| MR Egger | 134.4681 | 95 | 0.0048181 | Heterogeneity_tests | XL_VLDL_PL | MDD |
| Inverse variance weighted | 135.544 | 96 | 0.0049085 | Heterogeneity_tests | XL_VLDL_PL | MDD |
| MR Egger | 128.1853 | 90 | 0.0050978 | Heterogeneity_tests | XL_VLDL_FC | MDD |
| Inverse variance weighted | 129.3366 | 91 | 0.0051323 | Heterogeneity_tests | L_VLDL_L | MDD |
| MR Egger | 120.6561 | 84 | 0.0054282 | Heterogeneity_tests | M_HDL_TG | MDD |
| MR Egger | 132.5586 | 94 | 0.0054364 | Heterogeneity_tests | L_LDL_TG_pct | MDD |
| Inverse variance weighted | 128.659 | 91 | 0.0057543 | Heterogeneity_tests | XL_VLDL_FC | MDD |
| Inverse variance weighted | 133.1823 | 95 | 0.0059652 | Heterogeneity_tests | L_LDL_TG_pct | MDD |
| MR Egger | 109.8712 | 76 | 0.0066998 | Heterogeneity_tests | IDL_CE | MDD |
| MR Egger | 144.7668 | 106 | 0.0073861 | Heterogeneity_tests | S_VLDL_TG | MDD |
| Inverse variance weighted | 145.7819 | 107 | 0.0075611 | Heterogeneity_tests | S_VLDL_TG | MDD |
| MR Egger | 144.1183 | 106 | 0.0081617 | Heterogeneity_tests | MDD | IDL_TG |
| Inverse variance weighted | 109.8832 | 77 | 0.0082539 | Heterogeneity_tests | IDL_CE | MDD |
| Inverse variance weighted | 145.0994 | 107 | 0.0083936 | Heterogeneity_tests | MDD | VLDL_FC |
| Inverse variance weighted | 100.18603 | 69 | 0.0084156 | Heterogeneity_tests | Unsaturation | MDD |
| Inverse variance weighted | 145.0349 | 107 | 0.0084764 | Heterogeneity_tests | MDD | S_VLDL_L |
| Inverse variance weighted | 144.7795 | 107 | 0.0088117 | Heterogeneity_tests | MDD | IDL_TG |
| Inverse variance weighted | 144.7476 | 107 | 0.0088543 | Heterogeneity_tests | MDD | S_VLDL_P |
| MR Egger | 134.1435 | 98 | 0.0090105 | Heterogeneity_tests | L_VLDL_TG | MDD |
| MR Egger | 142.5843 | 106 | 0.010299 | Heterogeneity_tests | MDD | XL_VLDL_CE |
| MR Egger | 142.5219 | 106 | 0.0103957 | Heterogeneity_tests | MDD | L_VLDL_CE |
| Inverse variance weighted | 134.2897 | 99 | 0.0105577 | Heterogeneity_tests | L_VLDL_TG | MDD |
| MR Egger | 133.9465 | 99 | 0.0111284 | Heterogeneity_tests | XS_VLDL_TG_pct | MDD |
| MR Egger | 104.4001 | 74 | 0.0114847 | Heterogeneity_tests | IDL_C | MDD |
| Inverse variance weighted | 134.5772 | 100 | 0.0120635 | Heterogeneity_tests | XS_VLDL_TG_pct | MDD |
| MR Egger | 115.6377 | 84 | 0.0126089 | Heterogeneity_tests | S_HDL_FC_pct | MDD |
| MR Egger | 141.2209 | 106 | 0.012609 | Heterogeneity_tests | MDD | S_VLDL_L |
| MR Egger | 141.1155 | 106 | 0.0128056 | Heterogeneity_tests | MDD | Ala |
| MR Egger | 140.8013 | 106 | 0.0134081 | Heterogeneity_tests | MDD | S_VLDL_P |
| Inverse variance weighted | 104.4018 | 75 | 0.0140299 | Heterogeneity_tests | IDL_C | MDD |
| Inverse variance weighted | 116.1253 | 85 | 0.014068 | Heterogeneity_tests | S_HDL_FC_pct | MDD |
| Inverse variance weighted | 70.74003 | 47 | 0.0141482 | Heterogeneity_tests | Citrate | MDD |
| Inverse variance weighted | 141.4716 | 107 | 0.0143773 | Heterogeneity_tests | MDD | Ala |
| MR Egger | 69.28695 | 46 | 0.0148051 | Heterogeneity_tests | Citrate | MDD |
| MR Egger | 95.63135 | 68 | 0.0152505 | Heterogeneity_tests | Unsaturation | MDD |
| Inverse variance weighted | 140.9114 | 107 | 0.0155826 | Heterogeneity_tests | MDD | M_VLDL_L |
| MR Egger | 139.4552 | 106 | 0.0162866 | Heterogeneity_tests | MDD | VLDL_FC |
| Inverse variance weighted | 140.3049 | 107 | 0.0169886 | Heterogeneity_tests | MDD | XS_VLDL_PL_pct |
| MR Egger | 115.7518 | 86 | 0.0178997 | Heterogeneity_tests | VLDL_L | MDD |
| Inverse variance weighted | 116.4323 | 87 | 0.0192608 | Heterogeneity_tests | VLDL_L | MDD |
| MR Egger | 137.3432 | 106 | 0.0219158 | Heterogeneity_tests | MDD | XS_VLDL_PL_pct |
| MR Egger | 106.9872 | 81 | 0.0281922 | Heterogeneity_tests | M_VLDL_L | MDD |
| MR Egger | 135.4171 | 106 | 0.0284722 | Heterogeneity_tests | MDD | M_VLDL_L |

| | | | | | | |
|---|---|---|---|---|---|---|
| MR Egger | 107.7048 | 82 | 0.0300604 | Heterogeneity_tests | XL_VLDL_FC_pct | MDD |
| Inverse variance weighted | 107.3726 | 82 | 0.0315683 | Heterogeneity_tests | M_VLDL_L | MDD |
| Inverse variance weighted | 108.2014 | 83 | 0.033057 | Heterogeneity_tests | XL_VLDL_FC_pct | MDD |
| MR Egger | 87.67621 | 67 | 0.0458968 | Heterogeneity_tests | IDL_FC_pct | MDD |
| Inverse variance weighted | 88.41712 | 68 | 0.0487707 | Heterogeneity_tests | IDL_FC_pct | MDD |
| MR Egger | 92.63456 | 72 | 0.0512753 | Heterogeneity_tests | XS_VLDL_FC_pct | MDD |
| MR Egger | 107.1118 | 85 | 0.0528407 | Heterogeneity_tests | XS_VLDL_PL_pct | MDD |
| Inverse variance weighted | 92.67784 | 73 | 0.0598886 | Heterogeneity_tests | XS_VLDL_FC_pct | MDD |
| Inverse variance weighted | 107.142 | 86 | 0.0610446 | Heterogeneity_tests | XS_VLDL_PL_pct | MDD |
| MR Egger | 126.4409 | 106 | 0.0857 | Heterogeneity_tests | MDD | Citrate |
| Inverse variance weighted | 126.8679 | 107 | 0.092241 | Heterogeneity_tests | MDD | Citrate |
| MR Egger | 96.90448 | 80 | 0.0960684 | Heterogeneity_tests | Omega_6_pct | MDD |
| Inverse variance weighted | 97.07442 | 81 | 0.1075825 | Heterogeneity_tests | Omega_6_pct | MDD |
| MR Egger | 123.0445 | 106 | 0.1233505 | Heterogeneity_tests | MDD | Pyruvate |
| Inverse variance weighted | 100.21337 | 85 | 0.1242277 | Heterogeneity_tests | M_LDL_TG_pct | MDD |
| MR Egger | 99.05155 | 84 | 0.1252607 | Heterogeneity_tests | M_LDL_TG_pct | MDD |
| MR Egger | 94.08053 | 80 | 0.1343978 | Heterogeneity_tests | S_VLDL_C_pct | MDD |
| Inverse variance weighted | 123.147 | 107 | 0.1361599 | Heterogeneity_tests | MDD | Pyruvate |
| MR Egger | 105.4789 | 91 | 0.1423301 | Heterogeneity_tests | XXL_VLDL_TG | MDD |
| Inverse variance weighted | 94.08669 | 81 | 0.1516813 | Heterogeneity_tests | S_VLDL_C_pct | MDD |
| Inverse variance weighted | 105.518 | 92 | 0.1586198 | Heterogeneity_tests | XXL_VLDL_TG | MDD |
| MR Egger | 90.82898 | 81 | 0.2133238 | Heterogeneity_tests | S_HDL_C_pct | MDD |
| Inverse variance weighted | 91.34875 | 82 | 0.2249465 | Heterogeneity_tests | S_HDL_C_pct | MDD |
| MR Egger | 37.29465 | 32 | 0.2385853 | Heterogeneity_tests | Pyruvate | MDD |
| MR Egger | 100.9931 | 92 | 0.2446836 | Heterogeneity_tests | S_VLDL_TG_pct | MDD |
| Inverse variance weighted | 101.0899 | 93 | 0.2658653 | Heterogeneity_tests | S_VLDL_TG_pct | MDD |
| Inverse variance weighted | 37.58517 | 33 | 0.2671485 | Heterogeneity_tests | Pyruvate | MDD |
| MR Egger | 59.19662 | 54 | 0.2916545 | Heterogeneity_tests | XXL_VLDL_PL_pct | MDD |
| Inverse variance weighted | 60.12855 | 55 | 0.2953899 | Heterogeneity_tests | XXL_VLDL_PL_pct | MDD |

**Supplementary Table 9**

**Correlation between MDD and Microbiome metabolic profiles**

| microbe | id | beta_lmm | se_lmm | pval_lmm | beta_lmm_mdd | se_lmm_mdd | pval_lmm_mdd | Corr_lmm | T_corr_lmm | P_corr | FDR_corr |
|---|---|---|---|---|---|---|---|---|---|---|---|
| ChristensenellaceaeR7group | genus.ChristensenellaceaeR7group.id.11283 | -0.4549 | 0.0694 | 3.55E-04 | -1.0906 | 0.1587 | 3.31E-04 | -0.7043 | 11.5288 | 7.82E-22 | 2.46E-19 |
| Christensenellaceae | family.Christensenellaceae.id.1866 | -0.4147 | 0.0585 | 1.69E-04 | -1.1043 | 0.1684 | 5.46E-04 | -0.6767 | 10.6799 | 1.12E-19 | 1.76E-17 |
| Ruminiclostridium6 | genus.Ruminiclostridium6.id.11356 | -0.6898 | 0.1181 | 2.65E-04 | -0.6308 | 0.0675 | 1.48E-08 | -0.6596 | 10.1974 | 1.86E-18 | 1.58E-16 |
| LachnospiraceaeNC2004group | genus.LachnospiraceaeNC2004group.id.11316 | -0.6311 | 0.0814 | 2.86E-07 | -0.6885 | 0.1075 | 4.73E-03 | -0.6592 | 10.1847 | 2.00E-18 | 1.58E-16 |
| Eubacteriumxylanophilumgroup | genus..Eubacteriumxylanophilumgroup.id.14375 | -0.5621 | 0.0789 | 1.67E-03 | -0.7487 | 0.0914 | 4.77E-02 | -0.6487 | 9.9048 | 1.02E-17 | 4.59E-16 |
| Coprococcus1 | genus.Coprococcus1.id.11301 | -0.9752 | 0.1136 | 3.57E-05 | -0.4204 | 0.0497 | 3.11E-08 | -0.6403 | 9.6861 | 3.60E-17 | 1.42E-15 |
| RuminococcaceaeUCG014 | genus.RuminococcaceaeUCG014.id.11371 | -0.4728 | 0.0625 | 9.33E-04 | -0.8558 | 0.1024 | 3.42E-04 | -0.6361 | 9.5783 | 6.71E-17 | 2.35E-15 |
| LachnospiraceaeAC2044group | genus.LachnospiraceaeAC2044group.id.11313 | -1.0770 | 0.2874 | 3.48E-03 | -0.3637 | 0.0978 | 4.34E-02 | -0.6258 | 9.3231 | 2.91E-16 | 8.36E-15 |
| RuminococcaceaeUCG005 | genus.RuminococcaceaeUCG005.id.11363 | -0.3990 | 0.0558 | 3.19E-09 | -0.9816 | 0.1668 | 1.73E-03 | -0.6258 | 9.3222 | 2.92E-16 | 8.36E-15 |
| RuminococcaceaeUCG009 | genus.RuminococcaceaeUCG009.id.11366 | -1.0430 | 0.1394 | 6.89E-04 | -0.3519 | 0.0684 | 2.36E-03 | -0.6058 | 8.8470 | 4.40E-15 | 1.07E-13 |
| RuminococcaceaeUCG007 | genus.RuminococcaceaeUCG007.id.11364 | -0.8979 | 0.1125 | 1.19E-08 | -0.4037 | 0.0903 | 3.96E-03 | -0.6020 | 8.7608 | 7.16E-15 | 1.61E-13 |
| Lachnoclostridium | genus.Lachnoclostridium.id.11308 | 0.5713 | 0.0644 | 1.14E-05 | 0.6307 | 0.0832 | 2.62E-04 | 0.6003 | 8.7210 | 8.97E-15 | 1.88E-13 |
| Ruminococcaceae | family.Ruminococcaceae.id.2050 | -0.5702 | 0.0862 | 6.81E-09 | -0.6239 | 0.0964 | 1.29E-03 | -0.5965 | 8.6347 | 1.46E-14 | 2.87E-13 |
| FamilyXIIIAD3011group | genus.FamilyXIIIAD3011group.id.11293 | -0.7506 | 0.1458 | 1.04E-04 | -0.4632 | 0.1404 | 1.79E-02 | -0.5896 | 8.4816 | 3.45E-14 | 6.39E-13 |
| Thermoanaerobacteraceae | family.Thermoanaerobacteraceae.id.2126 | -1.0077 | 0.1266 | 7.44E-10 | -0.3421 | 0.0387 | 7.25E-04 | -0.5871 | 8.4270 | 4.69E-14 | 8.21E-13 |
| Clostridiaceae1 | family.Clostridiaceae1.id.1869 | -0.4221 | 0.0608 | 1.45E-06 | -0.8140 | 0.1510 | 6.21E-04 | -0.5861 | 8.4057 | 5.28E-14 | 8.75E-13 |
| Ruminococcus1 | genus.Ruminococcus1.id.11373 | -0.5564 | 0.0919 | 3.43E-07 | -0.6110 | 0.0951 | 1.60E-03 | -0.5831 | 8.3395 | 7.64E-14 | 1.20E-12 |
| Gelria | genus.Gelria.id.2134 | -1.0022 | 0.1249 | 1.77E-10 | -0.3385 | 0.0376 | 9.71E-04 | -0.5825 | 8.3256 | 8.26E-14 | 1.24E-12 |
| Clostridiumsensustricto1 | genus.Clostridiumsensustricto1.id.1873 | -0.4102 | 0.0581 | 1.29E-06 | -0.8199 | 0.1528 | 8.71E-04 | -0.5799 | 8.2714 | 1.12E-13 | 1.48E-12 |
| Terrisporobacter | genus.Terrisporobacter.id.11348 | -0.4810 | 0.0571 | 3.69E-11 | -0.6977 | 0.1111 | 1.77E-03 | -0.5793 | 8.2570 | 1.21E-13 | 1.52E-12 |
| RuminococcaceaeUCG010 | genus.RuminococcaceaeUCG010.id.11367 | -0.3760 | 0.0538 | 2.35E-10 | -0.8770 | 0.1197 | 4.57E-04 | -0.5742 | 8.1499 | 2.19E-13 | 2.65E-12 |
| RuminococcaceaeNK4A214group | genus.RuminococcaceaeNK4A214group.id.11358 | -0.4209 | 0.0600 | 3.33E-10 | -0.7534 | 0.0879 | 3.86E-03 | -0.5631 | 7.9172 | 7.91E-13 | 9.03E-12 |
| Anaerotruncus | genus.Anaerotruncus.id.2054 | -0.5188 | 0.0738 | 8.75E-10 | -0.6109 | 0.1140 | 9.26E-04 | -0.5630 | 7.9145 | 8.03E-13 | 9.03E-12 |
| Ruminococcusgnavusgroup | genus..Ruminococcusgnavusgroup.id.14376 | 0.5584 | 0.1033 | 7.35E-05 | 0.5620 | 0.1207 | 9.85E-04 | 0.5602 | 7.8573 | 1.10E-12 | 1.19E-11 |
| RuminococcaceaeUCG002 | genus.RuminococcaceaeUCG002.id.11360 | -0.3440 | 0.0569 | 4.55E-06 | -0.8875 | 0.1678 | 1.01E-03 | -0.5525 | 7.7024 | 2.56E-12 | 2.69E-11 |
| Flavonifractor | genus.Flavonifractor.id.2059 | 0.5967 | 0.1001 | 1.37E-06 | 0.5025 | 0.1006 | 1.15E-03 | 0.5475 | 7.6028 | 4.39E-12 | 4.46E-11 |
| Christensenella | genus.Christensenella.id.1867 | 0.9550 | 0.2145 | 3.53E-03 | 0.3034 | 0.0600 | 3.82E-03 | 0.5383 | 7.4213 | 1.17E-11 | 1.15E-10 |
| Fusobacteriaceae | family.Fusobacteriaceae.id.2208 | 0.7981 | 0.1198 | 1.83E-02 | 0.3584 | 0.0949 | 2.88E-02 | 0.5348 | 7.3547 | 1.67E-11 | 1.59E-10 |
| Coprococcus2 | genus.Coprococcus2.id.11302 | -0.4615 | 0.0758 | 1.72E-08 | -0.6180 | 0.1260 | 2.46E-03 | -0.5340 | 7.3390 | 1.81E-11 | 1.68E-10 |
| RuminococcaceaeUCG003 | genus.RuminococcaceaeUCG003.id.11361 | -0.4026 | 0.0819 | 6.27E-05 | -0.7059 | 0.1254 | 1.23E-03 | -0.5331 | 7.3208 | 2.00E-11 | 1.80E-10 |
| Marvinbryantia | genus.Marvinbryantia.id.2005 | -0.6872 | 0.0930 | 6.08E-09 | -0.4107 | 0.0594 | 4.33E-03 | -0.5312 | 7.2855 | 2.41E-11 | 2.11E-10 |

| | | | | | | | | | | | |
|---|---|---|---|---|---|---|---|---|---|---|---|
| RuminococcaceaeV9D2013group | genus.RuminococcaceaeV9D2013group.id.11372 | -0.7455 | 0.1236 | 6.87E-08 | -0.3735 | 0.0667 | 3.23E-03 | -0.5277 | 7.2183 | 3.45E-11 | 2.94E-10 |
| Eggerthella | genus.Eggerthella.id.819 | 0.6266 | 0.1196 | 7.72E-05 | 0.4375 | 0.1153 | 1.21E-02 | 0.5236 | 7.1401 | 5.22E-11 | 4.33E-10 |
| Subdoligranulum | genus.Subdoligranulum.id.2070 | -1.0483 | 0.2030 | 3.66E-03 | -0.2598 | 0.0507 | 8.14E-03 | -0.5219 | 7.1089 | 6.15E-11 | 4.97E-10 |
| LachnospiraceaeNK4A136group | genus.LachnospiraceaeNK4A136group.id.11319 | -0.6930 | 0.1353 | 4.62E-05 | -0.3926 | 0.0745 | 6.09E-04 | -0.5216 | 7.1034 | 6.33E-11 | 4.98E-10 |
| Intestinibacter | genus.Intestinibacter.id.11345 | -0.5059 | 0.0688 | 4.48E-11 | -0.5370 | 0.1103 | 3.25E-03 | -0.5212 | 7.0967 | 6.56E-11 | 5.04E-10 |
| Caproiciproducens | genus.Caproiciproducens.id.14382 | -0.8789 | 0.1273 | 3.29E-09 | -0.3070 | 0.0766 | 3.80E-03 | -0.5195 | 7.0634 | 7.81E-11 | 5.86E-10 |
| ClostridialesvadinBB60group | family.ClostridialesvadinBB60group.id.11286 | -0.4517 | 0.0707 | 2.19E-08 | -0.5894 | 0.0751 | 4.09E-05 | -0.5160 | 6.9988 | 1.10E-10 | 7.70E-10 |
| Romboutsia | genus.Romboutsia.id.11347 | -0.4785 | 0.0636 | 1.10E-11 | -0.5564 | 0.1005 | 1.37E-03 | -0.5160 | 6.9984 | 1.10E-10 | 7.70E-10 |
| Intestinimonas | genus.Intestinimonas.id.2062 | -0.7245 | 0.2015 | 1.42E-02 | -0.3661 | 0.0509 | 1.28E-04 | -0.5150 | 6.9806 | 1.21E-10 | 8.29E-10 |
| Megamonas | genus.Megamonas.id.2184 | 0.5076 | 0.1177 | 9.86E-03 | 0.5119 | 0.0711 | 2.24E-06 | 0.5098 | 6.8848 | 1.99E-10 | 1.33E-09 |
| Turicibacter | genus.Turicibacter.id.2162 | -0.4504 | 0.0671 | 8.28E-07 | -0.5739 | 0.0871 | 2.34E-06 | -0.5084 | 6.8601 | 2.26E-10 | 1.48E-09 |
| Faecalitalea | genus.Faecalitalea.id.11392 | 0.8681 | 0.2020 | 8.28E-04 | 0.2951 | 0.0978 | 1.28E-02 | 0.5061 | 6.8186 | 2.80E-10 | 1.80E-09 |
| Oxalobacteraceae | family.Oxalobacteraceae.id.2966 | -0.4509 | 0.0791 | 9.89E-07 | -0.5666 | 0.1144 | 2.58E-03 | -0.5055 | 6.8067 | 2.98E-10 | 1.88E-09 |
| Peptostreptococcaceae | family.Peptostreptococcaceae.id.2042 | -0.5191 | 0.0678 | 5.73E-10 | -0.4900 | 0.0987 | 1.47E-03 | -0.5043 | 6.7859 | 3.32E-10 | 2.05E-09 |
| Desulfovibrionaceae | family.Desulfovibrionaceae.id.3169 | -0.8813 | 0.2752 | 1.10E-02 | -0.2849 | 0.1076 | 3.01E-02 | -0.5011 | 6.7282 | 4.47E-10 | 2.71E-09 |
| BacteroidalesS247group | family.BacteroidalesS247group.id.11173 | -0.7184 | 0.1906 | 1.24E-02 | -0.3430 | 0.0987 | 1.29E-02 | -0.4964 | 6.6438 | 6.89E-10 | 4.02E-09 |
| Cellulosilyticum | genus.Cellulosilyticum.id.1995 | -0.8050 | 0.1266 | 5.09E-09 | -0.3041 | 0.0496 | 2.65E-03 | -0.4948 | 6.6153 | 7.97E-10 | 4.56E-09 |
| Senegalimassilia | genus.Senegalimassilia.id.11160 | -0.7631 | 0.1573 | 4.27E-05 | -0.3186 | 0.0571 | 1.04E-03 | -0.4930 | 6.5843 | 9.33E-10 | 5.25E-09 |
| Ruminococcustorquesgroup | genus..Ruminococcustorquesgroup.id.14377 | 0.4605 | 0.1285 | 1.51E-02 | 0.5252 | 0.1236 | 3.89E-03 | 0.4918 | 6.5623 | 1.04E-09 | 5.61E-09 |
| Eubacteriumruminantiumgroup | genus..Eubacteriumruminantiumgroup.id.11340 | -0.7105 | 0.1160 | 2.23E-08 | -0.3388 | 0.0695 | 2.78E-03 | -0.4906 | 6.5417 | 1.16E-09 | 6.09E-09 |
| Parvibacter | genus.Parvibacter.id.824 | -0.9893 | 0.1855 | 1.44E-05 | -0.2431 | 0.0542 | 3.37E-03 | -0.4904 | 6.5388 | 1.18E-09 | 6.09E-09 |
| Lactonifactor | genus.Lactonifactor.id.11332 | 0.6629 | 0.1276 | 9.71E-06 | 0.3615 | 0.0589 | 2.38E-02 | 0.4896 | 6.5233 | 1.27E-09 | 6.45E-09 |
| Eubacteriumeligensgroup | genus..Eubacteriumeligensgroup.id.14372 | -0.3853 | 0.1053 | 4.57E-03 | -0.6143 | 0.1066 | 5.79E-04 | -0.4865 | 6.4699 | 1.67E-09 | 8.35E-09 |
| Anaerofustis | genus.Anaerofustis.id.1931 | 0.6431 | 0.2136 | 9.94E-03 | 0.3646 | 0.0937 | 2.68E-03 | 0.4842 | 6.4298 | 2.04E-09 | 1.00E-08 |
| RuminococcaceaeUCG008 | genus.RuminococcaceaeUCG008.id.11365 | -0.5246 | 0.1125 | 4.19E-04 | -0.4456 | 0.1011 | 2.31E-03 | -0.4835 | 6.4177 | 2.17E-09 | 1.05E-08 |
| Fusobacterium | genus.Fusobacterium.id.2210 | 0.7839 | 0.1264 | 7.13E-09 | 0.2951 | 0.1058 | 3.44E-02 | 0.4809 | 6.3735 | 2.71E-09 | 1.29E-08 |
| Olsenella | genus.Olsenella.id.822 | -0.4951 | 0.0856 | 6.16E-07 | -0.4585 | 0.0864 | 1.01E-02 | -0.4764 | 6.2963 | 3.98E-09 | 1.87E-08 |
| Hydrogenoanaerobacterium | genus.Hydrogenoanaerobacterium.id.2060 | -0.7817 | 0.1281 | 3.12E-07 | -0.2881 | 0.0431 | 2.15E-08 | -0.4746 | 6.2648 | 4.66E-09 | 2.13E-08 |
| Varibaculum | genus.Varibaculum.id.427 | 0.9230 | 0.1971 | 8.57E-06 | 0.2440 | 0.0441 | 1.25E-02 | 0.4746 | 6.2646 | 4.66E-09 | 2.13E-08 |
| Eubacteriumtenuegroup | genus..Eubacteriumtenuegroup.id.14380 | -0.8376 | 0.1998 | 9.82E-03 | -0.2665 | 0.0443 | 6.85E-06 | -0.4724 | 6.2280 | 5.59E-09 | 2.52E-08 |
| Butyricimonas | genus.Butyricimonas.id.945 | -0.5059 | 0.1602 | 9.25E-03 | -0.4270 | 0.1505 | 2.08E-02 | -0.4648 | 6.0990 | 1.06E-08 | 4.70E-08 |
| Clostridiuminnocuumgroup | genus..Clostridiuminnocuumgroup.id.14397 | 0.9395 | 0.2094 | 2.40E-04 | 0.2296 | 0.0498 | 2.19E-03 | 0.4645 | 6.0940 | 1.08E-08 | 4.73E-08 |
| Eubacteriumoxidoreducensgroup | genus..Eubacteriumoxidoreducensgroup.id.11339 | -0.5390 | 0.1109 | 1.85E-05 | -0.3996 | 0.0868 | 3.40E-03 | -0.4641 | 6.0873 | 1.12E-08 | 4.77E-08 |
| Howardella | genus.Howardella.id.2000 | -0.6704 | 0.1134 | 2.65E-06 | -0.3212 | 0.1171 | 2.93E-02 | -0.4640 | 6.0865 | 1.12E-08 | 4.77E-08 |

| | | | | | | | | | | | |
|---|---|---|---|---|---|---|---|---|---|---|---|
| Holdemanella | genus.Holdemanella.id.11393 | -1.0346 | 0.2095 | 2.71E-03 | -0.2033 | 0.0992 | 8.07E-02 | -0.4586 | 5.9969 | 1.74E-08 | 7.02E-08 |
| Epulopiscium | genus.Epulopiscium.id.1998 | -0.5141 | 0.1065 | 8.61E-04 | -0.4088 | 0.0789 | 9.01E-05 | -0.4585 | 5.9939 | 1.76E-08 | 7.02E-08 |
| Parvimonas | genus.Parvimonas.id.1945 | -1.2489 | 0.3847 | 2.26E-02 | -0.1669 | 0.0471 | 7.18E-03 | -0.4566 | 5.9628 | 2.05E-08 | 7.97E-08 |
| Oxalobacter | genus.Oxalobacter.id.2978 | -0.3918 | 0.1525 | 4.28E-02 | -0.5313 | 0.1145 | 2.26E-03 | -0.4562 | 5.9571 | 2.11E-08 | 8.11E-08 |
| Defluviitaleaceae | family.Defluviitaleaceae.id.1924 | -0.6766 | 0.1356 | 1.59E-04 | -0.3050 | 0.0706 | 1.40E-02 | -0.4543 | 5.9245 | 2.47E-08 | 9.37E-08 |
| Atopobium | genus.Atopobium.id.814 | 0.7131 | 0.2930 | 8.94E-02 | 0.2874 | 0.0662 | 6.63E-02 | 0.4527 | 5.8989 | 2.79E-08 | 1.05E-07 |
| Scardovia | genus.Scardovia.id.440 | 0.8060 | 0.1386 | 6.98E-08 | 0.2533 | 0.0436 | 6.66E-07 | 0.4518 | 5.8844 | 2.99E-08 | 1.11E-07 |
| Ruminiclostridium | genus.Ruminiclostridium.id.11353 | -0.9096 | 0.1502 | 2.29E-07 | -0.2220 | 0.0340 | 5.70E-06 | -0.4494 | 5.8447 | 3.62E-08 | 1.33E-07 |
| PrevotellaceaeNK3B31group | genus.PrevotellaceaeNK3B31group.id.11185 | -0.7109 | 0.1698 | 2.85E-03 | -0.2788 | 0.0541 | 9.47E-07 | -0.4452 | 5.7767 | 5.01E-08 | 1.81E-07 |
| RuminococcaceaeUCG004 | genus.RuminococcaceaeUCG004.id.11362 | 0.6075 | 0.1980 | 1.74E-02 | 0.3144 | 0.0791 | 8.62E-03 | 0.4370 | 5.6456 | 9.32E-08 | 3.34E-07 |
| RikenellaceaeRC9gutgroup | genus.RikenellaceaeRC9gutgroup.id.11191 | -0.5565 | 0.1311 | 2.30E-02 | -0.3417 | 0.0808 | 1.02E-02 | -0.4361 | 5.6300 | 1.00E-07 | 3.54E-07 |
| DefluviitaleaceaeUCG011 | genus.DefluviitaleaceaeUCG011.id.11287 | -0.6732 | 0.1449 | 4.57E-04 | -0.2814 | 0.0667 | 4.00E-02 | -0.4352 | 5.6165 | 1.07E-07 | 3.75E-07 |
| Anaeroplasmataceae | family.Anaeroplasmataceae.id.3925 | -0.8210 | 0.2038 | 2.08E-03 | -0.2294 | 0.0544 | 8.77E-03 | -0.4340 | 5.5972 | 1.17E-07 | 4.01E-07 |
| Anaeroplasma | genus.Anaeroplasma.id.3926 | -0.8210 | 0.2038 | 2.08E-03 | -0.2294 | 0.0544 | 8.77E-03 | -0.4340 | 5.5972 | 1.17E-07 | 4.01E-07 |
| Clostridiumsensustricto13 | genus.Clostridiumsensustricto13.id.1877 | -0.8381 | 0.2883 | 2.01E-02 | -0.2232 | 0.0617 | 9.60E-03 | -0.4325 | 5.5730 | 1.31E-07 | 4.44E-07 |
| Rikenellaceae | family.Rikenellaceae.id.967 | -0.4650 | 0.0821 | 1.00E-07 | -0.4012 | 0.0899 | 5.10E-03 | -0.4320 | 5.5650 | 1.36E-07 | 4.56E-07 |
| Alistipes | genus.Alistipes.id.968 | -0.5992 | 0.1089 | 2.11E-07 | -0.3100 | 0.0652 | 6.32E-03 | -0.4310 | 5.5493 | 1.46E-07 | 4.84E-07 |
| ErysipelotrichaceaeUCG006 | genus.ErysipelotrichaceaeUCG006.id.11386 | -0.7927 | 0.2097 | 7.30E-03 | -0.2330 | 0.0430 | 6.10E-03 | -0.4298 | 5.5302 | 1.60E-07 | 5.25E-07 |
| Lachnospiraceae | family.Lachnospiraceae.id.1987 | 0.3260 | 0.0796 | 1.60E-02 | 0.5598 | 0.1465 | 7.57E-03 | 0.4272 | 5.4895 | 1.93E-07 | 6.27E-07 |
| Dielma | genus.Dielma.id.11380 | 1.0898 | 0.3206 | 8.94E-03 | 0.1627 | 0.0306 | 1.48E-05 | 0.4211 | 5.3940 | 3.00E-07 | 9.64E-07 |
| LachnospiraceaeFCS020group | genus.LachnospiraceaeFCS020group.id.11314 | -0.4152 | 0.0998 | 5.84E-04 | -0.4232 | 0.1266 | 1.41E-02 | -0.4192 | 5.3646 | 3.43E-07 | 1.09E-06 |
| Blautia | genus.Blautia.id.1992 | 0.2997 | 0.0942 | 3.36E-02 | 0.5831 | 0.1639 | 9.62E-03 | 0.4180 | 5.3464 | 3.72E-07 | 1.17E-06 |
| Prevotella | genus.Prevotella.id.963 | -0.7899 | 0.4041 | 8.17E-02 | -0.2202 | 0.0839 | 3.17E-02 | -0.4170 | 5.3314 | 3.99E-07 | 1.24E-06 |
| Paeniclostridium | genus.Paeniclostridium.id.14378 | -0.7940 | 0.2774 | 2.69E-02 | -0.2154 | 0.0823 | 3.55E-02 | -0.4136 | 5.2777 | 5.09E-07 | 1.57E-06 |
| Comamonadaceae | family.Comamonadaceae.id.2913 | 0.8121 | 0.3124 | 3.45E-02 | 0.2086 | 0.1530 | 2.34E-01 | 0.4116 | 5.2478 | 5.82E-07 | 1.78E-06 |
| RuminococcaceaeUCG011 | genus.RuminococcaceaeUCG011.id.11368 | -0.4410 | 0.1473 | 1.37E-02 | -0.3837 | 0.1309 | 1.79E-02 | -0.4113 | 5.2436 | 5.93E-07 | 1.80E-06 |
| Eubacteriumrectalegroup | genus..Eubacteriumrectalegroup.id.14374 | 0.4645 | 0.1096 | 1.82E-03 | 0.3615 | 0.0840 | 5.06E-03 | 0.4098 | 5.2194 | 6.61E-07 | 1.98E-06 |
| Peptococcaceae | family.Peptococcaceae.id.2024 | -0.4069 | 0.1006 | 1.58E-03 | -0.4076 | 0.1002 | 9.35E-03 | -0.4072 | 5.1806 | 7.87E-07 | 2.34E-06 |
| Phascolarctobacterium | genus.Phascolarctobacterium.id.2168 | -0.6332 | 0.1957 | 4.60E-03 | -0.2584 | 0.0975 | 3.19E-02 | -0.4045 | 5.1385 | 9.49E-07 | 2.79E-06 |
| Klebsiella | genus.Klebsiella.id.3507 | 0.4792 | 0.1001 | 6.87E-02 | 0.3394 | 0.1027 | 1.41E-02 | 0.4033 | 5.1206 | 1.03E-06 | 2.98E-06 |
| Kocuria | genus.Kocuria.id.642 | -0.7159 | 0.2493 | 2.95E-02 | -0.2271 | 0.0507 | 5.52E-02 | -0.4032 | 5.1193 | 1.03E-06 | 2.98E-06 |
| LachnospiraceaeUCG003 | genus.LachnospiraceaeUCG003.id.11323 | -0.4152 | 0.0838 | 4.07E-06 | -0.3908 | 0.0694 | 2.29E-06 | -0.4028 | 5.1128 | 1.06E-06 | 3.04E-06 |
| Sellimonas | genus.Sellimonas.id.14369 | 0.5607 | 0.1874 | 7.49E-03 | 0.2891 | 0.0709 | 3.09E-03 | 0.4026 | 5.1104 | 1.08E-06 | 3.06E-06 |
| Fastidiosipila | genus.Fastidiosipila.id.2058 | -0.8944 | 0.2416 | 2.01E-02 | -0.1801 | 0.0471 | 9.77E-02 | -0.4014 | 5.0920 | 1.17E-06 | 3.29E-06 |

| | | | | | | | | | | | |
|---|---|---|---|---|---|---|---|---|---|---|---|
| FamilyXIIIUCG001 | genus.FamilyXIIIUCG001.id.11294 | -0.5708 | 0.1283 | 2.76E-04 | -0.2819 | 0.1160 | 5.02E-02 | -0.4011 | 5.0882 | 1.19E-06 | 3.32E-06 |
| Neisseriaceae | family.Neisseriaceae.id.3010 | -0.5528 | 0.1398 | 9.90E-04 | -0.2868 | 0.0668 | 3.70E-03 | -0.3982 | 5.0438 | 1.44E-06 | 3.94E-06 |
| Tyzzerella3 | genus.Tyzzerella3.id.11335 | 0.4832 | 0.1162 | 4.93E-03 | 0.3276 | 0.0885 | 1.71E-02 | 0.3979 | 5.0389 | 1.47E-06 | 3.99E-06 |
| Desulfovibrio | genus.Desulfovibrio.id.3173 | -0.7418 | 0.2116 | 4.74E-02 | -0.2131 | 0.0825 | 7.56E-02 | -0.3976 | 5.0346 | 1.50E-06 | 4.04E-06 |
| PrevotellaceaeUCG004 | genus.PrevotellaceaeUCG004.id.11188 | -0.7507 | 0.2696 | 2.08E-02 | -0.2069 | 0.0824 | 4.30E-02 | -0.3941 | 4.9825 | 1.89E-06 | 5.00E-06 |
| Eubacteriumhalliigroup | genus..Eubacteriumhalliigroup.id.11338 | 0.4290 | 0.1199 | 2.32E-03 | 0.3604 | 0.1465 | 3.94E-02 | 0.3932 | 4.9693 | 2.00E-06 | 5.25E-06 |
| Anaerostipes | genus.Anaerostipes.id.1991 | 0.4057 | 0.0794 | 8.08E-03 | 0.3805 | 0.1224 | 1.95E-02 | 0.3929 | 4.9638 | 2.05E-06 | 5.34E-06 |
| LachnospiraceaeND3007group | genus.LachnospiraceaeND3007group.id.11317 | -0.9660 | 0.2897 | 1.07E-02 | -0.1593 | 0.0574 | 8.57E-02 | -0.3923 | 4.9558 | 2.12E-06 | 5.47E-06 |
| Ruminococcus2 | genus.Ruminococcus2.id.11374 | -0.6314 | 0.2193 | 2.69E-02 | -0.2408 | 0.0827 | 3.14E-02 | -0.3899 | 4.9199 | 2.48E-06 | 6.35E-06 |
| Ruminiclostridium1 | genus.Ruminiclostridium1.id.11354 | -0.7564 | 0.1646 | 9.77E-05 | -0.1996 | 0.0496 | 3.09E-03 | -0.3885 | 4.8993 | 2.71E-06 | 6.88E-06 |
| Butyrivibrio | genus.Butyrivibrio.id.1993 | -0.3954 | 0.0848 | 2.44E-02 | -0.3805 | 0.0815 | 9.52E-04 | -0.3879 | 4.8898 | 2.82E-06 | 7.04E-06 |
| Eubacteriumcoprostanoligenesgroup | genus..Eubacteriumcoprostanoligenesgroup.id.11375 | -0.4049 | 0.1545 | 2.11E-02 | -0.3715 | 0.1151 | 1.43E-02 | -0.3878 | 4.8891 | 2.83E-06 | 7.04E-06 |
| Pseudobutyrivibrio | genus.Pseudobutyrivibrio.id.2010 | -0.6167 | 0.2287 | 2.04E-02 | -0.2439 | 0.0842 | 1.91E-02 | -0.3878 | 4.8881 | 2.84E-06 | 7.04E-06 |
| Oscillibacter | genus.Oscillibacter.id.2063 | -0.4516 | 0.2394 | 1.34E-01 | -0.3288 | 0.0973 | 7.78E-03 | -0.3854 | 4.8525 | 3.31E-06 | 8.15E-06 |
| FamilyXIII | family.FamilyXIII.id.1957 | -0.4188 | 0.1704 | 4.71E-02 | -0.3541 | 0.1325 | 2.49E-02 | -0.3851 | 4.8482 | 3.38E-06 | 8.25E-06 |
| Rhodospirillaceae | family.Rhodospirillaceae.id.2717 | -0.7901 | 0.1650 | 4.74E-06 | -0.1827 | 0.0461 | 1.22E-02 | -0.3799 | 4.7722 | 4.67E-06 | 1.13E-05 |
| Neisseria | genus.Neisseria.id.3032 | -0.4892 | 0.1327 | 2.19E-03 | -0.2949 | 0.0680 | 3.81E-03 | -0.3798 | 4.7706 | 4.70E-06 | 1.13E-05 |
| Acidaminococcaceae | family.Acidaminococcaceae.id.2166 | -0.9547 | 0.2596 | 2.36E-03 | -0.1487 | 0.0629 | 5.08E-02 | -0.3769 | 4.7271 | 5.65E-06 | 1.34E-05 |
| Eubacteriaceae | family.Eubacteriaceae.id.1928 | 0.6405 | 0.2036 | 1.01E-02 | 0.2217 | 0.0574 | 2.13E-03 | 0.3768 | 4.7266 | 5.67E-06 | 1.34E-05 |
| Tyzzerella | genus.Tyzzerella.id.11334 | 0.4889 | 0.2351 | 9.97E-02 | 0.2891 | 0.0937 | 2.04E-02 | 0.3760 | 4.7147 | 5.96E-06 | 1.40E-05 |
| Anaerofilum | genus.Anaerofilum.id.2053 | -0.4521 | 0.2567 | 1.11E-01 | -0.3106 | 0.1050 | 1.81E-02 | -0.3747 | 4.6961 | 6.44E-06 | 1.50E-05 |
| Anaeroglobus | genus.Anaeroglobus.id.2176 | 0.5878 | 0.1299 | 2.20E-05 | 0.2348 | 0.0624 | 1.21E-02 | 0.3715 | 4.6498 | 7.82E-06 | 1.81E-05 |
| Odoribacter | genus.Odoribacter.id.952 | -0.4615 | 0.1387 | 1.54E-02 | -0.2971 | 0.1206 | 4.18E-02 | -0.3703 | 4.6313 | 8.45E-06 | 1.94E-05 |
| Methanosphaera | genus.Methanosphaera.id.124 | -0.4415 | 0.1514 | 3.95E-02 | -0.3067 | 0.0916 | 7.76E-03 | -0.3680 | 4.5982 | 9.69E-06 | 2.21E-05 |
| Gallicola | genus.Gallicola.id.1941 | 0.7756 | 0.2255 | 1.25E-03 | 0.1744 | 0.0377 | 1.03E-03 | 0.3678 | 4.5959 | 9.79E-06 | 2.22E-05 |
| Parascardovia | genus.Parascardovia.id.439 | 0.5738 | 0.1901 | 9.14E-03 | 0.2355 | 0.0774 | 2.16E-02 | 0.3676 | 4.5931 | 9.90E-06 | 2.23E-05 |
| PrevotellaceaeUCG001 | genus.PrevotellaceaeUCG001.id.11186 | -0.7057 | 0.3426 | 9.18E-02 | -0.1894 | 0.0421 | 9.06E-02 | -0.3656 | 4.5638 | 1.12E-05 | 2.50E-05 |
| Enterorhabdus | genus.Enterorhabdus.id.820 | -0.6041 | 0.1217 | 1.11E-02 | -0.2200 | 0.0849 | 5.20E-02 | -0.3645 | 4.5487 | 1.19E-05 | 2.64E-05 |
| Porphyromonadaceae | family.Porphyromonadaceae.id.943 | -0.4982 | 0.1305 | 1.69E-03 | -0.2662 | 0.1327 | 8.09E-02 | -0.3642 | 4.5434 | 1.22E-05 | 2.69E-05 |
| Haemophilus | genus.Haemophilus.id.3698 | -0.4875 | 0.1789 | 1.59E-02 | -0.2690 | 0.1262 | 7.26E-02 | -0.3621 | 4.5133 | 1.38E-05 | 3.02E-05 |
| ErysipelotrichaceaeUCG004 | genus.ErysipelotrichaceaeUCG004.id.11385 | -0.6295 | 0.1812 | 3.69E-02 | -0.2056 | 0.0478 | 8.90E-05 | -0.3598 | 4.4804 | 1.57E-05 | 3.41E-05 |
| Gordonibacter | genus.Gordonibacter.id.821 | 0.5679 | 0.2573 | 6.27E-02 | 0.2276 | 0.0731 | 4.87E-02 | 0.3595 | 4.4762 | 1.60E-05 | 3.45E-05 |
| Barnesiella | genus.Barnesiella.id.944 | -0.6140 | 0.1443 | 9.24E-05 | -0.2073 | 0.0613 | 1.97E-02 | -0.3568 | 4.4376 | 1.87E-05 | 4.01E-05 |
| Finegoldia | genus.Finegoldia.id.1940 | 0.5180 | 0.1448 | 2.39E-02 | 0.2452 | 0.0550 | 2.53E-02 | 0.3564 | 4.4318 | 1.92E-05 | 4.09E-05 |

| | | | | | | | | | | | |
|---|---|---|---|---|---|---|---|---|---|---|---|
| Prevotellaceae | family.Prevotellaceae.id.960 | -0.5554 | 0.1247 | 5.01E-05 | -0.2248 | 0.0528 | 2.58E-04 | -0.3534 | 4.3891 | 2.28E-05 | 4.82E-05 |
| CoriobacteriaceaeUCG003 | genus.CoriobacteriaceaeUCG003.id.11159 | -0.6086 | 0.2574 | 6.04E-02 | -0.2048 | 0.0528 | 9.58E-02 | -0.3531 | 4.3848 | 2.32E-05 | 4.84E-05 |
| Paenibacillaceae | family.Paenibacillaceae.id.1740 | 0.4538 | 0.4427 | 3.25E-01 | 0.2747 | 0.1261 | 1.72E-01 | 0.3531 | 4.3846 | 2.32E-05 | 4.84E-05 |
| Bacteroidespectinophilusgroup | genus..Bacteroidespectinophilusgroup.id.14371 | -0.5416 | 0.1353 | 2.31E-02 | -0.2253 | 0.0674 | 1.21E-02 | -0.3493 | 4.3320 | 2.86E-05 | 5.93E-05 |
| Slackia | genus.Slackia.id.825 | -0.6855 | 0.1684 | 8.18E-05 | -0.1764 | 0.0456 | 6.15E-03 | -0.3477 | 4.3089 | 3.14E-05 | 6.46E-05 |
| Holdemania | genus.Holdemania.id.2157 | 0.8109 | 0.1212 | 1.19E-08 | 0.1458 | 0.0801 | 1.10E-01 | 0.3438 | 4.2538 | 3.90E-05 | 7.93E-05 |
| CandidatusStoquefichus | genus.CandidatusStoquefichus.id.11379 | 0.4679 | 0.1522 | 1.45E-02 | 0.2526 | 0.0912 | 2.90E-02 | 0.3438 | 4.2537 | 3.90E-05 | 7.93E-05 |
| Pasteurellaceae | family.Pasteurellaceae.id.3689 | -0.4307 | 0.1731 | 2.50E-02 | -0.2584 | 0.1229 | 7.08E-02 | -0.3336 | 4.1120 | 6.78E-05 | 1.35E-04 |
| Solobacterium | genus.Solobacterium.id.2161 | -0.5718 | 0.1969 | 7.53E-03 | -0.1910 | 0.0648 | 9.25E-02 | -0.3305 | 4.0688 | 8.00E-05 | 1.58E-04 |
| Peptococcus | genus.Peptococcus.id.2037 | -0.4439 | 0.1533 | 2.09E-02 | -0.2443 | 0.1014 | 5.47E-02 | -0.3293 | 4.0521 | 8.52E-05 | 1.68E-04 |
| Oscillospira | genus.Oscillospira.id.2064 | -0.7630 | 0.1876 | 2.54E-04 | -0.1420 | 0.0612 | 6.48E-02 | -0.3292 | 4.0504 | 8.58E-05 | 1.68E-04 |
| Methanobacteriaceae | family.Methanobacteriaceae.id.121 | -0.3098 | 0.0834 | 1.22E-02 | -0.3370 | 0.1283 | 4.11E-02 | -0.3231 | 3.9670 | 1.18E-04 | 2.26E-04 |
| Sarcina | genus.Sarcina.id.1896 | 1.0866 | 0.2265 | 6.99E-05 | 0.0952 | 0.0609 | 2.11E-01 | 0.3216 | 3.9462 | 1.27E-04 | 2.41E-04 |
| Elusimicrobiaceae | family.Elusimicrobiaceae.id.1635 | 0.6329 | 0.1938 | 5.61E-03 | 0.1633 | 0.0444 | 3.39E-04 | 0.3215 | 3.9453 | 1.28E-04 | 2.41E-04 |
| Elusimicrobium | genus.Elusimicrobium.id.1636 | 0.6329 | 0.1938 | 5.61E-03 | 0.1633 | 0.0444 | 3.39E-04 | 0.3215 | 3.9453 | 1.28E-04 | 2.41E-04 |
| HafniaObesumbacterium | genus.HafniaObesumbacterium.id.14636 | -0.7346 | 0.2527 | 2.27E-02 | -0.1398 | 0.0669 | 7.02E-02 | -0.3205 | 3.9312 | 1.34E-04 | 2.52E-04 |
| Brochothrix | genus.Brochothrix.id.1738 | 0.5760 | 0.1961 | 1.08E-02 | 0.1778 | 0.0878 | 7.37E-02 | 0.3200 | 3.9247 | 1.38E-04 | 2.57E-04 |
| Dorea | genus.Dorea.id.1997 | 0.4552 | 0.1557 | 2.23E-02 | 0.2239 | 0.1026 | 7.30E-02 | 0.3193 | 3.9145 | 1.43E-04 | 2.65E-04 |
| Lachnospira | genus.Lachnospira.id.2004 | -0.3843 | 0.2366 | 1.45E-01 | -0.2581 | 0.0738 | 8.65E-03 | -0.3149 | 3.8551 | 1.78E-04 | 3.28E-04 |
| Oribacterium | genus.Oribacterium.id.2008 | -0.5673 | 0.1898 | 4.90E-03 | -0.1742 | 0.0508 | 1.09E-02 | -0.3144 | 3.8476 | 1.83E-04 | 3.36E-04 |
| Leuconostoc | genus.Leuconostoc.id.1841 | 0.4066 | 0.2008 | 7.46E-02 | 0.2422 | 0.0874 | 3.00E-02 | 0.3138 | 3.8406 | 1.88E-04 | 3.42E-04 |
| Methanobrevibacter | genus.Methanobrevibacter.id.123 | -0.3194 | 0.0978 | 3.03E-02 | -0.3059 | 0.1073 | 4.24E-02 | -0.3126 | 3.8235 | 2.00E-04 | 3.62E-04 |
| LachnospiraceaeUCG008 | genus.LachnospiraceaeUCG008.id.11328 | 0.4495 | 0.1463 | 2.52E-02 | 0.2146 | 0.0628 | 7.50E-03 | 0.3106 | 3.7963 | 2.21E-04 | 3.98E-04 |
| Victivallis | genus.Victivallis.id.2256 | -0.7463 | 0.2942 | 3.30E-02 | -0.1288 | 0.0551 | 5.54E-02 | -0.3100 | 3.7885 | 2.27E-04 | 4.07E-04 |
| Acetanaerobacterium | genus.Acetanaerobacterium.id.2051 | 0.6797 | 0.2841 | 4.99E-02 | 0.1409 | 0.0418 | 4.38E-02 | 0.3094 | 3.7807 | 2.34E-04 | 4.14E-04 |
| LachnospiraceaeUCG004 | genus.LachnospiraceaeUCG004.id.11324 | -0.3697 | 0.1269 | 1.58E-02 | -0.2588 | 0.1386 | 9.33E-02 | -0.3093 | 3.7792 | 2.35E-04 | 4.14E-04 |
| Prevotella9 | genus.Prevotella9.id.11183 | -0.7102 | 0.2185 | 1.67E-02 | -0.1345 | 0.1116 | 3.78E-01 | -0.3091 | 3.7765 | 2.38E-04 | 4.16E-04 |
| LachnospiraceaeNK4B4group | genus.LachnospiraceaeNK4B4group.id.11320 | -0.7038 | 0.1961 | 8.32E-03 | -0.1290 | 0.0546 | 4.01E-02 | -0.3013 | 3.6712 | 3.47E-04 | 6.03E-04 |
| FamilyXI | family.FamilyXI.id.1732 | 0.4786 | 0.2491 | 9.29E-02 | 0.1802 | 0.0810 | 7.36E-02 | 0.2937 | 3.5696 | 4.96E-04 | 8.53E-04 |
| Gemella | genus.Gemella.id.1733 | 0.4786 | 0.2491 | 9.29E-02 | 0.1802 | 0.0810 | 7.36E-02 | 0.2937 | 3.5696 | 4.96E-04 | 8.53E-04 |
| Isobaculum | genus.Isobaculum.id.1822 | -0.6885 | 0.2736 | 2.17E-02 | -0.1235 | 0.0505 | 3.58E-02 | -0.2917 | 3.5427 | 5.44E-04 | 9.30E-04 |
| Paraprevotella | genus.Paraprevotella.id.962 | -0.7006 | 0.1671 | 2.08E-04 | -0.1214 | 0.0592 | 2.91E-01 | -0.2916 | 3.5417 | 5.46E-04 | 9.30E-04 |
| Parasutterella | genus.Parasutterella.id.2892 | 0.4314 | 0.1333 | 4.07E-03 | 0.1855 | 0.0588 | 2.85E-03 | 0.2829 | 3.4266 | 8.10E-04 | 1.37E-03 |
| Rikenella | genus.Rikenella.id.973 | -0.4635 | 0.2408 | 9.16E-02 | -0.1723 | 0.0842 | 9.79E-02 | -0.2826 | 3.4235 | 8.19E-04 | 1.38E-03 |

| | | | | | | | | | | | |
|---|---|---|---|---|---|---|---|---|---|---|---|
| CoriobacteriaceaeUCG002 | genus.CoriobacteriaceaeUCG002.id.11158 | -0.4527 | 0.1369 | 1.35E-03 | -0.1761 | 0.0702 | 4.12E-02 | -0.2823 | 3.4193 | 8.31E-04 | 1.39E-03 |
| Megasphaera | genus.Megasphaera.id.2185 | 0.4752 | 0.1896 | 1.53E-02 | 0.1607 | 0.0729 | 1.43E-01 | 0.2764 | 3.3414 | 1.08E-03 | 1.78E-03 |
| Metascardovia | genus.Metascardovia.id.438 | 0.3610 | 0.2583 | 1.94E-01 | 0.2082 | 0.1149 | 1.98E-01 | 0.2742 | 3.3124 | 1.19E-03 | 1.95E-03 |
| Leuconostocaceae | family.Leuconostocaceae.id.1839 | 0.3795 | 0.1988 | 9.88E-02 | 0.1961 | 0.0963 | 7.60E-02 | 0.2728 | 3.2950 | 1.26E-03 | 2.05E-03 |
| Micrococcaceae | family.Micrococcaceae.id.636 | 0.3723 | 0.2335 | 1.77E-01 | 0.1971 | 0.1106 | 1.13E-01 | 0.2709 | 3.2695 | 1.37E-03 | 2.22E-03 |
| Leptotrichiaceae | family.Leptotrichiaceae.id.2215 | -0.6784 | 0.2834 | 2.94E-02 | -0.1063 | 0.0646 | 1.36E-01 | -0.2686 | 3.2398 | 1.51E-03 | 2.43E-03 |
| Papillibacter | genus.Papillibacter.id.2065 | -0.5476 | 0.1657 | 2.74E-03 | -0.1288 | 0.0778 | 1.33E-01 | -0.2656 | 3.2012 | 1.71E-03 | 2.74E-03 |
| Fusicatenibacter | genus.Fusicatenibacter.id.11305 | 0.4391 | 0.2241 | 9.31E-02 | 0.1600 | 0.0968 | 1.49E-01 | 0.2650 | 3.1934 | 1.75E-03 | 2.80E-03 |
| Succinatimonas | genus.Succinatimonas.id.3329 | 0.5761 | 0.1923 | 2.63E-02 | 0.1203 | 0.0814 | 1.98E-01 | 0.2633 | 3.1706 | 1.88E-03 | 3.00E-03 |
| Ruminiclostridium9 | genus.Ruminiclostridium9.id.11357 | -0.5763 | 0.1638 | 5.96E-04 | -0.1198 | 0.0473 | 2.87E-02 | -0.2627 | 3.1639 | 1.92E-03 | 3.04E-03 |
| Rothia | genus.Rothia.id.646 | 0.3555 | 0.2211 | 1.80E-01 | 0.1929 | 0.1099 | 1.19E-01 | 0.2619 | 3.1527 | 1.99E-03 | 3.14E-03 |
| Anaerosporobacter | genus.Anaerosporobacter.id.1990 | 0.3726 | 0.2235 | 1.88E-01 | 0.1839 | 0.0666 | 2.53E-02 | 0.2618 | 3.1514 | 2.00E-03 | 3.14E-03 |
| Cardiobacteriaceae | family.Cardiobacteriaceae.id.3410 | -0.3265 | 0.2514 | 2.37E-01 | -0.2084 | 0.0954 | 7.32E-02 | -0.2609 | 3.1399 | 2.08E-03 | 3.22E-03 |
| Cardiobacterium | genus.Cardiobacterium.id.3411 | -0.3265 | 0.2514 | 2.37E-01 | -0.2084 | 0.0954 | 7.32E-02 | -0.2609 | 3.1399 | 2.08E-03 | 3.22E-03 |
| Butyricicoccus | genus.Butyricicoccus.id.2055 | 0.4607 | 0.1889 | 8.41E-02 | 0.1422 | 0.0877 | 1.62E-01 | 0.2559 | 3.0759 | 2.54E-03 | 3.92E-03 |
| Pseudoflavonifractor | genus.Pseudoflavonifractor.id.2066 | 0.7911 | 0.3298 | 4.69E-02 | 0.0815 | 0.0403 | 1.13E-01 | 0.2540 | 3.0512 | 2.74E-03 | 4.22E-03 |
| Cloacibacillus | genus.Cloacibacillus.id.3908 | -0.5286 | 0.1690 | 2.43E-03 | -0.1189 | 0.0388 | 5.59E-03 | -0.2507 | 3.0088 | 3.13E-03 | 4.76E-03 |
| Abiotrophia | genus.Abiotrophia.id.1803 | 1.2637 | 0.3335 | 1.07E-02 | 0.0497 | 0.0566 | 4.05E-01 | 0.2506 | 3.0079 | 3.14E-03 | 4.76E-03 |
| Eubacteriumbrachygroup | genus..Eubacteriumbrachygroup.id.11296 | 0.3318 | 0.1856 | 1.17E-01 | 0.1892 | 0.1330 | 1.90E-01 | 0.2506 | 3.0074 | 3.14E-03 | 4.76E-03 |
| Brevibacteriaceae | family.Brevibacteriaceae.id.521 | -0.6533 | 0.3393 | 8.91E-02 | -0.0938 | 0.0681 | 2.22E-01 | -0.2476 | 2.9690 | 3.54E-03 | 5.30E-03 |
| Brevibacterium | genus.Brevibacterium.id.522 | -0.6533 | 0.3393 | 8.91E-02 | -0.0938 | 0.0681 | 2.22E-01 | -0.2476 | 2.9690 | 3.54E-03 | 5.30E-03 |
| Asaccharobacter | genus.Asaccharobacter.id.813 | 0.4935 | 0.2309 | 6.47E-02 | 0.1222 | 0.0554 | 6.85E-02 | 0.2456 | 2.9439 | 3.82E-03 | 5.68E-03 |
| Denitrobacterium | genus.Denitrobacterium.id.818 | -0.4745 | 0.3228 | 1.81E-01 | -0.1271 | 0.1007 | 2.76E-01 | -0.2456 | 2.9435 | 3.82E-03 | 5.68E-03 |
| Peptoniphilus | genus.Peptoniphilus.id.1946 | 0.6300 | 0.2476 | 5.43E-02 | 0.0934 | 0.0316 | 9.32E-02 | 0.2426 | 2.9052 | 4.29E-03 | 6.34E-03 |
| Catabacter | genus.Catabacter.id.11282 | -0.5443 | 0.4220 | 2.29E-01 | -0.1069 | 0.0594 | 1.32E-01 | -0.2413 | 2.8885 | 4.51E-03 | 6.64E-03 |
| possiblegenusSk018 | genus.possiblegenusSk018.id.2019 | -0.4554 | 0.2611 | 1.29E-01 | -0.1238 | 0.1015 | 2.71E-01 | -0.2375 | 2.8406 | 5.20E-03 | 7.62E-03 |
| Alcaligenaceae | family.Alcaligenaceae.id.2875 | 0.3576 | 0.1563 | 2.71E-02 | 0.1529 | 0.0517 | 7.38E-03 | 0.2338 | 2.7944 | 5.96E-03 | 8.69E-03 |
| Synergistes | genus.Synergistes.id.3913 | 0.4240 | 0.4252 | 3.48E-01 | 0.1287 | 0.0477 | 2.48E-02 | 0.2336 | 2.7920 | 6.00E-03 | 8.71E-03 |
| Eubacteriumventriosumgroup | genus..Eubacteriumventriosumgroup.id.11341 | 0.5004 | 0.3174 | 1.57E-01 | 0.1085 | 0.0673 | 1.55E-01 | 0.2330 | 2.7843 | 6.14E-03 | 8.87E-03 |
| RuminococcaceaeUCG013 | genus.RuminococcaceaeUCG013.id.11370 | 0.5322 | 0.2699 | 1.05E-01 | 0.0975 | 0.0577 | 1.56E-01 | 0.2278 | 2.7187 | 7.42E-03 | 1.07E-02 |
| Murimonas | genus.Murimonas.id.14368 | 0.2461 | 0.2150 | 2.80E-01 | 0.2060 | 0.1051 | 9.04E-02 | 0.2252 | 2.6856 | 8.15E-03 | 1.17E-02 |
| Eubacteriumfissicatenagroup | genus..Eubacteriumfissicatenagroup.id.14373 | 0.6660 | 0.3380 | 6.69E-02 | 0.0737 | 0.0542 | 2.24E-01 | 0.2215 | 2.6391 | 9.29E-03 | 1.32E-02 |
| Biostraticola | genus.Biostraticola.id.3471 | -0.6617 | 0.4286 | 1.53E-01 | -0.0729 | 0.0501 | 1.88E-01 | -0.2196 | 2.6153 | 9.93E-03 | 1.41E-02 |
| Pediococcus | genus.Pediococcus.id.1838 | 0.3662 | 0.1925 | 6.85E-02 | 0.1291 | 0.0648 | 8.95E-02 | 0.2175 | 2.5887 | 1.07E-02 | 1.51E-02 |

| | | | | | | | | | | | |
|---|---|---|---|---|---|---|---|---|---|---|---|
| Parabacteroides | genus.Parabacteroides.id.954 | -0.3704 | 0.1574 | 4.01E-02 | -0.1251 | 0.1806 | 5.16E-01 | -0.2152 | 2.5606 | 1.15E-02 | 1.62E-02 |
| Alloscardovia | genus.Alloscardovia.id.435 | 0.2192 | 0.2256 | 3.70E-01 | 0.2090 | 0.1087 | 9.05E-02 | 0.2140 | 2.5459 | 1.20E-02 | 1.68E-02 |
| PrevotellaceaeUCG003 | genus.PrevotellaceaeUCG003.id.11187 | -0.5024 | 0.2433 | 6.54E-02 | -0.0881 | 0.0595 | 1.97E-01 | -0.2104 | 2.5012 | 1.36E-02 | 1.88E-02 |
| LachnospiraceaeUCG001 | genus.LachnospiraceaeUCG001.id.11321 | -0.4238 | 0.1634 | 1.10E-02 | -0.1028 | 0.0433 | 1.90E-02 | -0.2087 | 2.4799 | 1.44E-02 | 1.99E-02 |
| Syntrophomonadaceae | family.Syntrophomonadaceae.id.2072 | -0.4028 | 0.3220 | 2.47E-01 | -0.1059 | 0.1000 | 3.20E-01 | -0.2065 | 2.4522 | 1.55E-02 | 2.13E-02 |
| Alloprevotella | genus.Alloprevotella.id.961 | -0.4233 | 0.2825 | 1.78E-01 | -0.0976 | 0.0577 | 1.34E-01 | -0.2033 | 2.4121 | 1.72E-02 | 2.36E-02 |
| Burkholderiaceae | family.Burkholderiaceae.id.2899 | -0.6488 | 0.4877 | 2.18E-01 | -0.0630 | 0.0948 | 5.26E-01 | -0.2022 | 2.3990 | 1.78E-02 | 2.43E-02 |
| Prevotella1 | genus.Prevotella1.id.11179 | -0.3778 | 0.3670 | 3.32E-01 | -0.1071 | 0.0703 | 1.69E-01 | -0.2011 | 2.3855 | 1.84E-02 | 2.50E-02 |
| Bifidobacteriaceae | family.Bifidobacteriaceae.id.433 | 0.3110 | 0.1402 | 2.86E-02 | 0.1245 | 0.0505 | 1.51E-02 | 0.1968 | 2.3317 | 2.12E-02 | 2.87E-02 |
| Aerococcaceae | family.Aerococcaceae.id.1802 | 0.6551 | 0.2522 | 3.26E-02 | 0.0576 | 0.0796 | 4.87E-01 | 0.1943 | 2.3016 | 2.29E-02 | 3.08E-02 |
| Gardnerella | genus.Gardnerella.id.437 | -0.4827 | 0.1605 | 4.06E-03 | -0.0775 | 0.0493 | 1.31E-01 | -0.1934 | 2.2899 | 2.36E-02 | 3.16E-02 |
| Staphylococcaceae | family.Staphylococcaceae.id.1775 | -0.4437 | 0.2754 | 1.81E-01 | -0.0827 | 0.0469 | 1.43E-01 | -0.1916 | 2.2683 | 2.49E-02 | 3.32E-02 |
| Porphyromonas | genus.Porphyromonas.id.956 | -0.7468 | 0.2610 | 8.20E-03 | -0.0480 | 0.0431 | 3.11E-01 | -0.1894 | 2.2414 | 2.66E-02 | 3.54E-02 |
| Bifidobacterium | genus.Bifidobacterium.id.436 | 0.3304 | 0.1595 | 5.04E-02 | 0.1080 | 0.0474 | 2.46E-02 | 0.1889 | 2.2348 | 2.71E-02 | 3.58E-02 |
| Eubacteriumnodatumgroup | genus..Eubacteriumnodatumgroup.id.11297 | -0.4067 | 0.2223 | 1.08E-01 | -0.0850 | 0.0658 | 2.52E-01 | -0.1859 | 2.1985 | 2.96E-02 | 3.89E-02 |
| Bacteroidaceae | family.Bacteroidaceae.id.917 | -0.5277 | 0.1867 | 1.84E-02 | -0.0628 | 0.1319 | 6.48E-01 | -0.1820 | 2.1503 | 3.33E-02 | 4.34E-02 |
| Bacteroides | genus.Bacteroides.id.918 | -0.5277 | 0.1867 | 1.84E-02 | -0.0628 | 0.1319 | 6.48E-01 | -0.1820 | 2.1503 | 3.33E-02 | 4.34E-02 |
| Prevotella7 | genus.Prevotella7.id.11182 | -0.5585 | 0.2108 | 1.89E-02 | -0.0583 | 0.1066 | 5.98E-01 | -0.1804 | 2.1314 | 3.49E-02 | 4.52E-02 |
| Lactobacillus | genus.Lactobacillus.id.1837 | -0.4572 | 0.3337 | 2.13E-01 | -0.0709 | 0.0424 | 1.97E-01 | -0.1801 | 2.1272 | 3.52E-02 | 4.55E-02 |
| Mogibacterium | genus.Mogibacterium.id.1960 | -0.3635 | 0.2060 | 1.13E-01 | -0.0870 | 0.0776 | 2.98E-01 | -0.1779 | 2.1001 | 3.76E-02 | 4.83E-02 |
| Bilophila | genus.Bilophila.id.3170 | -0.1613 | 0.3724 | 6.73E-01 | -0.1941 | 0.1348 | 1.91E-01 | -0.1770 | 2.0890 | 3.86E-02 | 4.94E-02 |
| Carnobacterium | genus.Carnobacterium.id.1818 | -0.2087 | 0.1180 | 8.13E-02 | -0.1469 | 0.0842 | 1.78E-01 | -0.1751 | 2.0665 | 4.07E-02 | 5.18E-02 |
| Prevotella2 | genus.Prevotella2.id.11180 | -0.3906 | 0.2300 | 1.54E-01 | -0.0784 | 0.0756 | 3.89E-01 | -0.1750 | 2.0652 | 4.08E-02 | 5.18E-02 |
| Roseburia | genus.Roseburia.id.2012 | 0.3847 | 0.1943 | 9.70E-02 | 0.0731 | 0.0701 | 3.09E-01 | 0.1677 | 1.9768 | 5.01E-02 | 6.34E-02 |
| Ezakiella | genus.Ezakiella.id.11289 | -0.2854 | 0.2133 | 2.38E-01 | -0.0952 | 0.0563 | 2.06E-01 | -0.1649 | 1.9420 | 5.42E-02 | 6.83E-02 |
| Planococcaceae | family.Planococcaceae.id.1753 | 0.2811 | 0.2185 | 2.20E-01 | 0.0923 | 0.0898 | 3.34E-01 | 0.1611 | 1.8960 | 6.01E-02 | 7.54E-02 |
| Anaerococcus | genus.Anaerococcus.id.1937 | 0.3047 | 0.3061 | 3.46E-01 | 0.0830 | 0.0665 | 2.53E-01 | 0.1590 | 1.8714 | 6.35E-02 | 7.90E-02 |
| Murdochiella | genus.Murdochiella.id.1944 | -0.3205 | 0.2842 | 3.04E-01 | -0.0789 | 0.0563 | 2.22E-01 | -0.1590 | 1.8712 | 6.35E-02 | 7.90E-02 |
| Faecalicoccus | genus.Faecalicoccus.id.11391 | 0.3563 | 0.3743 | 3.78E-01 | 0.0701 | 0.0601 | 2.99E-01 | 0.1580 | 1.8595 | 6.51E-02 | 8.08E-02 |
| Mitsuokella | genus.Mitsuokella.id.2186 | -0.3024 | 0.1490 | 5.20E-01 | -0.0758 | 0.0225 | 9.85E-04 | -0.1514 | 1.7792 | 7.75E-02 | 9.57E-02 |
| Coprococcus3 | genus.Coprococcus3.id.11303 | 0.3119 | 0.2666 | 2.78E-01 | 0.0717 | 0.0735 | 3.67E-01 | 0.1495 | 1.7573 | 8.11E-02 | 9.97E-02 |
| Granulicatella | genus.Granulicatella.id.1821 | 0.2997 | 0.1763 | 1.62E-01 | 0.0745 | 0.1266 | 5.87E-01 | 0.1494 | 1.7561 | 8.13E-02 | 9.97E-02 |
| Victivallaceae | family.Victivallaceae.id.2255 | -0.3569 | 0.2029 | 8.51E-02 | -0.0609 | 0.0356 | 8.98E-02 | -0.1475 | 1.7325 | 8.55E-02 | 1.04E-01 |
| Hungatella | genus.Hungatella.id.11306 | -0.4109 | 0.2889 | 1.61E-01 | -0.0523 | 0.0324 | 1.44E-01 | -0.1466 | 1.7221 | 8.73E-02 | 1.06E-01 |

| | | | | | | | | | | | |
|---|---|---|---|---|---|---|---|---|---|---|---|
| Lactococcus | genus.Lactococcus.id.1851 | -0.2971 | 0.2846 | 3.33E-01 | -0.0718 | 0.0767 | 3.90E-01 | -0.1461 | 1.7156 | 8.85E-02 | 1.07E-01 |
| Coprobacter | genus.Coprobacter.id.949 | -0.4500 | 0.1658 | 1.39E-02 | -0.0459 | 0.0813 | 5.90E-01 | -0.1438 | 1.6879 | 9.37E-02 | 1.13E-01 |
| Acidaminococcus | genus.Acidaminococcus.id.2167 | 0.1560 | 0.1180 | 1.94E-01 | 0.1297 | 0.0740 | 1.10E-01 | 0.1423 | 1.6702 | 9.72E-02 | 1.17E-01 |
| Staphylococcus | genus.Staphylococcus.id.1780 | -0.2199 | 0.3364 | 5.34E-01 | -0.0848 | 0.0562 | 1.73E-01 | -0.1366 | 1.6019 | 1.12E-01 | 1.34E-01 |
| LachnospiraceaeUCG010 | genus.LachnospiraceaeUCG010.id.11330 | 0.2649 | 0.1118 | 2.12E-02 | 0.0685 | 0.1026 | 5.40E-01 | 0.1347 | 1.5798 | 1.17E-01 | 1.39E-01 |
| Lactobacillaceae | family.Lactobacillaceae.id.1836 | -0.3807 | 0.3454 | 3.06E-01 | -0.0475 | 0.0452 | 3.67E-01 | -0.1345 | 1.5775 | 1.17E-01 | 1.39E-01 |
| Peptostreptococcus | genus.Peptostreptococcus.id.2045 | 0.4159 | 0.2099 | 5.02E-02 | 0.0425 | 0.0379 | 3.96E-01 | 0.1330 | 1.5592 | 1.21E-01 | 1.44E-01 |
| Actinomyces | genus.Actinomyces.id.423 | 0.1973 | 0.2855 | 5.13E-01 | 0.0870 | 0.1000 | 4.18E-01 | 0.1310 | 1.5357 | 1.27E-01 | 1.50E-01 |
| Actinomycetaceae | family.Actinomycetaceae.id.421 | 0.2695 | 0.2398 | 3.03E-01 | 0.0629 | 0.0880 | 5.01E-01 | 0.1302 | 1.5255 | 1.29E-01 | 1.52E-01 |
| Lawsonella | genus.Lawsonella.id.14206 | -0.1739 | 0.3349 | 6.14E-01 | -0.0972 | 0.0726 | 2.16E-01 | -0.1300 | 1.5236 | 1.30E-01 | 1.52E-01 |
| Coriobacteriaceae | family.Coriobacteriaceae.id.811 | -0.1932 | 0.3719 | 6.17E-01 | -0.0866 | 0.0546 | 1.54E-01 | -0.1294 | 1.5158 | 1.32E-01 | 1.54E-01 |
| Streptococcus | genus.Streptococcus.id.1853 | 0.1849 | 0.2143 | 4.12E-01 | 0.0895 | 0.1223 | 4.95E-01 | 0.1286 | 1.5071 | 1.34E-01 | 1.56E-01 |
| Bacillaceae | family.Bacillaceae.id.1680 | 0.3489 | 0.2554 | 1.82E-01 | 0.0471 | 0.0302 | 1.21E-01 | 0.1283 | 1.5027 | 1.35E-01 | 1.57E-01 |
| Faecalibacterium | genus.Faecalibacterium.id.2057 | -0.2517 | 0.2571 | 3.62E-01 | -0.0642 | 0.0942 | 5.16E-01 | -0.1271 | 1.4890 | 1.39E-01 | 1.60E-01 |
| Catenibacterium | genus.Catenibacterium.id.2153 | -0.1617 | 0.3168 | 6.27E-01 | -0.0998 | 0.0872 | 3.04E-01 | -0.1270 | 1.4878 | 1.39E-01 | 1.60E-01 |
| LachnospiraceaeNK3A20group | genus.LachnospiraceaeNK3A20group.id.11318 | 0.1995 | 0.2345 | 4.36E-01 | 0.0762 | 0.0421 | 1.53E-01 | 0.1233 | 1.4437 | 1.51E-01 | 1.73E-01 |
| Mobiluncus | genus.Mobiluncus.id.425 | -0.5256 | 0.3333 | 1.84E-01 | -0.0279 | 0.0397 | 4.91E-01 | -0.1211 | 1.4177 | 1.59E-01 | 1.80E-01 |
| Streptococcaceae | family.Streptococcaceae.id.1850 | 0.1468 | 0.2132 | 5.13E-01 | 0.0999 | 0.1271 | 4.62E-01 | 0.1211 | 1.4176 | 1.59E-01 | 1.80E-01 |
| Dialister | genus.Dialister.id.2183 | -0.3134 | 0.1719 | 8.84E-02 | -0.0418 | 0.0534 | 4.44E-01 | -0.1145 | 1.3387 | 1.83E-01 | 2.07E-01 |
| Corynebacterium | genus.Corynebacterium.id.449 | -0.6323 | 0.2101 | 8.57E-03 | -0.0195 | 0.1206 | 8.75E-01 | -0.1111 | 1.2992 | 1.96E-01 | 2.21E-01 |
| Ruminiclostridium5 | genus.Ruminiclostridium5.id.11355 | -0.1017 | 0.3340 | 7.68E-01 | -0.1169 | 0.0996 | 2.82E-01 | -0.1090 | 1.2742 | 2.05E-01 | 2.30E-01 |
| Bacillus | genus.Bacillus.id.1688 | 0.4958 | 0.5098 | 3.54E-01 | 0.0237 | 0.1109 | 8.36E-01 | 0.1083 | 1.2662 | 2.08E-01 | 2.32E-01 |
| Eisenbergiella | genus.Eisenbergiella.id.11304 | -0.2612 | 0.2392 | 2.82E-01 | -0.0409 | 0.0505 | 4.77E-01 | -0.1034 | 1.2073 | 2.29E-01 | 2.54E-01 |
| Aggregatibacter | genus.Aggregatibacter.id.3691 | 0.1336 | 0.4184 | 7.58E-01 | 0.0791 | 0.0852 | 3.87E-01 | 0.1028 | 1.2007 | 2.32E-01 | 2.56E-01 |
| Cryptobacterium | genus.Cryptobacterium.id.817 | -0.0941 | 0.2274 | 6.95E-01 | -0.1096 | 0.0805 | 2.14E-01 | -0.1015 | 1.1858 | 2.38E-01 | 2.62E-01 |
| Campylobacteraceae | family.Campylobacteraceae.id.3273 | -0.0804 | 0.2011 | 6.99E-01 | -0.1111 | 0.1111 | 3.40E-01 | -0.0945 | 1.1030 | 2.72E-01 | 2.97E-01 |
| Moraxellaceae | family.Moraxellaceae.id.3710 | 0.4794 | 0.3463 | 2.14E-01 | 0.0179 | 0.0327 | 6.42E-01 | 0.0925 | 1.0795 | 2.82E-01 | 3.08E-01 |
| ErysipelotrichaceaeUCG003 | genus.ErysipelotrichaceaeUCG003.id.11384 | 0.2126 | 0.1947 | 3.15E-01 | 0.0393 | 0.0851 | 6.65E-01 | 0.0915 | 1.0672 | 2.88E-01 | 3.13E-01 |
| Synergistaceae | family.Synergistaceae.id.3901 | -0.1700 | 0.3755 | 6.77E-01 | -0.0469 | 0.0514 | 4.00E-01 | -0.0893 | 1.0418 | 2.99E-01 | 3.24E-01 |
| Coprobacillus | genus.Coprobacillus.id.2154 | 0.0737 | 0.4239 | 8.66E-01 | 0.0959 | 0.0758 | 2.47E-01 | 0.0841 | 0.9801 | 3.29E-01 | 3.55E-01 |
| Verrucomicrobiaceae | family.Verrucomicrobiaceae.id.4036 | -0.0682 | 0.1761 | 7.12E-01 | -0.1024 | 0.1158 | 4.05E-01 | -0.0836 | 0.9743 | 3.32E-01 | 3.55E-01 |
| Akkermansia | genus.Akkermansia.id.4037 | -0.0682 | 0.1761 | 7.12E-01 | -0.1024 | 0.1158 | 4.05E-01 | -0.0836 | 0.9743 | 3.32E-01 | 3.55E-01 |
| Erysipelotrichaceae | family.Erysipelotrichaceae.id.2149 | 0.1606 | 0.1935 | 4.38E-01 | 0.0409 | 0.0545 | 5.09E-01 | 0.0811 | 0.9454 | 3.46E-01 | 3.69E-01 |
| Listeriaceae | family.Listeriaceae.id.1737 | -0.1336 | 0.1934 | 5.13E-01 | -0.0492 | 0.1022 | 6.43E-01 | -0.0811 | 0.9449 | 3.46E-01 | 3.69E-01 |

| | | | | | | | | | | | |
|---|---|---|---|---|---|---|---|---|---|---|---|
| Ruminococcusgauvreauiigroup | genus..Ruminococcusgauvreauiigroup.id.11342 | 0.2542 | 0.1669 | 2.10E-01 | 0.0238 | 0.0793 | 7.81E-01 | 0.0779 | 0.9075 | 3.66E-01 | 3.88E-01 |
| Collinsella | genus.Collinsella.id.815 | 0.1074 | 0.3026 | 7.32E-01 | 0.0417 | 0.0501 | 4.36E-01 | 0.0669 | 0.7793 | 4.37E-01 | 4.61E-01 |
| Adlercreutzia | genus.Adlercreutzia.id.812 | 0.4665 | 0.2029 | 5.64E-02 | 0.0095 | 0.2229 | 9.67E-01 | 0.0666 | 0.7754 | 4.39E-01 | 4.61E-01 |
| Erysipelatoclostridium | genus.Erysipelatoclostridium.id.11381 | -0.1103 | 0.1982 | 5.79E-01 | -0.0388 | 0.0368 | 2.96E-01 | -0.0654 | 0.7619 | 4.47E-01 | 4.68E-01 |
| Catenisphaera | genus.Catenisphaera.id.14395 | -0.2442 | 0.3013 | 4.51E-01 | -0.0170 | 0.0679 | 8.10E-01 | -0.0645 | 0.7513 | 4.54E-01 | 4.73E-01 |
| EscherichiaShigella | genus.EscherichiaShigella.id.3504 | 0.2336 | 0.2361 | 3.50E-01 | 0.0145 | 0.0803 | 8.61E-01 | 0.0582 | 0.6772 | 4.99E-01 | 5.19E-01 |
| Enterococcaceae | family.Enterococcaceae.id.1828 | -0.0621 | 0.2172 | 8.09E-01 | -0.0511 | 0.0592 | 4.32E-01 | -0.0563 | 0.6556 | 5.13E-01 | 5.32E-01 |
| Eubacteriumsaphenumgroup | genus..Eubacteriumsaphenumgroup.id.14363 | 0.1776 | 0.4386 | 6.97E-01 | 0.0157 | 0.0615 | 8.13E-01 | 0.0528 | 0.6143 | 5.40E-01 | 5.56E-01 |
| Stomatobaculum | genus.Stomatobaculum.id.2015 | -0.0945 | 0.3048 | 7.66E-01 | -0.0276 | 0.0461 | 5.67E-01 | -0.0511 | 0.5943 | 5.53E-01 | 5.66E-01 |
| Carnobacteriaceae | family.Carnobacteriaceae.id.1811 | 0.1656 | 0.1699 | 3.88E-01 | 0.0131 | 0.1131 | 9.14E-01 | 0.0466 | 0.5418 | 5.89E-01 | 6.00E-01 |
| Sutterella | genus.Sutterella.id.2896 | -0.2209 | 0.2431 | 3.85E-01 | -0.0088 | 0.1116 | 9.39E-01 | -0.0441 | 0.5133 | 6.09E-01 | 6.18E-01 |
| Campylobacter | genus.Campylobacter.id.3275 | -0.0325 | 0.2011 | 8.76E-01 | -0.0469 | 0.1187 | 7.00E-01 | -0.0390 | 0.4538 | 6.51E-01 | 6.59E-01 |
| Pseudomonas | genus.Pseudomonas.id.3723 | -0.2648 | 0.2068 | 2.14E-01 | -0.0033 | 0.0711 | 9.69E-01 | -0.0297 | 0.3448 | 7.31E-01 | 7.35E-01 |
| Lachnoanaerobaculum | genus.Lachnoanaerobaculum.id.2003 | 0.3856 | 0.2951 | 2.39E-01 | 0.0008 | 0.0646 | 9.91E-01 | 0.0170 | 0.1981 | 8.43E-01 | 8.46E-01 |
| Allisonella | genus.Allisonella.id.2174 | 0.0006 | 0.2950 | 9.99E-01 | 0.0031 | 0.0783 | 9.70E-01 | 0.0013 | 0.0152 | 9.88E-01 | 9.88E-01 |
| Pseudomonadaceae | family.Pseudomonadaceae.id.3718 | -0.2997 | 0.2129 | 1.79E-01 | 0.0473 | 0.1078 | 7.02E-01 | Inf | NA | NA | NA |
| Succinivibrio | genus.Succinivibrio.id.3331 | -0.1840 | 0.2358 | 4.64E-01 | 0.0733 | 0.0695 | 3.26E-01 | Inf | NA | NA | NA |
| Tyzzerella4 | genus.Tyzzerella4.id.11336 | 0.4029 | 0.4931 | 4.36E-01 | -0.0473 | 0.0729 | 5.33E-01 | Inf | NA | NA | NA |
| Prevotella6 | genus.Prevotella6.id.11181 | -0.0575 | 0.4232 | 8.98E-01 | 0.0426 | 0.0367 | 2.74E-01 | Inf | NA | NA | NA |
| Acinetobacter | genus.Acinetobacter.id.3711 | 0.3364 | 0.3114 | 3.26E-01 | -0.0067 | 0.0382 | 8.75E-01 | Inf | NA | NA | NA |
| Succiniclasticum | genus.Succiniclasticum.id.2169 | -0.2369 | 0.4658 | 6.26E-01 | 0.0310 | 0.0397 | 4.55E-01 | Inf | NA | NA | NA |
| LachnospiraceaeUCG007 | genus.LachnospiraceaeUCG007.id.11327 | 0.2575 | 0.3024 | 3.98E-01 | -0.0158 | 0.0435 | 7.51E-01 | Inf | NA | NA | NA |
| Veillonellaceae | family.Veillonellaceae.id.2172 | -0.0539 | 0.1578 | 7.35E-01 | 0.0401 | 0.0482 | 4.11E-01 | Inf | NA | NA | NA |
| Leptotrichia | genus.Leptotrichia.id.2216 | -0.3248 | 0.2963 | 3.38E-01 | 0.0003 | 0.0456 | 9.96E-01 | Inf | NA | NA | NA |
| Enterococcus | genus.Enterococcus.id.1831 | 0.0936 | 0.1872 | 6.20E-01 | -0.0403 | 0.0629 | 5.47E-01 | Inf | NA | NA | NA |
| Corynebacteriaceae | family.Corynebacteriaceae.id.448 | -0.2832 | 0.3173 | 4.03E-01 | 0.0244 | 0.1440 | 8.71E-01 | Inf | NA | NA | NA |
| vadinBE97 | family.vadinBE97.id.14446 | -0.2930 | 0.3828 | 4.68E-01 | 0.0200 | 0.0679 | 7.86E-01 | Inf | NA | NA | NA |
| Morganella | genus.Morganella.id.3512 | -0.1137 | 0.4085 | 7.88E-01 | 0.0484 | 0.0675 | 4.93E-01 | Inf | NA | NA | NA |
| Enterobacteriaceae | family.Enterobacteriaceae.id.3469 | 0.1283 | 0.1865 | 5.11E-01 | -0.0165 | 0.0910 | 8.60E-01 | Inf | NA | NA | NA |
| Succinivibrionaceae | family.Succinivibrionaceae.id.3326 | -0.0656 | 0.1976 | 7.51E-01 | 0.0353 | 0.0594 | 5.85E-01 | Inf | NA | NA | NA |
| Parasporobacterium | genus.Parasporobacterium.id.2009 | 0.2238 | 0.2991 | 4.79E-01 | -0.0047 | 0.0528 | 9.33E-01 | Inf | NA | NA | NA |
| Sneathia | genus.Sneathia.id.2218 | -0.1245 | 0.2229 | 5.83E-01 | 0.0168 | 0.0784 | 8.37E-01 | Inf | NA | NA | NA |
| CandidatusSoleaferrea | genus.CandidatusSoleaferrea.id.11350 | 0.0151 | 0.2002 | 9.42E-01 | -0.0720 | 0.1102 | 5.29E-01 | Inf | NA | NA | NA |
| FamilyXIIIUCG002 | genus.FamilyXIIIUCG002.id.11295 | 0.1112 | 0.3376 | 7.50E-01 | -0.0275 | 0.0645 | 6.80E-01 | Inf | NA | NA | NA |

| | | | | | | | | | | | |
|---|---|---|---|---|---|---|---|---|---|---|---|
| Eubacterium | genus.Eubacterium.id.1932 | -0.0183 | 0.2559 | 9.47E-01 | 0.0796 | 0.1059 | 5.47E-01 | Inf | NA | NA | NA |
| Weissella | genus.Weissella.id.1843 | 0.0178 | 0.4372 | 9.69E-01 | -0.0475 | 0.0773 | 5.53E-01 | Inf | NA | NA | NA |
| Veillonella | genus.Veillonella.id.2198 | -0.1271 | 0.2605 | 6.38E-01 | 0.0307 | 0.1475 | 8.43E-01 | Inf | NA | NA | NA |
| Corynebacterium1 | genus.Corynebacterium1.id.11143 | -0.1097 | 0.4306 | 8.05E-01 | 0.0393 | 0.0995 | 7.10E-01 | Inf | NA | NA | NA |
