## Supplementary Figures for "Interplay of Human Metabolome and Gut Microbiome in Major Depression"

### Supplementary Figure 1: Metabolome-wide association results (model 1)

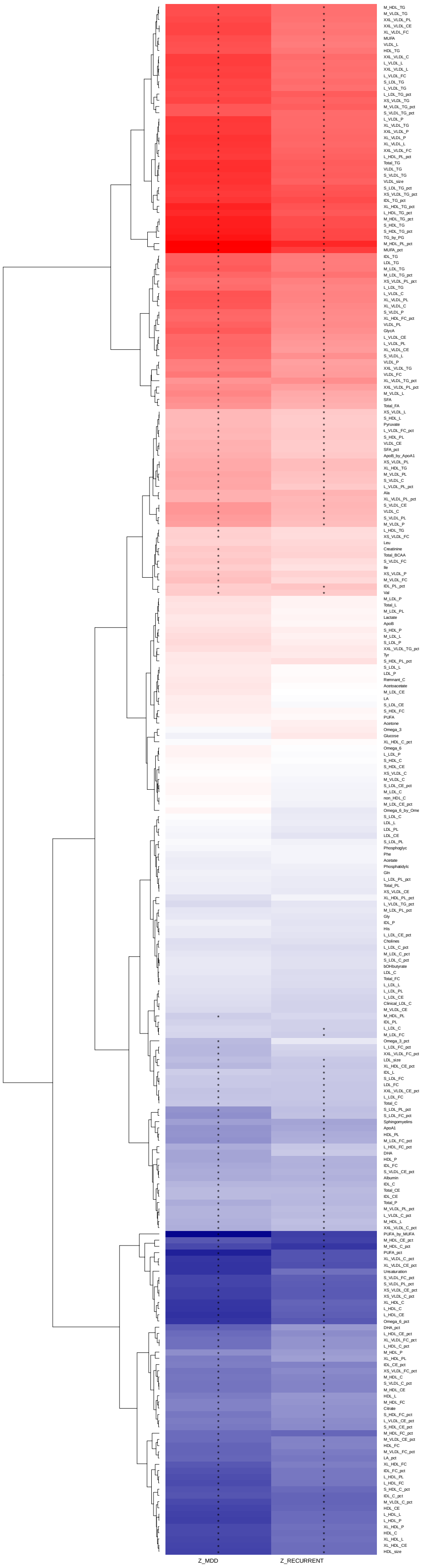

### Supplementary Figure 2: Metabolome-wide association results (model 2)

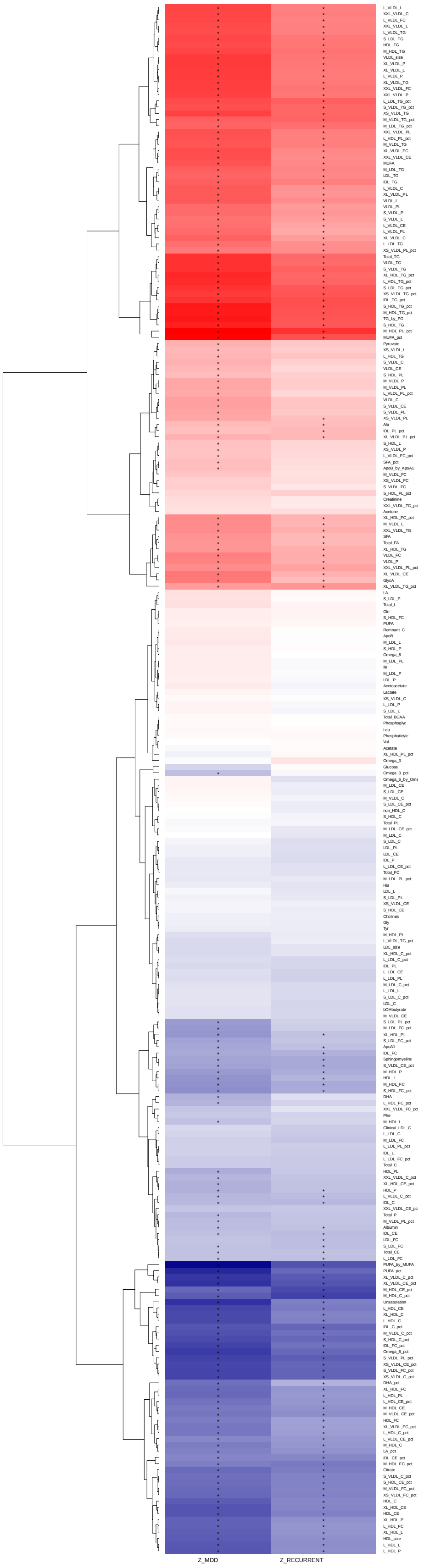

Supplementary Figure 3: Metabolome-wide association results (model 3)

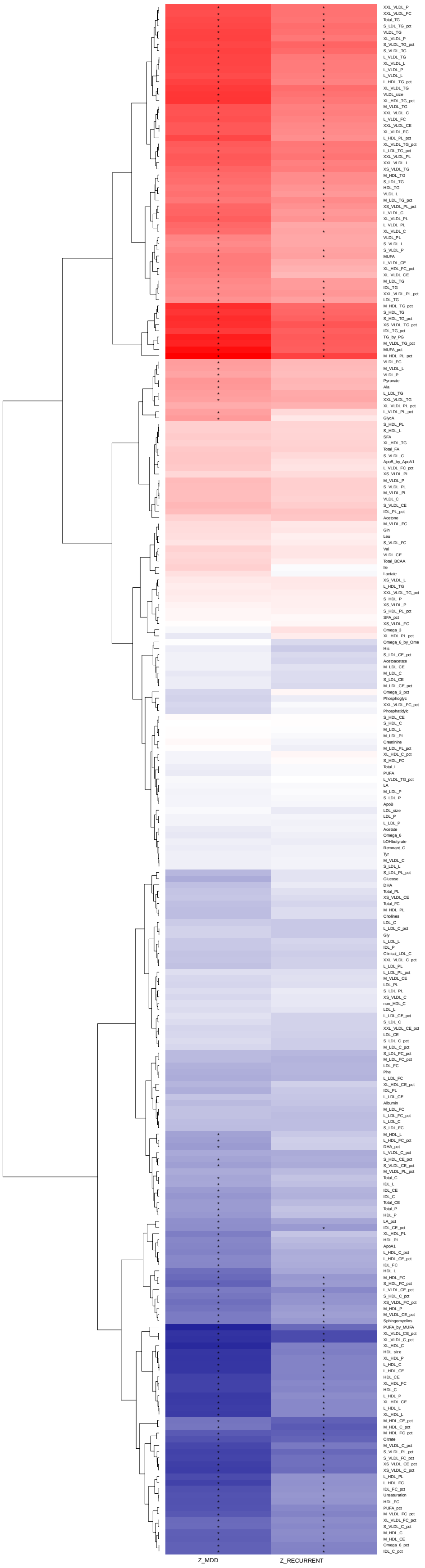

### Supplementary Figure 4: Metabolome-wide association results (model 4)

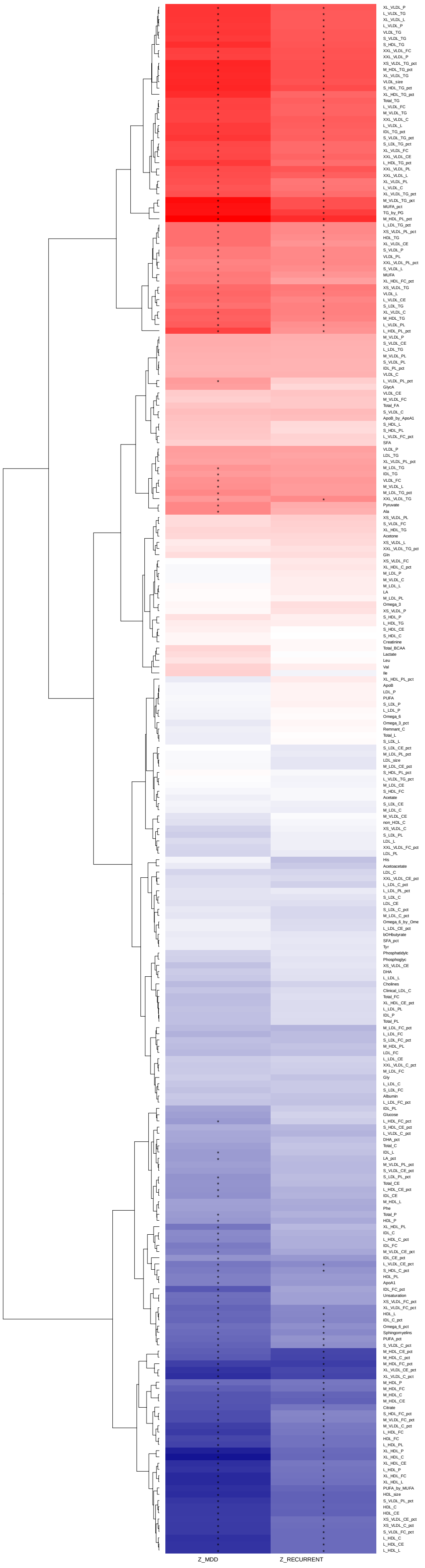

Supplementary Figure 5: Scatter plot of effect estimates (MDD) from model 4 and sensitivity analysis by removing individuals on antidepressants

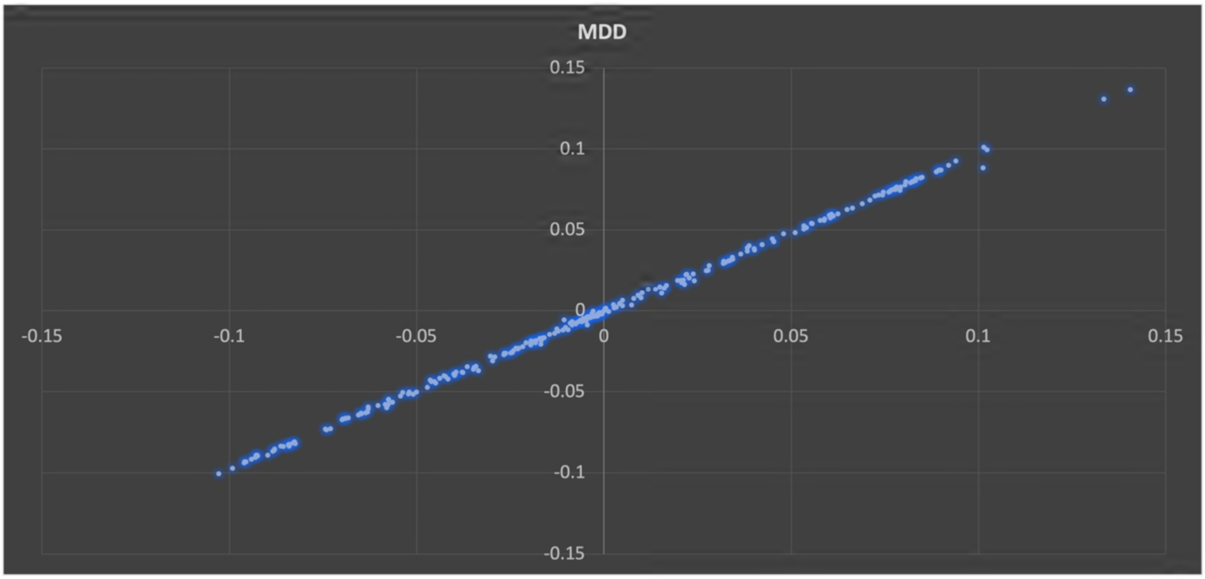

#### Supplementary Figure 6: Significantly associated metabolites with MDD in model 4 and the results of replication in BBMRI-NL

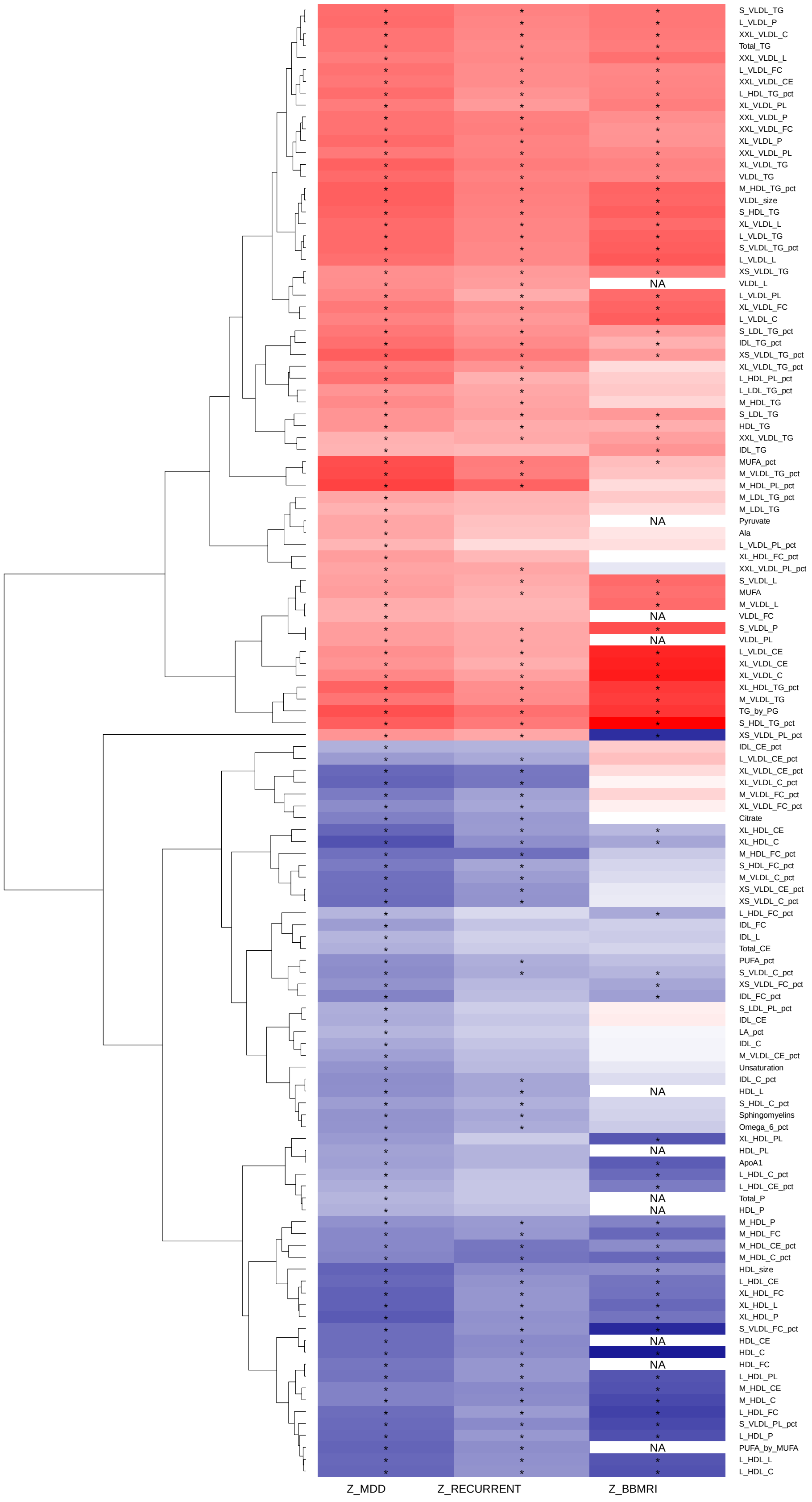
